# Effectiveness of non-pharmacological Interventions For Fatigue in Long term conditions (EIFFEL)-systematic review and network meta-analysis

**DOI:** 10.1101/2025.07.23.25332061

**Authors:** Joanna Leaviss, Andrew Booth, David Coyle, George Daly, Sarah Davis, Helen Dawes, Vincent Deary, Kritica Dwivedi, Jessica Forsyth, Kate Fryer, Samantha McCormick, Marissa Martyn-St James, Julia Newton, Shijie Ren, Gillian Rooney, Anthea Sutton, Mon Mon-Yee, Christopher Burton

## Abstract

**Objective:** To assess the clinical effectiveness of non-pharmacological interventions for fatigue in adults with long term medical conditions.

**Design:** Systematic review and network meta-analysis

**Data sources:** All searches were performed on the following databases: MEDLINE, Embase, CINAHL, APA PsycINFO, Web of Science Core Collection and the Cochrane Central Register of Controlled Trials.

**Methods:** Screening of eligible studies was performed independently and in duplicate, with data extraction and risk of bias assessments conducted by one of two reviewers and validated by the other. Random effects network meta-analyses were conducted for the primary analyses. The primary outcome was self-reported fatigue at end of treatment, short term (up to 3 months after end of treatment) and long term (more than 3 months). The primary network meta-analyses pooled data from all conditions for each time point; a secondary analysis was carried out for separate condition categories. Three rounds of focus groups of people with lived experience of fatigue informed decisions about aggregating data across interventions and conditions, and interpretation of the findings.

**Eligibility criteria for selecting studies:** Randomised controlled trials of non-pharmacological interventions for fatigue in long term medical conditions where fatigue was either a criterion for inclusion, the primary target of the intervention, or the primary or co-primary outcome. We excluded studies of post-infectious, post-traumatic, cancer-related or idiopathic fatigue and limited inclusion to European-style healthcare systems.

**Results:** 88 randomised controlled trials were included, comprising 6636 participants for end of treatment analyses, 1849 (short term) and 2322 (long term), allocated to one of 27 interventions. The most common condition studied was multiple sclerosis (51 studies). Compared to usual care, cognitive behavioural therapy (CBT)-based interventions showed statistically significant reductions in fatigue at end of treatment (standardised mean difference −0.63, 95% credible interval (CrI) −0.87 to −0.4, 17 studies) and long term follow up (−0.4, −0.63 to −0.21,9 studies). Physical activity promotion showed significant reduction in fatigue at all three time points: end of treatment (−0.32, −0.62 to −0.01,7 studies), short term (−0.51, −0.84 to −0.17, 1 study) and long term (−0.52, −0.86 to −0.18, 2 studies). Self-management focusing on energy conservation showed no statistically significant benefit at end of treatment (−0.2, −0.52 to 0.12, 10 studies), short term (−0.13, −0.51 to 0.25, 7 studies) or long term (−0.42, −0.9 to 0.09, 3 studies).

**Conclusions:** Interventions which support individuals to increase physical activity or that are based on cognitive behavioural are effective in reducing fatigue in people with long-term medical conditions. The strength of the evidence for these is moderate to low. Although there are relatively few studies in any condition other than multiple sclerosis, the magnitude of effect appears similar across different conditions.

**Systematic review registration:** PROSPERO CRD42023440141

## Introduction

Persistent fatigue is common in long-term medical conditions^1^. Alongside feelings of tiredness, fatigue includes a sense of needing to rest, or of difficulty in initiating or sustaining voluntary effort^2^ ^3^. People with medical conditions typically describe their fatigue as “more than ordinary tiredness” ^4^ with impacts that go beyond the feeling of fatigue ^5^ ^6^. In addition to wanting their fatigue reduced, and a return to meaningful activities^7^, patients want their experience of fatigue to be validated ^8^. However, many patients report feeling that others, including clinicians, do not take fatigue seriously^9^

While fatigue is common in medical conditions, its presence correlates poorly with disease severity ^10–13^ and it commonly persists after the disease has been brought under control^14^. There appear to be similarities in fatigue across medical conditions, including similarities in experience and impairment ^9^. Current models of fatigue include biological ^15^ and psychosocial factors^1^ ^11^, with increasing interest in the role of altered signalling between the brain and body^16–19^. There are currently no licensed drug treatments for fatigue in long-term conditions.

Non-pharmacological interventions have been developed to overcome fatigue in medical conditions. These include interventions focusing on physical activity (either managing or increasing activity), those that are more psychologically based, as well as a range of forms of non-invasive stimulation, body-mind practices and nutritional supplementation. In practice, many fatigue rehabilitation and self-management programmes contain multiple components. As fatigue is increasingly understood in terms of processes in the body, brain, and signalling between the two^16^ ^18^ ^19^, these different types of non-pharmacological interventions described above are scientifically plausible. However, to many patients with fatigue this rationale is often not apparent.

Thus, proposed interventions may be seen as illogical (physical exercise when they are already exhausted), stigmatising (psychological interventions implying fatigue is “all in the mind” or can be overcome just by thinking differently) or inappropriate (body-mind interventions being too “alternative”). These conceptual barriers to engagement with interventions are an important aspect of this problem^20^.

We found two published network meta-analyses (NMA) of non-pharmacological interventions for fatigue in specific conditions: multiple sclerosis (113 studies) ^21^and post-stroke (10 studies)^22^ as well as one meta-analysis of physical activity interventions across multiple conditions^23^. We found no examples of NMA of the same intervention type across different conditions, suggesting that generalisability across conditions is a largely unanswered question. We therefore conducted a systematic review and network meta-analysis to investigate the clinical effectiveness of non-pharmacological interventions for fatigue in long term conditions more generally. This study was conducted in response to a commissioned call from the UK National Institute of Health & Care Research and comprises one part of a larger evidence synthesis regarding fatigue in long term conditions that includes health economic and qualitative components; these have been submitted for publication separately.

## Methods

This systematic review was conducted and reported in accordance with the Cochrane Handbook for Systematic Reviews of interventions ^24^ and the Preferred Reporting Items for Systematic review and Meta-Analysis guidelines. ^25^ The study eligibility criteria used the PICOS framework. The protocol for this review was registered with the CRD PROSPERO database CRD42023440141. The following alteration from the published protocol was applied: a limitation on included studies to countries with comparable healthcare systems to the UK.

### Patient and public involvement

This review included extensive patient and public involvement (PPI). Two of the investigators were appointed on the basis of their lived experience of fatigue in long term medical conditions. In addition, we convened 5 focus groups involving 25 people with fatigue associated with long term conditions with the primary purpose of ensuring that any assumptions made about grouping interventions or conditions in the statistical analysis were compatible with patients’ experiences.

Participants of focus groups were recruited by advertisement through national peer support organisations and community organisations in South Yorkshire. We invited and recruited purposively to obtain a diverse mixture of long-term conditions and ethnic heritage. Ethics approval was obtained for the focus group study (HRA and Health and Care Research Wales, reference 23/SC/0292).

Focus groups were co-led by PPI investigators (DC and SM) and KF, participants consented to participation and groups were recorded and transcribed for analysis. The focus groups explored important issues in relation to the conduct of the review, particularly the similarities and differences in experience of fatigue between conditions and between interventions. From this, focus groups discussed the appropriateness of combining studies across different conditions. Discussions also considered issues around acceptability and feasibility of different interventions and guidance on framing and content of dissemination materials for patients and professionals.

### Study eligibility criteria

To be eligible for inclusion, studies had to be randomised controlled trials that met the following criteria for population, intervention comparator, outcome and setting.

#### Population

Adults with a long-term condition, using the NHS definition as “an illness that cannot be cured but that can usually be controlled with medicines or other treatments”. The commissioning brief specifically excluded fatigue in people with cancer, in relation to or following from infection (HIV, Hepatitis C, Long Covid and ME/Chronic Fatigue Syndrome) or resulting from injuries or developmental disorders. It also excluded conditions in which symptoms, rather than observable pathology, were the defining features (e.g. fibromyalgia or irritable bowel syndrome).

#### Interventions

We included studies of any non-pharmacological intervention in which a stated explicit aim or primary outcome was to address fatigue. These included behavioural, exercise based, and nutritional interventions as well as a range of forms of non-invasive stimulation. We excluded interventions that were specific to a condition (e.g. pulmonary rehabilitation in lung disease) or to a problem other than fatigue (e.g. vestibular rehabilitation for balance problems in people with multiple sclerosis). Interventions could be delivered face-to-face or at a distance and included technology-assisted interventions.

#### Comparators

Comparators were “usual care”, waiting list control, sham or placebo (for stimulation or nutritional interventions), another non-pharmacological intervention or attentional control such as education or information.

#### Outcomes

Primary outcome: we required that studies reported an established measure for fatigue. We allocated three time points for follow up. These were end of treatment, short term (up to 3 months after the end of treatment), and long term (more than 3 months after the end of treatment). Where studies reported multiple long term time points, we extracted data for each of these, with the primary analysis using the longest follow up data.

#### Setting

Studies could be conducted in primary, secondary, or community-based settings, however we only included studies which could feasibly be delivered in an outpatient or community-based setting. We excluded studies set in countries with healthcare systems that are not comparable to the UK.

### Information sources

A comprehensive search of bibliographic databases to identify randomised controlled trials (RCTs) was conducted in September-October 2023 and updated in September 2024. Search strategies combined free-text and thesaurus terms related to long-term conditions (both specific conditions and general terms such as “chronic disease” and “long-term illness”), and terms for fatigue measures (specifically named scales, and general terms for fatigue and assessment). Methodological search filters were used to identify RCTs. No date or language limits were applied to the search. All searches were performed on the following databases: Ovid MEDLINE, Embase (via Ovid), CINAHL (via EBSCO), APA Psycinfo (via Ovid), Web of Science Core Collection (Science Citation Index and Social Sciences Citation Index). Additionally, the Cochrane Central Register of Controlled Trials (CENTRAL) was searched for RCTs, and the Cochrane Database of Systematic Reviews (CDSR) was searched for systematic reviews. All databases were searched from inception to the respective search dates.

There was no limit on date of study inclusion. Details of the search strategies are in Supplemental methods 1

### Study selection and data collection

We carried out a two-stage sifting process for inclusion of studies, (title/abstract then full paper sift), using Covidence (Veritas Health Innovation) to manage the selection process. 3 reviewers initially reviewed 10% of titles and abstracts according to pre-specified inclusion and exclusion criteria. Issues relating to ambiguity of any criteria were resolved by team discussion. All remaining titles and abstracts were then scrutinised independently by two reviewers (Cohen’s kappa 0.58). Full texts of potentially eligible studies were then assessed for eligibility. Discrepancies were resolved by discussion between the two reviewers in consultation with a third investigator (CB) if required. The most common discrepancies were concerned with the cut-off criteria for inclusion where boundaries were blurred, for example whether the intervention focus was managing fatigue or whether fatigue was one of multiple secondary outcomes in a general condition self-management intervention. The update search was sifted using the same eligibility criteria.

Two reviewers (JL & GR) extracted the following data, with intervention characteristics using the ‘Template for Intervention Description and Replication statement’ (TIDieR) ^26^. Data extraction aimed to reflect sources of complexity such as: population differences e.g. diagnostic criteria of the included long-term conditions; the use of multiple components within interventions; the expertise and skills of those delivering and receiving the intervention; the intervention context including method and intensity of delivery; settings; timepoints of outcome measurement; attrition. Results (estimates and corresponding standard errors (SE), standard deviations (SD), confidence intervals (CI) or inter quartile ranges (IQR)) were also extracted by one of two reviewers (JL & GR), and double checked by the other. Given the large numbers of included studies it was not feasible to contact the authors of included studies to enquire about missing or incomplete data or data that was only included graphically. Interventions were coded into categories using the method described below.

### Risk of Bias Assessment of included studies

Risk of bias assessment of all studies included in the NMA of the present review was undertaken using an adapted version 2 of the Cochrane risk-of-bias tool (RoB2) for RCTs ^27^. For pragmatic reasons relating to the volume of studies included in the NMA in the present review we adapted the RoB2 tool to facilitate quicker completion, reducing the number of signalling questions from 22 to 15 within five domains. A full description of these methods is presented in Supplemental methods 2.

### GRADE assessment

Review findings were synthesised using an adaptation of the Grading of Recommendations Assessment, Development and Evaluation (GRADE) framework^28^ to assess the quality of the evidence (certainty in the evidence) for fatigue for each intervention compared to usual care, at each of the three timepoints analysed in the NMA. We adapted GRADE to incorporate elements of CINeMA,^29^ a framework largely based on GRADE, modified to facilitate network meta-analysis. Whilst we adopted the assessment framework of CINeMA (e.g. methods of assessing heterogeneity and inconsistency), we used a Bayesian approach to analysis rather than the current CINeMA analysis platform. We used a framework based on risk of bias, inconsistency, imprecision and heterogeneity. We used a threshold of an SMD of 0.34 as clinically meaningful (see below for rationale). A full description of the methods used for the GRADE assessment is provided in Supplemental methods 3.

### Classification of conditions

An initial description of the condition (checked against exclusion criteria) was generated for all extracted studies. These were aggregated into broad disease categories (e.g. musculoskeletal disorders). Within neurological disorders, multiple sclerosis and stroke were kept separate from other neurological disorders.

### Classification of interventions

This followed an iterative inductive approach. First a simple description of the interventions in each arm was recorded by during data extraction. Next a clinical investigator reviewed these to generate an initial classification with draft criteria for each category. The same investigator then reviewed the full-text descriptions of interventions and classified them using the draft criteria: during this process the criteria were edited and refined following discussions with other clinical investigators. These criteria were then reviewed and tested (for a sample of behavioural interventions) by independent checking of categorisation. Differences were resolved by discussion. The final criteria (Supplemental methods 4) were then re-applied to all included studies. In parallel with this, we grouped the individual intervention categories into higher level groups to produce a hierarchical taxonomy. We took this approach as many interventions had multiple (often overlapping) components although in varying amounts.

### Use of patient focus group and other qualitative data to inform our analysis

From the patient focus groups and a parallel qualitative evidence synthesis (Booth, personal communication) we identified three key observations to guide decisions about inclusion of interventions and conditions for analysis. These were: (1) The experience of fatigue is multifaceted and different for each individual; differences (and similarities) are as evident within conditions as between conditions, (2) while few focus group participants had experience of specific interventions for fatigue, none of those discussed was unacceptable to most participants, (3) personal circumstances and experience were important in valuing interventions. These observations informed our study design choices to carry out the primary analysis across conditions, to have no a priori restriction on interventions and to recognise the importance of personal context in recommendations arising from the analysis. Additional data relating to the focus groups is provided in Supplemental methods 5

### Statistical Analysis

The primary analysis consisted of three separate NMAs, each corresponding to a different follow-up time point. We used standardised mean difference (SMD) of the change in fatigue outcomes from baseline as the measure of effect, evaluated using Hedge’s correction for small studies. The detailed methodology is described in Supplemental methods 6. We generated networks of evidence at the three time points (end of treatment, short term and longer term) and conducted an NMA at each time point, using a random-effects model in view of the heterogeneity of study design, intervention and population^30^. Parameters of the random-effects model were estimated using a Bayesian framework, with non-informative parameter priors. All analyses were conducted using WinBUGS ^31^ via the R package, R2WinBUGS ^32^. Results are presented as the posterior median treatment effects and 95% credible intervals (CrI). Study heterogeneity was assessed and interpreted using established categories ^33^. Consistency was checked by comparing the posterior mean residual deviance from the unrelated mean effects model and the NMA model; and node-splitting analysis^34^

We conducted three secondary analyses. These were (1) to examine the sensitivity of the findings to different rules about preferred time point in longer term follow up studies; (2) to examine condition (or condition-group) specific networks and (3) to exclude studies identified as pilot or feasibility studies. Finally, in order to translate findings from the SMD into clinically meaningful values we took the estimated clinically important difference on the Fatigue Severity Scale (FSS) ^35^ and mapped it via an estimate of the baseline SD of studies within the EOT network which used FSS in order to calculate the corresponding clinically meaningful SMD.

## Results

After de-duplication, 10108 titles and abstracts were reviewed. From these, 1068 full-text articles were assessed for eligibility, of which 118 studies reported in 120 manuscripts were eligible for inclusion. Of these, 88 studies, reported in 90 manuscripts, were included in the NMA (see figure 1 and Supplemental results 1). The 30 studies not included in the NMA are listed in Supplemental results 2 with reasons for non-inclusion in Supplemental results 3. Timepoints at which included studies reported results are in Supplemental results 4

**Figure 1.**
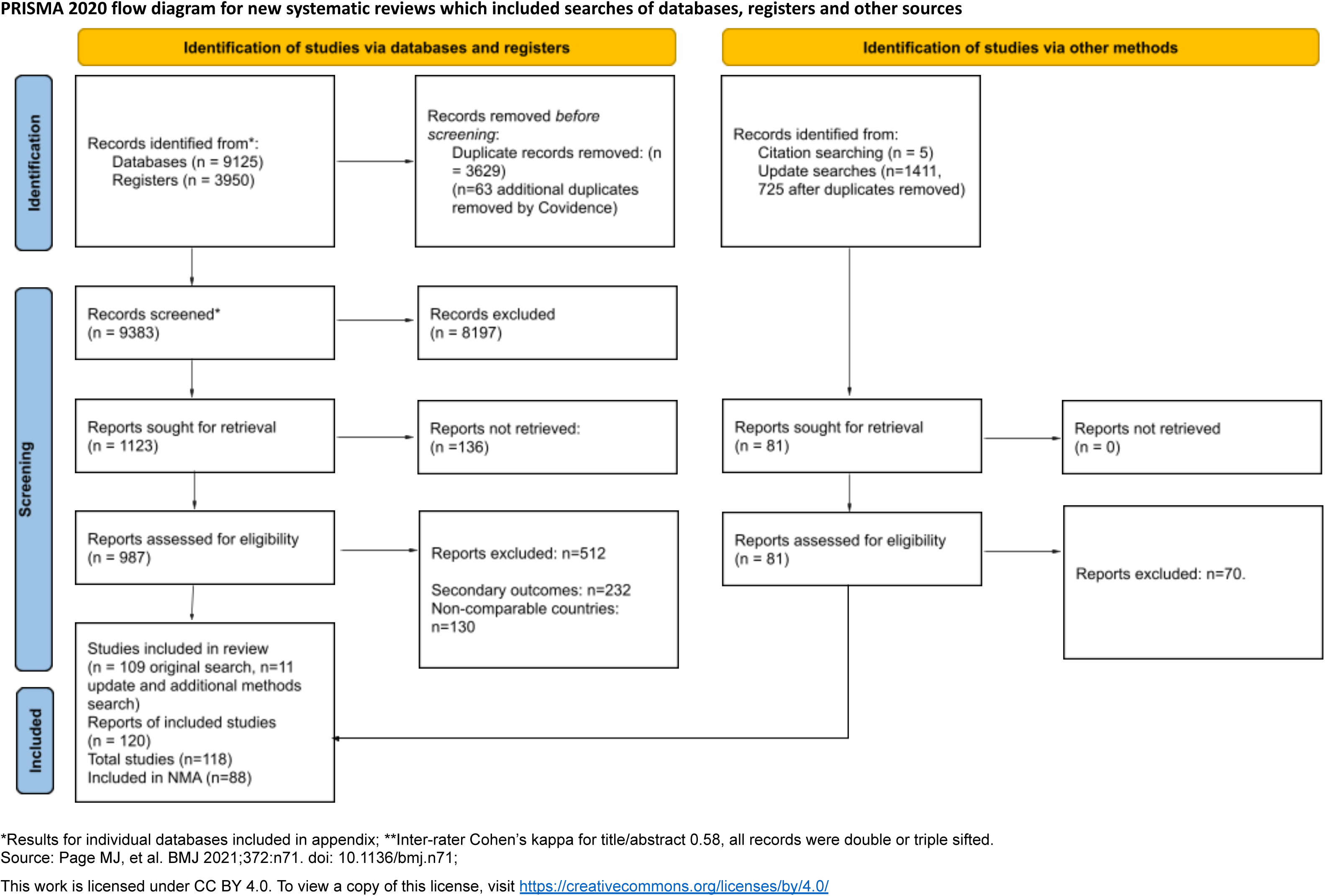
PRISMA Flow Diagram.

### Medical Conditions

The most common condition was multiple sclerosis (51 studies). There were 6 studies in stroke and 5 in other neurological conditions. 20 studies involved a range of musculoskeletal and connective tissue disorders ranging from osteoarthritis to systemic sclerosis. The remaining studies included inflammatory bowel disease (6) chronic kidney disease (3) and diabetes, hypothyroidism, heart disease and psoriasis (1 each).

### Interventions

Table 1 shows the distribution of interventions by conditions. The most common intervention was CBT based interventions (19 studies). Other self-management interventions were energy conserving fatigue management (11), activating fatigue management (3) and general self-management (6). 28 interventions were focused on physical activity including supervised exercise (14), unsupervised exercise (7) and physical activity promotion (7). Other intervention categories were less common, often with single instances of distinct interventions within a category. Interventions were delivered to individuals and to groups, using in person, phone and online formats.

**Table 1.** Distribution of interventions by medical condition.

|  | Intervention | Long term Condition |  |  |  |  |  |  |  | Total |
| --- | --- | --- | --- | --- | --- | --- | --- | --- | --- | --- |
|  |  | MS | Stroke | Neuro | MSK | Kidney | IBD | Endo. | Other |  |
| **Exercise-based**<br>**(N=30)** | Exercise Supervised | 12<sup>36-45</sup> | - | 1<sup>46</sup> | 1<sup>47</sup> | - | - | - | 2<sup>48-49</sup> | 16 |
|  | Exercise Unsupervised | 4<sup>42-50-52</sup> | - | - | 3<sup>53-55</sup> | - | - | - | - | 7 |
|  | Physical activity promotion | 3<sup>52-56-57</sup> | - | - | 4<sup>47-54-58-59</sup> | - | - | - | - | 7 |
| **Self -**<br>**management**<br>**(N=40)** | CBT-based intervention | 7<sup>60-66</sup> | 3<sup>67-69</sup> | 1<sup>70</sup> | 3<sup>20-58-71</sup> | 2<sup>72-73</sup> | 2<sup>74-75</sup> | 1<sup>76</sup> | - | 19 |
|  | Active Fatigue Self-Management | 1<sup>77</sup> | 1<sup>78</sup> | - | 1<sup>79</sup> | - | - | - | - | 3 |
|  | Conservative Fatigue Self-Management | 8<sup>80-87</sup> | - | 1<sup>88</sup> | 1<sup>89</sup> | 1<sup>90</sup> | - | - | - | 11 |
|  | General Self-Management | 1<sup>86</sup> | 1<sup>78</sup> | - | 4<sup>91-94</sup> | - | - | - | - | 6 |
|  | Rehabilitation | 1<sup>95</sup> | - | - | - | - | - | - | - | 1 |
| **Mind & Body**<br>**(N=14)** | Mind-Body | 7<sup>65-84-87-96-98</sup> | - | 1<sup>99</sup> | 2<sup>55-100</sup> | - | - | - | - | 10 |
|  | Mindfulness based | 2<sup>52-101</sup> | - | - | - | - | 1<sup>102</sup> | - | - | 3 |
|  | Other Psychological | - | 1<sup>69</sup> | - | - | - | - | - | - | 1 |
| **Stimulation**<br>**(N=14)** | External stimulation | 4<sup>103-106</sup> | - | - | - | - | - | - | - | 4 |
|  | Acupuncture-type | - | - | 1<sup>107</sup> | - | - | 1<sup>108</sup> | - | - | 2 |
|  | Aromatherapy | - | - | - | - | - | - | 1<sup>109</sup> | - | 1 |
|  | Transcranial Stimulation | 4<sup>110-113</sup> | - | - | - | - | - | - | - | 4 |
|  | Vagal Nerve Stimulation | - | - | - | 2<sup>114-115</sup> | - | - | - | - | 2 |
|  | Remote Ischaemic Conditioning | - | 1<sup>116</sup> | - | - | - | - | - | - | 1 |
| **Nutritional**<br>**(N=6)** | Nutritional Supplement | 1<sup>117</sup> | - | - | 1<sup>118</sup> | - | 2<sup>119-120</sup> | - | - | 4 |
|  | Diet | 1<sup>121</sup> | - | - | - | - | - | - | - | 1 |
|  | Plant-based | 1<sup>122</sup> | - | - | - | - | - | - | - | 1 |
|  | Total | 57 | 7 | 5 | 22 | 3 | 6 | 2 | 2 | 104 |

Duration of the interventions ranged from three weeks to six months, although most behavioural interventions lasted between 6 and 12 weeks. More detailed descriptions of interventions content and delivery by study are provided in Supplemental results 5-7.

### Risk of Bias

An overall summary of the risk of bias assessments is presented in Figure 2. Individual study risk of bias is reported in Supplemental results 6 and 7. Overall, whilst the body of evidence contains some larger trials, many of the studies are small, under-powered or pilot/feasibility studies. Furthermore, the large majority of trials involved at least one behavioural arm (e.g. physical activity or self-management) for which blinding was impossible because of the nature of the intervention, resulting in high risk of bias judgements in accordance with the RoB v2.0 guidance^27^. A summary of key findings by domain is presented below.

**Figure 2.**
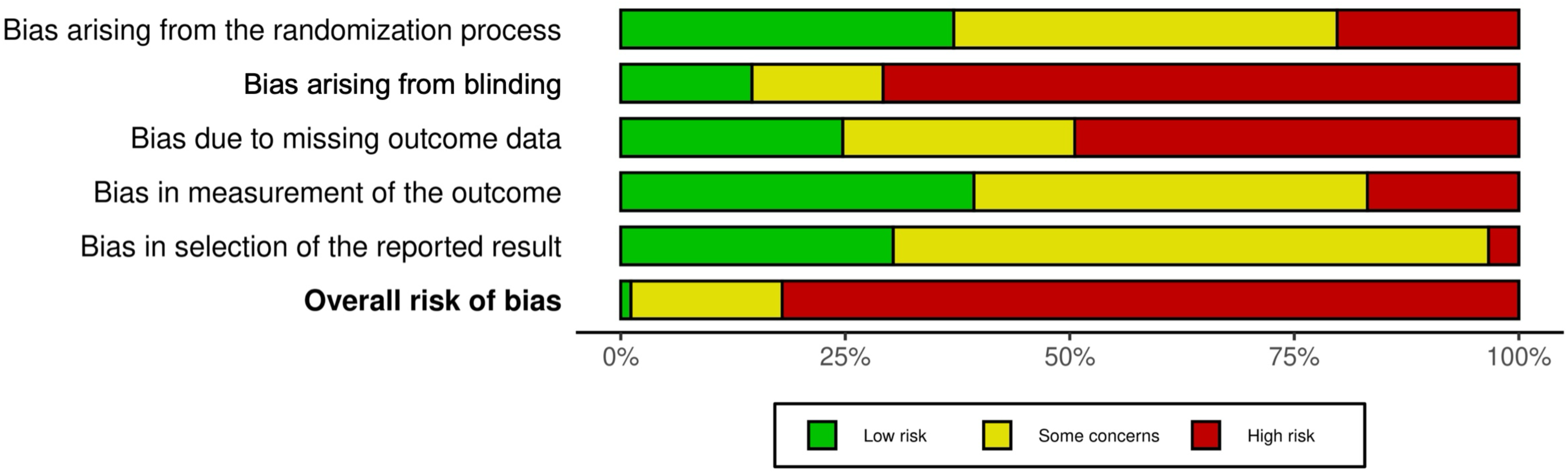
Summary of risk of bias for all included studies.

#### Risk of bias from the randomisation process

Whilst all included studies described themselves as randomised controlled trials, nearly a quarter of the studies did not provide enough detail on the method of randomisation to make a judgement on whether there was a potential risk of bias. Of the studies that were judged to be at high risk of bias for randomisation, this was mostly due to the use of simple randomisation (alternate or manual). One study used a matched control group as a third arm, and another created an additional control arm after randomisation for participants who declined their allocated interventions.

#### Risk of bias due to blinding

Blinding was rarely possible due to the nature of the interventions, most of which were behavioural. Whilst we acknowledge this practical restriction on study design, this still introduces a risk of bias. Lack of blinding of participants and care givers was the most common risk of bias across all studies. Where blinding was possible, e.g. studies of interventions with placebo or sham controls, it was not conducted in all cases.

#### Risk of bias due to missing outcome data

The sample size in only half of the studies was based on a power calculation. Many of the studies that did not use a power calculation were reported by the authors to be underpowered. Around a quarter of studies reported high attrition (>20%), and for many of these, the withdrawals were not balanced between study arms.

#### Risk of bias from measurement of the outcome

Around a half of the studies were at low risk of bias for measurement of outcome, due to blinding of outcome assessment. In the remaining studies, blinding of those conducting the outcome assessment was either specifically reported to have *not* been conducted, or did not provide details on the process.

#### Risk of bias from selective reporting

The majority of studies were reported to be on trials registries, mostly NCT or ISRCTN. It was not possible to locate protocols for many studies within the time and resources of the review, and we therefore loosened our criteria, using the study plans on the trials registries to make our judgements where a full protocol was not readily accessible.

Where there was a protocol or study plan identified, outcomes were mostly analysed as per the protocol.

### Primary analysis

#### Network geometry

Network diagrams for the primary analysis at the different time points are shown in Figure 3. The networks contained 27 connected interventions (including control interventions) at the end of treatment, 16 at short term and 13 at long term follow up. They were evidenced from 84 studies (6636 participants), 24 studies (1849 participants) and 18 studies (2322 participants) respectively. To ensure connectivity of the networks, the “Control” node includes “Control”, “Placebo” and “Sham” interventions. “Information and education” is also included as a comparator rather than an intervention.

**Figure 3.**
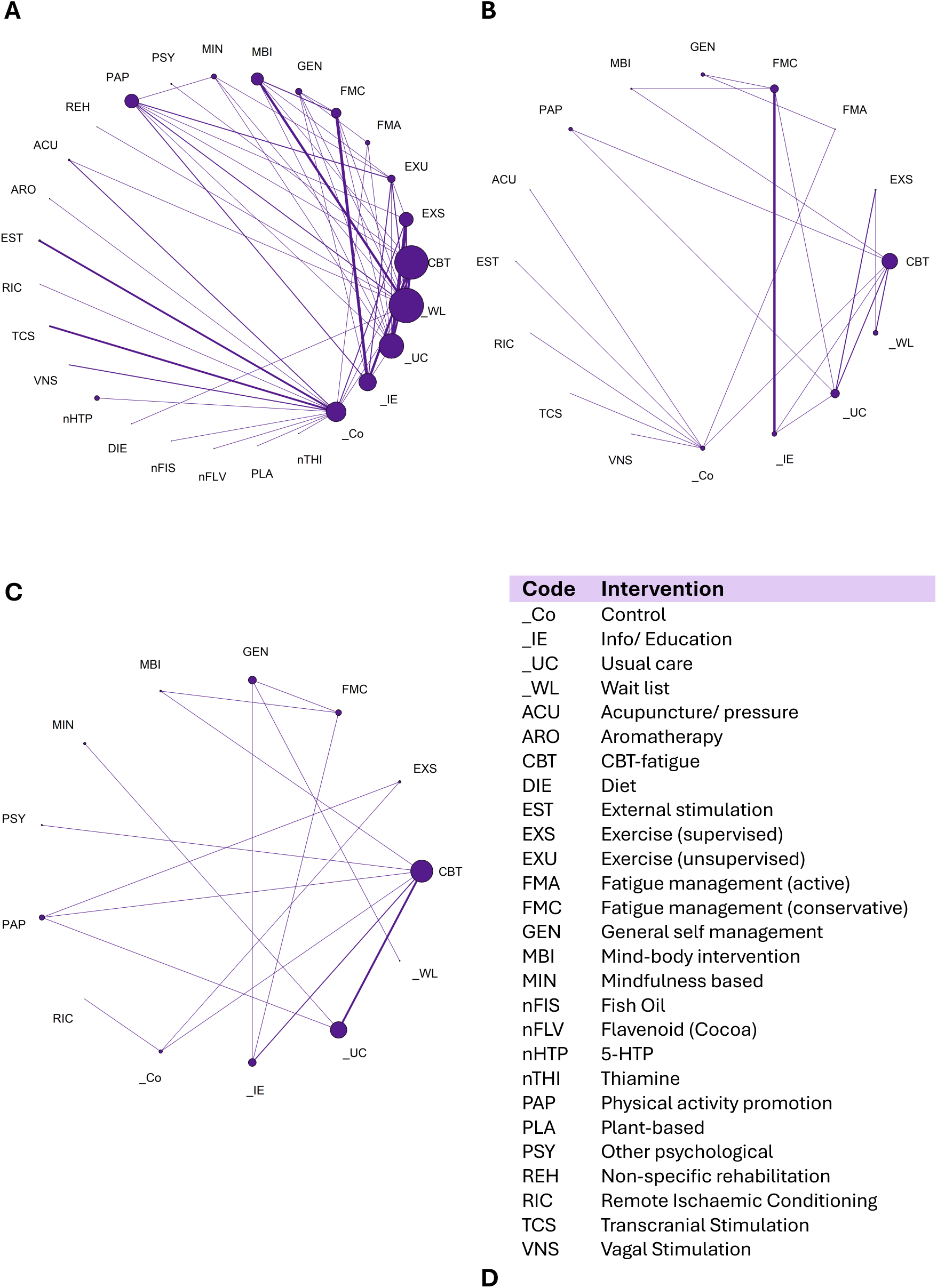
Network geometry for A) end of treatment, B) short term, and C) long term analyses, respectively, indicating the number of participants who received each intervention (size of node) and the number of studies contributing to the direct evidence and comparisons between interventions (thickness of line). D) Key of intervention coding used in network geometry.

#### Synthesis of results

Figures 4 and 5 show the predicted SMDs for each intervention within the networks at each time point. A negative SMD indicates a reduction in fatigue relative to usual care. Within each of the forest plots, the final group (“Other”) represents interventions typically included as comparator interventions. When assessing for inconsistency within the networks, no statistically significant inconsistency was detected within the primary analysis, (see Supplemental statistical data).

**Figure 4.**
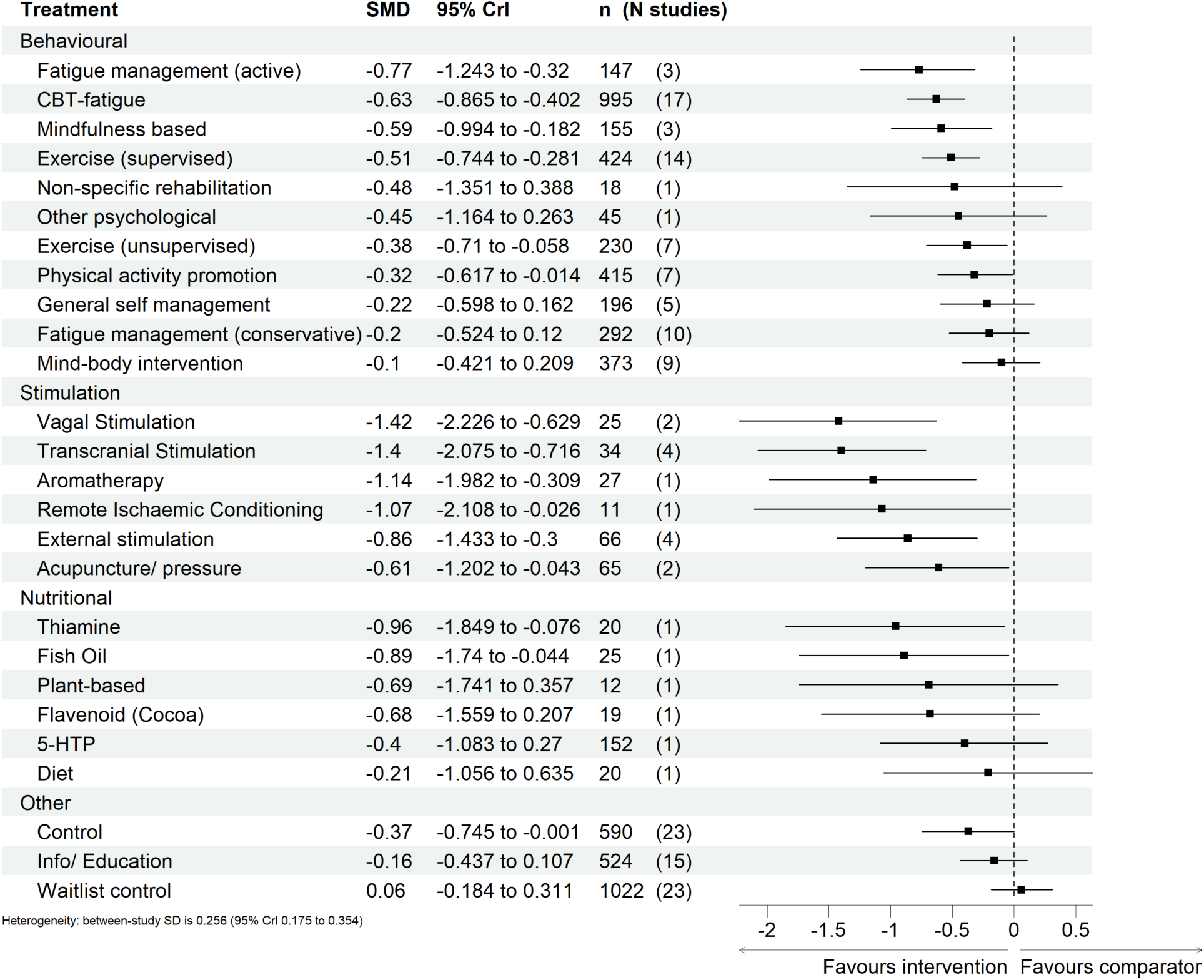
Predicted effects on fatigue outcomes of interventions, relative to usual care, at end of treatment, with 95% credible intervals (CrI). The number of participants (n) and the number of studies (N studies) are given for context. Broad intervention categorisation is also presented to aid interpretation (Behavioural, Stimulation, Nutritional, and Other). The “control” node is displayed as this functioned to ensure connectivity of the network, but this is not an active intervention for consideration/recommendation.

**Figure 5.**
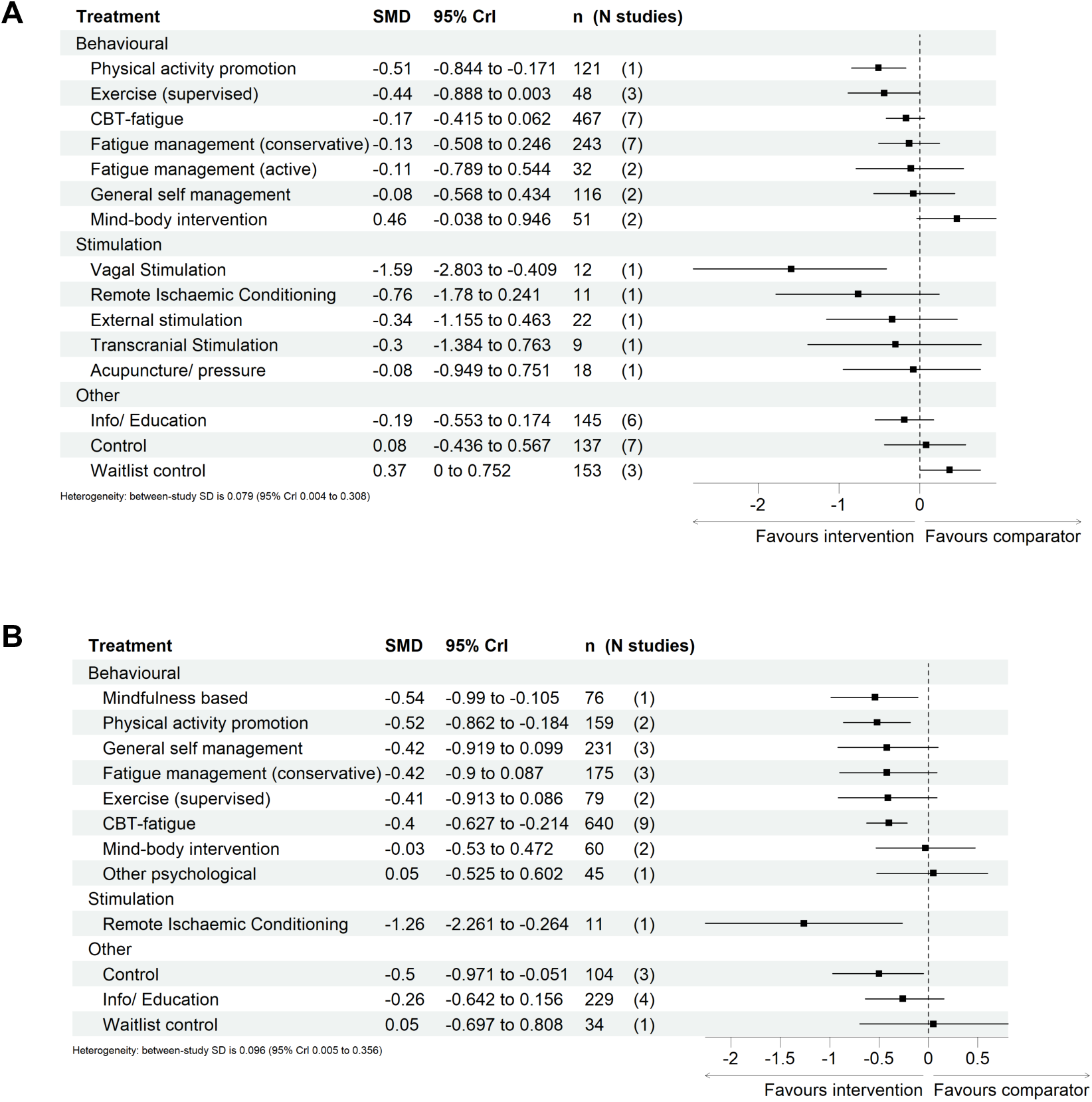
Predicted effects on fatigue outcomes of interventions, relative to usual care, at A) short term and B) long term follow up, with 95% credible intervals (CrI). The number of participants (n) and the number of studies (N studies) are given for context. The “control” node is displayed as this functioned to ensure connectivity of the network, but this is not an active intervention for consideration/recommendation.

Relative to usual care, CBT-based interventions showed statistically significant reductions in fatigue at end of treatment (SMD −0.63, 95% CrI −0.87 to −0.4, 17 studies) and long term follow up (−0.4, −0.63 to −0.21,9 studies). The reduction at short term, with fewer studies was smaller and not statistically significant (−0.17, −0.42 to 0.06, 7 studies). Active fatigue management showed a statistically significant reductions in fatigue at end of treatment −0.77 (−1.2 to −0.32, 3 studies) but this was not sustained to short term (2 studies) and no studies reported long term follow up. Conservative self-management showed no statistically significant change in fatigue at end of treatment (− 0.2, −0.52 to 0.12, 10 studies), short term (−0.13, −0.51 to 0.25, 7 studies) or long term (−0.42, −0.9 to 0.09, 3 studies). Mindfulness-based interventions showed statistically significant reductions in fatigue at end of treatment (−0.59, −0.99 to −0.18, 3 studies) and long term (−0.54, −0.99 to −0.11, 1 study).

Physical activity promotion showed significant reduction in fatigue at all three time points: end of treatment (−0.32, −0.62 to −0.01,7 studies), short term (−0.51, −0.84 to - 0.17, 1 study) and long term (−0.52, −0.86 to −0.18, 2 studies). Supervised exercise showed statistically significant reductions in fatigue at end of treatment (−0.51, −0.74 to −0.28, 14 studies) but SMDs at short term (−0.44, −0.89 to 0.003, 3 studies) and long term (−0.41, −0.91 to 0.09, 2 studies), while of comparable magnitude, were not statistically significant.

Non-invasive stimulation studies were few in number and small in size (14 studies, 228 participants). While observed effects at end of treatment were large, only 5 studies (72 participants) reported effects at short term and 1 study (11 participants) reported longer term follow up. Nutritional studies reported end of treatment results only. Estimated effect sizes in the NMA for non-invasive stimulation and nutritional studies appear larger than reported in the original papers because SMDs are estimated relative to usual care, while these were compared to sham or placebo which in turn had a greater effect than usual care.

In each of the primary analyses, there was moderate heterogeneity indicating potentially varying treatment effects between studies. The standard deviation of the between study heterogeneity was greatest in the end of treatment analysis (0.256, 0.175 to 0.354) and comparable within the short and longer term networks (0.079, 0.004 to 0.308) and (0.096, 0.005 to 0.356) respectively. This suggests that there are generally smaller differences between the study design, interventions and populations in the short and longer term networks compared to the end of treatment network.

#### Sensitivity analysis

Results of the sensitivity analysis are presented in Supplemental statistical data

#### Studies with multiple follow up time points after 3 months

Five studies included within the long-term analysis had data available from more than one time point after 3 months. Re-analysing the longer term data, instead using the shortest follow up point after 3 months had minimal impact on the results of the NMA

#### Sensitivity analysis: condition specific analyses

Due to the sparsity of evidence other than for multiple sclerosis, meaningful networks could only be constructed for the following conditions or condition groups: multiple sclerosis (at all three time points), musculoskeletal (end of treatment and long term), and inflammatory bowel disease, kidney disease and stroke (end of treatment only). The networks were small other than for multiple sclerosis so predicted treatment effects were generally associated with large uncertainty.

#### Sensitivity analysis: exclusion of pilot and feasibility studies

Reanalysis after removal of pilot and feasibility studies had minimal impact of the results of the NMA, although it did result in the exclusion of some interventions from the networks.

#### Clinically important difference

We estimated that a clinically important difference of 3.6 points on the Fatigue Severity Scale (range 9-63) ^35^ was equivalent to a SMD in the network meta-analysis of 0.34.

This indicates that the effect sizes which reached statistical significance were also likely to be clinically meaningful.

The certainty of the evidence for the observed intervention effects using our adapted GRADE framework is summarised below for all timepoints.

The certainty of the evidence for the observed intervention effects at short term and long term follow up is summarised in table 2. In summary, at long term (more than 3 months after end of treatment) the evidence for physical activity promotion (interventions which supported individuals to increase physical activity) was rated as moderate. The evidence for CBT-based interventions and mindfulness was rated as low. The strength of evidence for all other interventions was rated as very low.

**Table 2.** Summary of risk of GRADE evidence: fatigue as outcome at long term follow up (at least 13 weeks after end of treatment).

| **Intervention** | **Summary of findings** | **Quality of evidence** | **Reason for grading up or down** |
| --- | --- | --- | --- |
| **Behavioural** |  |  |  |
| Physical activity promotion | Evidence of a medium effect from 2 studies with 159 participants | Moderate | RoB - down 1 |
| CBT based intervention | Evidence of a small effect from 9 studies with 640 participants | Low | RoB - down 1; Inconsistency - down 1 |
| Mindfulness –based intervention | Evidence of a medium effect from 1 study with 76 participants | Low | RoB - down 1; Inconsistency - down 1 |
| General self-management | Non significant evidence of a small effect from 3 studies with 231 participants | Very low | RoB - down 1; Inconsistency - down 1; Imprecision - down 1; Heterogeneity - down 1 |
| Fatigue management (conservative) | Non significant evidence of a small effect from 3 studies with 175 participants | Very low | RoB - down 1; Inconsistency - down 1; Imprecision - down 1; Heterogeneity - down 1 |
| Exercise (supervised) | Non significant evidence of a small effect from 2 studies with 79 participants | Very low | RoB - down 1; Inconsistency - down 1; Imprecision - down 1; Heterogeneity - down 1 |
| Mind-body intervention | Non significant evidence of unsubstantial effect from 2 studies with 60 participants | Very low | RoB - down 1; Inconsistency - down 1; Imprecision - down 2; Heterogeneity - down 2 |
| Other psychological | Non significant evidence of unsubstantial effect from 1 study with 45 participants | Very low | RoB - down 1; Inconsistency - down 1; Imprecision - down 2; Heterogeneity - down 2 |
| **Non-invasive Stimulation** |  |  |  |
| Remote ischaemic conditioning | Evidence of a large effect from 1 study with 11 participants | Very low | RoB - down 1; Inconsistency - down 1; Heterogeneity - down 1 |

## Discussion

### Summary of main findings

Non-pharmacological interventions for fatigue in long term conditions other than multiple sclerosis have received relatively little attention in terms of large well conducted randomised studies, and have rarely been conducted across conditions. Nevertheless, we found evidence of effectiveness of interventions that increase physical activity or are based on cognitive behavioural therapy. We found no significant benefit from approaches to self-management in fatigue which focused on energy conservation. These findings appeared relatively consistent across conditions in keeping with other evidence of similarities in fatigue across conditions. The evidence generally carries high risk of bias, although this is at least due to taking a strict approach to judgement of risk of bias involving blinding.

### Strengths and limitations

Strengths of this review include the broad scope both of eligible conditions and non-pharmacological interventions. This was underpinned by extensive patient and public involvement to ensure that assumptions made by researchers were concordant with the lived experience of people with fatigue in long term conditions. We used network meta-analysis to combine evidence across multiple conditions and interventions to maximise the available information in light of our focus groups and qualitative evidence synthesis that identified substantial similarities across conditions.

This review was limited by issues common to other reviews of complex interventions relating to eligibility of studies, choice of time points, categorisation of interventions and the large number of small studies. We identified many studies that included fatigue as one of multiple outcomes. Our inclusion criteria were restricted to studies where fatigue was the primary focus (in terms of either the population, the proposed mechanism of intervention, or the primary outcome). However, this created a grey area, particularly with self-management type interventions, where subjective judgement and resolution through discussion was often required. It is possible that another review team might have operationalised this differently. Although we took this relatively strict approach to inclusion, the small number of studies of general condition self-management which did meet our inclusion criteria showed no significant effect, suggesting this approach was justified.

The timepoints at which outcomes were measured varied between studies. We anchored timepoints to the expected end of treatment rather than enrolment. In practice this meant that while the boundary between short term and long term treatment had a similar relationship to time of enrolment within studies with similar durations of intervention, this was not necessarily the case comparing across interventions. As the networks are sparse, results of the statistical analyses may be affected were the timepoint categories different. In particular, we considered that studies with longer follow up may be penalised relative to studies with shorter long term follow up due to attrition of effect or follow up, however the sensitivity analysis found no significant evidence of this.

The majority of studies were of behavioural interventions, often using pragmatic designs, and therefore blinding of participants was not possible. Risk of bias was therefore rated as high for almost all studies. The evidence base includes studies with heterogenous interventions, comparators and timepoints. Many interventions comprised of multiple components, some of which were common to interventions across categories. We were not able to conduct a component NMA due to limited data. Rather we developed a classification of interventions and applied a best-fit principle of allocation. In some cases, this may have obscured results – for instance the small category of Mind-body interventions included several studies where relaxation was used as a low intensity comparator intervention. Further, many different fatigue scales were used to measure outcomes across studies. This necessitated statistical standardisation and may have increased uncertainty due to the inherent differences between scales used due to the potential variability in focus of the different fatigue scales. Future trials in this area would benefit from more standardised methodology, to reduce the observed uncertainty and enable more confident interpretation of results across studies. Many studies were small, including pilot/feasibility studies, however excluding pilot and feasibility studies had little effect on the key findings. A few studies evaluated emerging treatments, in particular non-invasive transcranial and vagus nerve stimulation. These showed potentially large short term effects, and whilst they are still at an experimental stage they do appear to warrant further research.

### Relationship to existing research

This was the first transdiagnostic review of multiple non-pharmacological interventions for long term conditions although we were aware of two previously published reviews in multiple sclerosis ^21^and stroke ^22^. We applied stricter inclusion criteria in comparison to these reviews, in order to focus on fatigue outcomes. We identified one transdiagnostic review of physical activity promotion for fatigue which found sustained benefits with an estimated SMD slightly larger than those from our NMA^23^. We also restricted studies to those conducted within Western healthcare systems and culture to maximise transferability to those systems.

### Implications for practice, policy and research

The review findings provide evidence for the effectiveness of interventions that promoted an increase in physical activity or were based on CBT. While effects were observed across different long-term conditions, all interventions were delivered within single conditions. We found some evidence for mindfulness based intervention and some forms of non-invasive stimulation which may warrant further research. We found no single clear best intervention and from our parallel focus group work and qualitative evidence synthesis recognise that offering patients a choice of interventions is better than aiming for a single best treatment for all. We recommend that further research focuses on delivery of interventions in a transdiagnostic format.

### Conclusion

Interventions which support individuals to increase physical activity or that are based on cognitive behavioural are effective in reducing fatigue in people with long-term medical conditions. Strength of the evidence for these is moderate to low. Although there are relatively few studies in any condition other than multiple sclerosis, the magnitude of effect appears similar across different conditions.

## Data Availability

Search and data extraction tables from the systematic review will be made available on a public server at the University of Sheffield on publication of relevant studies.

## Acknowledgements

We wish to acknowledge the valuable support and wisdom of the members of our project oversight committee, Professors, Neil Basu, Hans Knoop, Bernd Löwe, and Kate Seers and the practical administrative support of Lauren Hartley. HD is supported by the NIHR Exeter Biomedical Research Centre.

## Conflict of Interest

The authors declare that they have o conflict of interests

## Ethics statement

Ethics approval was obtained for the focus group study from HRA and Health and Care Research Wales, reference 23/SC/0292.

## EIFFEL Supplementary Methods

## 1 Supplementary Methods 1: Detailed search strategy

Search Strategies: RCT search

Ovid MEDLINE(R) ALL <1946 to September 27, 2023>

- exp Chronic Disease/ 625076
- ((chronic or long-term or long term) adj (condition* or disease* or illness*)).ti,ab. 129174
- chronically ill.ti,ab. 6151
- exp Rheumatic Diseases/ 261192
- rheumati*.ti,ab. 63183
- exp Diabetes Mellitus/ 511411
- diabet*.ti,ab. 776423
- exp Endocrine System Diseases/ 1133836
- exp Thyroid Diseases/ 163788
- exp Adrenal Gland Diseases/ or exp Adrenal Insufficiency/ 72739
- exp Autoimmune Diseases/ 546702
- ((endocrine or thyroid or adrenal or autoimmune or auto-immune or auto immune) adj1 (disorder* or disease* or condition*)).ti,ab. 122720
- adrenal insufficiency.ti,ab. 7311
- exp Heart Failure/ 148655
- heart failure*.ti,ab. 209132
- exp Coronary Disease/ 236955
- coronary heart disease*.ti,ab. 55465
- exp Renal Insufficiency, Chronic/ 135178
- exp Kidney Failure, Chronic/ 101277
- (chronic adj (renal or kidney) adj (insuffucien* or failure* or disease*)).ti,ab. 98172
- exp Renal Dialysis/ 126720
- dialysis.ti,ab. 122294
- exp Transplants/ 31895
- (transplant* adj3 (heart* or kidney* or liver* or lung*)).ti,ab. 180737
- exp Multiple Sclerosis/ 70469
- multiple sclerosis.ti,ab. 90255
- exp Stroke/ 174658
- stroke.ti,ab. 305418
- exp Neurodegenerative Diseases/ 371816
- ((neurodegenerative or neuro-degenerative or neuro degenerative) adj (disease* or disorder* or condition*)).ti,ab. 99788
- exp Parkinson Disease/ 82813
- (parkinson* adj disease).ti,ab. 116256
- exp Arthritis, Rheumatoid/ 126357
- rheumatoid arthritis.ti,ab. 120734
- exp Osteoarthritis/ 78077
- osteoarthritis.ti,ab. 84060
- exp Lupus Erythematosus, Systemic/ 67452
- lupus.ti,ab. 88490
- exp Scleroderma, Systemic/ 23231
- (systemic sclerosis or scleroderma).ti,ab. 28912
- exp Inflammatory Bowel Diseases/ 97805
- (inflammatory bowel disease* or IBD).ti,ab. 67194
- exp Liver Cirrhosis, Biliary/ 8772
- (primary biliary cirrhosis or PBS).ti,ab. 36509
- exp Cholangitis, Sclerosing/ 4703
- sclerosing cholangiti*.ti,ab. 7289
- exp Lung Diseases/ 1236542
- ((lung or pulmonary) adj (disease* or disorder* or condition*)).ti,ab. 142936
- exp Pulmonary Disease, Chronic Obstructive/ 67441
- ((chronic obstructive adj (pulmonary or lung or airway) adj (disease* or obstruction*)) or (COPD or COAD)).ti,ab. 83179
- exp Asthma/ 143083
- (asthma or asthmatic).ti,ab. 176832
- exp Muscular Diseases/ 196410
- (((muscle or muscular or myopathic) adj (disorder* or disease* or condition*)) or (myopathy or myopathies)).ti,ab. 36194
- exp Muscular Dystrophies/ 30020
- (muscular dystroph* or myodystroph*).ti,ab. 27210
- or/1-56 5558763
- ”Fatigue Questionnaire”.ti,ab. 382
- ”Fatigue Severity Scale”.ti,ab. 1808
- ”Multidimensional Assessment of Fatigue”.ti,ab. 139
- ”Short Form-36 Vitality”.ti,ab. 34
- (”Functional Assessment of Chronic Illness Therapy Fatigue” or FACIT F).ti,ab. 633
- ”Brief Fatigue Inventory”.ti,ab. 472
- ”Numerical Rating Scale Fatigue”.ti,ab. 7
- (”Visual Analog Scale Fatigue” or VAS F).ti,ab. 105
- ”Checklist Individual Strength”.ti,ab. 323
- ”Chalder Fatigue Scale”.ti,ab. 201
- ”Multidimensional Fatigue Inventory Scale”.ti,ab. 8
- ”Piper Fatigue Scale”.ti,ab. 268
- (PROMIS-29 or PROMIS 29 or PROMIS29).ti,ab. 235
- Pittsburgh Fatigability Scale.ti,ab. 32
- Fatigue Descriptive Scale.ti,ab. 10
- Modified Fatigue Impact Scale.ti,ab. 517
- (”40-item Fatigue Impact Scale” or “40 item Fatigue Impact Scale”).ti,ab. 4
- (”29-item Fatigue Assessment Instrument” or “29 item Fatigue Assessment Instrument”).ti,ab. 1
- (”Functional Assessment of Multiple Sclerosis” or FAMS).ti,ab. 172
- or/58-76 4995
- 78 *Fatigue/ 16397
- 79 (fatigue adj7 (scale* or subscale* or sub-scale* or questionnaire* or assessment* or inventor* or measure* or tool*)).ti,ab. 17355
- 80 (scale or subscale or sub-scale or questionnaire or assessment or inventory or measure or measurement).ti,ab. 3456677
- 81 (fatigability or fatigable).ti,ab. 3406
- 78 or 81 19535
- 80 and 82 6761
- 79 or 83 19584
- 77 or 84 20135
- 57 and 85 8442
- exp randomized controlled trial/ 602157
- controlled clinical trial.pt. 95425
- randomized.ab. 618304
- placebo.ab. 241721
- clinical trials as topic/ 201321
- randomly.ab. 417343
- trial.ti. 293560
- or/87-93 1550631
- exp animals/ not humans/ 5158236
- 94 not 95 1427530
- 86 and 96 1938

Embase <1974 to 2023 Week 38>

- *chronic disease/ 32846
- ((chronic or long-term or long term) adj (condition* or disease* or illness*)).ti,ab. 179677
- chronically ill.ti,ab. 7536
- *rheumatic disease/ 32096
- rheumati*.ti,ab. 80760
- *diabetes mellitus/ 242661
- diabet*.ti,ab. 1174089
- *endocrine disease/ 7079
- *thyroid disease/ 13779
- *adrenal disease/ 2041
- *adrenal insufficiency/ 4392
- *autoimmune disease/ 35243
- ((endocrine or thyroid or adrenal or autoimmune or auto-immune or auto immune) adj1 (disorder* or disease* or condition*)).ti,ab. 182768
- adrenal insufficiency.ti,ab. 10682
- *heart failure/ 126614
- heart failure*.ti,ab. 350732
- *coronary artery disease/ 96075
- coronary heart disease*.ti,ab. 75791
- *chronic kidney failure/ 61974
- (chronic adj (renal or kidney) adj (insuffucien* or failure* or disease*)).ti,ab. 155133
- *hemodialysis/ 63019
- dialysis.ti,ab. 181634
- *transplantation/ 64169
- (transplant* adj3 (heart* or kidney* or liver* or lung*)).ti,ab. 304389
- *multiple sclerosis/ 102769
- multiple sclerosis.ti,ab. 141053
- *cerebrovascular accident/ 105016
- stroke.ti,ab. 488091
- *degenerative disease/ 18573
- ((neurodegenerative or neuro-degenerative or neuro degenerative) adj (disease* or disorder* or condition*)).ti,ab. 131356
- *Parkinson disease/ 121773
- (parkinson* adj disease).ti,ab. 168860
- *rheumatoid arthritis/ 127396
- rheumatoid arthritis.ti,ab. 180981
- *osteoarthritis/ 52154
- osteoarthritis.ti,ab. 120269
- *systemic lupus erythematosus/ 63341
- lupus.ti,ab. 126358
- *systemic sclerosis/ 22876
- (systemic sclerosis or scleroderma).ti,ab. 44582
- *inflammatory bowel disease/ 27489
- (inflammatory bowel disease* or IBD).ti,ab. 119395
- *biliary cirrhosis/ 2200
- (primary biliary cirrhosis or PBS).ti,ab. 59132
- *sclerosing cholangitis/ 1978
- sclerosing cholangiti*.ti,ab. 12483
- *lung disease/ 34450
- ((lung or pulmonary) adj (disease* or disorder* or condition*)).ti,ab. 217538
- *chronic obstructive lung disease/ 82916
- ((chronic obstructive adj (pulmonary or lung or airway) adj (disease* or obstruction*)) or (COPD or COAD)).ti,ab. 143936
- *asthma/ 152110
- (asthma or asthmatic).ti,ab. 258285
- *muscle disease/ 9985
- (((muscle or muscular or myopathic) adj (disorder* or disease* or condition*)) or (myopathy or myopathies)).ti,ab. 51568
- *muscular dystrophy/ 9507
- (muscular dystroph* or myodystroph*).ti,ab. 36327
- or/1-56 4496914
- ”Fatigue Questionnaire”.ti,ab. 625
- ”Fatigue Severity Scale”.ti,ab. 3460
- ”Multidimensional Assessment of Fatigue”.ti,ab. 269
- ”Short Form-36 Vitality”.ti,ab. 39
- (”Functional Assessment of Chronic Illness Therapy Fatigue” or FACIT F).ti,ab. 1676
- ”Brief Fatigue Inventory”.ti,ab. 881
- ”Numerical Rating Scale Fatigue”.ti,ab. 11
- (”Visual Analog Scale Fatigue” or VAS F).ti,ab. 163
- ”Checklist Individual Strength”.ti,ab. 452
- ”Chalder Fatigue Scale”.ti,ab. 310
- ”Multidimensional Fatigue Inventory Scale”.ti,ab. 14
- ”Piper Fatigue Scale”.ti,ab. 376
- (PROMIS-29 or PROMIS 29 or PROMIS29).ti,ab. 617
- Pittsburgh Fatigability Scale.ti,ab. 44
- Fatigue Descriptive Scale.ti,ab. 17
- Modified Fatigue Impact Scale.ti,ab. 1077
- (”40-item Fatigue Impact Scale” or “40 item Fatigue Impact Scale”).ti,ab. 4
- (”29-item Fatigue Assessment Instrument” or “29 item Fatigue Assessment Instrument”).ti,ab. 2
- (”Functional Assessment of Multiple Sclerosis” or FAMS).ti,ab. 355
- exp Fatigue Severity Scale/ or exp “Functional Assessment of Chronic Illness Therapy Fatigue Scale”/ or exp Multidimensional Fatigue Inventory/ or exp Chalder Fatigue Scale/ or exp Piper fatigue scale/ or exp “fatigue scale for motor and cognitive functions”/ or exp Fatigue Impact Scale/ 6541
- or/58-77 12125
- 79 *fatigue/ 25859
- (fatigue adj7 (scale* or subscale* or sub-scale* or questionnaire* or assessment* or inventor* or measure* or tool*)).ti,ab. 29052
- (scale or subscale or sub-scale or questionnaire or assessment or inventory or measure or measurement).ti,ab. 4694159
- (fatigability or fatigable).ti,ab. 5144
- 79 or 82 30552
- 81 and 83 12023
- 80 or 84 32495
- 78 or 85 35039
- 57 and 86 12690
- 88 exp randomized controlled trial/ 785235
- 89 controlled clinical trial/ 470992
- 90 random$.ti,ab. 1975440
- 91 randomization/ 98376
- 92 intermethod comparison/ 300743
- 93 placebo.ti,ab. 365413
- 94 (compare or compared or comparison).ti,ab. 7636197
- 95 ((evaluated or evaluate or evaluating or assessed or assess) and (compare or compared or comparing or comparison)).ab. 2778695
- 96 (open adj label).ti,ab. 108778
- 97 ((double or single or doubly or singly) adj (blind or blinded or blindly)).ti,ab. 273962
- 98 double blind procedure/ 210736
- 99 parallel group$1.ti,ab. 32147
- 100 (crossover or cross over).ti,ab. 124632
- 101 ((assign$ or match or matched or allocation) adj5 (alternate or group$1 or intervention$1 or patient$1 or subject$1 or participant$1)).ti,ab. 415356
- 102 (assigned or allocated).ti,ab. 490792
- 103 (controlled adj7 (study or design or trial)).ti,ab. 450700
- 104 (volunteer or volunteers).ti,ab. 282605
- 105 human experiment/ 642664
- 106 trial.ti. 401617
- 107 or/88-106 10036592
- 108 (random$ adj sampl$ adj7 (”cross section$” or questionnaire$1 or survey$ or database$1)).ti,ab. not (comparative study/ or controlled study/ or randomi?ed controlled.ti,ab. or randomly assigned.ti,ab.) 9609
- 109 cross-sectional study/ not (exp randomized controlled trial/ or controlled clinical trial/ or controlled study/ or randomi?ed controlled.ti,ab. or control group$1.ti,ab.) 361562search
- 110 (((case adj control$) and random$) not randomi?ed controlled).ti,ab. 21555
- 111 systematic review.ti,ab. not (trial or study).ti. 325581
- 112 (nonrandom$ not random$).ti,ab. 18945
- 113 “random field$”.ti,ab. 2966
- 114 (random cluster adj3 sampl$).ti,ab. 1583
- 115 (review.ab. and review.pt.) not trial.ti. 1131497
- 116 “we searched”.ab. and (review.ti. or review.pt.) 49364
- 117 “update review”.ab. 136
- 118 (databases adj4 searched).ab. 62550
- 119 (rat or rats or mouse or mice or swine or porcine or murine or sheep or lambs or pigs or piglets or rabbit or rabbits or cat or cats or dog or dogs or cattle or bovine or monkey or monkeys or trout or marmoset$1).ti. and animal experiment/ 1220906
- 120 animal experiment/ not (human experiment/ or human/) 2564767
- 121 or/108-120 4382762
- 122 107 not 121 8767282
- 123 87 and 122 6538
- 124 limit 123 to “remove medline records” 4030

CINAHL via EBSCO

Monday, October 02, 2023 4:18:56 PM S1 (MH “Chronic Disease+”)

S2 TI ((chronic or long-term or long term) N1 (condition* or disease* or illness*)) OR AB ((chronic or long-term or long term) N1 (condition* or disease* or illness*))

S3 TI chronically ill OR AB chronically ill S4 (MH “Rheumatic Diseases+”)

S5 TI rheumati* OR AB rheumati* S6 (MH “Diabetes Mellitus+”)

S7 TI diabet* OR AB diabet*

S8 (MH “Endocrine Diseases+”) S9 (MH “Thyroid Diseases+”)

S10 (MH “Adrenal Gland Diseases+”) S11 (MH “Adrenal Insufficiency+”) S12 (MH “Autoimmune Diseases+”)

S13 TI ((endocrine or thyroid or adrenal or autoimmune or auto-immune or auto immune) N1 (disorder* or disease* or condition*)) OR AB ((endocrine or thyroid or adrenal or autoimmune or auto-immune or auto immune) N1 (disorder* or disease* or condition*))

S14 TI adrenal insufficiency OR AB adrenal insufficiency S15 (MH “Heart Failure+”)

S16 TI heart failure* OR AB heart failure* S17 (MH “Coronary Disease+”)

S18 TI coronary heart disease* OR AB coronary heart disease* S19 (MH “Renal Insufficiency, Chronic+”)

S20 (MH “Kidney Failure, Chronic+”)

S21 TI (chronic adj (renal or kidney) N1 (insuffucien* or failure* or disease*)) OR AB (chronic adj (renal or kidney) N1 (insuffucien* or failure* or disease*))

S22 (MH “Dialysis Patients”) S23 TI dialysis OR AB dialysis

S24 TI (transplant* N3 (heart* or kidney* or liver* or lung*)) OR AB (transplant* N3 (heart* or kidney* or liver* or lung*))

S25 (MH “Multiple Sclerosis+”)

S26 TI multiple sclerosis OR AB multiple sclerosis S27 (MH “Stroke+”)

S28 TI stroke OR AB stroke

S29 (MH “Neurodegenerative Diseases+”)

S30 TI ((neurodegenerative or neuro-degenerative or neuro degenerative) N1 (disease* or disorder* or condition*)) OR AB ((neurodegenerative or neuro-degenerative or neuro degenerative) N1 (disease* or disorder* or condition*))

S31 (MH “Parkinson Disease”)

S32 TI (parkinson* N1 disease) OR AB (parkinson* N1 disease) S33 (MH “Arthritis, Rheumatoid+”)

S34 TI rheumatoid arthritis OR AB rheumatoid arthritis S35 (MH “Osteoarthritis+”)

S36 TI osteoarthritis OR AB osteoarthritis S37 (MH “Lupus Erythematosus, Systemic+”)

S38 TI lupus OR AB lupus

S39 (MH “Scleroderma, Systemic+”)

S40 TI (systemic sclerosis or scleroderma) OR AB (systemic sclerosis or scleroderma) S41 (MH “Inflammatory Bowel Diseases+”)

S42 TI (inflammatory bowel disease* or IBD) OR AB (inflammatory bowel disease* or IBD) S43 (MH “Liver Cirrhosis+”)

S44 TI (primary biliary cirrhosis or PBS) OR AB (primary biliary cirrhosis or PBS) S45 (MH “Cholangitis, Sclerosing”)

S46 TI sclerosing cholangiti* OR AB sclerosing cholangiti* S47 (MH “Lung Diseases+”)

S48 TI ((lung or pulmonary) N1 (disease* or disorder* or condition*)) OR AB ((lung or pulmonary) N1 (disease* or disorder* or condition*))

S49 (MH “Pulmonary Disease, Chronic Obstructive+”)

S50 TI ((chronic obstructive N1 (pulmonary or lung or airway) N1 (disease* or obstruction*)) or (COPD or COAD)) OR AB ((chronic obstructive N1 (pulmonary or lung or airway) N1 (disease* or obstruction*)) or (COPD or COAD))

S51 (MH “Asthma+”)

S52 TI (asthma or asthmatic) OR AB (asthma or asthmatic) S53 (MH “Muscular Diseases+”)

S54 TI (((muscle or muscular or myopathic) ADJ1 (disorder* or disease* or condition*)) or (myopathy or myopathies)) OR AB (((muscle or muscular or myopathic) ADJ1 (disorder* or disease* or condition*)) or (myopathy or myopathies))

S55 (MH “Muscular Dystrophy+”)

S56 TI (muscular dystroph* or myodystroph*) OR AB (muscular dystroph* or myodystroph*)

S57 S1 OR S2 OR S3 OR S4 OR S5 OR S6 OR S7 OR S8 OR S9 OR S10 OR S11 OR S12 OR S13 OR S14 OR S15 OR S16 OR S17 OR S18 OR S19 OR S20 OR S21 OR S22 OR S23 OR S24 OR S25 OR S26 OR S27 OR S28 OR S29 OR S30 OR S31 OR S32 OR S33 OR S34 OR S35 OR S36 OR S37 OR S38 OR S39 OR S40 OR S41 OR S42 OR S43 OR S44 OR S45 OR S46 OR S47 OR S48 OR S49 OR S50 OR S51 OR S52 OR S53 OR S54 OR S55 OR S56

S58 TI “Fatigue Questionnaire” OR AB “Fatigue Questionnaire” S59 TI “Fatigue Severity Scale” OR AB “Fatigue Severity Scale”

S60 TI “Multidimensional Assessment of Fatigue” OR AB “Multidimensional Assessment of Fatigue” S61 TI “Short Form-36 Vitality” OR AB “Short Form-36 Vitality”

S62 TI (”Functional Assessment of Chronic Illness Therapy Fatigue” or FACIT F) OR AB (”Functional Assessment of Chronic Illness Therapy Fatigue” or FACIT F)

S63 TI “Brief Fatigue Inventory” OR AB “Brief Fatigue Inventory”

S64 TI “Numerical Rating Scale Fatigue” OR AB “Numerical Rating Scale Fatigue”

S65 TI (”Visual Analog Scale Fatigue” or VAS F) OR AB (”Visual Analog Scale Fatigue” or VAS F) S66 TI “Checklist Individual Strength” OR AB “Checklist Individual Strength”

S67 TI “Chalder Fatigue Scale” OR AB “Chalder Fatigue Scale”

S68 TI “Multidimensional Fatigue Inventory Scale” OR AB “Multidimensional Fatigue Inventory Scale” S69 TI “Piper Fatigue Scale” OR AB “Piper Fatigue Scale”

S70 TI (PROMIS-29 or PROMIS 29 or PROMIS29) OR AB (PROMIS-29 or PROMIS 29 or PROMIS29)

S71 TI Pittsburgh Fatigability Scale OR AB Pittsburgh Fatigability Scale S72 TI Fatigue Descriptive Scale OR AB Fatigue Descriptive Scale

S73 TI Modified Fatigue Impact Scale OR AB Modified Fatigue Impact Scale

S74 TI (”40-item Fatigue Impact Scale” or “40 item Fatigue Impact Scale”) OR AB (”40-item Fatigue Impact Scale” or “40 item Fatigue Impact Scale”)

S75 TI (”29-item Fatigue Assessment Instrument” or “29 item Fatigue Assessment Instrument”) OR AB (”29-item Fatigue Assessment Instrument” or “29 item Fatigue Assessment Instrument”)

S76 TI (”Functional Assessment of Multiple Sclerosis” or FAMS) OR AB (”Functional Assessment of Multiple Sclerosis” or FAMS)

S77 S58 OR S59 OR S60 OR S61 OR S62 OR S63 OR S64 OR S65 OR S66 OR S67 OR S68 OR S69 OR S70 OR S71 OR S72 OR S73 OR S74 OR S75 OR S76

S78 (MM “Fatigue”)

S79 TI (fatigue N7 (scale* or subscale* or sub-scale* or questionnaire* or assessment* or inventor* or measure* or tool*)) OR AB (fatigue N7 (scale* or subscale* or sub-scale* or questionnaire* or assessment* or inventor* or measure* or tool*))

S80 TI (scale or subscale or sub-scale or questionnaire or assessment or inventory or measure or measurement) OR AB (scale or subscale or sub-scale or questionnaire or assessment or inventory or measure or measurement)

S81 TI (fatigability or fatigable) OR AB (fatigability or fatigable) S82 S78 OR S81

S83 S80 AND S82 S84 S79 OR S83 S85 S77 OR S84 S86 S57 AND S85

S87 MH “Clinical Trials+”

S88 PT Clinical trial S89 TX clinic* n1 trial*

S90 TX ((singl* n1 blind*) or (singl* n1 mask*)) or TX ((doubl* n1 blind*) or (doubl* n1 mask*)) or TX ((tripl* n1 blind*) or (tripl* n1 mask*)) or TX ((trebl* n1 blind*) or (trebl* n1 mask*))

S91 TX randomi* control* trial*

S92 MH “Random Assignment” S93 TX random* allocat* S94 TX placebo*

S95 MH “Placebos”

S96 MH “Quantitative Studies” S97 TX allocat* random*

S98 S87 OR S88 OR S89 OR S90 OR S91 OR S92 OR S93 OR S94 OR S95 OR S96 OR S97 S99 S86 AND S98 Results 1905

APA PsycInfo <1806 to September Week 4 2023>

- exp Chronic Illness/ 34600
- ((chronic or long-term or long term) adj (condition* or disease* or illness*)).ti,ab. 29719
- chronically ill.ti,ab. 3184
- exp Rheumatoid Arthritis/ 2100
- rheumati*.ti,ab. 1112
- exp Diabetes Mellitus/ 10460
- diabet*.ti,ab. 36523
- exp Thyroid Disorders/ 1539
- exp Adrenal Gland Disorders/ 422
- exp Immunologic Disorders/ 53042
- ((endocrine or thyroid or adrenal or autoimmune or auto-immune or auto immune) adj1 (disorder* or disease* or condition*)).ti,ab. 3899
- adrenal insufficiency.ti,ab. 148
- exp Heart Disorders/ 16450
- heart failure*.ti,ab. 4536
- exp Cardiovascular Disorders/ 72080
- coronary heart disease*.ti,ab. 4403
- exp Kidney Diseases/ 2674
- (chronic adj (renal or kidney) adj (insuffucien* or failure* or disease*)).ti,ab. 1543
- exp Dialysis/ 2247
- dialysis.ti,ab. 2381
- exp Organ Transplantation/ 5421
- (transplant* adj3 (heart* or kidney* or liver* or lung*)).ti,ab. 2086
- exp Multiple Sclerosis/ 14366
- multiple sclerosis.ti,ab. 17136
- exp Cerebrovascular Accidents/ 24837
- stroke.ti,ab. 37683
- exp Neurodegenerative Diseases/ 95903
- ((neurodegenerative or neuro-degenerative or neuro degenerative) adj (disease* or disorder* or condition*)).ti,ab. 19229
- (parkinson* adj disease).ti,ab. 32763
- exp Rheumatoid Arthritis/ 2100
- rheumatoid arthritis.ti,ab. 2806
- exp Arthritis/ 4810
- osteoarthritis.ti,ab. 2288
- exp Lupus/ 883
- lupus.ti,ab. 1629
- (systemic sclerosis or scleroderma).ti,ab. 219
- exp Colon Disorders/ 5096
- (inflammatory bowel disease* or IBD).ti,ab. 1201
- exp Liver Disorders/ 5073
- (primary biliary cirrhosis or PBS).ti,ab. 1564
- sclerosing cholangiti*.ti,ab. 19
- exp Lung Disorders/ 5469
- ((lung or pulmonary) adj (disease* or disorder* or condition*)).ti,ab. 3973
- exp Chronic Obstructive Pulmonary Disease/ 1788
- ((chronic obstructive adj (pulmonary or lung or airway) adj (disease* or obstruction*)) or (COPD or COAD)).ti,ab. 3014
- exp Asthma/ 5266
- (asthma or asthmatic).ti,ab. 8470
- exp Muscular Disorders/10761
- (((muscle or muscular or myopathic) adj (disorder* or disease* or condition*)) or (myopathy or myopathies)).ti,ab. 1828
- exp Muscular Dystrophy/ 1524
- (muscular dystroph* or myodystroph*).ti,ab. 1629
- or/1-51 380568
- ”Fatigue Questionnaire”.ti,ab. 128
- ”Multidimensional Assessment of Fatigue”.ti,ab. 35
- ”Short Form-36 Vitality”.ti,ab. 7
- (”Functional Assessment of Chronic Illness Therapy Fatigue” or FACIT F).ti,ab. 75
- ”Brief Fatigue Inventory”.ti,ab. 105
- ”Numerical Rating Scale Fatigue”.ti,ab. 4
- (”Visual Analog Scale Fatigue” or VAS F).ti,ab. 21
- ”Checklist Individual Strength”.ti,ab. 110
- ”Chalder Fatigue Scale”.ti,ab. 73
- ”Multidimensional Fatigue Inventory Scale”.ti,ab. 0
- ”Piper Fatigue Scale”.ti,ab. 74
- (PROMIS-29 or PROMIS 29 or PROMIS29).ti,ab. 54
- Pittsburgh Fatigability Scale.ti,ab. 8
- Fatigue Descriptive Scale.ti,ab. 1
- Modified Fatigue Impact Scale.ti,ab. 139
- (”40-item Fatigue Impact Scale” or “40 item Fatigue Impact Scale”).ti,ab. 2
- (”29-item Fatigue Assessment Instrument” or “29 item Fatigue Assessment Instrument”).ti,ab. 1
- (”Functional Assessment of Multiple Sclerosis” or FAMS).ti,ab. 62
- or/52-70 380976
- 72 exp Fatigue/ 11615
- (fatigue adj7 (scale* or subscale* or sub-scale* or questionnaire* or assessment* or inventor* or measure* or tool*)).ti,ab. 5324
- (scale or subscale or sub-scale or questionnaire or assessment or inventory or measure or measurement).ti,ab. 1060391
- (fatigability or fatigable).ti,ab. 536
- 72 or 75 11998
- 74 and 76 4608
- 73 or 77 7340
- 52 and 78 2359
- (double-blind or random: assigned or control).tw. 554265
- 79 and 80 497

Web of Science Core Collection (Science and Social Sciences Citation Indexes – SCI-EXPANDED, SSCI)

TS=(((chronic) NEAR/1 (condition* or disease* or illness*))) — 3,672,386 TS=(((long-term) NEAR/1 (condition* or disease* or illness*))) — 16,292 TS=(chronically ill) — 6,072

TS=(rheumati*) — 54,425

TS=(diabet*) — 879,022

TS=(((endocrine) NEAR/1 (disorder* or disease* or condition*))) — 8,914

TS=(((thyroid) NEAR/1 (disorder* or disease* or condition*))) — 20,043 TS=(((adrenal) NEAR/1 (disorder* or disease* or condition*))) — 1,557 TS=(((autoimmune) NEAR/1 (disorder* or disease* or condition*))) — 106,203 TS=(((auto-immune) NEAR/1 (disorder* or disease* or condition*))) — 1,931 TS=(((auto immune) NEAR/1 (disorder* or disease* or condition*))) — 3,507 TS=adrenal insufficiency — 8,383

TS=heart failure* — 326,475 TS=coronary heart disease* — 192,572

TS=(chronic NEAR/1 (renal or kidney) NEAR/1 (insuffucien* or failure* or disease*)) — 121,026 TS=dialysis — 131,347

TS=(transplant* NEAR/3 (heart* or kidney* or liver* or lung*)) — 257,475 TS=multiple sclerosis — 146,702

TS=stroke — 409,574

TS=((neurodegenerative) NEAR/1 (disease* or disorder* or condition*)) — 106,263 TS=((neuro-degenerative) NEAR/1 (disease* or disorder* or condition*)) — 786 TS=((neuro degenerative) NEAR/1 (disease* or disorder* or condition*)) — 996 TS=(parkinson* NEAR/1 disease) — 176,729

TS=rheumatoid arthritis — 192,358 TS=osteoarthritis — 111,937

TS=lupus — 123,376

TS=(systemic sclerosis or scleroderma) — 45,897 TS=(inflammatory bowel disease* or IBD) — 113,244 TS=(primary biliary cirrhosis or PBS) — 55,103 TS=sclerosing cholangiti* — 11,277

TS=((lung or pulmonary) NEAR/1 (disease* or disorder* or condition*)) — 167,964

TS=((chronic obstructive NEAR/1 (pulmonary or lung or airway) NEAR/1 (disease* or obstruction*)) or (COPD or COAD)) — 104,320

TS=(asthma or asthmatic) — 225,022

TS=(((muscle or muscular or myopathic) NEAR/1 (disorder* or disease* or condition*)) or (myopathy or myopathies)) — 49,529

TS=(muscular dystroph* or myodystroph*) — 38,613

#1 OR #2 OR #3 OR #4 OR #5 OR #6 OR #7 OR #8 OR #9 OR #10 OR #11 OR #12 OR #13 OR #14

OR #15 OR #16 OR #17 OR #18 OR #19 OR #20 OR #21 OR #22 OR #23 OR #24 OR #25 OR #26

OR #27 OR #28 OR #29 OR #30 OR #31 OR #32 OR #33 OR #34 OR #35 – 4,071,294

TS=(”Fatigue Questionnaire”) — 349 TS=(”Fatigue Severity Scale”) — 1,806

TS=”Multidimensional Assessment of Fatigue” — 126 TS=”Short Form-36 Vitality” — 34

TS=(”Functional Assessment of Chronic Illness Therapy Fatigue” or FACIT F) — 598 TS=”Brief Fatigue Inventory” — 438

TS=(”Visual Analog Scale Fatigue” or VAS F) — 1,168 TS=”Checklist Individual Strength” — 318 TS=”Chalder Fatigue Scale” — 189 TS=”Multidimensional Fatigue Inventory Scale” — 6 TS=”Piper Fatigue Scale” — 234

TS=(PROMIS-29 or PROMIS 29 or PROMIS29) — 464

TS=Pittsburgh Fatigability Scale — 35 TS=Fatigue Descriptive Scale — 785 TS=Modified Fatigue Impact Scale — 852

TS=(”40-item Fatigue Impact Scale” or “40 item Fatigue Impact Scale”) — 4

TS=(”29-item Fatigue Assessment Instrument” or “29 item Fatigue Assessment Instrument”) — 1 TS=(”Functional Assessment of Multiple Sclerosis” or FAMS) — 266

TS=(fatigue NEAR/7 (scale* or subscale* or sub-scale* or questionnaire* or assessment* or inventor* or measure* or tool*)) — 26,432

TS=(scale or subscale or sub-scale or questionnaire or assessment or inventory or measure or measurement) — 10,552,202

TS=(fatigue or fatigability or fatigable) — 260,526 #56 AND #57 — 7,113

#55 OR #58 — 93,806

#32 OR #33 OR #34 OR #35 OR #36 OR #37 OR #38 OR #39 OR #40 OR #41 OR #42 OR #43 OR

#44 OR #45 OR #46 OR #47 OR #48 OR #49 OR #50 OR #51 OR #52 OR #53 OR #54-7,673

TI=(randomi?ed controlled trial) — 153,872 #36 AND #60 AND #61 - 756

Cochrane

Date Run: 03/10/2023 15:24:43

ID Search Hits

#1 MeSH descriptor: [Chronic Disease] explode all trees 38848

#2 ((chronic or long-term or long term) NEXT (condition* or disease* or illness*)):ti OR ((chronic or long-term or long term) NEXT (condition* or disease* or illness*)):ab 14796

#3 chronically ill:ti OR chronically ill:ab 553

#4 MeSH descriptor: [Rheumatic Diseases] explode all trees 21037 #5 rheumati*:ti OR rheumati*:ab 4521

#6 MeSH descriptor: [Diabetes Mellitus] explode all trees 46685 #7 diabet*:ti OR diabet*:ab 109423

#8 MeSH descriptor: [Endocrine System Diseases] explode all trees 61968 #9 MeSH descriptor: [Thyroid Diseases] explode all trees 2994

#10 MeSH descriptor: [Adrenal Gland Diseases] explode all trees 755

#11 MeSH descriptor: [Adrenal Insufficiency] explode all trees 327

#12 MeSH descriptor: [Autoimmune Diseases] explode all trees 25146

#13 ((endocrine or thyroid or adrenal or autoimmune or auto-immune or auto immune) NEAR/1 (disorder* or disease* or condition*)):ti OR ((endocrine or thyroid or adrenal or autoimmune or auto-immune or auto immune) NEAR/1 (disorder* or disease* or condition*)):ab 6114

#14 adrenal insufficiency:ti OR adrenal insufficiency:ab 517

#15 MeSH descriptor: [Heart Failure] explode all trees 14623 #16 heart failure*:ti OR heart failure*:ab 38262

#17 MeSH descriptor: [Coronary Disease] explode all trees 18489

#18 coronary heart disease*:ti OR coronary heart disease*:ab 21575 #19 MeSH descriptor: [Renal Insufficiency, Chronic] explode all trees 8653 #20 MeSH descriptor: [Kidney Failure, Chronic] explode all trees 5550

#21 (chronic NEXT (renal or kidney) NEXT (insuffucien* or failure* or disease*)):ti OR (chronic NEXT (renal or kidney) NEXT (insuffucien* or failure* or disease*)):ab 12353

#22 MeSH descriptor: [Renal Dialysis] explode all trees 6578 #23 dialysis:ti OR dialysis:ab13972

#24 MeSH descriptor: [Transplantation] explode all trees 16684

#25 (transplant* NEAR/3 (heart* or kidney* or liver* or lung*)):ti OR (transplant* NEAR/3 (heart* or kidney* or liver* or lung*)):ab 13405

#26 MeSH descriptor: [Multiple Sclerosis] explode all trees 5959 #27 multiple sclerosis:ti OR multiple sclerosis:ab 11695

#28 MeSH descriptor: [Stroke] explode all trees 15152 #29 stroke:ti OR stroke:ab 63149

#30 MeSH descriptor: [Neurodegenerative Diseases] explode all trees 14828

#31 ((neurodegenerative or neuro-degenerative or neuro degenerative) NEXT (disease* or disorder* or condition*)):ti OR ((neurodegenerative or neuro-degenerative or neuro degenerative)

NEXT (disease* or disorder* or condition*)):ab 2697

#32 MeSH descriptor: [Parkinson Disease] explode all trees 6233

#33 (parkinson* NEXT disease):ti OR (parkinson* NEXT disease):ab 11158 #34 MeSH descriptor: [Arthritis, Rheumatoid] explode all trees 7374 #35 rheumatoid arthritis:ti OR rheumatoid arthritis:ab 17496

#36 MeSH descriptor: [Osteoarthritis] explode all trees 10596 #37 osteoarthritis:ti OR osteoarthritis:ab 19163

#38 MeSH descriptor: [Lupus Erythematosus, Systemic] explode all trees 1448

#39 lupus:ti OR lupus:ab 3797

#40 MeSH descriptor: [Scleroderma, Systemic] explode all trees 731

#41 (systemic sclerosis or scleroderma):ti OR (systemic sclerosis or scleroderma):ab 2068 #42 MeSH descriptor: [Inflammatory Bowel Diseases] explode all trees 4872

#43 (inflammatory bowel disease* or IBD):ti OR (inflammatory bowel disease* or IBD):ab 4800

#44 MeSH descriptor: [Liver Cirrhosis, Biliary] explode all trees 368

#45 (primary biliary cirrhosis or PBS):ti OR (primary biliary cirrhosis or PBS):ab 1704

#46 MeSH descriptor: [Cholangitis, Sclerosing] explode all trees 135

#47 sclerosing cholangiti*:ti OR sclerosing cholangiti*:ab 365

#48 MeSH descriptor: [Lung Diseases] explode all trees 58875

#49 ((lung or pulmonary) NEXT (disease* or disorder* or condition*)):ti OR ((lung or pulmonary) NEXT (disease* or disorder* or condition*)):ab 20401

#50 MeSH descriptor: [Pulmonary Disease, Chronic Obstructive] explode all trees 7303

#51 ((chronic obstructive NEXT (pulmonary or lung or airway) NEXT (disease* or obstruction*)) or (COPD or COAD)):ti OR ((chronic obstructive NEXT (pulmonary or lung or airway) NEXT (disease* or obstruction*)) or (COPD or COAD)):ab 22726

#52 MeSH descriptor: [Asthma] explode all trees 15046

#53 (asthma or asthmatic):ti OR (asthma or asthmatic):ab 34442 #54 MeSH descriptor: [Muscular Diseases] explode all trees 11960

#55 (((muscle or muscular or myopathic) NEXT (disorder* or disease* or condition*)) or (myopathy or myopathies)):ti OR (((muscle or muscular or myopathic) NEXT (disorder* or disease* or condition*)) or (myopathy or myopathies)):ab 1361

#56 MeSH descriptor: [Muscular Dystrophies] explode all trees 595

#57 (muscular dystroph* or myodystroph*):ti OR (muscular dystroph* or myodystroph*):ab 1101

#58 {OR #1-#57} 494013

#59 “Fatigue Questionnaire”:ti OR “Fatigue Questionnaire”:ab 240

#60 “Fatigue Severity Scale”:ti OR “Fatigue Severity Scale”:ab 1152

#61 “Multidimensional Assessment of Fatigue”:ti OR “Multidimensional Assessment of Fatigue”:ab 56

#62 “Short Form-36 Vitality”:ti OR “Short Form-36 Vitality”:ab 8

#63 (”Functional Assessment of Chronic Illness Therapy Fatigue” or FACIT F):ti OR (”Functional Assessment of Chronic Illness Therapy Fatigue” or FACIT F):ab 954

#64 “Brief Fatigue Inventory”:ti OR “Brief Fatigue Inventory”:ab 412

#65 “Numerical Rating Scale Fatigue”:ti OR “Numerical Rating Scale Fatigue”:ab 4

#66 (”Visual Analog Scale Fatigue” or VAS F):ti OR (”Visual Analog Scale Fatigue” or VAS F):ab 2248

#67 “Checklist Individual Strength”:ti OR “Checklist Individual Strength”:ab 167

#68 “Chalder Fatigue Scale”:ti OR “Chalder Fatigue Scale”:ab 190

#69 “Multidimensional Fatigue Inventory Scale”:ti OR “Multidimensional Fatigue Inventory Scale”:ab 3

#70 “Piper Fatigue Scale”:ti OR “Piper Fatigue Scale”:ab 205

#71 (PROMIS-29 or PROMIS 29 or PROMIS29):ti OR (PROMIS-29 or PROMIS 29 or PROMIS29):ab 258

#72 Pittsburgh Fatigability Scale:ti OR Pittsburgh Fatigability Scale:ab 10

#73 Fatigue Descriptive Scale:ti OR Fatigue Descriptive Scale:ab 493

#74 Modified Fatigue Impact Scale:ti OR Modified Fatigue Impact Scale:ab 700

#75 (”40-item Fatigue Impact Scale” or “40 item Fatigue Impact Scale”):ti OR (”40-item Fatigue Impact Scale” or “40 item Fatigue Impact Scale”):ab 1

#76 (”29-item Fatigue Assessment Instrument” or “29 item Fatigue Assessment Instrument”):ti OR (”29-item Fatigue Assessment Instrument” or “29 item Fatigue Assessment Instrument”):ab 1

#77 (”Functional Assessment of Multiple Sclerosis” or FAMS):ti OR (”Functional Assessment of Multiple Sclerosis” or FAMS):ab 50

#78 {OR #59-#77} 6577

#79 MeSH descriptor: [Fatigue] this term only 8377

#80 (fatigue NEXT/7 (scale* or subscale* or sub-scale* or questionnaire* or assessment* or inventor* or measure* or tool*)):ti OR (fatigue NEXT/7 (scale* or subscale* or sub-scale* or questionnaire* or assessment* or inventor* or measure* or tool*)):ab 7123

#81 (scale or subscale or sub-scale or questionnaire or assessment or inventory or measure or measurement):ti OR (scale or subscale or sub-scale or questionnaire or assessment or inventory or measure or measurement):ab 512278

#82 (fatigability or fatigable):ti OR (fatigability or fatigable):ab 393 #83 #79 OR #82 8708

#84 #81 AND #83 3579

#85 #80 OR #84 9134

#86 #58 AND #85 3981

Search Strategies: Systematic Reviews search

Ovid MEDLINE(R) Epub Ahead of Print and In-Process, In-Data-Review & Other Non-Indexed Citations and Daily <November 27, 2024>

- exp Chronic Disease/ 654158
- ((chronic or long-term or long term) adj (condition* or disease* or illness*)).ti,ab. 141815
- chronically ill.ti,ab. 6323
- exp Rheumatic Diseases/ 272346
- rheumati*.ti,ab. 66215
- exp Diabetes Mellitus/ 538090
- diabet*.ti,ab. 834172
- exp Endocrine System Diseases/ 1181269
- exp Thyroid Diseases/ 168992
- exp Adrenal Gland Diseases/ or exp Adrenal Insufficiency/ 74467
- exp Autoimmune Diseases/ 570667
- ((endocrine or thyroid or adrenal or autoimmune or auto-immune or auto immune) adj1 (disorder* or disease* or condition*)).ti,ab. 132811
- adrenal insufficiency.ti,ab. 7868
- exp Heart Failure/ 157212
- heart failure*.ti,ab. 226915
- exp Coronary Disease/ 242628
- coronary heart disease*.ti,ab. 57626
- exp Renal Insufficiency, Chronic/ 141810
- exp Kidney Failure, Chronic/ 103657
- (chronic adj (renal or kidney) adj (insuffucien* or failure* or disease*)).ti,ab. 107957
- exp Renal Dialysis/ 130524
- dialysis.ti,ab. 127861
- exp Transplants/ 33321
- (transplant* adj3 (heart* or kidney* or liver* or lung*)).ti,ab. 191883
- exp Multiple Sclerosis/ 73877
- multiple sclerosis.ti,ab. 96133
- exp Stroke/ 186713
- stroke.ti,ab. 330970
- exp Neurodegenerative Diseases/ 394228
- ((neurodegenerative or neuro-degenerative or neuro degenerative) adj (disease* or disorder* or condition*)).ti,ab. 111431
- exp Parkinson Disease/ 88202
- (parkinson* adj disease).ti,ab. 126026
- exp Arthritis, Rheumatoid/ 130626
- rheumatoid arthritis.ti,ab. 126907
- exp Osteoarthritis/ 82888
- osteoarthritis.ti,ab. 92680
- exp Lupus Erythematosus, Systemic/ 69932
- lupus.ti,ab. 93222
- exp Scleroderma, Systemic/ 24020
- (systemic sclerosis or scleroderma).ti,ab. 30500
- exp Inflammatory Bowel Diseases/ 103655
- (inflammatory bowel disease* or IBD).ti,ab. 73818
- exp Liver Cirrhosis, Biliary/ 9010
- (primary biliary cirrhosis or PBS).ti,ab. 38807
- exp Cholangitis, Sclerosing/ 4920
- sclerosing cholangiti*.ti,ab. 7766
- exp Lung Diseases/ 1312727
- ((lung or pulmonary) adj (disease* or disorder* or condition*)).ti,ab. 154066
- exp Pulmonary Disease, Chronic Obstructive/ 70946
- ((chronic obstructive adj (pulmonary or lung or airway) adj (disease* or obstruction*)) or (COPD or COAD)).ti,ab. 89722
- exp Asthma/ 147301
- (asthma or asthmatic).ti,ab. 184955
- exp Muscular Diseases/ 203180
- (((muscle or muscular or myopathic) adj (disorder* or disease* or condition*)) or (myopathy or myopathies)).ti,ab. 38290
- exp Muscular Dystrophies/ 30943
- (muscular dystroph* or myodystroph*).ti,ab. 28443
- or/1-56 5874536
- ”Fatigue Questionnaire”.ti,ab. 421
- ”Fatigue Severity Scale”.ti,ab. 2067
- ”Multidimensional Assessment of Fatigue”.ti,ab. 144
- ”Short Form-36 Vitality”.ti,ab. 34
- (”Functional Assessment of Chronic Illness Therapy Fatigue” or FACIT F).ti,ab. 752
- ”Brief Fatigue Inventory”.ti,ab. 517
- ”Numerical Rating Scale Fatigue”.ti,ab. 7
- (”Visual Analog Scale Fatigue” or VAS F).ti,ab. 120
- ”Checklist Individual Strength”.ti,ab. 352
- ”Chalder Fatigue Scale”.ti,ab. 240
- ”Multidimensional Fatigue Inventory Scale”.ti,ab. 10
- ”Piper Fatigue Scale”.ti,ab. 289
- (PROMIS-29 or PROMIS 29 or PROMIS29).ti,ab. 335
- Pittsburgh Fatigability Scale.ti,ab. 43
- Fatigue Descriptive Scale.ti,ab. 11
- Modified Fatigue Impact Scale.ti,ab. 606
- (”40-item Fatigue Impact Scale” or “40 item Fatigue Impact Scale”).ti,ab. 4
- (”29-item Fatigue Assessment Instrument” or “29 item Fatigue Assessment Instrument”).ti,ab. 1
- (”Functional Assessment of Multiple Sclerosis” or FAMS).ti,ab. 193
- or/58-76 5751
- *Fatigue/ 17037
- (fatigue adj7 (scale* or subscale* or sub-scale* or questionnaire* or assessment* or inventor* or measure* or tool*)).ti,ab. 19310
- (scale or subscale or sub-scale or questionnaire or assessment or inventory or measure or measurement).ti,ab. 3753921
- (fatigability or fatigable).ti,ab. 3630
- 78 or 81 20385
- 80 and 82 7203
- 79 or 83 21678
- 77 or 84 22367
- 57 and 85 9455
- (MEDLINE or systematic review).tw. or meta analysis.pt. 500343
- 86 and 87 325

Embase <1974 to 2024 Week 47>

1 *chronic disease/ 34954

2 ((chronic or long-term or long term) adj (condition* or disease* or illness*)).ti,ab. 196339

3 chronically ill.ti,ab. 7801

4 *rheumatic disease/ 33333

5 rheumati*.ti,ab. 86414

6 *diabetes mellitus/ 255008

7 diabet*.ti,ab. 1266395

8 *endocrine disease/ 7508

9 *thyroid disease/ 14564

10 *adrenal disease/ 2112

11 *adrenal insufficiency/ 4871

12 *autoimmune disease/ 37088

13 ((endocrine or thyroid or adrenal or autoimmune or auto-immune or auto immune) adj1 (disorder* or disease* or condition*)).ti,ab. 199767

14 adrenal insufficiency.ti,ab. 12291

15 *heart failure/ 136001

16 heart failure*.ti,ab. 381715

17 *coronary artery disease/ 100373

18 coronary heart disease*.ti,ab. 79147

19 *chronic kidney failure/ 69844

20 (chronic adj (renal or kidney) adj (insuffucien* or failure* or disease*)).ti,ab. 172600

21 *hemodialysis/ 67261

22 dialysis.ti,ab. 193744

23 *transplantation/ 64725

24 (transplant* adj3 (heart* or kidney* or liver* or lung*)).ti,ab. 329022

25 *multiple sclerosis/ 108475

26 multiple sclerosis.ti,ab. 149971

27 *cerebrovascular accident/ 115826

28 stroke.ti,ab. 529463

29 *degenerative disease/ 20667

30 ((neurodegenerative or neuro-degenerative or neuro degenerative) adj (disease* or disorder* or condition*)).ti,ab. 145110

31 *Parkinson disease/ 129695

32 (parkinson* adj disease).ti,ab. 181036

33 *rheumatoid arthritis/ 133978

34 rheumatoid arthritis.ti,ab. 191238

35 *osteoarthritis/ 55895

36 osteoarthritis.ti,ab. 131490

37 *systemic lupus erythematosus/ 67671

38 lupus.ti,ab. 135191

39 *systemic sclerosis/ 24539

40 (systemic sclerosis or scleroderma).ti,ab. 47556

41 *inflammatory bowel disease/ 33488

42 (inflammatory bowel disease* or IBD).ti,ab. 131258

43 *biliary cirrhosis/ 2407

44 (primary biliary cirrhosis or PBS).ti,ab. 62982

45 *sclerosing cholangitis/ 2054

46 sclerosing cholangiti*.ti,ab. 13366

47 *lung disease/ 35911

48 ((lung or pulmonary) adj (disease* or disorder* or condition*)).ti,ab. 237743

49 *chronic obstructive lung disease/ 88575

50 ((chronic obstructive adj (pulmonary or lung or airway) adj (disease* or obstruction*)) or (COPD or COAD)).ti,ab. 155701

51 *asthma/ 157738

52 (asthma or asthmatic).ti,ab. 271814

53 *muscle disease/ 10264

54 (((muscle or muscular or myopathic) adj (disorder* or disease* or condition*)) or (myopathy or myopathies)).ti,ab. 55279

55 *muscular dystrophy/ 9781

56 (muscular dystroph* or myodystroph*).ti,ab. 38669

57 or/1-56 4825307

58 “Fatigue Questionnaire”.ti,ab. 685

59 “Fatigue Severity Scale”.ti,ab. 3809

60 “Multidimensional Assessment of Fatigue”.ti,ab. 278

61 “Short Form-36 Vitality”.ti,ab. 41

62 (”Functional Assessment of Chronic Illness Therapy Fatigue” or FACIT F).ti,ab. 1940

63 “Brief Fatigue Inventory”.ti,ab. 951

64 “Numerical Rating Scale Fatigue”.ti,ab. 11

65 (”Visual Analog Scale Fatigue” or VAS F).ti,ab. 181

66 “Checklist Individual Strength”.ti,ab. 500

67 “Chalder Fatigue Scale”.ti,ab. 365

68 “Multidimensional Fatigue Inventory Scale”.ti,ab. 18

69 “Piper Fatigue Scale”.ti,ab. 401

70 (PROMIS-29 or PROMIS 29 or PROMIS29).ti,ab. 804

71 Pittsburgh Fatigability Scale.ti,ab. 56

72 Fatigue Descriptive Scale.ti,ab. 18

73 Modified Fatigue Impact Scale.ti,ab. 1198

74 (”40-item Fatigue Impact Scale” or “40 item Fatigue Impact Scale”).ti,ab. 4

75 (”29-item Fatigue Assessment Instrument” or “29 item Fatigue Assessment Instrument”).ti,ab. 2

76 (”Functional Assessment of Multiple Sclerosis” or FAMS).ti,ab. 374

77 exp Fatigue Severity Scale/ or exp “Functional Assessment of Chronic Illness Therapy Fatigue Scale”/ or exp Multidimensional Fatigue Inventory/ or exp Chalder Fatigue Scale/ or exp Piper fatigue scale/ or exp “fatigue scale for motor and cognitive functions”/ or exp Fatigue Impact Scale/ 7913

78 or/58-77 13999

79 *fatigue/ 27870

80 (fatigue adj7 (scale* or subscale* or sub-scale* or questionnaire* or assessment* or inventor* or measure* or tool*)).ti,ab. 31993

81 (scale or subscale or sub-scale or questionnaire or assessment or inventory or measure or measurement).ti,ab. 5085498

82 (fatigability or fatigable).ti,ab. 5536

83 83 79 or 82 32922

84 81 and 83 13199

85 80 or 84 35785

86 78 or 85 38984

87 57 and 86 14155

88 exp review/ 3355072

89 (literature adj3 review$).ti,ab. 494495

90 exp meta analysis/ 338595

91 exp “Systematic Review”/ 496268

92 88 or 89 or 90 or 91 3753315

93 (medline or medlars or embase or pubmed or cinahl or amed or psychlit or psyclit or psychinfo or psycinfo or scisearch or cochrane).ti,ab. 523890

94 RETRACTED ARTICLE/ 14987

95 93 or 94 538405

96 92 and 95 424367

97 (systematic$ adj2 (review$ or overview)).ti,ab. 458236

98 (meta?anal$ or meta anal$ or meta-anal$ or metaanal$ or metanal$).ti,ab. 411264

99 96 or 97 or 98 771746

100 87 and 99 519

101 limit 100 to “remove medline records” 247

CINAHL via EBSCO

S1 (MH “Chronic Disease+”)

S2 TI ((chronic or long-term or long term) N1 (condition* or disease* or illness*)) OR AB ((chronic or long-term or long term) N1 (condition* or disease* or illness*))

S3 TI chronically ill OR AB chronically ill S4 (MH “Rheumatic Diseases+”)

S5 TI rheumati* OR AB rheumati* S6 (MH “Diabetes Mellitus+”)

S7 TI diabet* OR AB diabet*

S8 (MH “Endocrine Diseases+”) S9 (MH “Thyroid Diseases+”)

S10 (MH “Adrenal Gland Diseases+”) S11 (MH “Adrenal Insufficiency+”) S12 (MH “Autoimmune Diseases+”)

S13 TI ((endocrine or thyroid or adrenal or autoimmune or auto-immune or auto immune) N1 (disorder* or disease* or condition*)) OR AB ((endocrine or thyroid or adrenal or autoimmune or auto-immune or auto immune) N1 (disorder* or disease* or condition*))

S14 TI adrenal insufficiency OR AB adrenal insufficiency S15 (MH “Heart Failure+”)

S16 TI heart failure* OR AB heart failure* S17 (MH “Coronary Disease+”)

S18 TI coronary heart disease* OR AB coronary heart disease*

S19 (MH “Renal Insufficiency, Chronic+”) S20 (MH “Kidney Failure, Chronic+”)

S21 TI (chronic adj (renal or kidney) N1 (insuffucien* or failure* or disease*)) OR AB (chronic adj (renal or kidney) N1 (insuffucien* or failure* or disease*))

S22 (MH “Dialysis Patients”) S23 TI dialysis OR AB dialysis

S24 TI (transplant* N3 (heart* or kidney* or liver* or lung*)) OR AB (transplant* N3 (heart* or kidney* or liver* or lung*))

S25 (MH “Multiple Sclerosis+”)

S26 TI multiple sclerosis OR AB multiple sclerosis S27 (MH “Stroke+”)

S28 TI stroke OR AB stroke

S29 (MH “Neurodegenerative Diseases+”)

S30 TI ((neurodegenerative or neuro-degenerative or neuro degenerative) N1 (disease* or disorder* or condition*)) OR AB ((neurodegenerative or neuro-degenerative or neuro degenerative) N1 (disease* or disorder* or condition*))

S31 (MH “Parkinson Disease”)

S32 TI (parkinson* N1 disease) OR AB (parkinson* N1 disease) S33 (MH “Arthritis, Rheumatoid+”)

S34 TI rheumatoid arthritis OR AB rheumatoid arthritis S35 (MH “Osteoarthritis+”)

S36 TI osteoarthritis OR AB osteoarthritis S37 (MH “Lupus Erythematosus, Systemic+”) S38 TI lupus OR AB lupus

S39 (MH “Scleroderma, Systemic+”)

S40 TI (systemic sclerosis or scleroderma) OR AB (systemic sclerosis or scleroderma) S41 (MH “Inflammatory Bowel Diseases+”)

S42 TI (inflammatory bowel disease* or IBD) OR AB (inflammatory bowel disease* or IBD) S43 (MH “Liver Cirrhosis+”)

S44 TI (primary biliary cirrhosis or PBS) OR AB (primary biliary cirrhosis or PBS) S45 (MH “Cholangitis, Sclerosing”)

S46 TI sclerosing cholangiti* OR AB sclerosing cholangiti* S47 (MH “Lung Diseases+”)

S48 TI ((lung or pulmonary) N1 (disease* or disorder* or condition*)) OR AB ((lung or pulmonary) N1 (disease* or disorder* or condition*))

S49 (MH “Pulmonary Disease, Chronic Obstructive+”)

S50 TI ((chronic obstructive N1 (pulmonary or lung or airway) N1 (disease* or obstruction*)) or (COPD or COAD)) OR AB ((chronic obstructive N1 (pulmonary or lung or airway) N1 (disease* or obstruction*)) or (COPD or COAD))

S51 (MH “Asthma+”)

S52 TI (asthma or asthmatic) OR AB (asthma or asthmatic) S53 (MH “Muscular Diseases+”)

S54 TI (((muscle or muscular or myopathic) ADJ1 (disorder* or disease* or condition*)) or (myopathy or myopathies)) OR AB (((muscle or muscular or myopathic) ADJ1 (disorder* or disease* or condition*)) or (myopathy or myopathies))

S55 (MH “Muscular Dystrophy+”)

S56 TI (muscular dystroph* or myodystroph*) OR AB (muscular dystroph* or myodystroph*)

S57 S1 OR S2 OR S3 OR S4 OR S5 OR S6 OR S7 OR S8 OR S9 OR S10 OR S11 OR S12 OR S13 OR S14 OR S15 OR S16 OR S17 OR S18 OR S19 OR S20 OR S21 OR S22 OR S23 OR S24 OR S25 OR S26 OR S27 OR S28 OR S29 OR S30 OR S31 OR S32 OR S33 OR S34 OR S35 OR S36 OR S37 OR S38 OR S39 OR S40 OR S41 OR S42 OR S43 OR S44 OR S45 OR S46 OR S47 OR S48 OR S49 OR S50 OR S51 OR S52 OR S53 OR S54 OR S55 OR S56

S58 TI “Fatigue Questionnaire” OR AB “Fatigue Questionnaire” S59 TI “Fatigue Severity Scale” OR AB “Fatigue Severity Scale”

S60 TI “Multidimensional Assessment of Fatigue” OR AB “Multidimensional Assessment of Fatigue” S61 TI “Short Form-36 Vitality” OR AB “Short Form-36 Vitality”

S62 TI (”Functional Assessment of Chronic Illness Therapy Fatigue” or FACIT F) OR AB (”Functional Assessment of Chronic Illness Therapy Fatigue” or FACIT F)

S63 TI “Brief Fatigue Inventory” OR AB “Brief Fatigue Inventory”

S64 TI “Numerical Rating Scale Fatigue” OR AB “Numerical Rating Scale Fatigue”

S65 TI (”Visual Analog Scale Fatigue” or VAS F) OR AB (”Visual Analog Scale Fatigue” or VAS F) S66 TI “Checklist Individual Strength” OR AB “Checklist Individual Strength”

S67 TI “Chalder Fatigue Scale” OR AB “Chalder Fatigue Scale”

S68 TI “Multidimensional Fatigue Inventory Scale” OR AB “Multidimensional Fatigue Inventory Scale” S69 TI “Piper Fatigue Scale” OR AB “Piper Fatigue Scale”

S70 TI (PROMIS-29 or PROMIS 29 or PROMIS29) OR AB (PROMIS-29 or PROMIS 29 or PROMIS29)

S71 TI Pittsburgh Fatigability Scale OR AB Pittsburgh Fatigability Scale S72 TI Fatigue Descriptive Scale OR AB Fatigue Descriptive Scale

S73 TI Modified Fatigue Impact Scale OR AB Modified Fatigue Impact Scale

S74 TI (”40-item Fatigue Impact Scale” or “40 item Fatigue Impact Scale”) OR AB (”40-item Fatigue Impact Scale” or “40 item Fatigue Impact Scale”)

S75 TI (”29-item Fatigue Assessment Instrument” or “29 item Fatigue Assessment Instrument”) OR AB (”29-item Fatigue Assessment Instrument” or “29 item Fatigue Assessment Instrument”)

S76 TI (”Functional Assessment of Multiple Sclerosis” or FAMS) OR AB (”Functional Assessment of Multiple Sclerosis” or FAMS)

S77 S58 OR S59 OR S60 OR S61 OR S62 OR S63 OR S64 OR S65 OR S66 OR S67 OR S68 OR S69 OR S70 OR S71 OR S72 OR S73 OR S74 OR S75 OR S76

S78 (MM “Fatigue”)

S79 TI (fatigue N7 (scale* or subscale* or sub-scale* or questionnaire* or assessment* or inventor* or measure* or tool*)) OR AB (fatigue N7 (scale* or subscale* or sub-scale* or questionnaire* or assessment* or inventor* or measure* or tool*))

S80 TI (scale or subscale or sub-scale or questionnaire or assessment or inventory or measure or measurement) OR AB (scale or subscale or sub-scale or questionnaire or assessment or inventory or measure or measurement)

S81 TI (fatigability or fatigable) OR AB (fatigability or fatigable) S82 S78 OR S81

S83 S79 AND S80 S84 (S82 OR S83)

S85 (TI (systematic* n3 review*)) or (AB (systematic* n3 review*)) or (TI (systematic* n3 bibliographic*)) or (AB (systematic* n3 bibliographic*)) or (TI (systematic* n3 literature)) or (AB (systematic* n3 literature)) or (TI (comprehensive* n3 literature)) or (AB (comprehensive* n3 literature)) or (TI (comprehensive* n3 bibliographic*)) or (AB (comprehensive* n3 bibliographic*)) or (TI (integrative n3 review)) or (AB (integrative n3 review)) or (JN “Cochrane Database of Systematic Reviews”) or (TI (information n2 synthesis)) or (TI (data n2 synthesis)) or (AB (information n2 synthesis)) or (AB (data n2 synthesis)) or (TI (data n2 extract*)) or (AB (data n2 extract*)) or (TI (medline or pubmed or psyclit or cinahl or (psycinfo not “psycinfo database”) or “web of science” or scopus or embase)) or (AB (medline or pubmed or psyclit or cinahl or (psycinfo not “psycinfo database”) or “web of science” or scopus or embase)) or (MH “Systematic Review”) or (MH “Meta Analysis”) or (TI (meta-analy* or metaanaly*)) or (AB (meta-analy* or metaanaly*)) 319,272

S86 (S84 AND S85) 266

APA PsycInfo <1806 to November 2024 Week 4>

- exp Chronic Illness/ 38747
- ((chronic or long-term or long term) adj (condition* or disease* or illness*)).ti,ab. 31815
- chronically ill.ti,ab. 3266
- exp Rheumatoid Arthritis/ 2191
- rheumati*.ti,ab. 1171
- exp Diabetes Mellitus/ 10907
- diabet*.ti,ab. 38686
- exp Thyroid Disorders/ 1601
- exp Adrenal Gland Disorders/ 434
- exp Immunologic Disorders/ 62844
- ((endocrine or thyroid or adrenal or autoimmune or auto-immune or auto immune) adj1 (disorder* or disease* or condition*)).ti,ab. 4101
- adrenal insufficiency.ti,ab. 152
- exp Heart Disorders/ 17285
- heart failure*.ti,ab. 4886
- exp Cardiovascular Disorders/ 75870
- coronary heart disease*.ti,ab. 4513
- exp Kidney Diseases/ 2855
- (chronic adj (renal or kidney) adj (insuffucien* or failure* or disease*)).ti,ab. 1678
- exp Dialysis/ 2398
- dialysis.ti,ab. 2467
- exp Organ Transplantation/ 5681
- (transplant* adj3 (heart* or kidney* or liver* or lung*)).ti,ab. 2172
- exp Multiple Sclerosis/ 14944
- multiple sclerosis.ti,ab. 17760
- exp Cerebrovascular Accidents/ 26271
- stroke.ti,ab. 39801
- exp Neurodegenerative Diseases/ 102451
- ((neurodegenerative or neuro-degenerative or neuro degenerative) adj (disease* or disorder* or condition*)).ti,ab. 20837
- (parkinson* adj disease).ti,ab. 34494
- exp Rheumatoid Arthritis/ 2191
- rheumatoid arthritis.ti,ab. 2904
- exp Arthritis/ 5077
- osteoarthritis.ti,ab. 2449
- exp Lupus/ 922
- lupus.ti,ab. 1691
- (systemic sclerosis or scleroderma).ti,ab. 232
- exp Colon Disorders/ 6783
- (inflammatory bowel disease* or IBD).ti,ab. 1324
- exp Liver Disorders/ 5368
- (primary biliary cirrhosis or PBS).ti,ab. 1666
- sclerosing cholangiti*.ti,ab. 21
- exp Lung Disorders/ 6793
- ((lung or pulmonary) adj (disease* or disorder* or condition*)).ti,ab. 4200
- exp Chronic Obstructive Pulmonary Disease/ 1930
- ((chronic obstructive adj (pulmonary or lung or airway) adj (disease* or obstruction*)) or (COPD or COAD)).ti,ab. 3223
- exp Asthma/ 5507
- (asthma or asthmatic).ti,ab. 8811
- exp Muscular Disorders/11266
- (((muscle or muscular or myopathic) adj (disorder* or disease* or condition*)) or (myopathy or myopathies)).ti,ab. 1877
- exp Muscular Dystrophy/ 1575
- (muscular dystroph* or myodystroph*).ti,ab. 1679
- or/1-51 403622
- ”Fatigue Questionnaire”.ti,ab. 135
- ”Multidimensional Assessment of Fatigue”.ti,ab. 36
- ”Short Form-36 Vitality”.ti,ab. 8
- (”Functional Assessment of Chronic Illness Therapy Fatigue” or FACIT F).ti,ab. 85
- ”Brief Fatigue Inventory”.ti,ab. 112
- ”Numerical Rating Scale Fatigue”.ti,ab. 4
- (”Visual Analog Scale Fatigue” or VAS F).ti,ab. 26
- ”Checklist Individual Strength”.ti,ab. 121
- ”Chalder Fatigue Scale”.ti,ab. 84
- ”Multidimensional Fatigue Inventory Scale”.ti,ab. 0
- ”Piper Fatigue Scale”.ti,ab. 76
- (PROMIS-29 or PROMIS 29 or PROMIS29).ti,ab. 83
- Pittsburgh Fatigability Scale.ti,ab. 14
- Fatigue Descriptive Scale.ti,ab. 1
- Modified Fatigue Impact Scale.ti,ab. 151
- (”40-item Fatigue Impact Scale” or “40 item Fatigue Impact Scale”).ti,ab. 2
- (”29-item Fatigue Assessment Instrument” or “29 item Fatigue Assessment Instrument”).ti,ab. 1
- (”Functional Assessment of Multiple Sclerosis” or FAMS).ti,ab. 62
- or/52-70 404079
- exp Fatigue/ 12644
- (fatigue adj7 (scale* or subscale* or sub-scale* or questionnaire* or assessment* or inventor* or measure* or tool*)).ti,ab. 5752
- (scale or subscale or sub-scale or questionnaire or assessment or inventory or measure or measurement).ti,ab. 1120968
- (fatigability or fatigable).ti,ab. 558
- 72 or 75 13033
- 74 and 76 5113
- 73 or 77 8002
- 52 and 78 2577
- (meta-analysis or search:).tw. 169257
- 79 and 80 93

Web of Science Core Collection (Science and Social Sciences Citation Indexes – SCI-EXPANDED, SSCI)

TS=(((chronic) NEAR/1 (condition* or disease* or illness*))) Results: 359541 (TS=(((long-term) NEAR/1 (condition* or disease* or illness*)))) Results: 18258 TS=(chronically ill) Results: 6642

TS=(rheumati*) Results: 60176

TS=(diabet*) Results: 950365

TS=(((endocrine) NEAR/1 (disorder* or disease* or condition*))) Results: 10304 TS=(((thyroid) NEAR/1 (disorder* or disease* or condition*))) Results: 22399 TS=(((adrenal) NEAR/1 (disorder* or disease* or condition*))) Results: 1817 TS=(((autoimmune) NEAR/1 (disorder* or disease* or condition*))) Results: 116974 TS=(((auto-immune) NEAR/1 (disorder* or disease* or condition*))) Results: 2123 TS=(((auto immune) NEAR/1 (disorder* or disease* or condition*))) Results: 3843 TS=adrenal insufficiency Results: 9764

TS=heart failure* Results: 358758 TS=coronary heart disease* Results: 204650

TS=(chronic NEAR/1 (renal or kidney) NEAR/1 (insuffucien* or failure* or disease*)) Results: 133763

TS=dialysis Results: 149587

TS=(transplant* NEAR/3 (heart* or kidney* or liver* or lung*)) Results: 275387 TS=multiple sclerosis Results: 156702

TS=stroke Results: 450117

TS=((neurodegenerative) NEAR/1 (disease* or disorder* or condition*)) Results: 117914 TS=((neuro-degenerative) NEAR/1 (disease* or disorder* or condition*)) Results: 826 TS=((neuro degenerative) NEAR/1 (disease* or disorder* or condition*)) Results: 1058 TS=(parkinson* NEAR/1 disease) Results: 190306

TS=rheumatoid arthritis Results: 205223 TS=osteoarthritis Results: 123264

TS=lupus Results: 132734

TS=(systemic sclerosis or scleroderma) Results: 49435 TS=(inflammatory bowel disease* or IBD) Results: 123692 TS=(primary biliary cirrhosis or PBS) Results: 60151 TS=sclerosing cholangiti* Results: 12099

TS=((lung or pulmonary) NEAR/1 (disease* or disorder* or condition*)) Results: 185254 TS=((chronic obstructive NEAR/1 (pulmonary or lung or airway) NEAR/1 (disease* or obstruction*)) or (COPD or COAD)) Results: 112767

TS=(asthma or asthmatic) Results: 239662

TS=(((muscle or muscular or myopathic) NEAR/1 (disorder* or disease* or condition*)) or (myopathy or myopathies)) Results: 54161

TS=(muscular dystroph* or myodystroph*) Results: 41449

#1 OR #2 OR #3 OR #4 OR #5 OR #6 OR #7 OR #8 OR #9 OR #10 OR #11 OR #12 OR #13 OR #14

OR #15 OR #16 OR #17 OR #18 OR #19 OR #20 OR #21 OR #22 OR #23 OR #24 OR #25 OR #26

OR #27 OR #28 OR #29 OR #30 OR #31 OR #32 OR #33 OR #34 OR #35 Results: 3991303

TS=(fatigue or fatigability or fatigable) Results: 288108

TS=(fatigue NEAR/7 (scale* or subscale* or sub-scale* or questionnaire* or assessment* or inventor* or measure* or tool*)) Results: 29495

TS=(scale or subscale or sub-scale or questionnaire or assessment or inventory or measure or measurement) Results: 11727743

#37 AND #39 Results: 103757

#38 OR #40 Results: 104634

#36 AND #41 Results: 17495

TI=(systematic NEAR/3 (review OR overview)) OR TI=(methodologic NEAR/3 (review OR overview)) OR TI=(quantitative NEAR/3 (review OR overview OR synthesis)) OR TI=(research NEAR/3 (integrative OR overview)) OR TI=(integrative NEAR/3 (review OR overview)) OR TI=(collaborative NEAR/3 (review OR overview)) Results: 316800

#42 AND #43 Results: 627

Cochrane Database of Systematic Reviews Issue 11 of 12, November 2024

ID Search

#1 MeSH descriptor: [Chronic Disease] explode all trees

#2 ((chronic or long-term or long term) NEXT (condition* or disease* or illness*)):ti OR ((chronic or long-term or long term) NEXT (condition* or disease* or illness*)):ab

#3 chronically ill:ti OR chronically ill:ab

#4 MeSH descriptor: [Rheumatic Diseases] explode all trees #5 rheumati*:ti OR rheumati*:ab

#6 MeSH descriptor: [Diabetes Mellitus] explode all trees #7 diabet*:ti OR diabet*:ab

#8 MeSH descriptor: [Endocrine System Diseases] explode all trees #9 MeSH descriptor: [Thyroid Diseases] explode all trees

#10 MeSH descriptor: [Adrenal Gland Diseases] explode all trees #11 MeSH descriptor: [Adrenal Insufficiency] explode all trees #12 MeSH descriptor: [Autoimmune Diseases] explode all trees

#13 ((endocrine or thyroid or adrenal or autoimmune or auto-immune or auto immune) NEAR/1 (disorder* or disease* or condition*)):ti OR ((endocrine or thyroid or adrenal or autoimmune or auto-immune or auto immune) NEAR/1 (disorder* or disease* or condition*)):ab

#14 adrenal insufficiency:ti OR adrenal insufficiency:ab #15 MeSH descriptor: [Heart Failure] explode all trees #16 heart failure*:ti OR heart failure*:ab

#17 MeSH descriptor: [Coronary Disease] explode all trees #18 coronary heart disease*:ti OR coronary heart disease*:ab

#19 MeSH descriptor: [Renal Insufficiency, Chronic] explode all trees #20 MeSH descriptor: [Kidney Failure, Chronic] explode all trees

#21 (chronic NEXT (renal or kidney) NEXT (insuffucien* or failure* or disease*)):ti OR (chronic NEXT (renal or kidney) NEXT (insuffucien* or failure* or disease*)):ab

#22 MeSH descriptor: [Renal Dialysis] explode all trees #23 dialysis:ti OR dialysis:ab

#24 MeSH descriptor: [Transplantation] explode all trees

#25 (transplant* NEAR/3 (heart* or kidney* or liver* or lung*)):ti OR (transplant* NEAR/3 (heart* or kidney* or liver* or lung*)):ab

#26 MeSH descriptor: [Multiple Sclerosis] explode all trees #27 multiple sclerosis:ti OR multiple sclerosis:ab

#28 MeSH descriptor: [Stroke] explode all trees #29 stroke:ti OR stroke:ab

#30 MeSH descriptor: [Neurodegenerative Diseases] explode all trees

#31 ((neurodegenerative or neuro-degenerative or neuro degenerative) NEXT (disease* or disorder* or condition*)):ti OR ((neurodegenerative or neuro-degenerative or neuro degenerative)

NEXT (disease* or disorder* or condition*)):ab

#32 MeSH descriptor: [Parkinson Disease] explode all trees

#33 (parkinson* NEXT disease):ti OR (parkinson* NEXT disease):ab #34 MeSH descriptor: [Arthritis, Rheumatoid] explode all trees

#35 rheumatoid arthritis:ti OR rheumatoid arthritis:ab #36 MeSH descriptor: [Osteoarthritis] explode all trees #37 osteoarthritis:ti OR osteoarthritis:ab

#38 MeSH descriptor: [Lupus Erythematosus, Systemic] explode all trees #39 lupus:ti OR lupus:ab

#40 MeSH descriptor: [Scleroderma, Systemic] explode all trees

#41 (systemic sclerosis or scleroderma):ti OR (systemic sclerosis or scleroderma):ab #42 MeSH descriptor: [Inflammatory Bowel Diseases] explode all trees

#43 (inflammatory bowel disease* or IBD):ti OR (inflammatory bowel disease* or IBD):ab #44 MeSH descriptor: [Liver Cirrhosis, Biliary] explode all trees

#45 (primary biliary cirrhosis or PBS):ti OR (primary biliary cirrhosis or PBS):ab #46 MeSH descriptor: [Cholangitis, Sclerosing] explode all trees

#47 sclerosing cholangiti*:ti OR sclerosing cholangiti*:ab #48 MeSH descriptor: [Lung Diseases] explode all trees

#49 ((lung or pulmonary) NEXT (disease* or disorder* or condition*)):ti OR ((lung or pulmonary) NEXT (disease* or disorder* or condition*)):ab

#50 MeSH descriptor: [Pulmonary Disease, Chronic Obstructive] explode all trees

#51 ((chronic obstructive NEXT (pulmonary or lung or airway) NEXT (disease* or obstruction*)) or (COPD or COAD)):ti OR ((chronic obstructive NEXT (pulmonary or lung or airway) NEXT (disease* or obstruction*)) or (COPD or COAD)):ab

#52 MeSH descriptor: [Asthma] explode all trees

#53 (asthma or asthmatic):ti OR (asthma or asthmatic):ab #54 MeSH descriptor: [Muscular Diseases] explode all trees

#55 (((muscle or muscular or myopathic) NEXT (disorder* or disease* or condition*)) or (myopathy or myopathies)):ti OR (((muscle or muscular or myopathic) NEXT (disorder* or disease* or condition*)) or (myopathy or myopathies)):ab

#56 MeSH descriptor: [Muscular Dystrophies] explode all trees

#57 (muscular dystroph* or myodystroph*):ti OR (muscular dystroph* or myodystroph*):ab #58 {OR #1-#57}

#59 “Fatigue Questionnaire”:ti OR “Fatigue Questionnaire”:ab #60 “Fatigue Severity Scale”:ti OR “Fatigue Severity Scale”:ab

#61 “Multidimensional Assessment of Fatigue”:ti OR “Multidimensional Assessment of Fatigue”:ab #62 “Short Form-36 Vitality”:ti OR “Short Form-36 Vitality”:ab

#63 (”Functional Assessment of Chronic Illness Therapy Fatigue” or FACIT F):ti OR (”Functional Assessment of Chronic Illness Therapy Fatigue” or FACIT F):ab

#64 “Brief Fatigue Inventory”:ti OR “Brief Fatigue Inventory”:ab

#65 “Numerical Rating Scale Fatigue”:ti OR “Numerical Rating Scale Fatigue”:ab

#66 (”Visual Analog Scale Fatigue” or VAS F):ti OR (”Visual Analog Scale Fatigue” or VAS F):ab #67 “Checklist Individual Strength”:ti OR “Checklist Individual Strength”:ab

#68 “Chalder Fatigue Scale”:ti OR “Chalder Fatigue Scale”:ab

#69 “Multidimensional Fatigue Inventory Scale”:ti OR “Multidimensional Fatigue Inventory Scale”:ab

#70 “Piper Fatigue Scale”:ti OR “Piper Fatigue Scale”:ab

#71 (PROMIS-29 or PROMIS 29 or PROMIS29):ti OR (PROMIS-29 or PROMIS 29 or PROMIS29):ab

#72 Pittsburgh Fatigability Scale:ti OR Pittsburgh Fatigability Scale:ab #73 Fatigue Descriptive Scale:ti OR Fatigue Descriptive Scale:ab

#74 Modified Fatigue Impact Scale:ti OR Modified Fatigue Impact Scale:ab

#75 (”40-item Fatigue Impact Scale” or “40 item Fatigue Impact Scale”):ti OR (”40-item Fatigue Impact Scale” or “40 item Fatigue Impact Scale”):ab

#76 (”29-item Fatigue Assessment Instrument” or “29 item Fatigue Assessment Instrument”):ti OR (”29-item Fatigue Assessment Instrument” or “29 item Fatigue Assessment Instrument”):ab

#77 (”Functional Assessment of Multiple Sclerosis” or FAMS):ti OR (”Functional Assessment of Multiple Sclerosis” or FAMS):ab

#78 {OR #59-#77}

#79 MeSH descriptor: [Fatigue] this term only

#80 (fatigue NEXT/7 (scale* or subscale* or sub-scale* or questionnaire* or assessment* or inventor* or measure* or tool*)):ti OR (fatigue NEXT/7 (scale* or subscale* or sub-scale* or questionnaire* or assessment* or inventor* or measure* or tool*)):ab

#81 (scale or subscale or sub-scale or questionnaire or assessment or inventory or measure or measurement):ti OR (scale or subscale or sub-scale or questionnaire or assessment or inventory or measure or measurement):ab

#82 (fatigability or fatigable):ti OR (fatigability or fatigable):ab #83 #79 OR #82

#84 #81 AND #83

#85 #80 OR #84

#86 #58 AND #85

## 2 Supplementary Methods 2 risk of bias assessment

### 2.1 Detailed description of methods

Risk of bias assessment of all studies included in the network meta-analysis (NMA) of the present review was undertaken by two experienced reviewers (MMSJ and JL). Any disagreements were resolved through discussion.

Version 2 of the Cochrane risk-of-bias tool (RoB2) for randomised trials (RCTs) [(Sterne et al. 2019] is the recommended tool to assess the risk of bias in RCTs included in Cochrane Reviews. We selected RoB2 as this provides a domain-based approach to identifying biases in RCTs. The tool is structured into five domains through which bias might be introduced into an RCT’s results:

Domain1: bias arising from the randomisation process,

Domain 2: bias due to deviations from intended interventions,

Domain 3: bias due to missing outcome data,

Domain 4: bias in measurement of the outcome; and Domain 5: bias in selection of the reported result.

The judgment for each domain (high risk, low risk, some concerns) is then used to inform an overall risk of bias judgement for each RCT (high risk, low risk, some concerns).

These domains focus on different aspects of trial design, conduct, and reporting. Within each domain, a series of questions (’signalling questions’) aim to elicit information about features of the RCT that are relevant to risk of bias. On the RoB2 tool, each signalling question has up to six possible responses - yes, partial yes, no, partial no, not applicable, no information.

There are 22 signalling questions in total in RoB2, which means that the completion rate for each RCT included in a review can be lengthy. We estimate ≤4 RCT reports per day. For pragmatic reasons relating the volume of studies included in the NMA in the present review, we adapted the RoB2 tool to facilitate quicker completion, whilst still capturing the issues with methodological conduct, reporting and other potential biases we observed in some of the RCTs (independent, small-scale, unregistered, minimally reported) that were included in the present NMA.

Two reviewers with RoB2 experience adapted the RoB2 tool so that some signalling questions that would be redundant for the RCTs included in the present NMA were omitted. The remaining signalling questions (n=15) responses were still yes, no, or unclear. We then adapted the RoB2 algorithms to be able to still judge each domain as low risk, high risk, or some concerns. We also retained the overall RoB2 tool risk of bias judgement algorithm for each RCT as follows:

All domains judged as ‘low risk’ = overall low risk,

Any domain judged as ‘high risk’ = overall high risk,

All domains judged as ‘some concerns’ = overall some concerns

Some domains ‘low risk’ and some domains ‘unclear risk’ = some concerns

Through the adaptation process we were also able to combine some domains resulting in four assessment domains, with a total of 15 signalling questions, as follows:

Domain1: bias arising from the randomisation process – three signalling questions,

Domain 2: bias due to blinding– three signalling questions,

Domain 3: bias due to missing outcome data– five signalling questions; and

Domain 4: Selection of the reported result and analysis of the outcome– four signalling questions

Agreement of the domains and signalling questions to be included in the adapted RoB2 tool was reached through discussion. We developed the adapted RoB2 tool in Excel and the two reviewers independently piloted this on 10 of the RCTs included in the NMA. Any amendments needed to the tool were discussed and agreed through discussion. As a result of this process, a key adjustment was made to the criteria used for risk of bias judgements for ‘attrition’. In consultation with the subject experts on the team we amended our prior threshold of >10% attrition to equal high risk of bias to >20%, due to this level of attrition being within the bounds of normal expectations for behavioural interventions. A copy of the adapted RoB2 tool is presented below.

Reference:

Sterne JAC, Savović J, Page MJ, Elbers RG, Blencowe NS, Boutron I, Cates CJ, Cheng H-Y, Corbett MS, Eldridge SM, Hernán MA, Hopewell S, Hróbjartsson A, Junqueira DR, Jüni P, Kirkham JJ, Lasserson T, Li T, McAleenan A, Reeves BC, Shepperd S, Shrier I, Stewart LA, Tilling K, White IR, Whiting PF, Higgins JPT. RoB 2: a revised tool for assessing risk of bias in randomised trials. BMJ 2019; 366: l4898.

### 2.2 Adapted Risk of Bias 2 assessment criteria

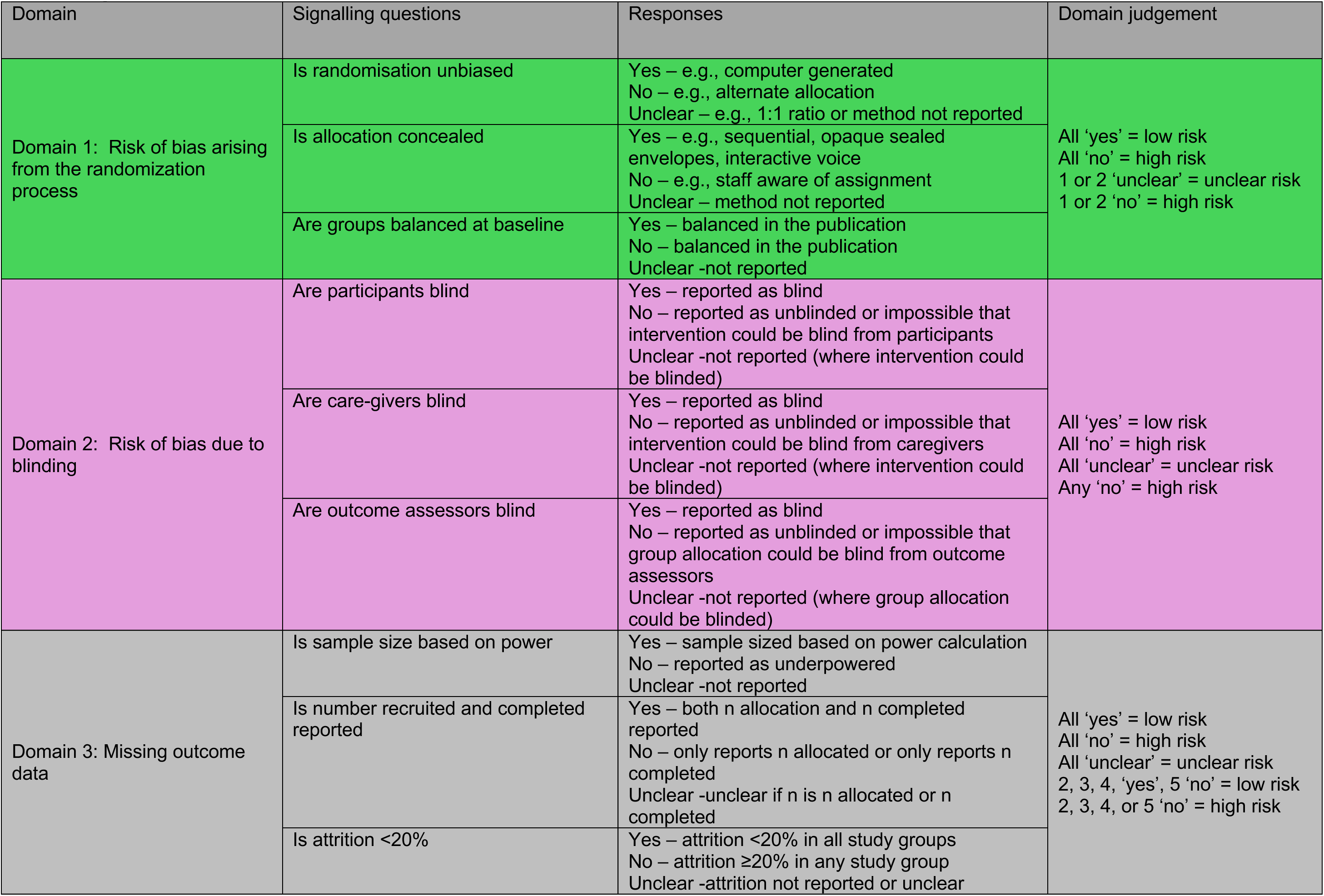
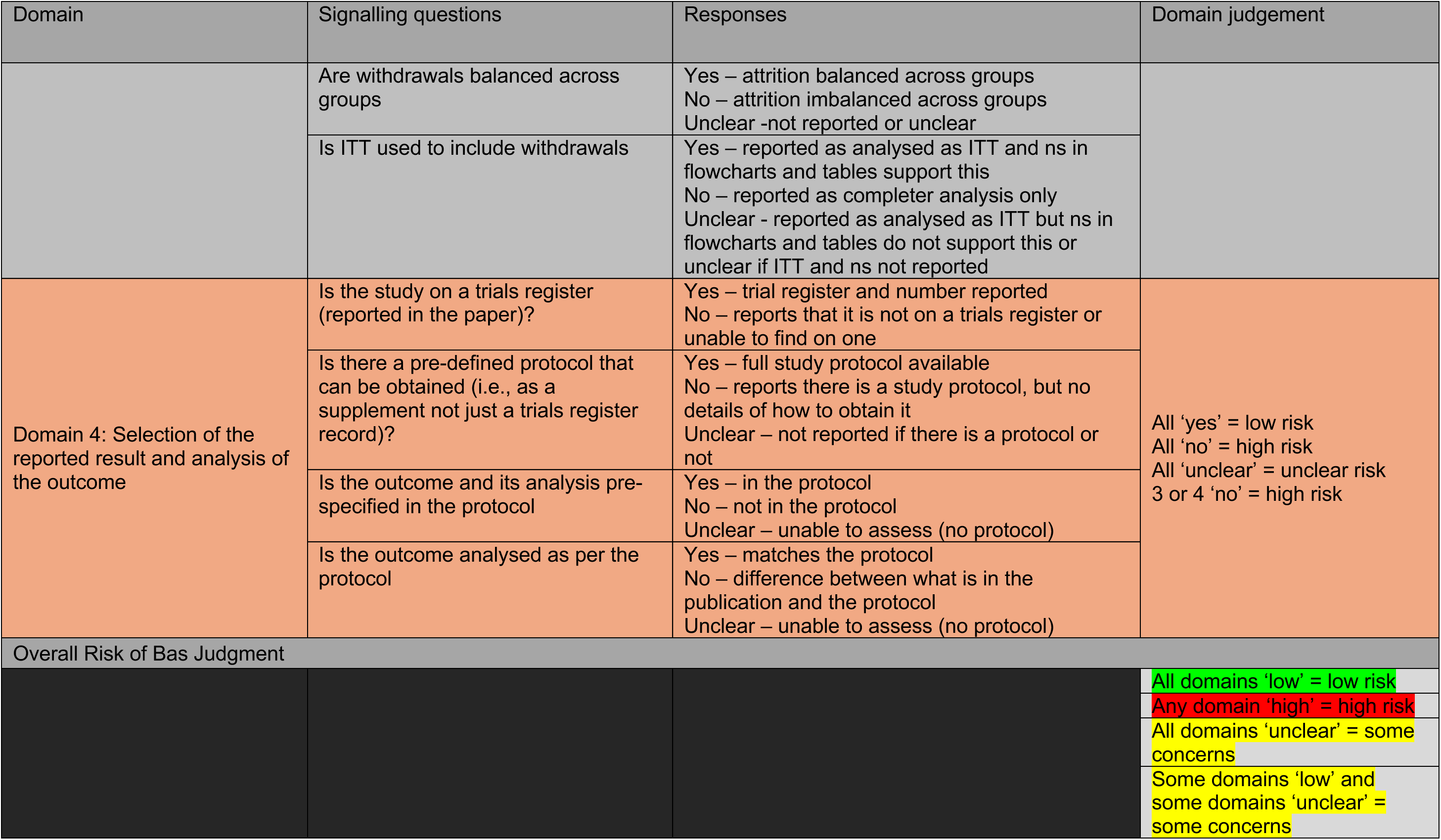

## 3 Supplementary Methods 3 GRADE Classification

### 3.1 Adapted GRADE methods

We assessed the certainty of the effect estimates of non-pharmacological interventions for fatigue compared to usual care using an adaption of GRADE and CINeMA methodology. Methods are described in detail below, and were designed to be appropriate for interpreting results from the body of evidence identified in this review, with judgements based on the data generated by the analyses of our network meta-analysis. The CINeMA framework is largely based on the GRADE framework, with modifications to facilitate assessment of network meta-analyses.

As part of the introduction of CINeMA, a web application has been developed, which enables the conduct of network meta-analyses via the netmeta package in R within the application. As our analyses were conducted using a Bayesian approach and the R2WinBUGS package, we were unable to utilise the CINeMA web application for assessment of the network. Therefore, we used a modified GRADE assessment, incorporating elements of the CINeMA framework for assessment of heterogeneity and inconsistency.

All evidence was derived from randomised controlled trials (RCTs), which were considered to be high quality as a starting point. As per GRADE methodology, the quality of evidence was to be upgraded for large effect size (up one or two levels depending on the magnitude of the effect size) and dose response (up one level). Quality was downgraded for high risk of bias, imprecision, inconsistency and heterogeneity. Number of participants given in the summary tables refers to total N in the relevant intervention arm, not the total in the studies. This is due to inclusion of indirect evidence which may be categorised in another intervention arm (not usual care). The evidence for each outcome was assessed using this framework by JL (RoB, publication bias) and JF (inconsistency, heterogeneity) and validated by the other, or independently in duplicate by JL and JF (imprecision). Disagreements were resolved through discussion. Any uncertainties were discussed with CB. Effect sizes were graded using Cohen’s categories; not substantial (SMD<0.2), small (0.2<=SMD<0.5), medium (0.5<=SMD<0.8), large (0.8<=SMD) (J. Cohen, *Statistical power analysis for the behavioral sciences*. Academic press, 2013). Final ratings were high, moderate, low or very low.

GRADE judgements were based on the following domains of the GRADE book (Neumann et al. 2024):

#### Limitations in the design or execution of randomized trials (RoB)

Individual studies were assessed using Cochrane RoB v2.0 tool. Overall ratings were then clustered by intervention group, as analysed in the NMA. For a variety of reasons, outlined in our detailed RoB section, the evidence included in most intervention groups was rated as high risk of bias and was downgraded by 1. The reason for an overall high RoB rating in many studies was for ‘blinding’, which may not be possible in behavioural interventions. Lack of blinding is more problematic with outcomes that have a subjective component, and as the fatigue measures in the included studies were self-reported and were administered in situations where participants or investigators could influence the probability of the outcomes, studies of these interventions were judged to be at high risk of bias as per the RoB v 2.0 handbook. The few studies with overall ‘some concerns’ were judged to also be at risk of bias and were also downgraded by 1.

#### Inconsistency

For the purpose of applying the GRADE rating, inconsistency was interpreted as any meaningful differences between the direct evidence (provided by the study data used within the NMA) and indirect evidence (resulting from indirect comparisons within the network). Comparisons of interventions relative to usual care were assessed as these are the primary results presented within forest plots. Other comparisons within the network may exhibit potential inconsistencies but not have been within this assessment. Agreement of indirect and direct evidence was assessed via node-splitting, if any of the predicted treatment effects from direct evidence were statistically significantly different from indirect estimates or no direct evidence was available, the comparison was downgraded by 1. If the difference between the two estimates (direct and indirect) differed by an amount greater than 0.34, chosen as a clinically meaningful SMD, the comparison was downgraded by one. At all three timepoints, the majority of interventions could not be assessed for inconsistency of evidence relative to usual care, due to other comparators being used and connecting within the network.

#### Imprecision

Ratings were based around thresholds for the minimal important difference for fatigue. In consultation with subject experts, we used a threshold of an SMD of 0.34 as clinically meaningful, as described in the methods section.

Imprecision was judged on whether the credible intervals of predicted treatment effects spanned both the lower and upper bounds of the clinically meaningful SMD, i.e. −0.34 and +0.34. If a credible interval spanned both −0.34 and +0.34, we rated down by 2 levels (major concerns), if the 95% credible interval spanned SMD=0 and one clinically meaningful threshold, we rated down by 1 (serious imprecision) and if the 95% credible interval was entirely included within the shaded region we did not downgrade.

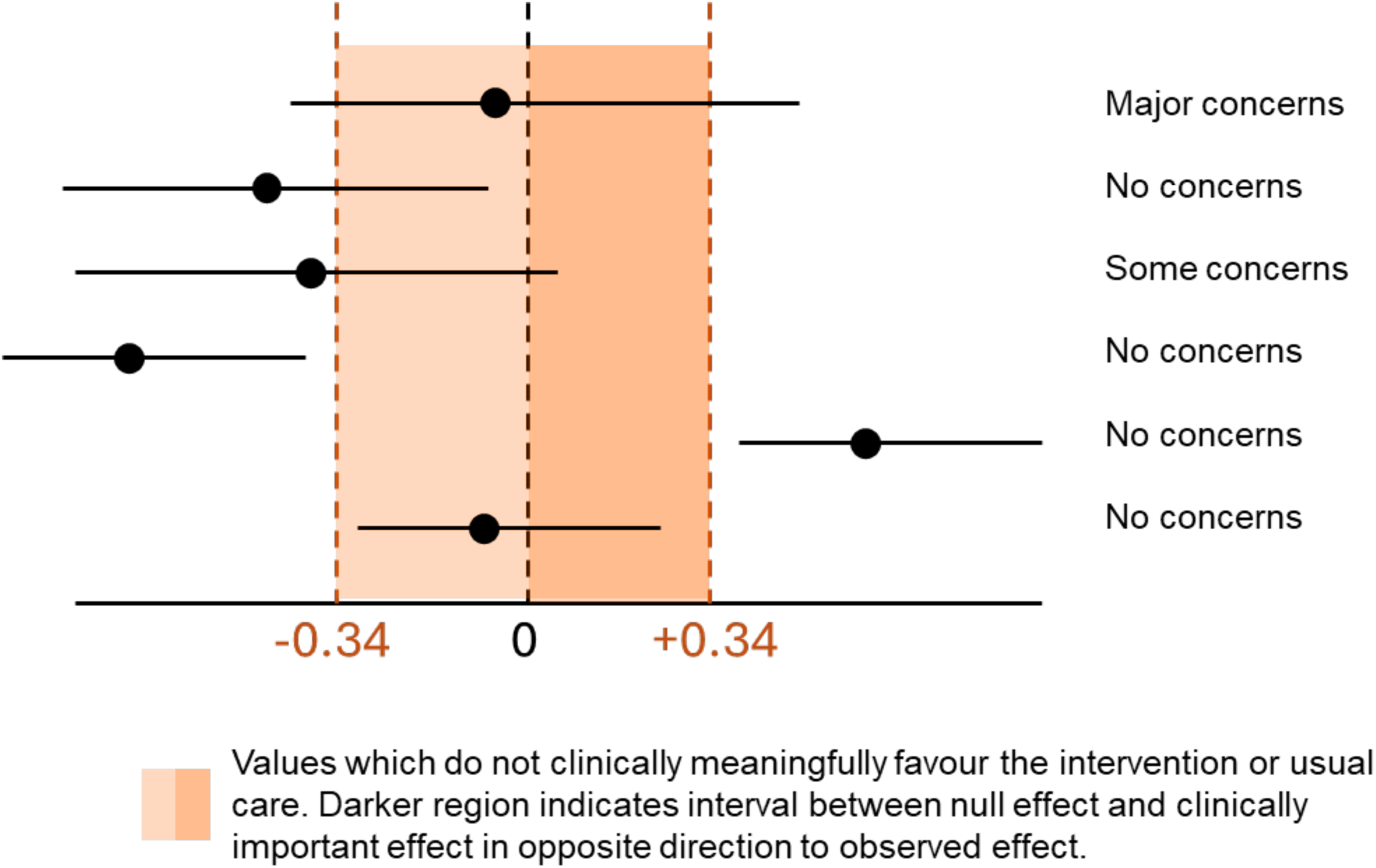

#### Heterogeneity

The 95% prediction intervals were compared to the clinically meaningful thresholds. The predicted intervals capture the uncertainty in the modelled treatment effect but also the heterogeneity between studies. The prediction intervals were graded as for the 95% credible intervals for the imprecision domain, as shown in the figure above.

#### Publication bias

Funnel plots were not created due to too few studies comparing the same two interventions. Any remaining assessments regarding publication bias are by necessity based on subjective judgements around the likelihood that evidence has been missed, for example: non-inclusion of conference abstracts or grey literature; non-publication of negative studies without an external funder; non-publication of negative studies in novel/emerging interventions. We decided therefore not to include publication bias as a formal domain in our overall assessment. Instead we offer the following observations: The body of evidence for non-pharmacological interventions comes from studies that are generally small in nature. These were not always externally funded or published on trials registries and it is therefore possible that other, similar studies with negative results may not have been published. This is also possible for some emerging interventions, for example the stimulation interventions, where there is a risk of publication bias due to studies with negative results potentially remaining unpublished. It is also important to note that just under half of the intervention groups consist of only one study - in fact all of those included in the nutritional group are single study interventions. Because of the small, exploratory nature of some of these single study interventions, we chose not to upgrade for large effect size where a large effect was observed.

References

Schünemann H, Brożek J, Guyatt G, Oxman A, editors. GRADE handbook for grading quality of evidence and strength of recommendations. Updated October 2013. The GRADE Working Group, 2013. Available from guidelinedevelopment.org/handbook.

Nikolakopoulou, A., Higgins, J. P., Papakonstantinou, T., Chaimani, A., Del Giovane, C., Egger, M., & Salanti, G. (2020). CINeMA: an approach for assessing confidence in the results of a network meta-analysis. *PLoS medicine*, *17*(4), e1003082.

## 4 Supplementary Methods 4: Intervention classification criteria

### 4.1 Physical activity-oriented interventions

#### 4.1.1 Exercise Supervised

Multiple sessions involving physical activity in a supervised / observed environment. Can include familiarisation with exercise, addressing barriers to exercise in addition to PA aimed at increasing exercise capacity / strength or fitness. May include recommendations to continue between sessions. May be delivered in groups or 1:1. May be in a gym, community resource, or outdoors. May include advice to repeat at home. Excludes specific forms of activity around training e.g. balance.

#### 4.1.2 Exercise Home

Unsupervised exercise at home aimed at increasing exercise capacity / strength or fitness. May involve initial explanation of physical activity and addressing barriers to exercise.

Includes ongoing contact / review to adapt exercise through the programme. May include initial or occasional observed sessions. May take place at home or at other personally relevant location.

#### 4.1.3 Physical activity promotion

One or more sessions aimed at increasing physical activity by addressing barriers to exercise, goal setting, and encouraging greater physical activity. May involve motivational techniques, cognitive / behavioural features, reporting and feedback, or the use of activity sensors. Less focused on structured exercise regimen than Exercise-Home,

#### 4.1.4 Active Recreational

Engage in physical activity generally used as recreation – includes hippotherapy, dance and Interventions involving specific therapeutic environment. Combine body based activity with indirect positive mental well-being.

### 4.2 Self-management interventions

#### 4.2.1 CBT-based fatigue intervention

Multiple sessions, focused on reducing and adapting to fatigue. May include other symptoms or aspects of the condition. Content includes (1) discussion of helpful / unhelpful thoughts and beliefs (2) behavioural activation - this may include increasing physical activity, management of time / resources, and body/emotion regulating activities (3) tasks / homework between sessions. May be 1:1 or in groups, in person or online. May include second generation features such as Acceptance and Commitment. Can include mindfulness as long as clearly meets CBT definition

#### 4.2.2 Fatigue-self-management-activation

One or more sessions focused on adapting to fatigue and increasing overall activity / engagement. Has only limited emphasis on energy conservation. Includes encouragement to increase activity – either social (behavioural activation) or physical (explicitly increasing physical activity). May include isolated CBT component such as thought challenges but does not meet sufficient criteria for CBT

#### 4.2.3 Fatigue self-management – energy conservation

One or more sessions focused on adapting to fatigue. Primary focus is on energy conservation and prudent allocation. Does not explicitly encourage increase in overall physical or social activity or set out to challenge thoughts. Includes activity pacing and other energy conservation concepts

#### 4.2.4 General / condition specific self-management

Multiple sessions focused on self-management of specific medical condition / disability. May include condition-related fatigue but that is not the primary focus (see Fatigue Self-Management). Includes condition monitoring / specific self-care.

#### 4.2.5 Rehabilitation

Multiple sessions focused on rehab from medical condition / disability. Has specific focus on either function or condition.

### 4.3 Mind / Mind-body interventions

#### 4.3.1 Body-Mind

Multiple sessions at least partly supervised which use approaches to maximise body-mind connection. Can involve traditional methods (yoga, tai-chi) or “scientific” methods e.g. neurofeedback. Emphasises control of the body (contrast with mindfulness which emphasises control of the mind). Also includes Pilates and Exercise-Breathing where slow movement and controlled breathing are combined.

#### 4.3.2 Mind-Body

Interventions focused on mental relaxation / control (contrast with body-mind). Includes relaxation, imagery etc

#### 4.3.3 Mindfulness based stress reduction

Multiple contacts focusing on learning and applying mindfulness-based techniques (meditation / breathwork). May include general guidance on living within energy resources, sleep, mental health and social interaction. Main focus is on applying mindfulness to daily life (rather than explicitly on addressing fatigue – which would be categorised as CBT with mindfulness)

#### 4.3.4 Psychosocial adaptation to condition

Psychosocial intervention focused on adapting to emotional consequences of medical condition. Less explicit structure and content than CBT, more focus on emotional consequences and less on other behavioural factors than Living Well / rehabilitation.

#### 4.3.5 Other specific psychological therapy

Multiple sessions, focused adapting to medical condition without specific focus on fatigue. Includes condition focused cognitive therapy and problem solving

### 4.4 Non-invasive stimulation

#### 4.4.1 CNS Stimulation

Use of one or more sessions of external stimulation of the nervous system either transcranially or via peripheral nerves.

#### 4.4.2 External stimulation

Application of detectable or undetectable external stimulation of the body (includes vibration, heat, light, electromagnetic force

#### 4.4.3 Aromatherapy

Intervention defined as aromatherapy

#### 4.4.4 Touch-based

Therapies that involve the direct (or indirect) use of human touch / interaction. Includes massage, reiki etc. Typically delivered in CAMH settings. May include passive movement.

#### 4.4.5 Acupuncture-type

Interventions using traditional chinese anatomical framework to deliver stimulation - acupuncture, acupressure etc.

### 4.5 Oral Interventions

#### 4.5.1 Plant based

Non-pharmacological supplement described by source rather than ingredient (e.g. paeony extract, ginseng)

#### 4.5.2 Nutritional supplement

Non-pharmacological supplement described by ingredient rather than source (l-carnitine, vitamins)

#### 4.5.3 Diet

Specific dietary intervention (e.g. low-GI, anti-inflammatory)

### 4.6 Education / information

#### 4.6.1 Information

Provision of written / digital information with no more than one session of personal contact

#### 4.6.2 Education

Provision and discussion / tailoring of written / digital information with more than one session of personal contact

### 4.7 Control definitions

#### 4.7.1 Usual care

Usual care or equivalent term either explicit or clearly implied.

#### 4.7.2 Waiting list control

Use of wait list control. Note can include both cross-over design (where arms cross over and all followed to final FU and parallel with no follow up of 2^nd^ arm active intervention.

#### 4.7.3 Placebo

Use of inert ingested substance

#### 4.7.4 Sham

Use of inert external procedure

#### 4.7.5 Control

Includes attentional control (presumed inert activity to adjust for time / attention), unfocused discussion meetings and activities (e.g. writing). Also used as default term if not sufficiently clear

## 5 Supplementary Methods 5: Focus Groups

### 5.1 Patient focus groups

We recruited 5 focus groups in order to reflect diversity of participants, clinical conditions, and location. Each group met on three occasions during the study, in early and mid 2024 and in early 2025. We conducted the focus groups using participatory approaches that we had previously found effective in PPI work with diverse patient groups, including using concise information summaries to inform interactive discussion and activities such as preference sorting. Ethics approval was obtained from the NHS Health Research Authority (23/SC/0292). The focus groups were co-led by an academic researcher (KF) and patient-researchers (DC & SM).

#### 5.1.1 Participants and recruitment

Inclusion and exclusion criteria matched those of the systematic review. We used multiple approaches to ensure a diverse sample involving contacting patients through specialist clinics and recruitment through patient and other community organisations with a focus of ethnic minority heritage. We recruited through the patient organisations and community groups, particularly in communities of minority ethnic heritage. Invitations were sent as posters / flyers as organisations permitted with the opportunity to respond via email or by a dedicated phone number. Individuals who expressed an interest were then contacted and received further information prior to enrolment. Participants provided written consent before the first focus group and this was verbally confirmed at the start of each focus group.

#### 5.1.2 Focus Groups

Focus groups were held online (3) and in community settings (2). The first round of focus groups gave participants the opportunity to describe their experiences of fatigue and to compare and contrast experiences across conditions. The second round focused on potential interventions and involved both description of experiences and discussion about a set of vignettes of potential fatigue interventions. The third round focused on communicating results of the evidence synthesis. The content of the groups was audio-recorded and transcribed before analysis which used thematic analysis. Developing findings were discussed within the study team.

### 5.2 Participant characteristics

The focus groups were recruited through patient organisations and community groups, specifically targeted non-white ethnic heritage. From 44 respondents, we recruited 25 (18 women 7 men) who were able to attend focus group which were held in person (2 groups) and online (3 groups) each on three separate occasions. While some individuals missed one of the series of three groups, none actively withdrew. Although we intended that people stay in the same group allowed people to move between groups and were struck that people were keen to continue their engagement.

Ages of participants ranged from under 30 to over 70, with the most represented age group being 50-59 years. We recruited from diverse ethnic heritages, with 10 identifying as South Asian, 8 as White, 5 as African-Caribbean and 2 others. Long term medical conditions reported included kidney or liver disease (6 participants), arthritis (5) diabetes (5) diabetes, heart conditions and neurological disorders (3 each).

### 5.3 Focus group findings informing the clinical effectiveness analysis

We drew on three themes in framing our analysing and reporting of clinical effectiveness.

1. Fatigue is an invisible problem. Few people had talked constructively about their fatigue with peers or with clinicians. There was little common language or models for explanation about fatigue and few people were aware that interventions for fatigue had been developed.
2. The experience of fatigue crosses diagnostic boundaries. Each individual’s experience of fatigue was personal to them. Where similarities occurred with others, these were as much across conditions as within. Nonetheless, certain features of conditions affected the experience of fatigue or constrained the approaches that might be taken to reduce it.
3. There is no one-size-fits-all solution. Differences in the experience of fatigue extended to differences in what people had found helpful for them (or saw as appropriate to try). There were some instances of scepticism, particularly where an intervention conflicted with prior experiences or beliefs, however in general people were open to considering new information and to trying interventions if these were made available. Participants often ranked availability or accessibility as more important than the particular name or content of an intervention.

## 6 Supplementary Methods 6 - additional statistical analysis methods

### 6.1 Multiple fatigue measures

Some studies reported multiple measures of fatigue. The scale “FSS” was prioritised, following input from the clinical experts on the team, followed by “MFIS”. Therefore, if a study reported multiple fatigue outcomes using different scales, and if one of the outcomes was reported using the “FSS” scale, this data was selected and used for the network meta-analysis (NMA). However, if they did not use the “FSS” scale but used the “MFIS” scale, the data reported using the MFIS scale was selected for the NMA. This ruling resolved most cases, however in cases where there were multiple scales not including “FSS” or “MFIS”, clinical experts were consulted to obtain the most appropriate scale for inclusion within the NMA.

### 6.2 Evaluation of the SMD

As mentioned above, fatigue outcomes were measured using different scoring methods across studies. To facilitate the analysis of studies using different scoring methods within a single NMA, standardised mean differences (SMDs) of the change from baseline of fatigue, were calculated for each study. The use of SMDs is based on the assumption that all scoring scales are quantifying the same treatment effect and can be transformed onto a common scale by dividing the mean difference in change from baseline between the intervention and comparator within each study by the standard deviation of the difference.^1^

Raw data extracted from study results in the form of means, standard deviations, standard errors, inter quartile ranges and confidence intervals were used to evaluate the SMD and subsequently the standard error (SE) of the SMD for each study using Hedge’s correction.^1^ In some studies, change from baseline was reported as opposed to pre and post intervention data, where available this data was extracted and used within the network.

Using intervention 1 as the reference treatment (for most analyses in this work, this corresponds to “usual care”), the SMD for the interventions in arm *t*, at follow-up *f* is given by

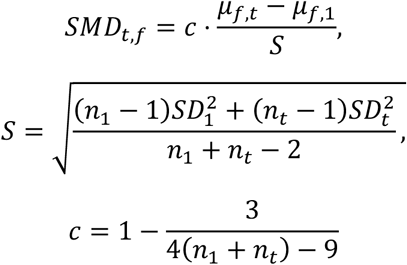

where *μ*_*f*,*t*_ and *μ*_*f*,1_ is the change in fatigue score before and after treatment in arm *t* and arm 1 at follow-up *f*, respectively; *S* is the within group standard deviation pooled across arms, *SD*_*t*_ and *SD*_1_ are the standard deviations in arm t and arm 1, respectively; *n*_*t*_ and *n*_1_ are the number of participants at baseline in arm *t* and arm 1, respectively; and *c* is Hedges’ correction factor. The standard error (SE) of the SMD is given by

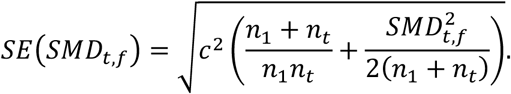

For studies where only the 95% confidence interval was presented alongside the mean instead of the standard deviation, the standard deviation was evaluated using

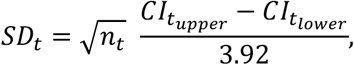

where *n*_*t*_ is the number of participants in study arm *t*, *CI*_*tupper*_ and *CI*_*tlo*w*er*_ are the upper and lower 95% confidence intervals. For cases where only the inter-quartile range was recorded, the standard deviation was evaluated as

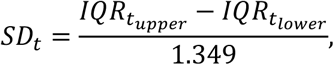

where *IQR*_*t*_ and *IQR*_*t*_ are the upper and lower quartiles.^2^ In most cases, fatigue scores were presented at baseline and then post-treatment, the mean change from baseline was therefore evaluated and the standard deviation of the mean change from baseline calculated using

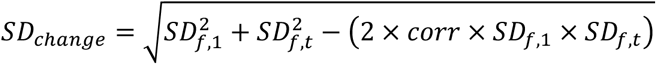

where the correlation coefficient (*corr*) was assumed to be equal to 0.5 as a conservative estimate.^3–5^

For studies where multiple arms presented data for the same intervention, according to the intervention classification conducted by the clinical experts, the data were combined. The mean change from baseline was evaluated as a weighted average, according to the number of participants in the arms being combined. The standard deviation was then evaluated as

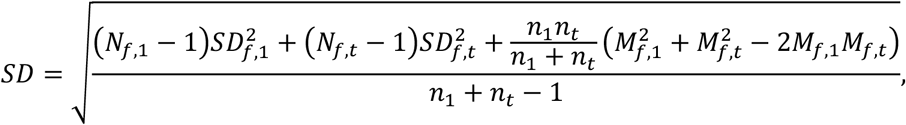

where *M*_*f*,*t*_ is the weighted average of the mean change from baseline in arm *t* at follow-up *f*.^5^

In the case where no available data were available to evaluate the mean and standard deviation of the change from baseline of the fatigue score for each arm, the study was not included within the NMA. No studies which only presented data graphically were included within the NMA due to the high number of studies.

### 6.3 Statistical model for the NMA

A random-effects NMA model was used to account for between study heterogeneity.^6^ Let *y*_*ik*_ denote the standardised mean difference (SMD) of arm *k* of trial *i*, where *k* = 1,…, *na* and *i* = 1,…, *ns*, with variance *V*_*ik*_. Here, *na* and *ns* correspond to the number of arms and the number of studies respectively. We assume that the treatment effects are normally distributed according to

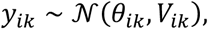

where **θ** are the parameters of interest. The individual *θ*_*ik*_ are modelled using the identity link function as they are continuous on the entire real line

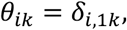

where *δ*_*i*,1*k*_ is the individual study treatment effect of intervention *k* relative to intervention 1 in study *i*. To allow for heterogeneity of treatment effects across studies, a random-effects model was assumed. The random-effects model was structured such that all individual study treatment effects arise from a common normal distribution centred about a mean population treatment effect, with some variance *τ*^2^

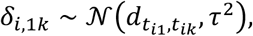

where *d*_*t*_*i*_,*t*_*ik*__ is the mean effect of the intervention in arm *k* of study *i* (*t*_*ik*_) compared to the intervention in arm 1 of study *i* (*t*_*i*1_).

In the case of studies with more than two arms, adjustment to the likelihood (function) was necessary to account for the correlation between multiple comparisons to arm 1 and was included via the assumption that the covariance between two comparisons relative to treatment 1 can be approximated as^1^/_*ni*,1_, where *n*_*i*,1_ is the number of participants in arm 1 of study *i* at baseline.^7^

### 6.4 Definition of priors

Parameters were estimated using a Bayesian framework, as such, non-informative priors were chosen for the between-study variance of treatment effects

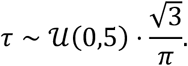

The 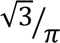 factor is included to account for the transformation of *τ* between the odds-ratio scale and the SMD scale.^8^ An informative prior, a truncated log-normal prior on *τ*^2^ was used in cases where there were less than 5 studies within the connected network.^8^

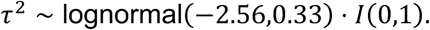

The prior on the mean treatment effects were defined as

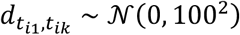

### 6.5 Implementation

All analyses were conducted using the freely available software package WinBUGS^9^ and R, via the R2Winbugs^10^ interface package. Model code was modified from NICE technical support document 2.^11^ Convergence to the target posterior distributions was assessed using the Gelman-Rubin statistic, as modified by Brooks and Gelman, for three chains with different initial values.^12^ The autocorrelation of samples from the burn-in period was also assessed for any significant autocorrelation which requires sample thinning. A burn-in period of 50,000 samples was implemented, with a further 1,000,000 samples after the burn-in period. The samples after the burn-in were subject to a thinning by a factor of 10.

Results are presented using the posterior median treatment effects and 95% credible intervals (CrI).

The validity of the inconsistency assumption was assessed by comparing the posterior mean residual deviance from the unrelated mean effects model and the NMA model; and node-splitting analysis. The posterior means of the deviance contributions for the unrelated mean effects model and the NMA model were plotted, cases where the posterior means lie across the *y* = *x* line, demonstrated that the inconsistency assumption held. Cases where the posterior deviance contributions deviated from this line (indicating an improvement greater than 0.5 points in the unrelated mean effects model) were investigated using node-splitting via the gemtc package in R.^13^

### 6.6 References

Higgins JPT, Altman DG, Sterne JAC (editors). Chapter 8: Assessing risk of bias in included studies. In: Higgins JPT, Churchill R, Chandler J, Cumpston MS (editors), Cochrane Handbook for Systematic Reviews of Interventions version 5.2.0 (updated June 2017), Cochrane, 2017. Available from www.training.cochrane.org/handbook.

## EIFFEL Supplementary Results

## Supplemental Results 1. Studies included in Network Meta-analysis

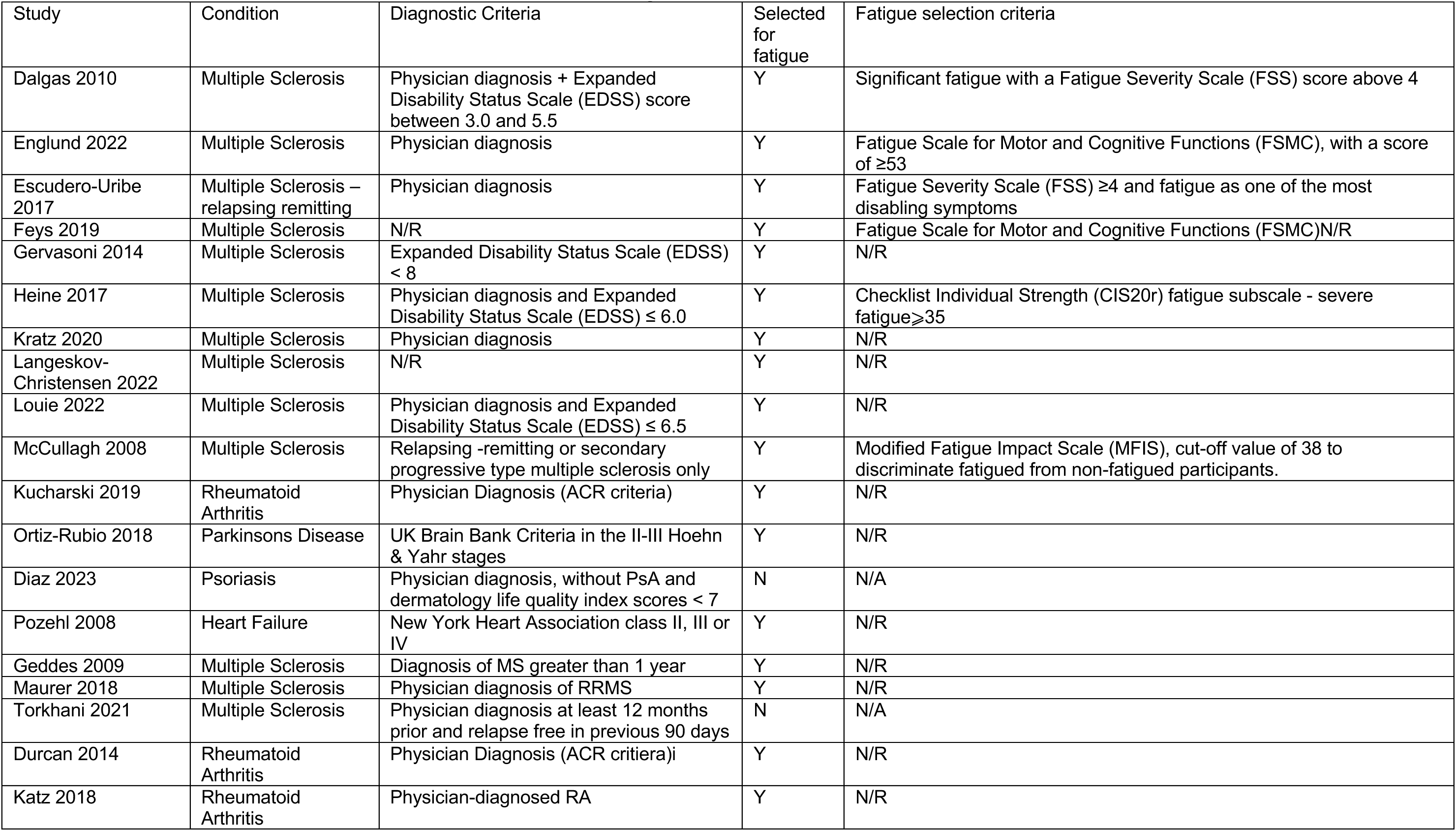
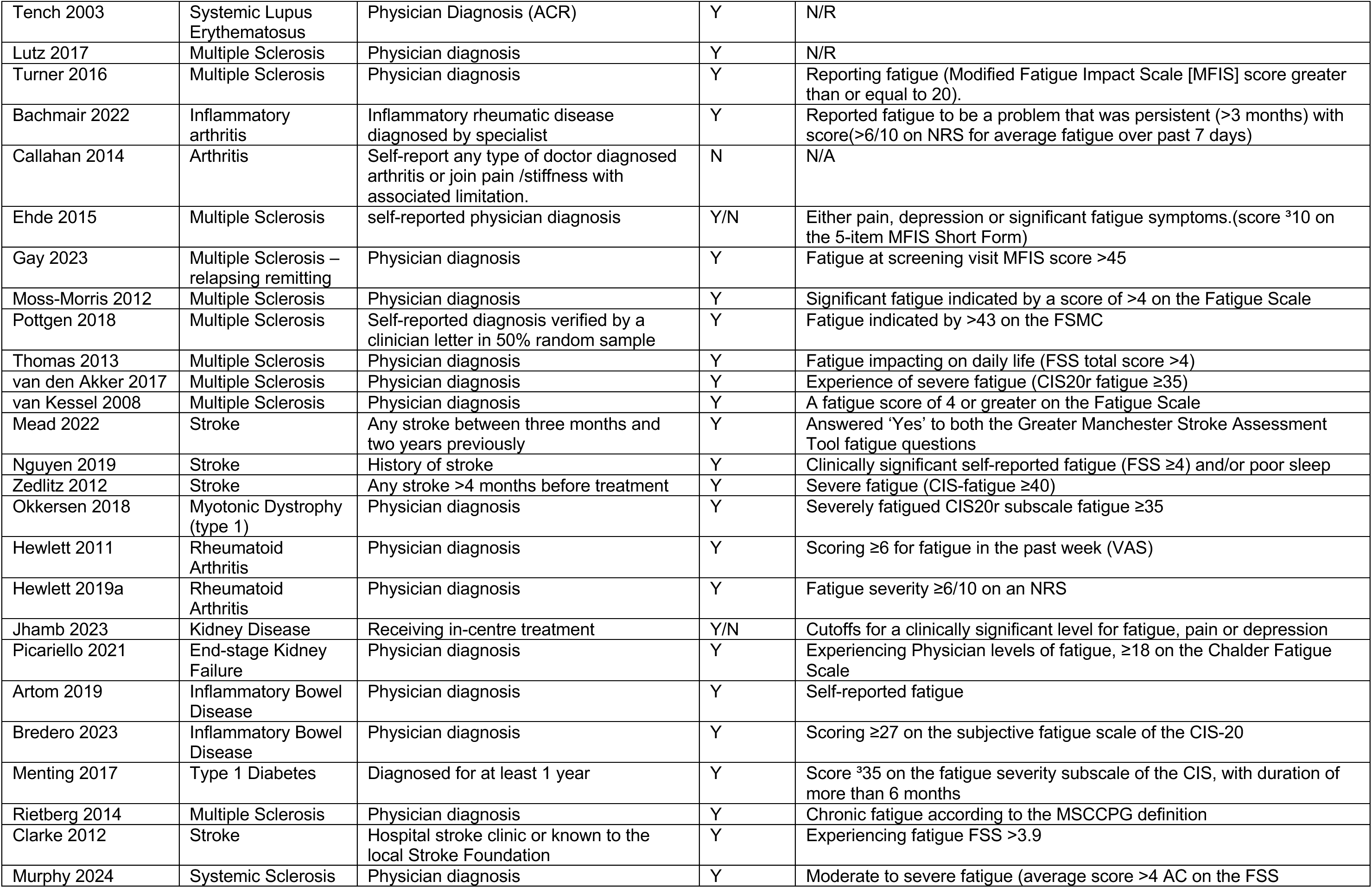
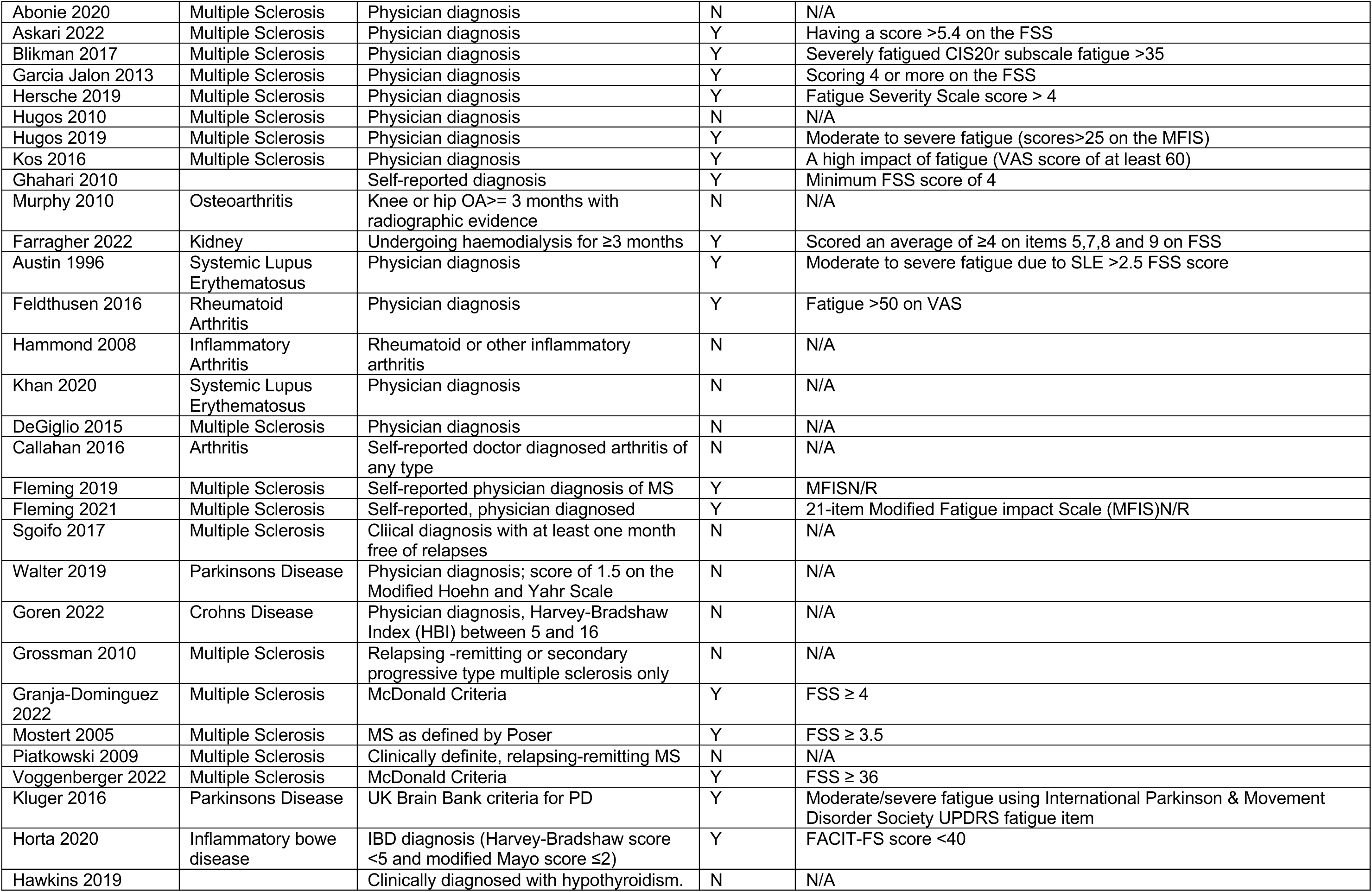
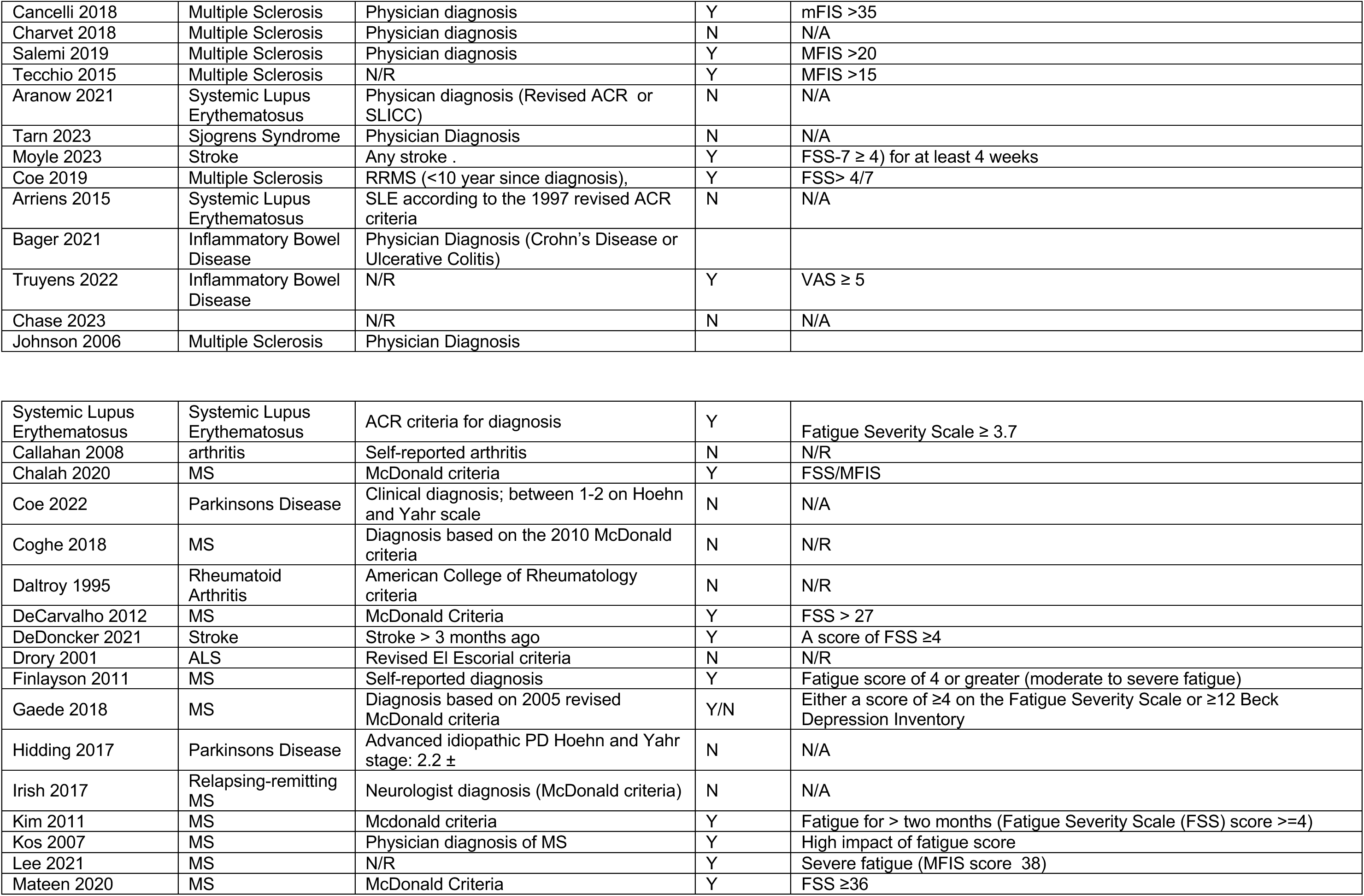
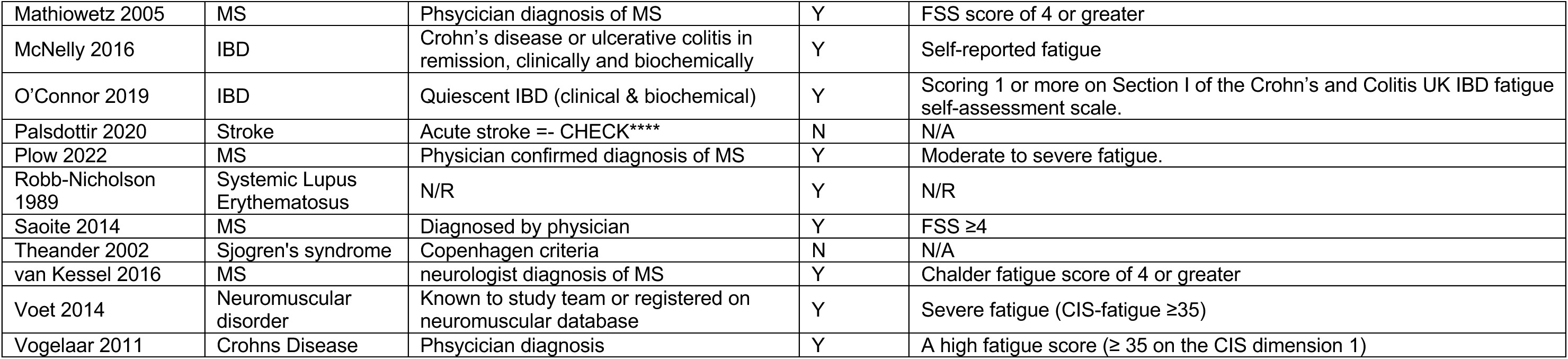

## Supplemental Results 2. Characteristics of potentially eligible studies, not included in network meta- analysis

Characteristics of potentially eligible studies, not included in network meta-analysis
|  |  |  |  |  |
| --- | --- | --- | --- | --- |
| Avaux 2016 | Systemic Lupus Erythematosus | ACR criteria for diagnosis | Y | Fatigue Severity Scale $\geq 3.7$ |
| Callahan 2008 | arthritis | Self-reported arthritis | N | N/R |
| Chalah 2020 | MS | McDonald criteria | Y | FSS/MFIS |
| Coe 2022 | Parkinsons Disease | Clinical diagnosis; between 1-2 on Hoehn and Yahr scale | N | N/A |
| Coghe 2018 | MS | Diagnosis based on the 2010 McDonald criteria | N | N/A |
| Daltroy 1995 | Rheumatoid Arthritis | American College of Rheumatology criteria | N | N/A |
| DeCarvalho 2012 | MS | McDonald Criteria | Y | FSS > 27 |
| DeDoncker 2021 | Stroke | Stroke > 3 months ago | Y | A score of FSS $\geq 4$ |
| Drory 2001 | ALS | Revised El Escorial criteria | N | N/R |
| Finlayson 2011 | MS | Self-reported diagnosis | Y | Fatigue score of 4 or greater (moderate to severe fatigue) |
| Gaede 2018 | MS | Diagnosis based on 2005 revised McDonald criteria | Y/N | Either a score of $\geq 4$ on the Fatigue Severity Scale or $\geq 12$ Beck Depression Inventory |
| Hidding 2017 | Parkinsons Disease | Advanced idiopathic PD Hoehn and Yahr stage: $2.2 \pm$ | N | N/A |
| Irish 2017 | Relapsing-remitting MS | Neurologist diagnosis (McDonald criteria) | N | N/A |
| Kim 2011 | MS | McDonald criteria | Y | Fatigue for > two months (Fatigue Severity Scale (FSS) score $\geq 4$) |
| Kos 2007 | MS | Physician diagnosis of MS | Y | High impact of fatigue score |
| Lee 2021 | MS | N/R | Y | Severe fatigue (MFIS score 38) |
| Mateen 2020 | MS | McDonald Criteria | Y | FSS $\geq 36$ |
| Mathiowetz 2005 | MS | Physician diagnosis of MS | Y | FSS score of 4 or greater |
| McNelly 2016 | IBD | Crohn's disease or ulcerative colitis in remission, clinically and biochemically | Y | Self-reported fatigue |
| O'Connor 2019 | IBD | Quiescent IBD (clinical & biochemical) | Y | Scoring 1 or more on Section I of the Crohn's and Colitis UK IBD fatigue self-assessment scale. |
| Palsdottir 2020 | Stroke | Admitted to hospital with acute stroke or in the chronic phase (1 year post stroke) | N | N/A |
| Plow 2022 | MS | Physician confirmed diagnosis of MS | Y | Moderate to severe fatigue. |
| Robb-Nicholson 1989 | Systemic Lupus Erythematosus | N/R | Y | N/R |
| Sabapathy 2011 | MS | N/R | N | N/A |
| Saoite 2014 | MS | Diagnosed by physician | Y | FSS $\geq 4$ |
| Theander 2002 | Sjogren's syndrome | Copenhagen criteria | N | N/A |
| van Kessel 2016 | MS | Neurologist diagnosis of MS | Y | Chalder fatigue score of 4 or greater |
| Voet 2014 | Neuromuscular disorder | Known to study team or registered on neuromuscular database | Y | Severe fatigue (CIS-fatigue $\geq 35$) |
| Vogelaar 2011 | Crohns Disease | Physician diagnosis | Y | A high fatigue score ($\geq 35$ on the CIS dimension 1) |
| Vogelaar 2014 | IBD | Diagnosis of IBD of at least 6 months | Y | CIS-fatigue score of $\geq 35$ |

## Supplemental Results 3. Reasons for non-inclusion of studies in NMA

Reasons for non-inclusion of studies in NMA
| Studies not included in NMA |  |
| --- | --- |
| Study | Reason for exclusion from NMA |
| Avaux 2016 | Graphical only |
| Callahan 2008 | No SDs for EOT, means only |
| Chalah 2020 | No Ns for each intervention group |
| Coe 2022 | Graphical data only |
| Coghe 2018 | Wilcoxon test statistics only, no means |
| Daltroy 1995 | Means only or group difference no SDs |
| De Carvelho 2012 | Graphical only |
| De Doncker 2021 | Graphical only |
| Drory 2001 | Graphical only |
| Finlayson 2011 | Between group t-test only, no means only Cohen's D |
| Gaede 2018 | No means and SDs, graphs and reduction numbers |
| Hidding 2017 | Individual patient data only |
| Irish 2017 | Graphs and percentage increases only |
| Kim 2011 | Graphical only, means only for baseline |
| Kos 2007 | Change scores compares groups |
| Lee 2021 | Graphical only |
| Mateen 2020 | No SDs |
| Mathiowetz 2005 | Data is difference between groups |
| McNelly 2016 | 4 x 4 factorial with merged groups |
| O'Connor 2019 | No SDs |
| Palsdottir 2020 | No SDs |
| Plow 2022 | Beta coefficients or graphical only |
| Robb-Nicholson 1987 | Correlations plus baseline data only |
| Sabapathy | Intervention arms the same category |
| Saiote 2014 | Fatigue scores graphical or means by responders/non responders |
| Theander 2002 | Correlations only |
| van Kessel 2016 | 2 arms same intervention category |
| Voet 2014 | Median and ranges only |
| Vogelaar 2011 | No Means, % of participants with decreased fatigue |
| Vogelaar 2014 | No Means, % of participants with decreased fatigue |

## Supplemental Results 4. Timing of outcome measures

### 4.1 Self-management interventions

| Behavioural Interventions |  |  |  |  |  |
| --- | --- | --- | --- | --- | --- |
| Study | Population | Intervention duration (EOT) | Short term | Long term | Short Long term (if any) |
| **Self-Management** |  |  |  |  |  |
| **Fatigue self-management - conservative** |  |  |  |  |  |
| Abonie 2020 | MS | 4 weeks |  |  |  |
| Askari 2022 | MS | 12 weeks |  |  |  |
| Blikman 2017 | MS | 4 months | 10 weeks | 36 weeks |  |
| Farragher 2022 | Kidney | 8 weeks | 12 weeks |  |  |
| Finlayson 2011 | MS | 6 weeks | 3 months | 3 months |  |
| GarciaJalon 2013 | MS | 5 weeks | 3 months |  |  |
| Ghahari 2010 | Chronic | 7 weeks | 3 months |  |  |
| Hersche 2019 | MS | 3 weeks | 3 months |  |  |
| Hugos 2010 | MS | 6 weeks |  |  |  |
| Hugos 2019a | MS | 6 weeks | 3 months | 6 months |  |
| Kos 2016 | MS | 3 weeks | 3 months |  |  |
| Mathiowetz 2005 |  |  |  |  |  |
| Murphy 2010 | OA | 2 weeks | 10 weeks |  |  |
| **Fatigue self-management - active** |  |  |  |  |  |
| Clarke 2012 | Stroke | 6 weeks | 3 months |  |  |
| Murphy 2024 | Sys. Sclerosis | 12 weeks |  |  |  |
| O'Connor 2019 | IBD | 6 months |  |  |  |
| Rietberg 2014 | MS | 12 weeks | 12 weeks |  |  |
| Vogelaar 2014 | IBD | 3 months |  | 3 months |  |
| **General self-management** |  |  |  |  |  |
| Austin 1996 | SLE | 6 months |  |  |  |
| Feldthusen 2016 | RA | 12 weeks |  | 6 months |  |
| Hammond 2008 | RA |  |  | 12 months | 6 months |
| Khan 2020 | SLE | 16 weeks |  |  |  |
| **CBT – fatigue** |  |  |  |  |  |
| Artom 2019 | IBD | 8 weeks |  | 10 months | 4 months |
| Bredero 2023 | IBD | 8 weeks |  |  |  |
| Ehde 2015 | MS |  |  | 10 months | 4 months |
| Gay 2023 | MS | 6 weeks |  | 12 months | 6 months |
| Hewlett 2011 | RA | 6 weeks |  |  |  |
| Hewlett 2019a | RA | 6 weeks |  | 46 weeks | 20 weeks |
| Jhamb 2023 | Kidney | 12 weeks |  |  |  |
| Mead 2022 | Stroke | 4 months | 2 months |  |  |
| Menting 2017 | T1 Diabetes | 5 months |  |  |  |
| Moss-Morris 2012 | MS | 10 weeks |  |  |  |
| Nguyen 2019 | Stroke | 2 months | 2 months |  |  |
| Okkersen 2018 | MD | 10 months |  |  |  |
| Picariello 2021 | Kidney | 12 weeks |  |  |  |
| Pottgen 2018 | MS | 12 weeks | 12 weeks |  |  |
| Thomas 2013 | MS | 6 weeks |  | 1 month | 4 months |
| van Kessel 2008 | MS | 8 weeks | 3 months | 6 months |  |
| Van Kessel 2016 | MS | 10 weeks |  |  |  |
| van den Akker 2017 | MS | 4 months | 10 weeks | 36 weeks |  |
| Zedlitz 2012 | Stroke | 12 weeks |  | 6 months |  |
| **Physical Activity** |  |  |  |  |  |
| **Physical activity promotion** |  |  |  |  |  |
| Bachmair 2022 | Inflammatory | 22 weeks | 6 weeks | 34 weeks |  |
| Callahan 2014 | Arthritis | 20 weeks |  |  |  |
| Lutz 2017 | MS | 6 weeks |  |  |  |
| Turner 2016 | MS | 6 months |  |  |  |
| **Exercise – supervised** |  |  |  |  |  |
| Dalgas 2010 | MS | 12 weeks | 12 weeks |  |  |
| Diaz 2023 | PsO | 16 weeks |  |  |  |
| Englund 2022 | MS | 12 weeks |  |  |  |
| Escudero-Urbe 2017 | MS | 12 weeks |  |  |  |
| Heine 2017 | MS | 16 weeks |  | 36 weeks |  |
| Feys 2019 | MS | 12 weeks |  |  |  |
| Gervasoni 2014 | MS | 2 weeks |  |  |  |
| Kratz 2020 | MS | 8 weeks |  |  |  |
| Kucharski 2019 | RA | 20 weeks |  | 12 months |  |
| Langeskov-Christensen 2022 | MS | 24 weeks |  |  |  |
| Louie 2022 | MS | 12 weeks | 12 weeks |  |  |
| McCullagh 2008 | MS | 12 weeks | 3 months |  |  |
| Ortiz-Rubio 2018 | PD | 8 weeks |  |  |  |
| Pozehl 2008 | HF | 24 weeks |  |  |  |
| **Exercise – unsupervised** |  |  |  |  |  |
| Durcan 2014 | Arthritis | 12 weeks |  |  |  |
| Geddes 2009 | MS | 12 weeks |  |  |  |
| Katz 2018 | RA | 21 weeks |  |  |  |
| Maurer 2018 | MS | 6 months |  |  |  |
| Tench 2003 | SLE | 12 weeks |  |  |  |
| **Active recreational** |  |  |  |  |  |
| **Rehabilitation** |  |  |  |  |  |
| DeGiglio 2015 | MS | 8 weeks |  |  |  |
| **Mindbody** |  |  |  |  |  |
| Callahan 2016 | Arthritis | 8 weeks |  |  |  |
| Fleming 2019 | MS | 8 weeks |  |  |  |
| Fleming 2021 | MS | 8 weeks |  |  |  |
| Walter 2019 | PD | 8 weeks |  |  |  |
| Sgoifo 2017 | MS | 8 weeks |  |  |  |
| **Mindfulness** |  |  |  |  |  |
| Goren 2022 | Crohn's | 3 months |  |  |  |
| Grossman 2010 | MS | 8 weeks |  | 6 months |  |
| Torkhani 2021 | MS | 8 weeks |  |  |  |

4.2 Stimulation interventions
| Study | Population | Intervention duration (EOT) | Short term | Long term | Short Long term (if any) |
| --- | --- | --- | --- | --- | --- |
| **Vagal stimulation** |  |  |  |  |  |
| Aranow 2021 | SLE | 5 days | 1 week |  |  |
| Tarn 2023 | Sjogren's | 54 days |  |  |  |
| **Trans Cranial stimulation** |  |  |  |  |  |
| Cancelli 2018 | MS | 5 days |  |  |  |
| Chalah 2020 | MS | 5 days |  |  |  |
| Charvet 2018 | MS | 4 weeks |  |  |  |
| Salemi 2019 | MS | 2 weeks | 1 month |  |  |
| Tecchio 2015 | MS | 5 days |  |  |  |
| **External stimulation** |  |  |  |  |  |
| Granja-Dominguez 2022 | MS | 4 weeks | 3 months |  |  |
| Mostert 2005 | MS | 4 weeks |  |  |  |
| Piatkowski 2009 | MS | 12 weeks |  |  |  |
| Voggenberger 2022 | MS | 2 weeks |  |  |  |
| **Aromatherapy** |  |  |  |  |  |
| Hawkins 2019 | Hypothyroidism | 14 days |  |  |  |
| **Acupuncture/acupressure** |  |  |  |  |  |
| Horta 2020 | IBD | 8 weeks | 8 weeks |  |  |
| Kluger 2016 | PD | 6 weeks |  |  |  |
| **RIC** |  |  |  |  |  |
| Moyle 2023 | Stroke | 6 weeks | 6 weeks | 18 weeks |  |

4.3 Nutritional Interventions
| **Study** | **Population** | **Intervention duration (EOT)** | **Short term** | **Long term** | **Short Long term (if any)** |
| --- | --- | --- | --- | --- | --- |
| **Fish oil** |  |  |  |  |  |
| Arriens 2015 | SLE | 6 months |  |  |  |
| **Thiamine HD** |  |  |  |  |  |
| Bager 2021 | IBD | 4 weeks |  |  |  |
| **5-HTP** |  |  |  |  |  |
| Truyens 2022 | IBD | 8 weeks |  |  |  |
| **Flavenoid - cocoa** |  |  |  |  |  |
| Coe 2019 | MS | 6 weeks |  |  |  |
| **Diet** |  |  |  |  |  |
| Chase 2023 | MS | 16 weeks |  |  |  |
| **Plant** |  |  |  |  |  |
| Johnson 2006 | MS | 4 weeks |  |  |  |

### Supplemental Results 5. Intervention content: studies included in NMA

#### 5.1 Behavioural Interventions

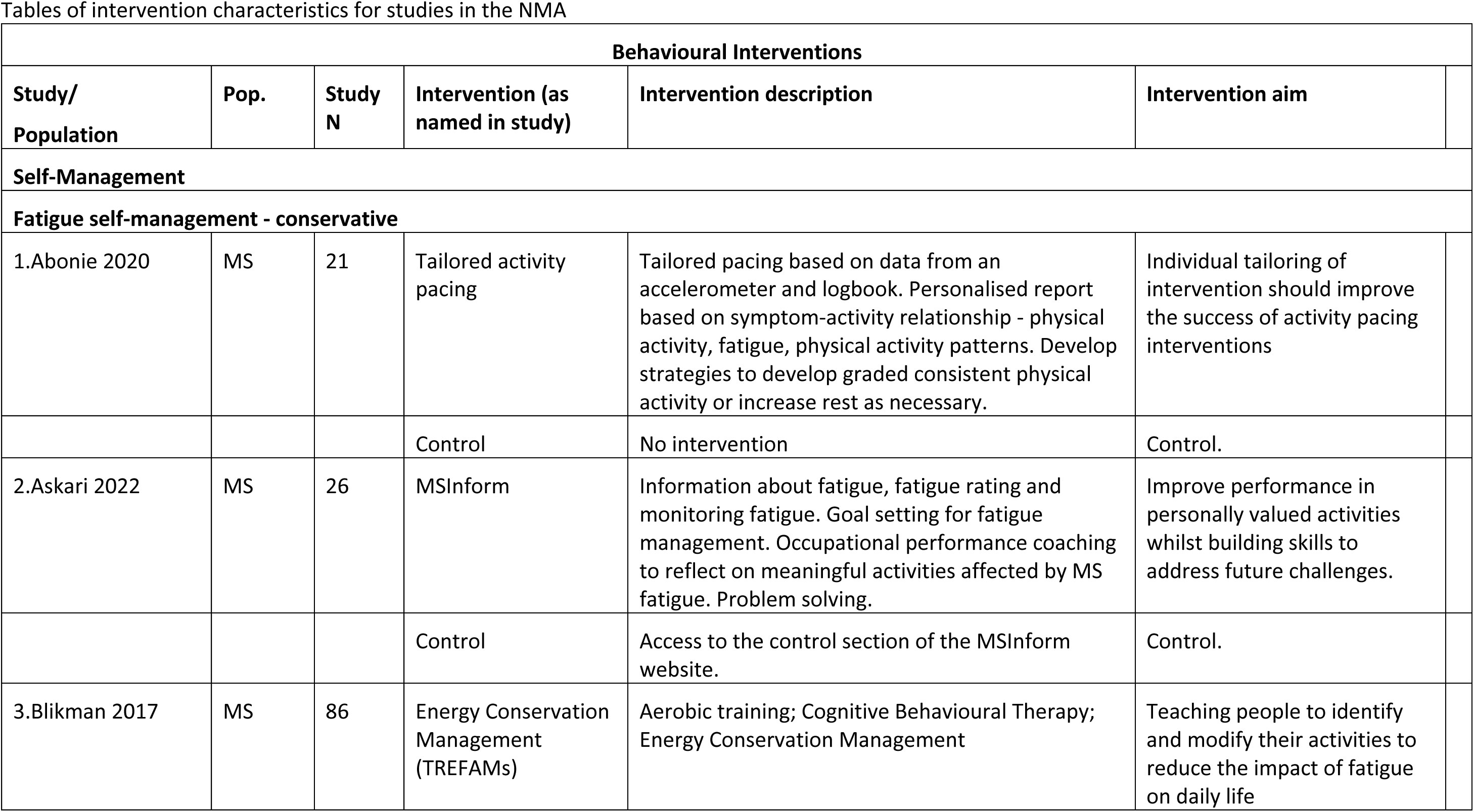
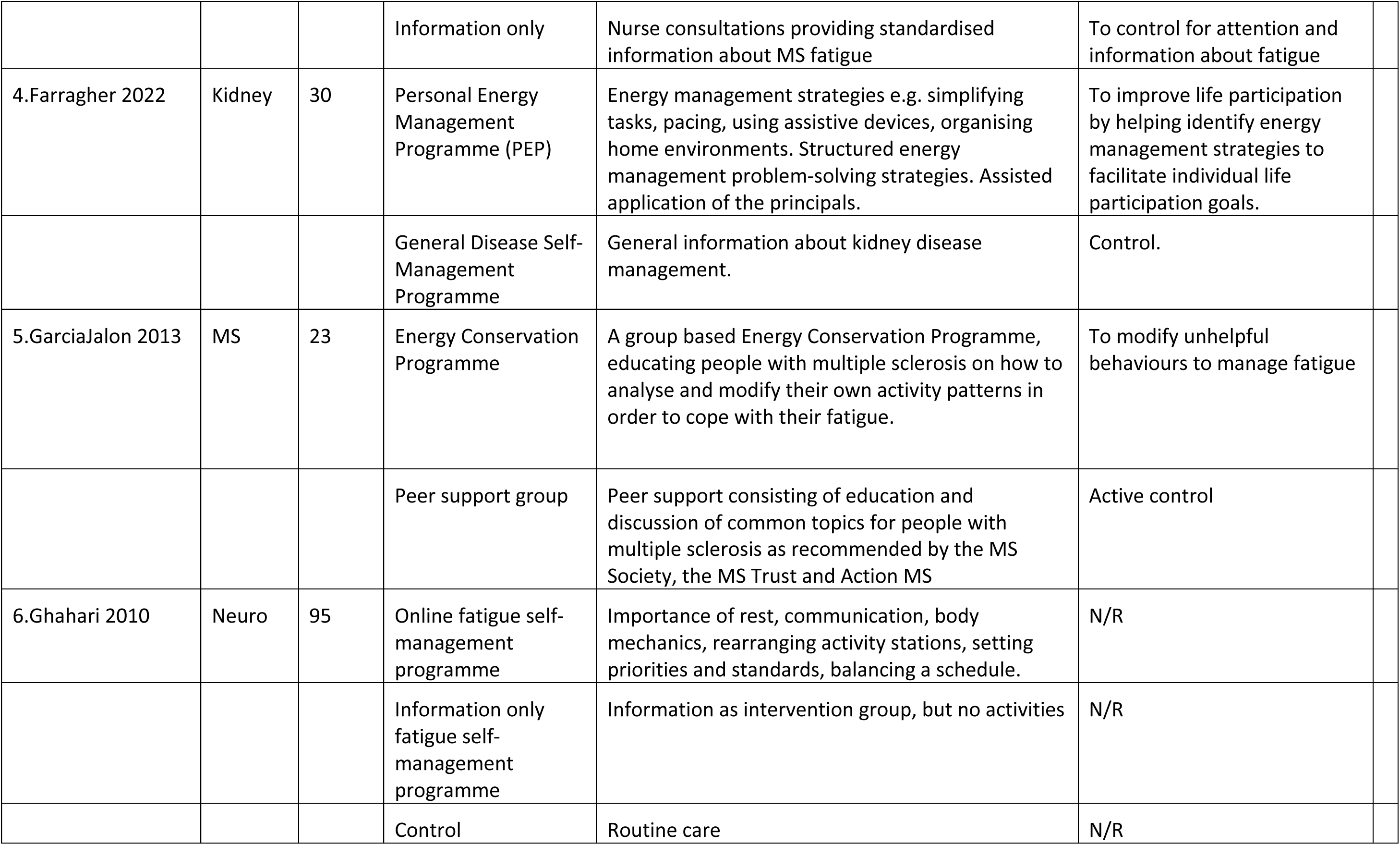
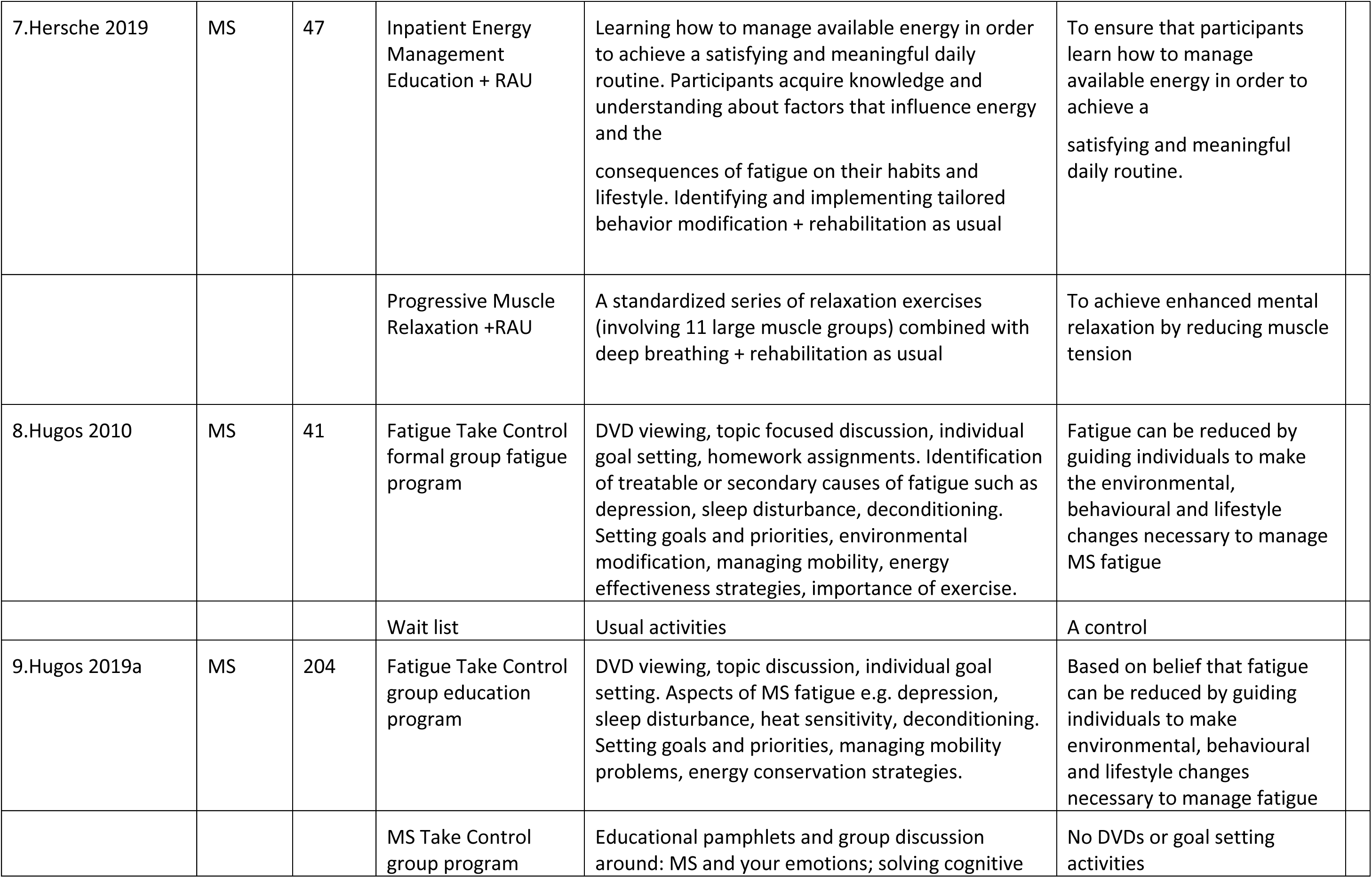
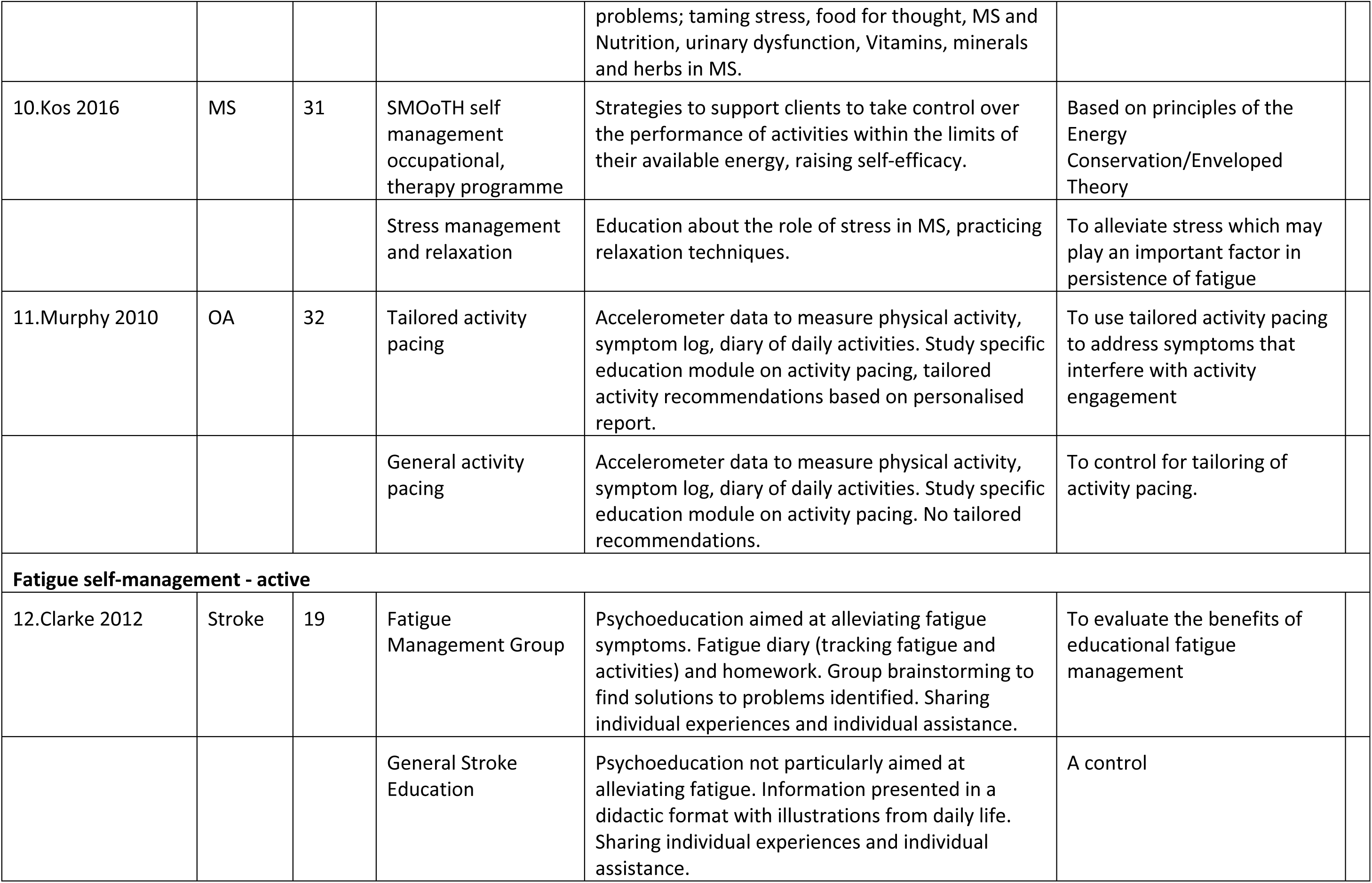
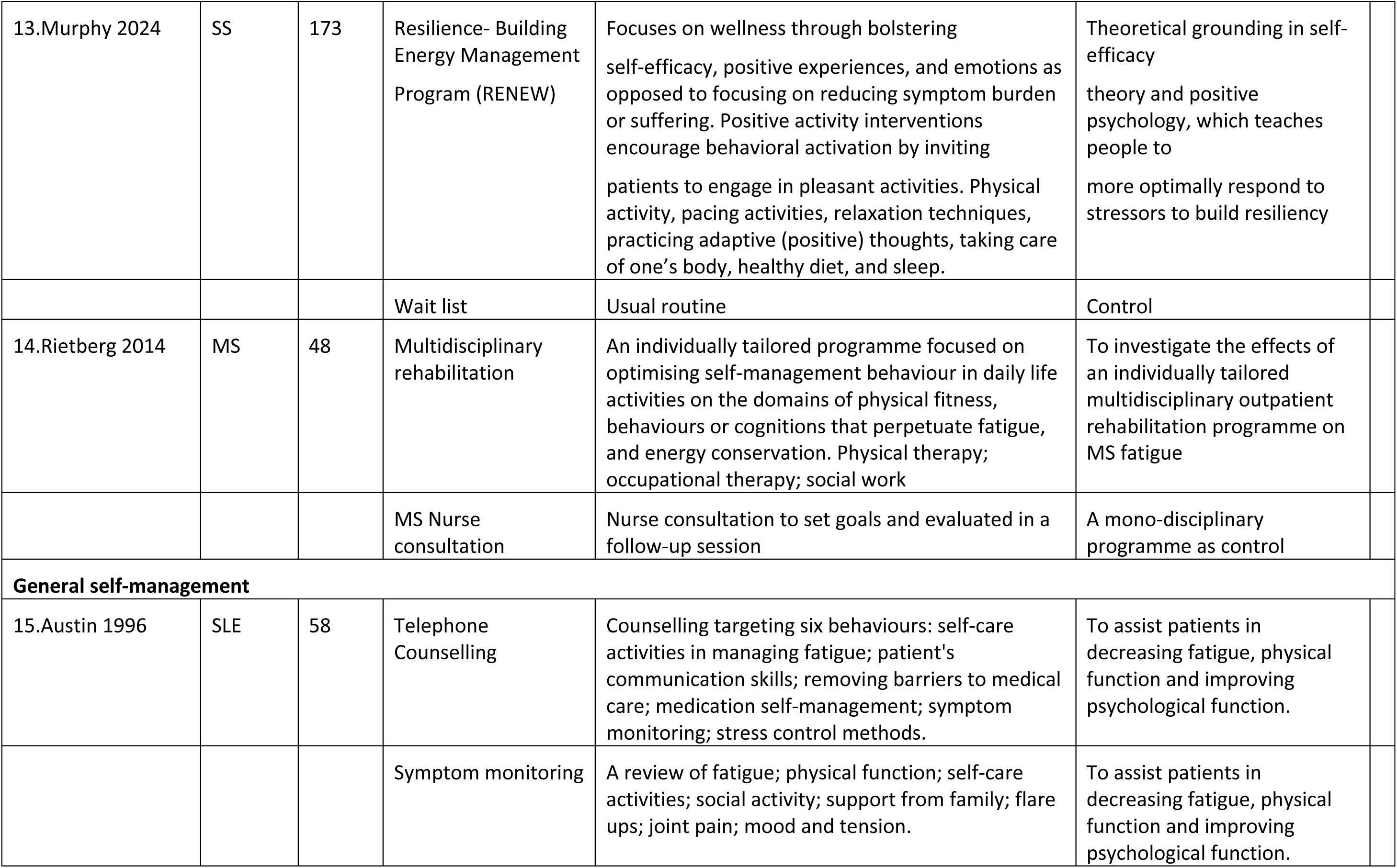
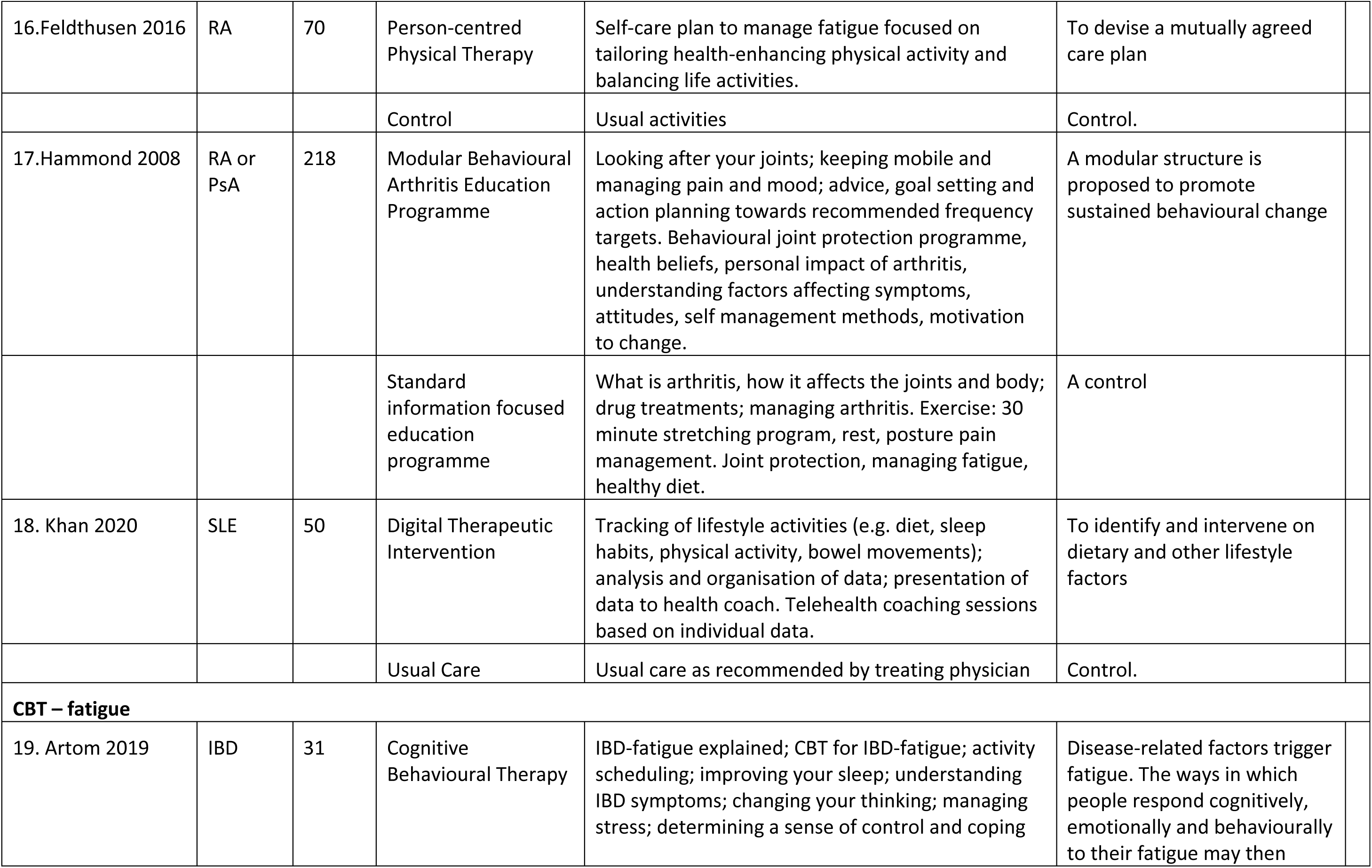
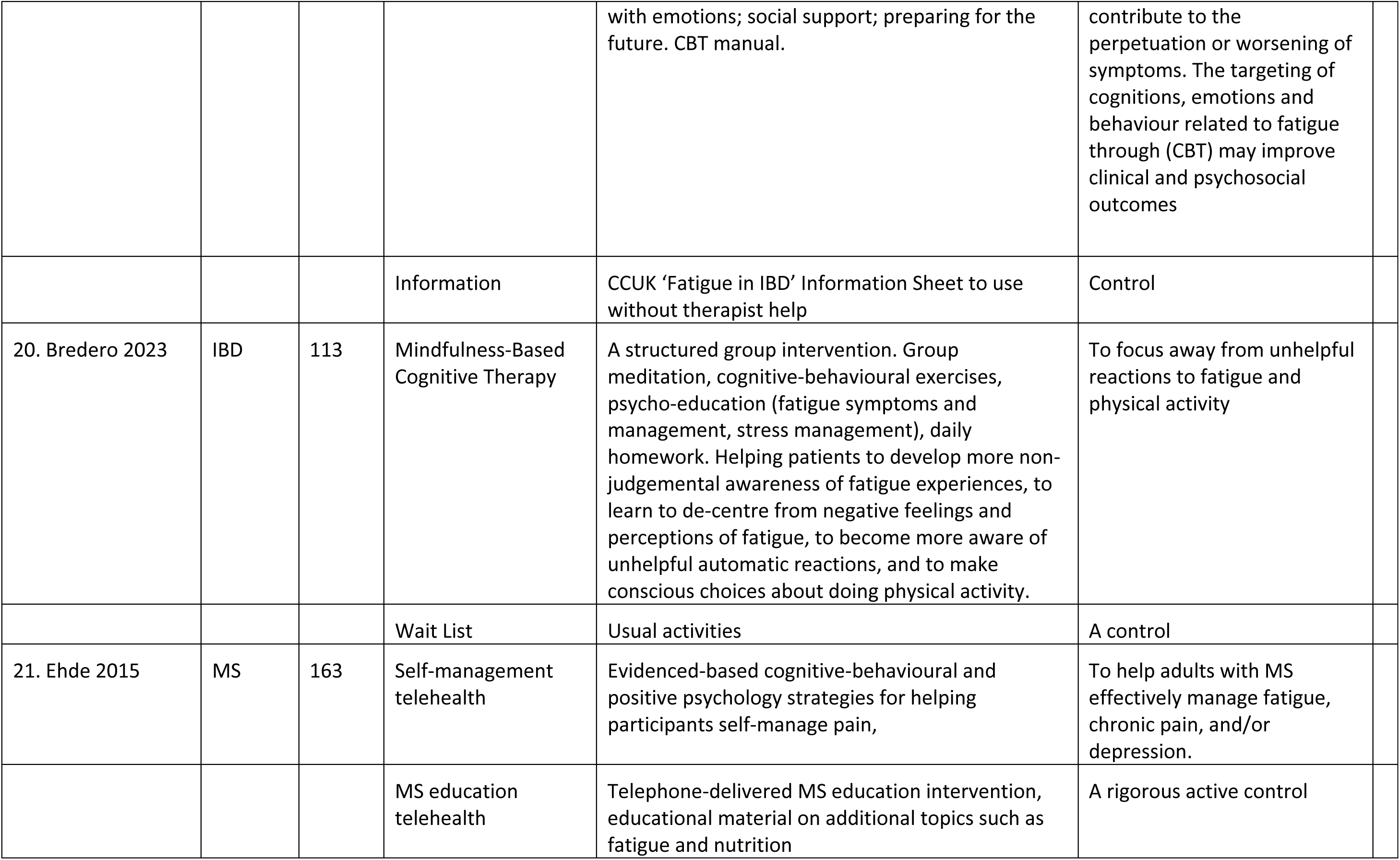
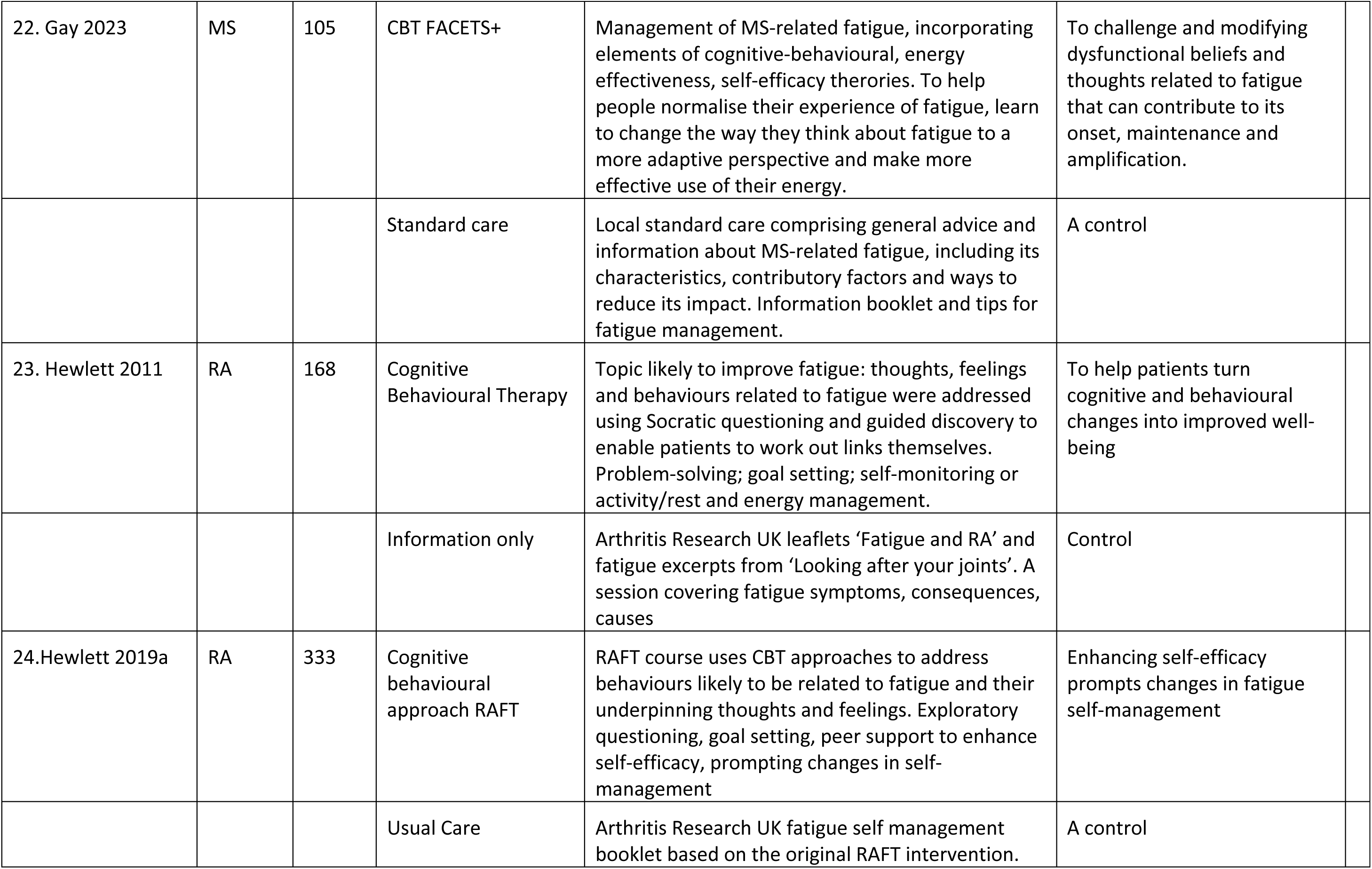
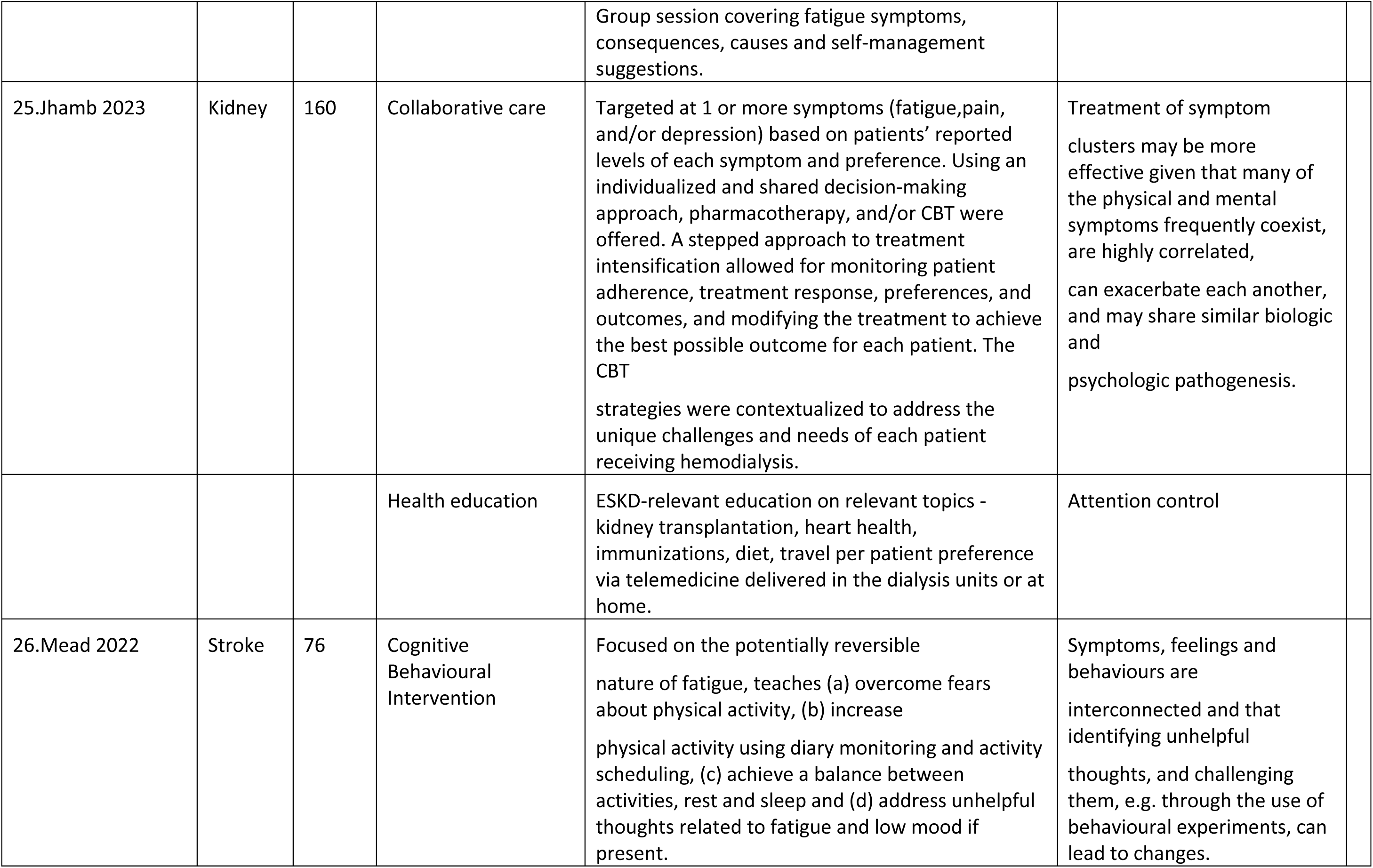
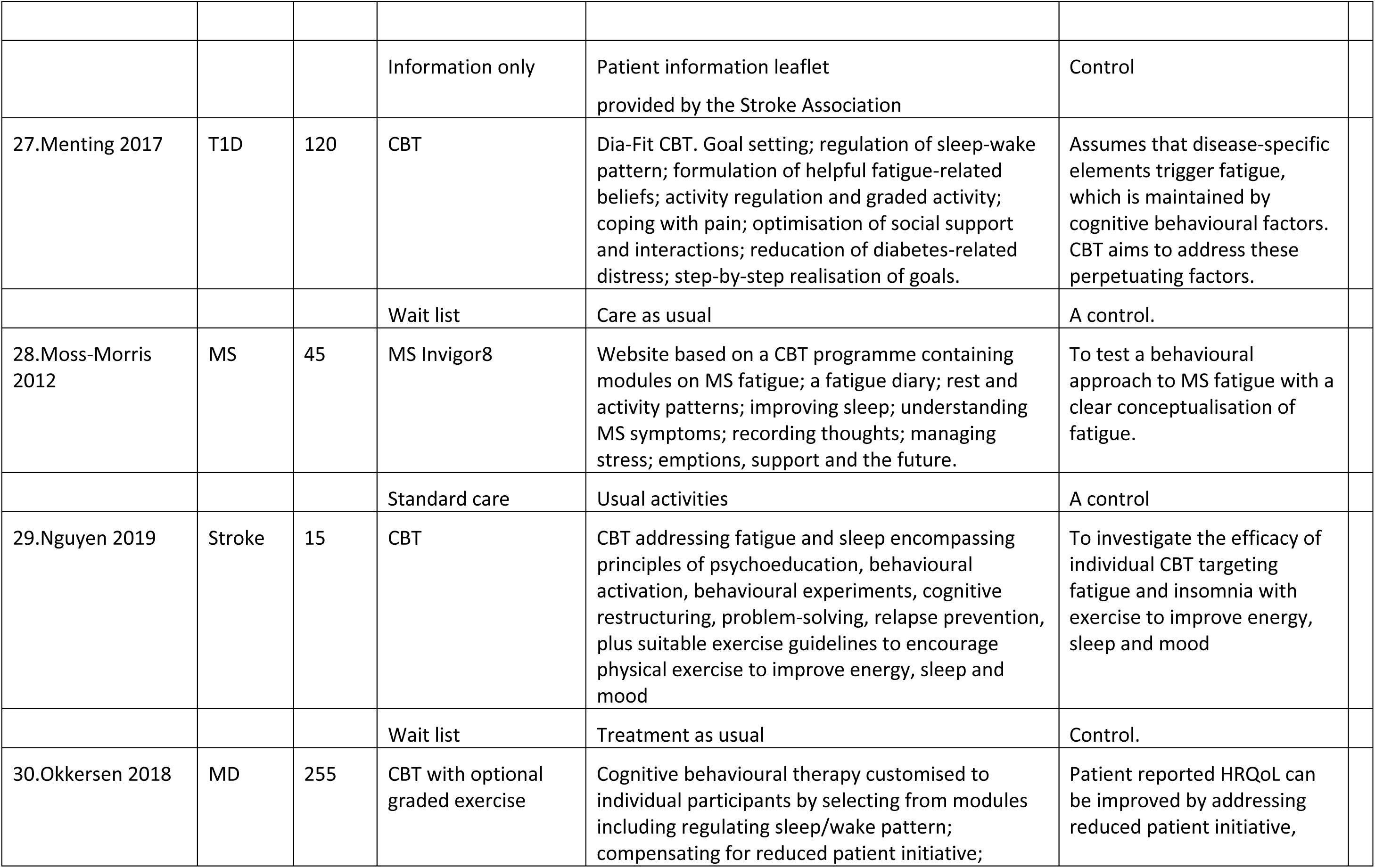
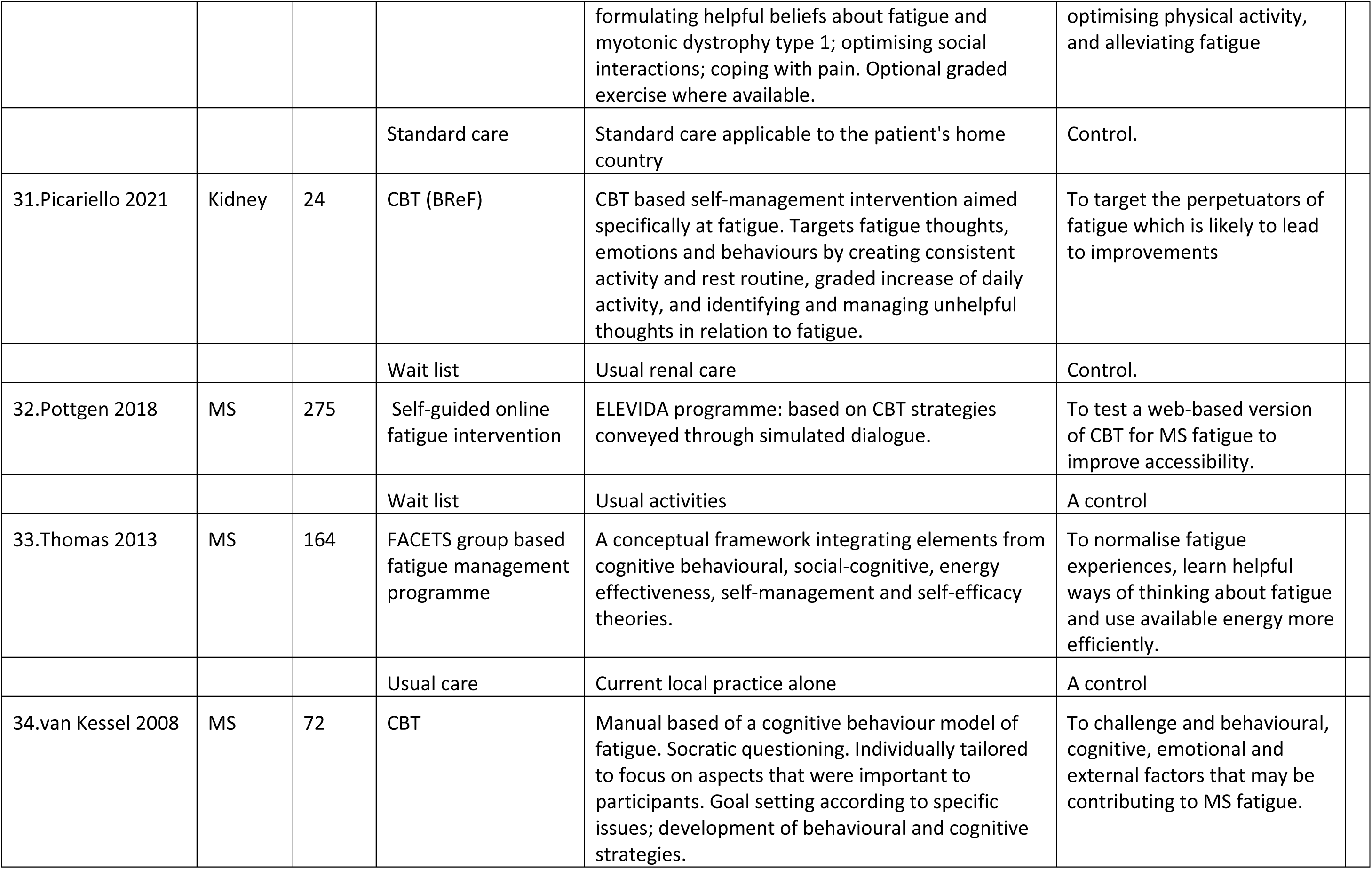
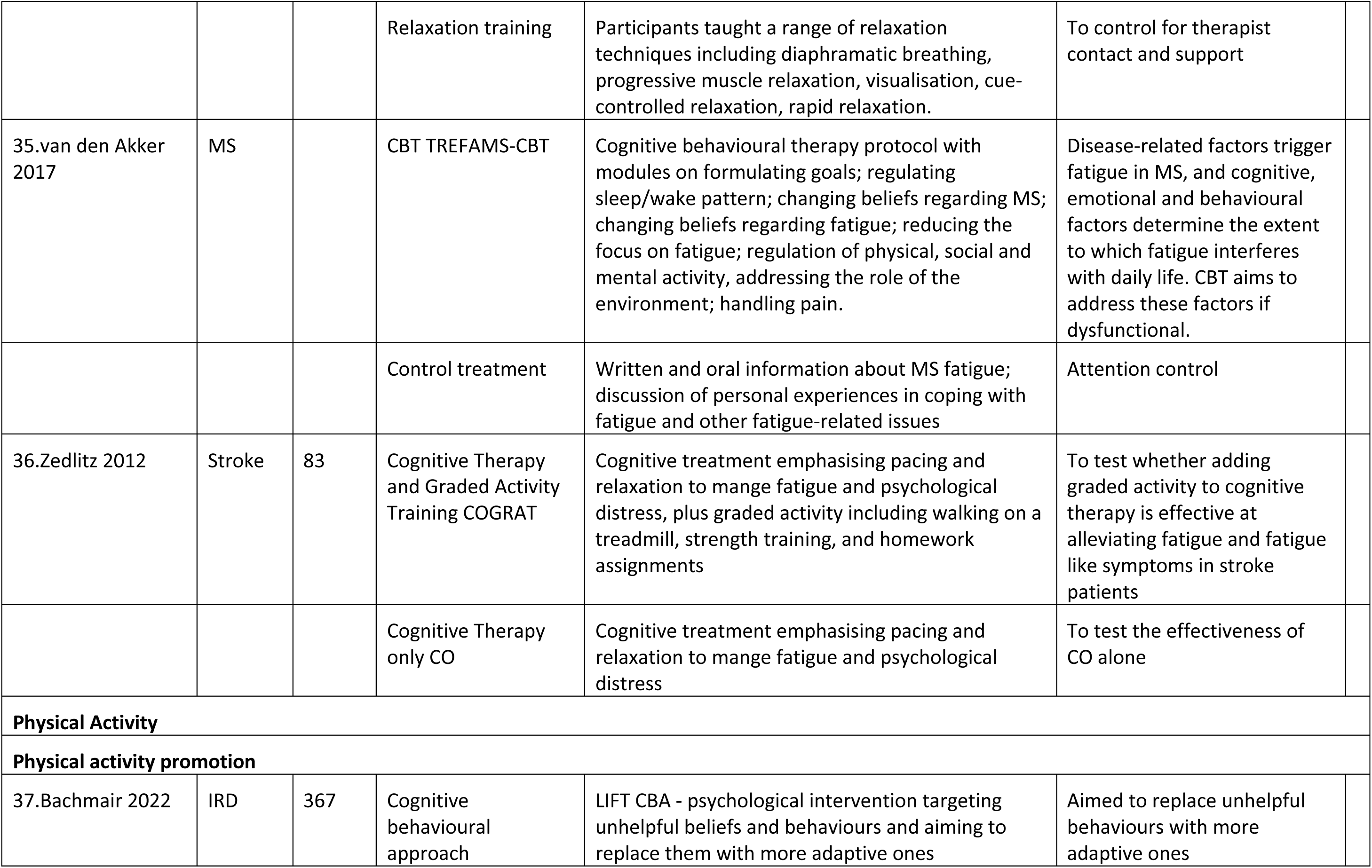
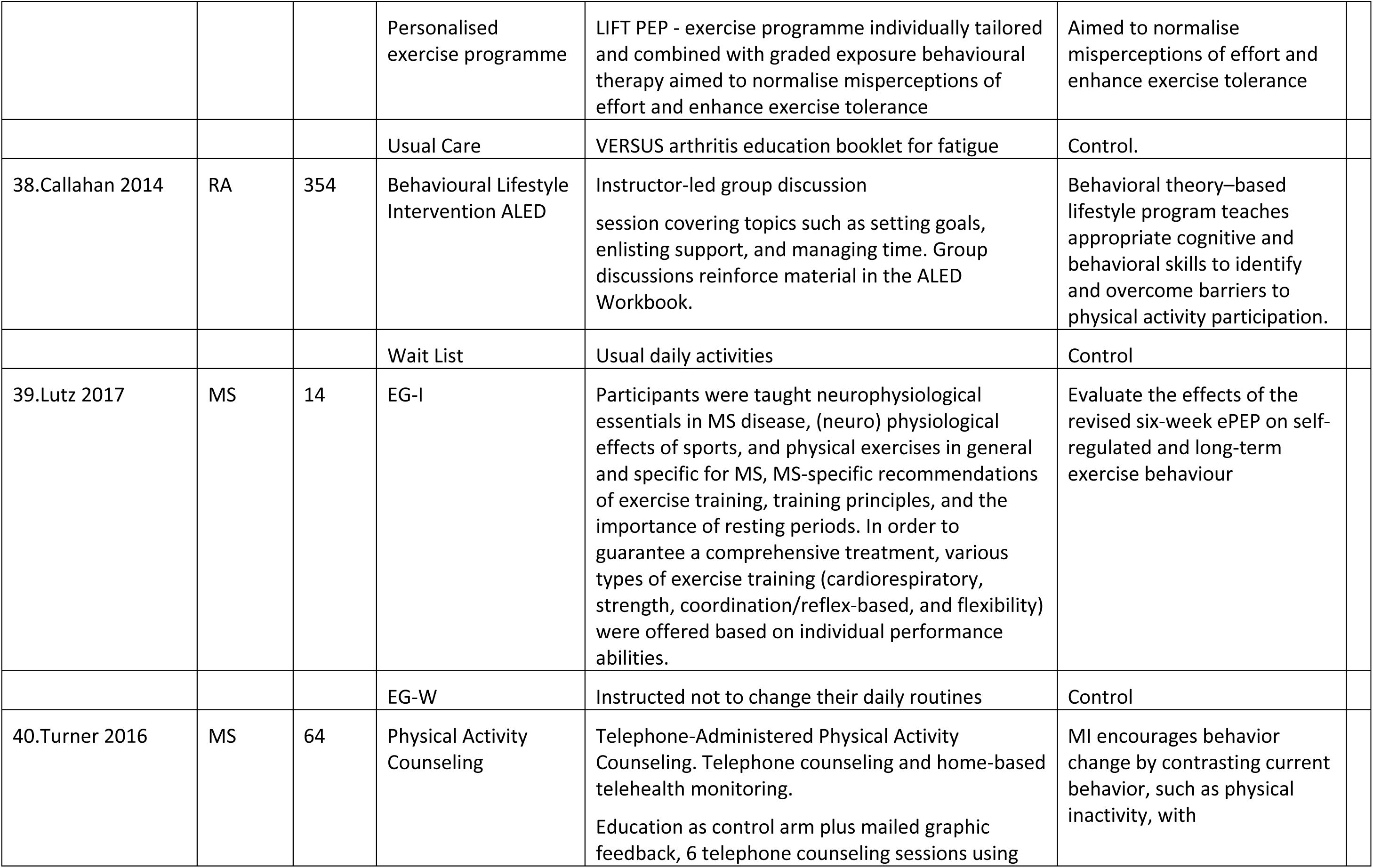
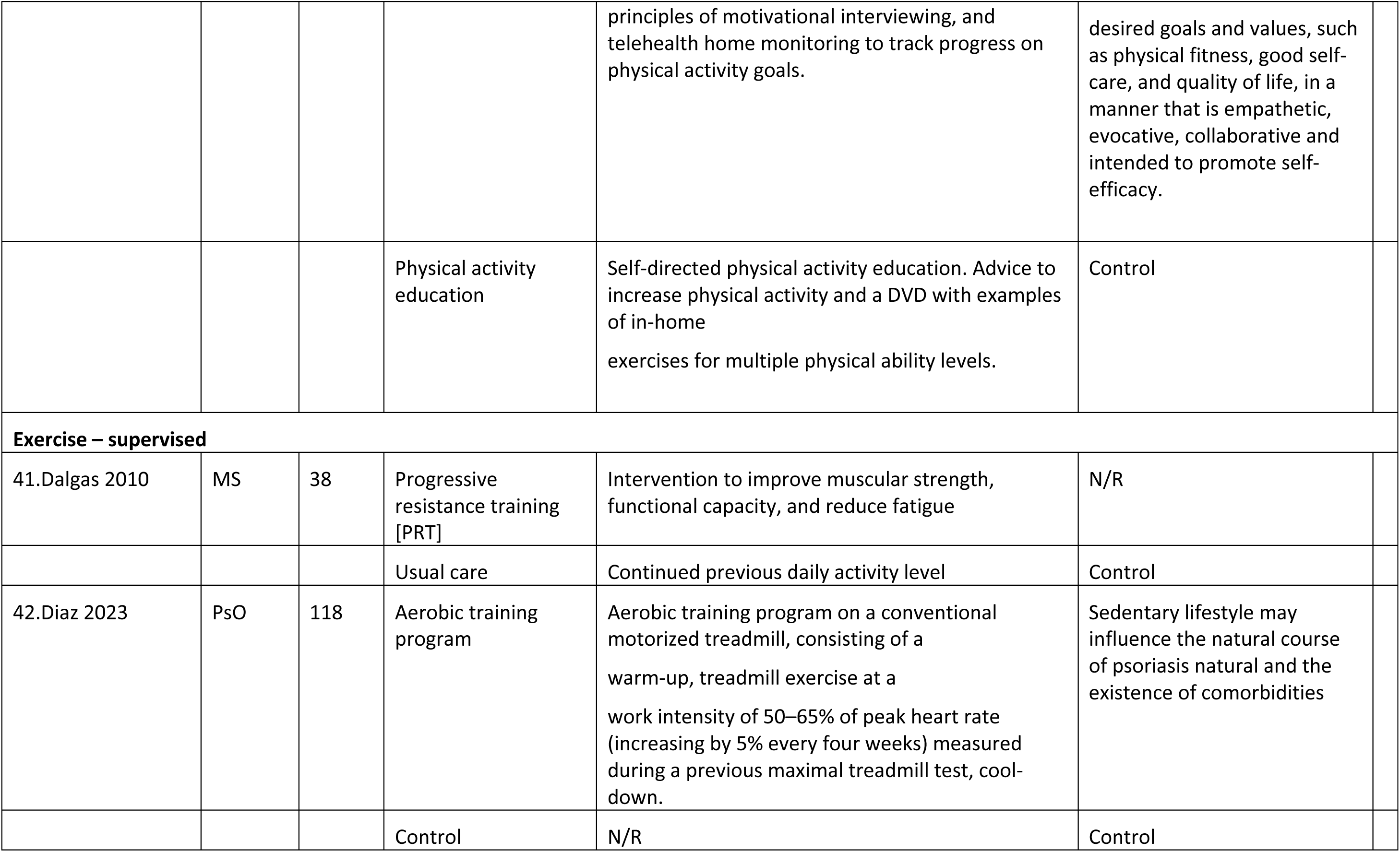
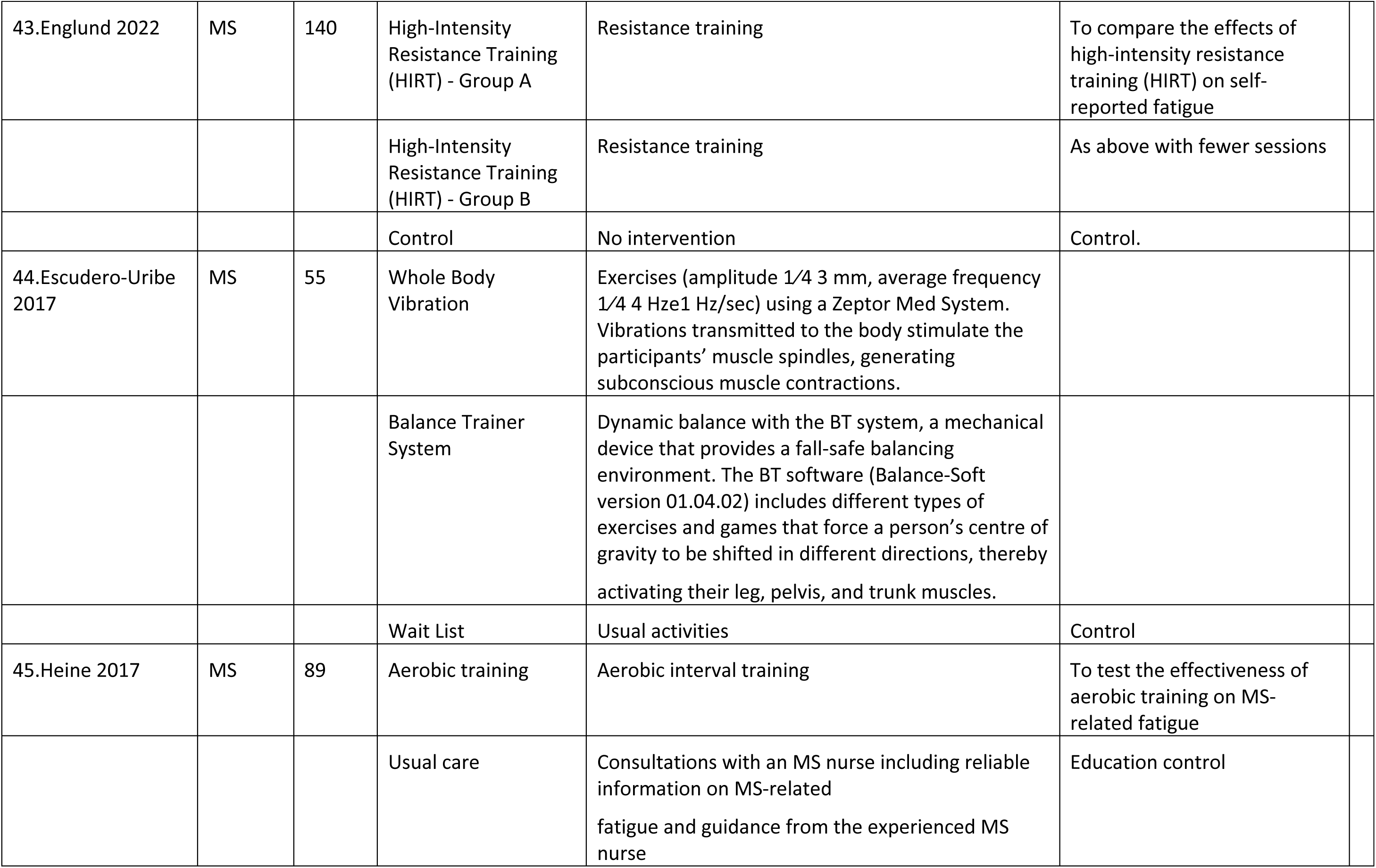
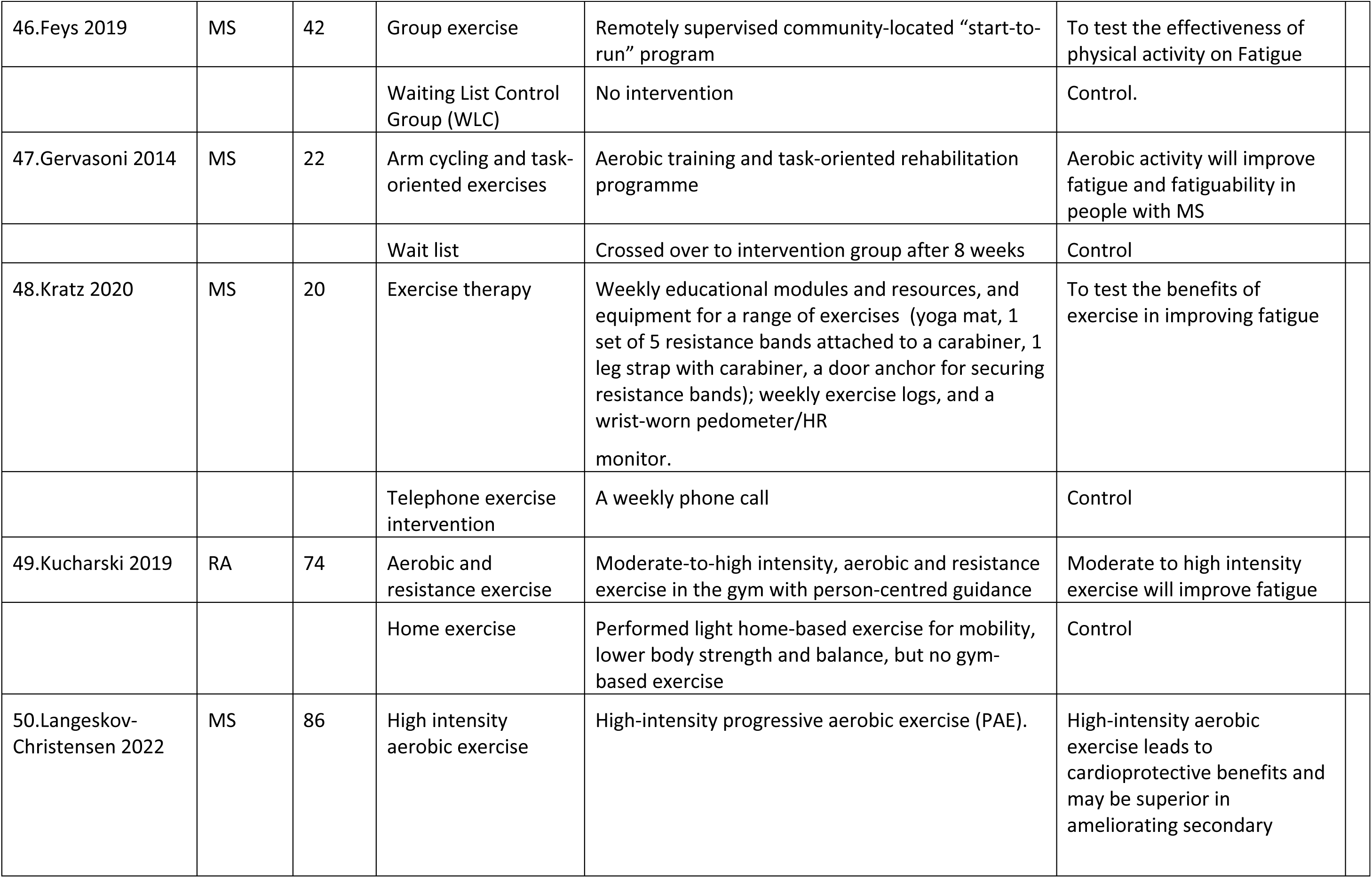
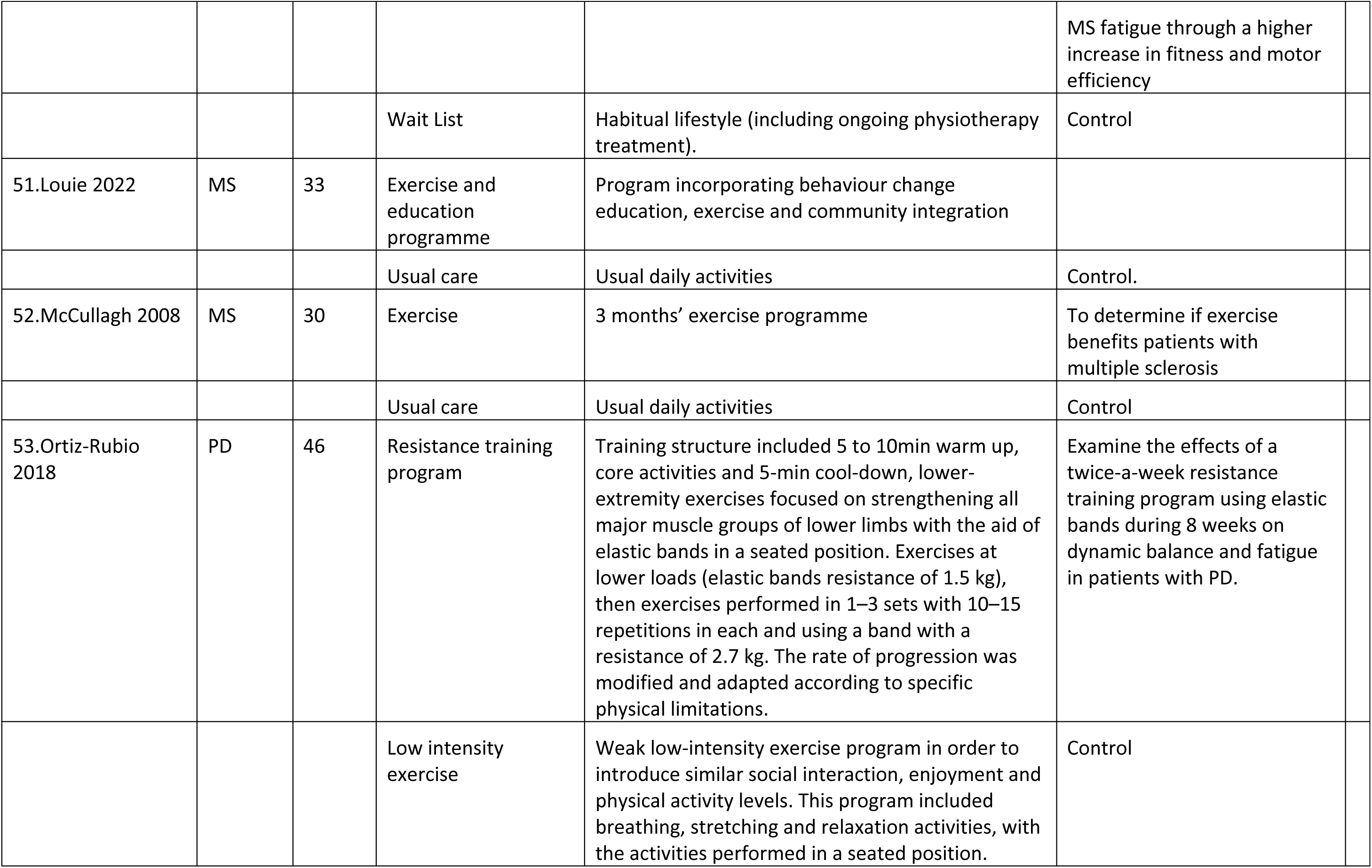
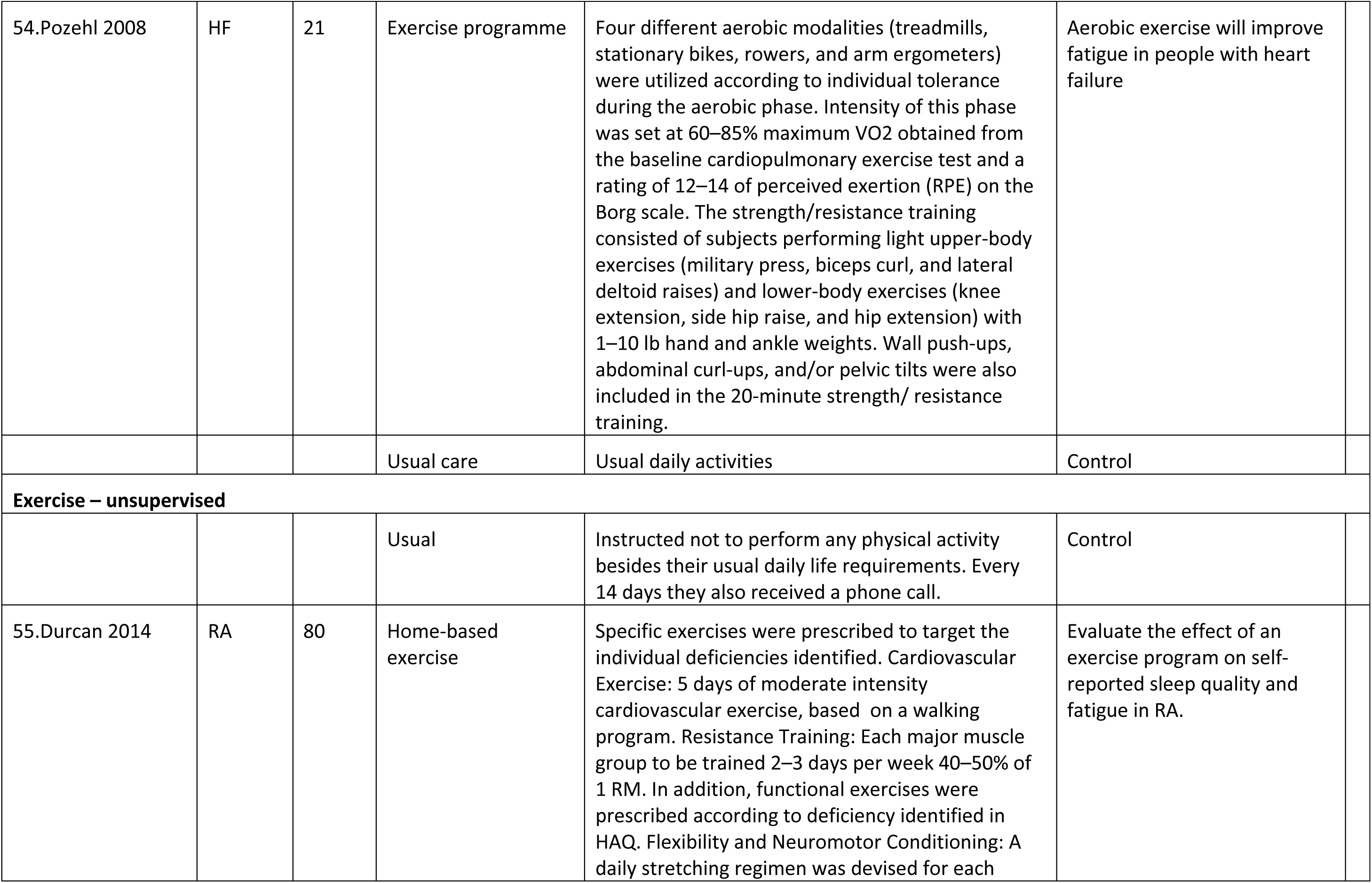
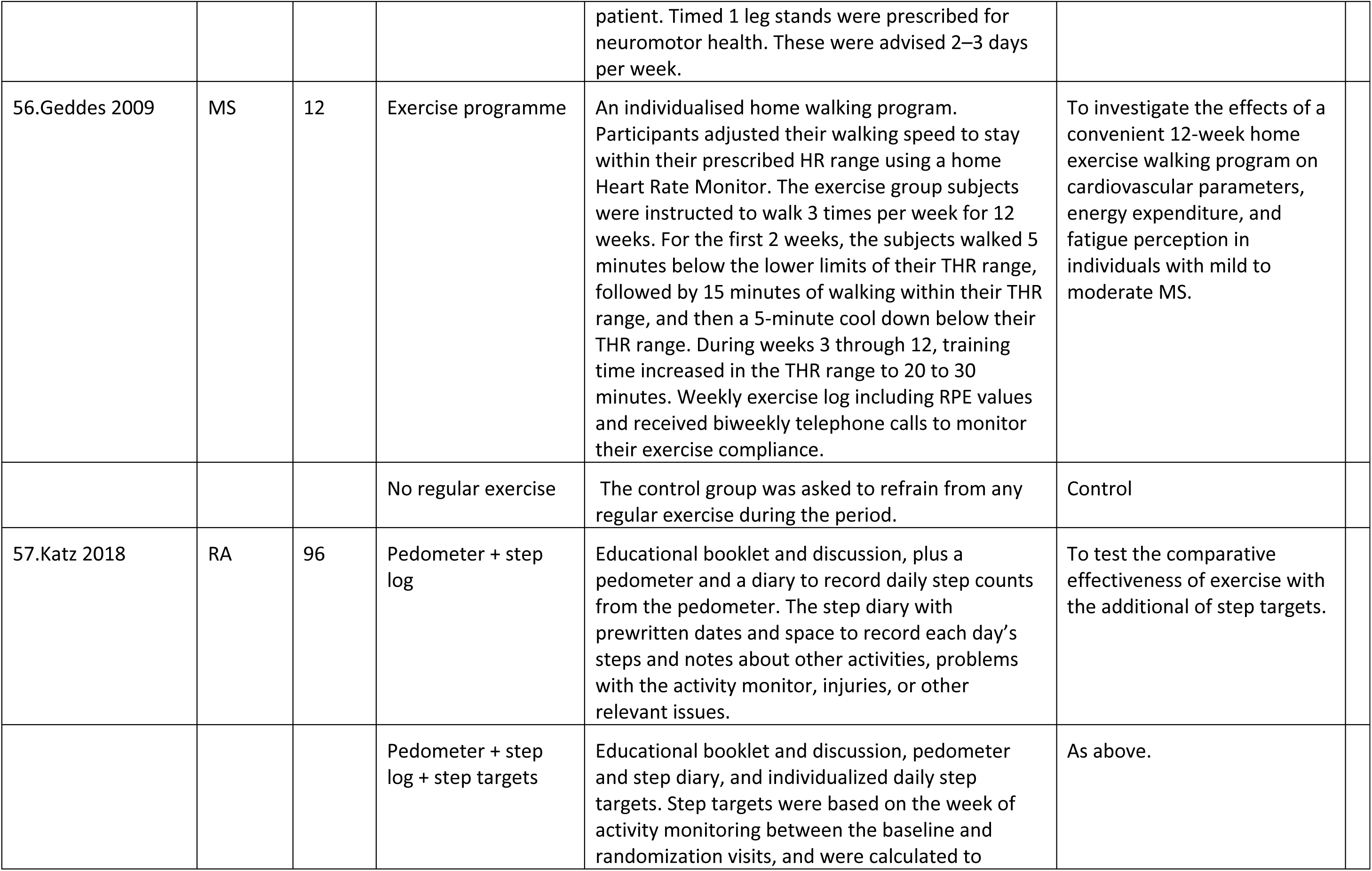
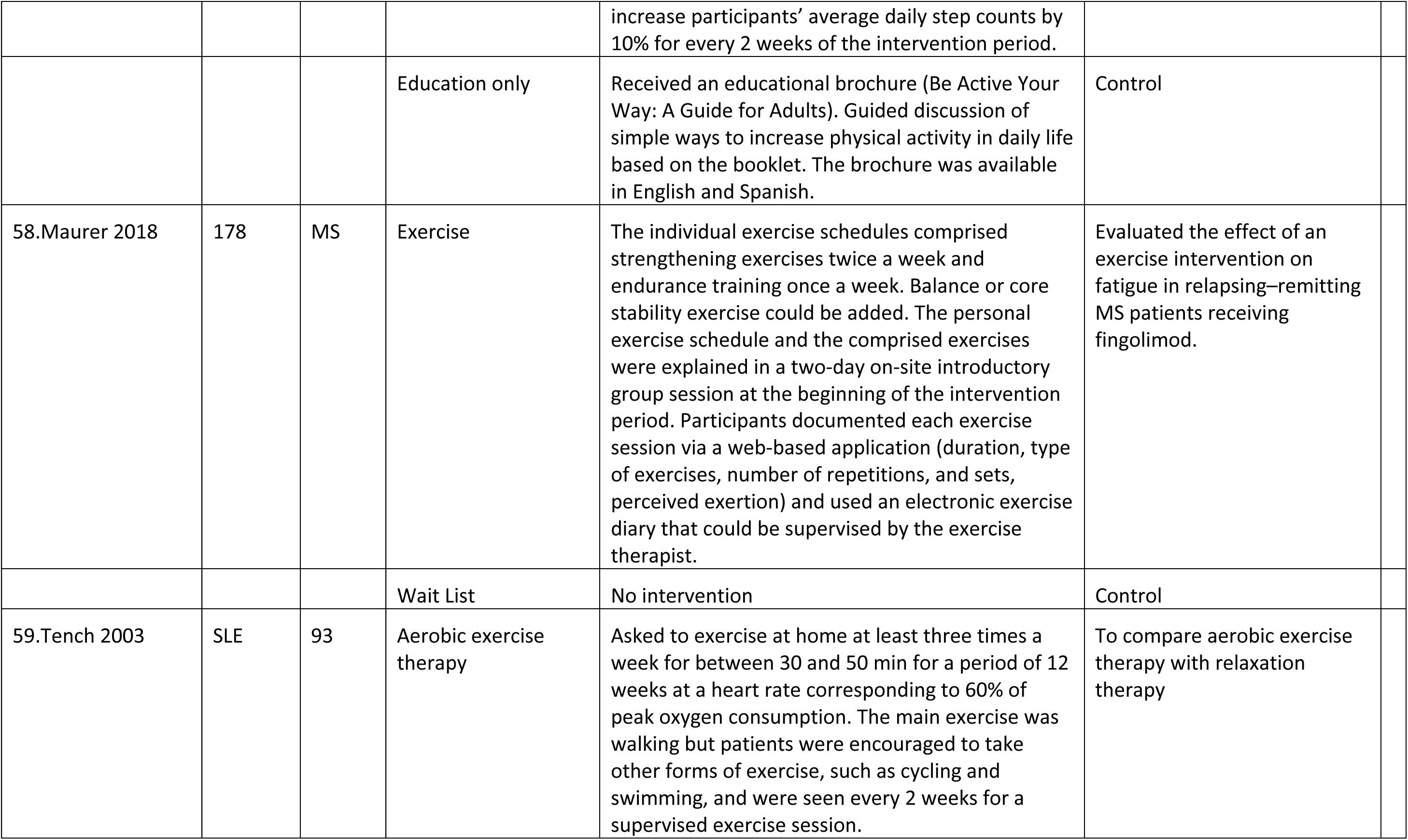
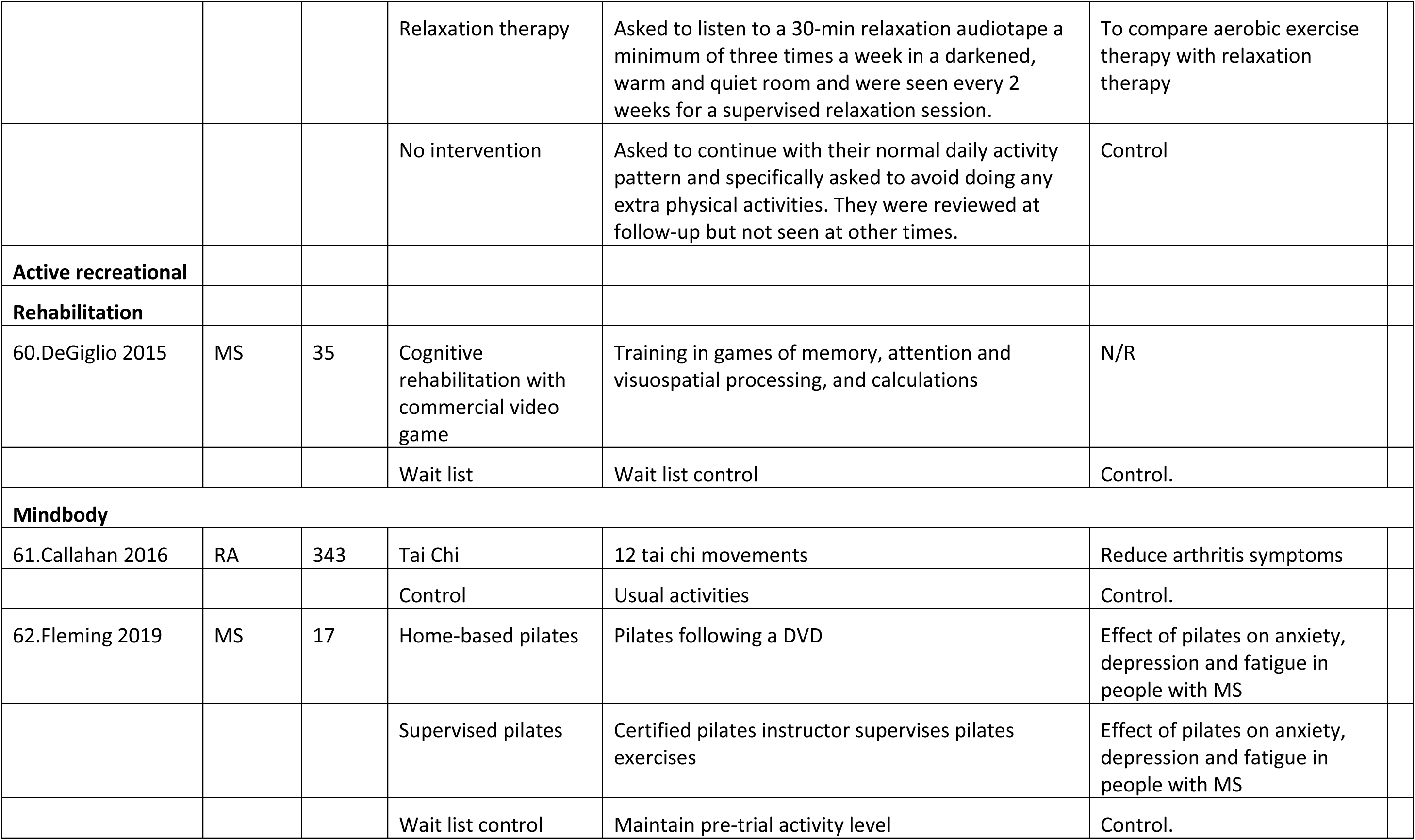
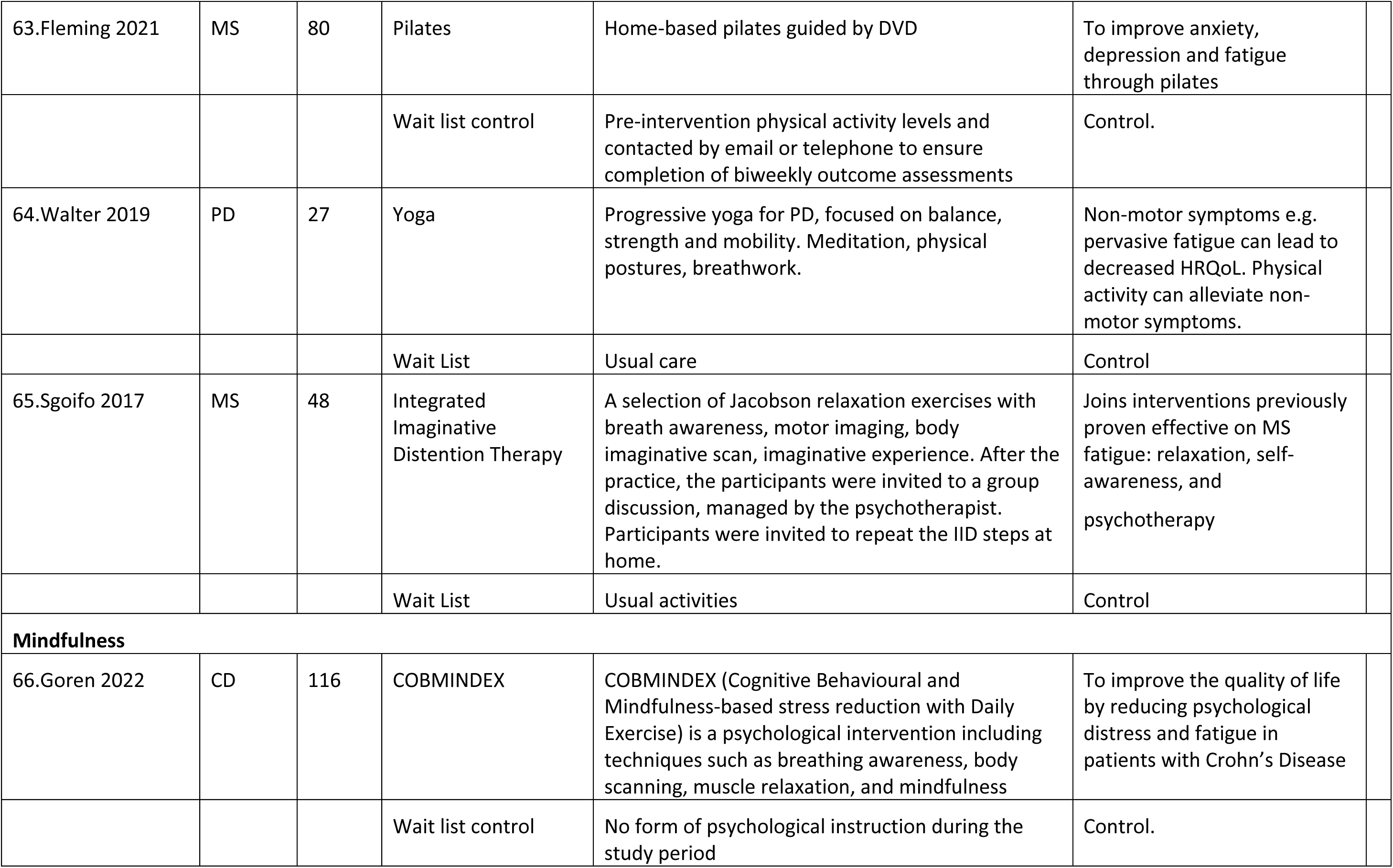
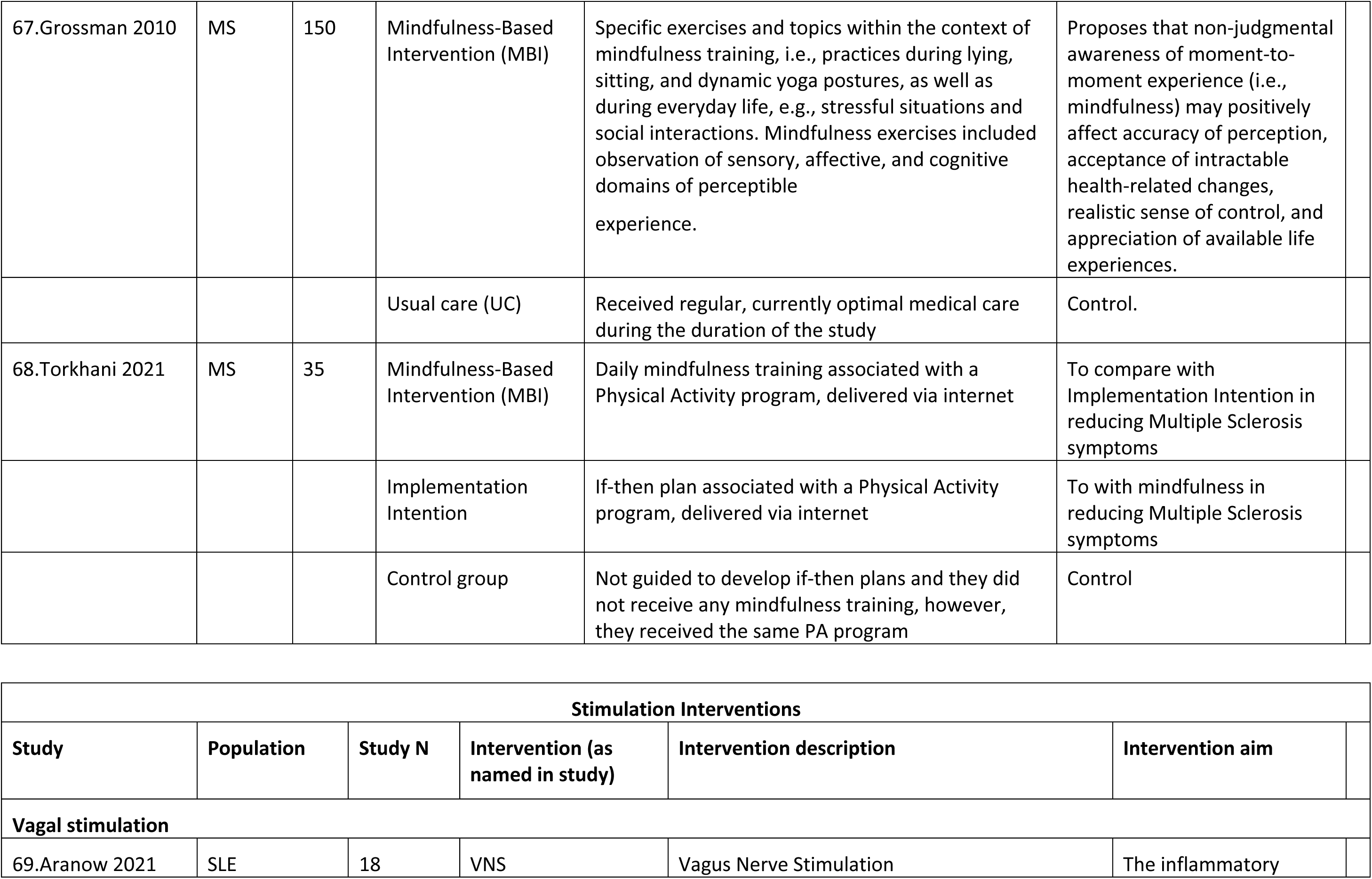
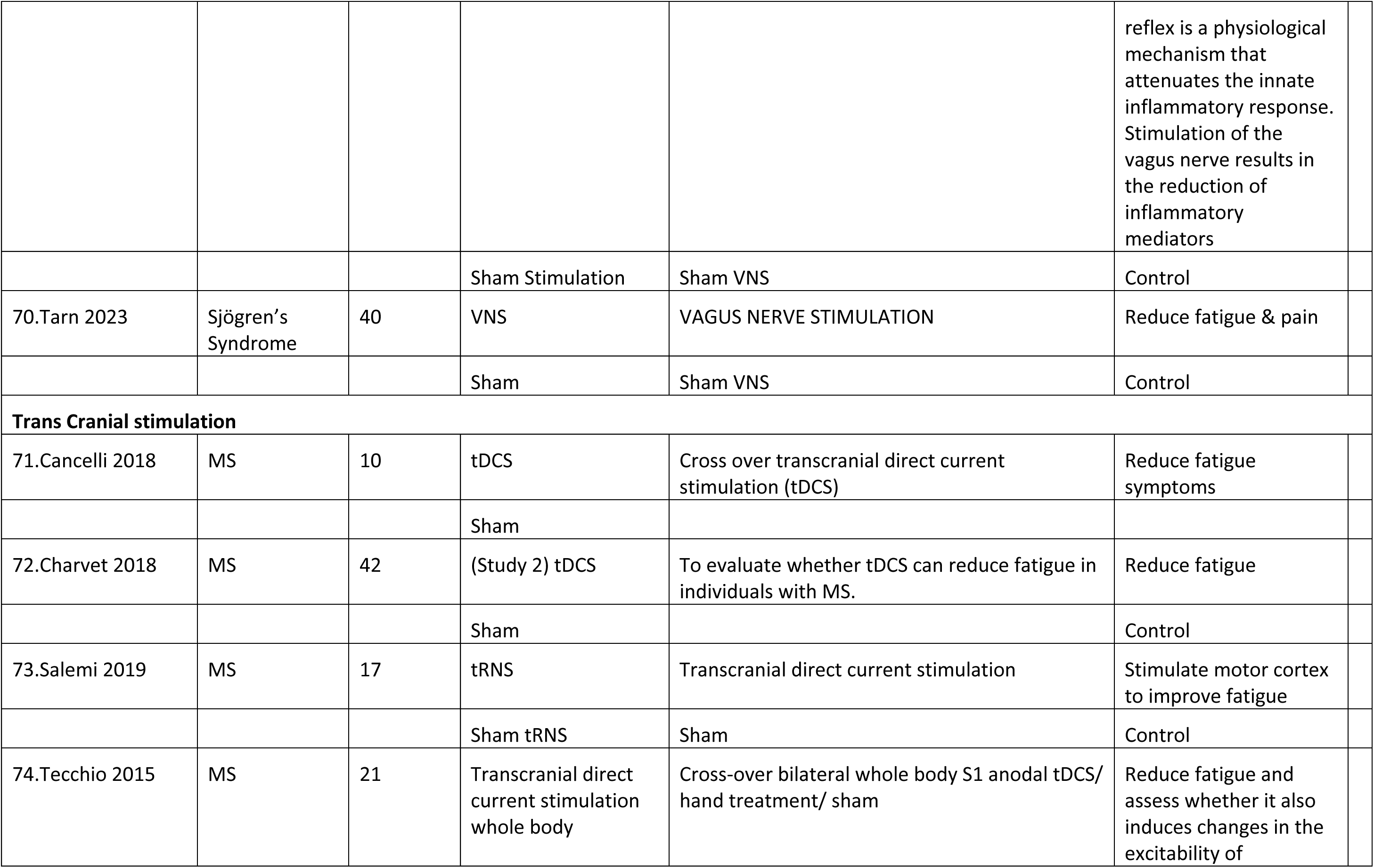
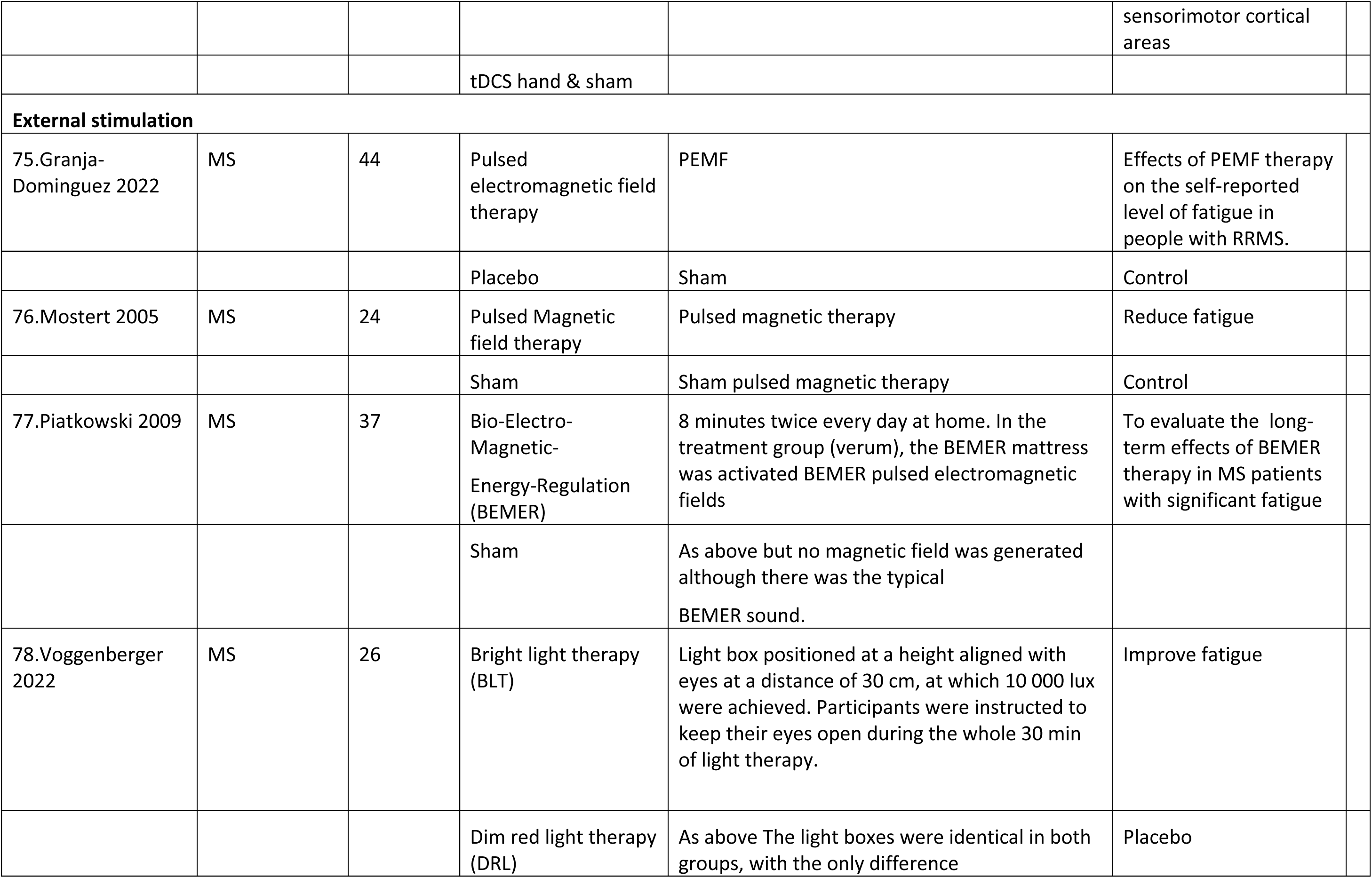
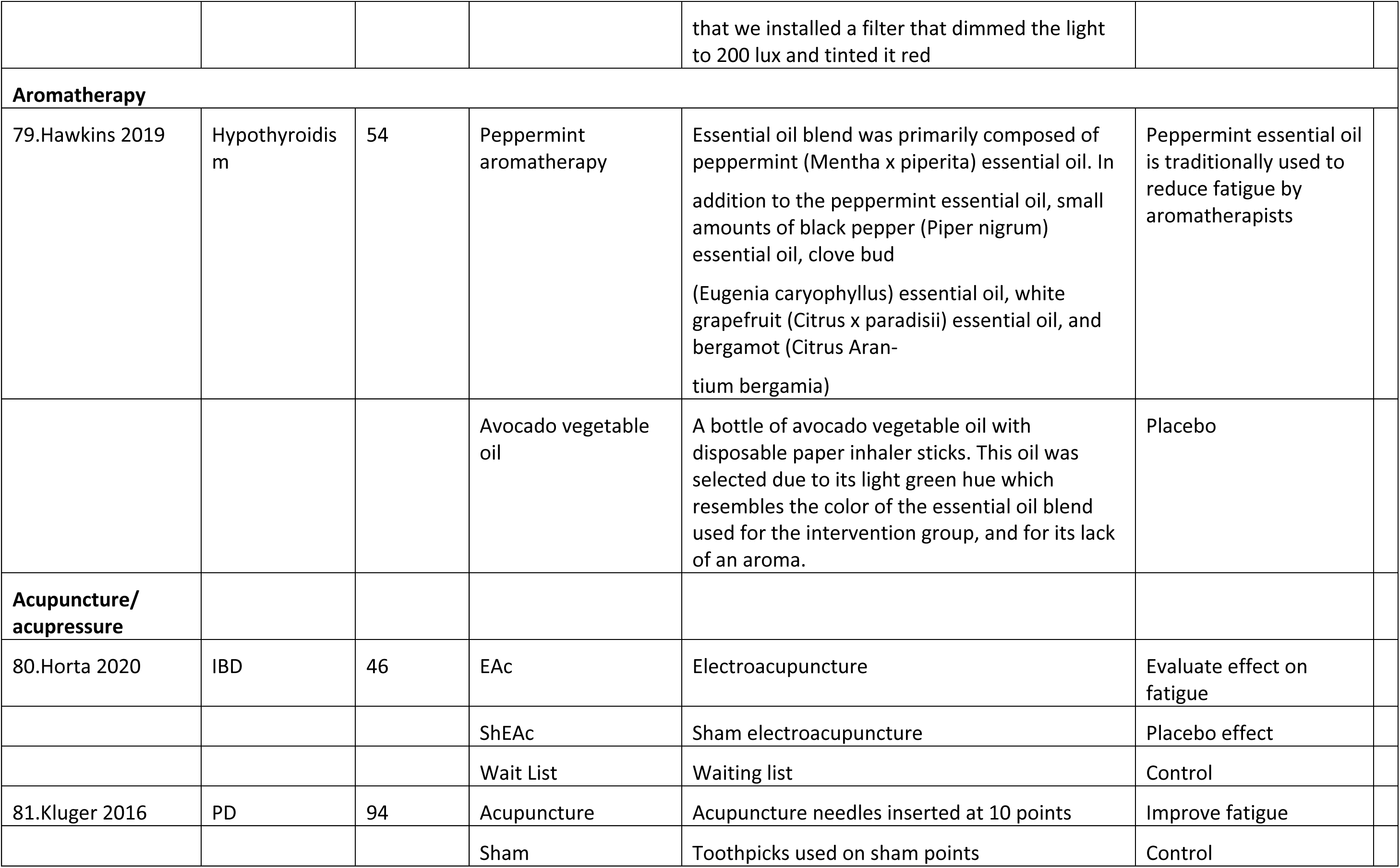
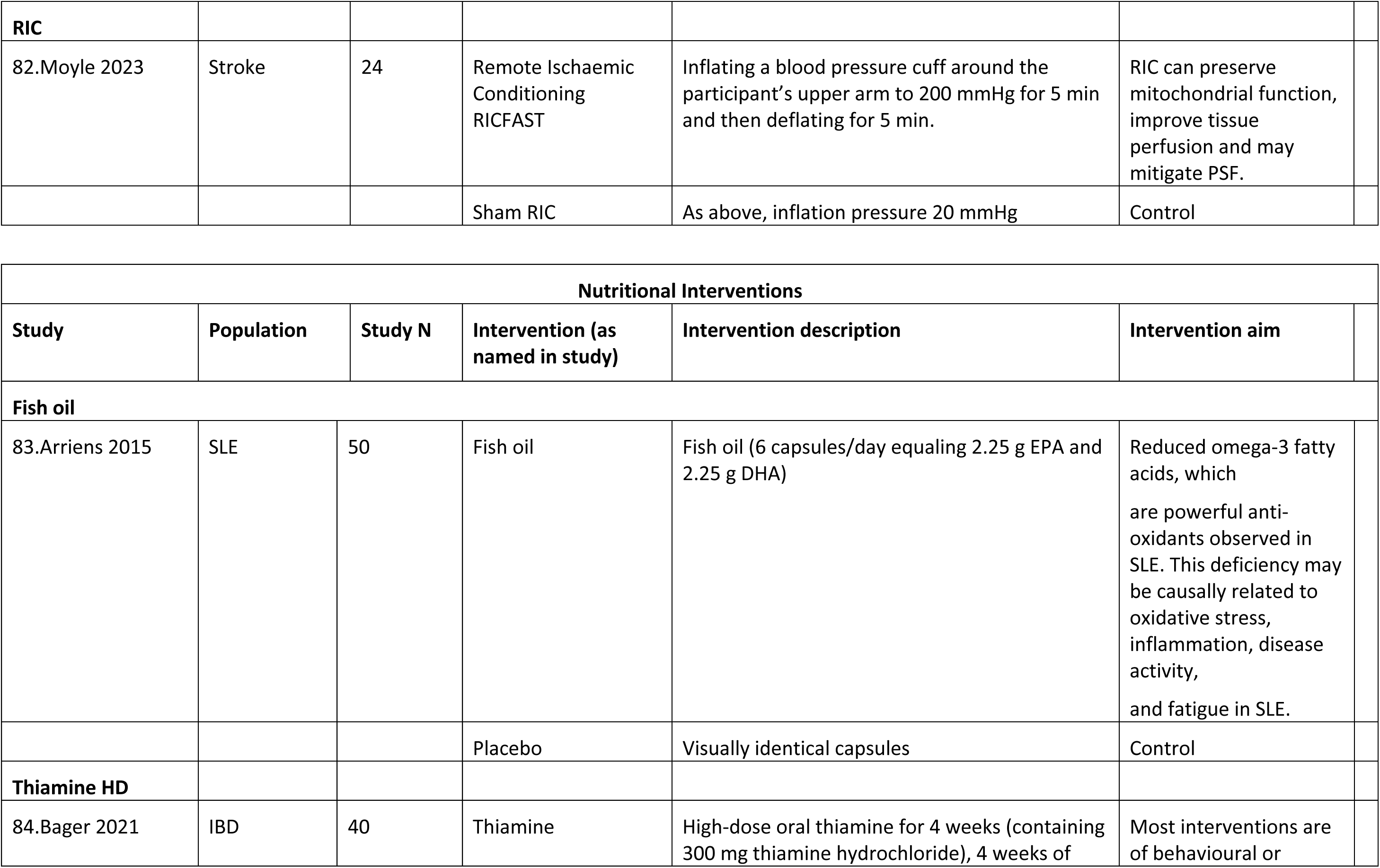
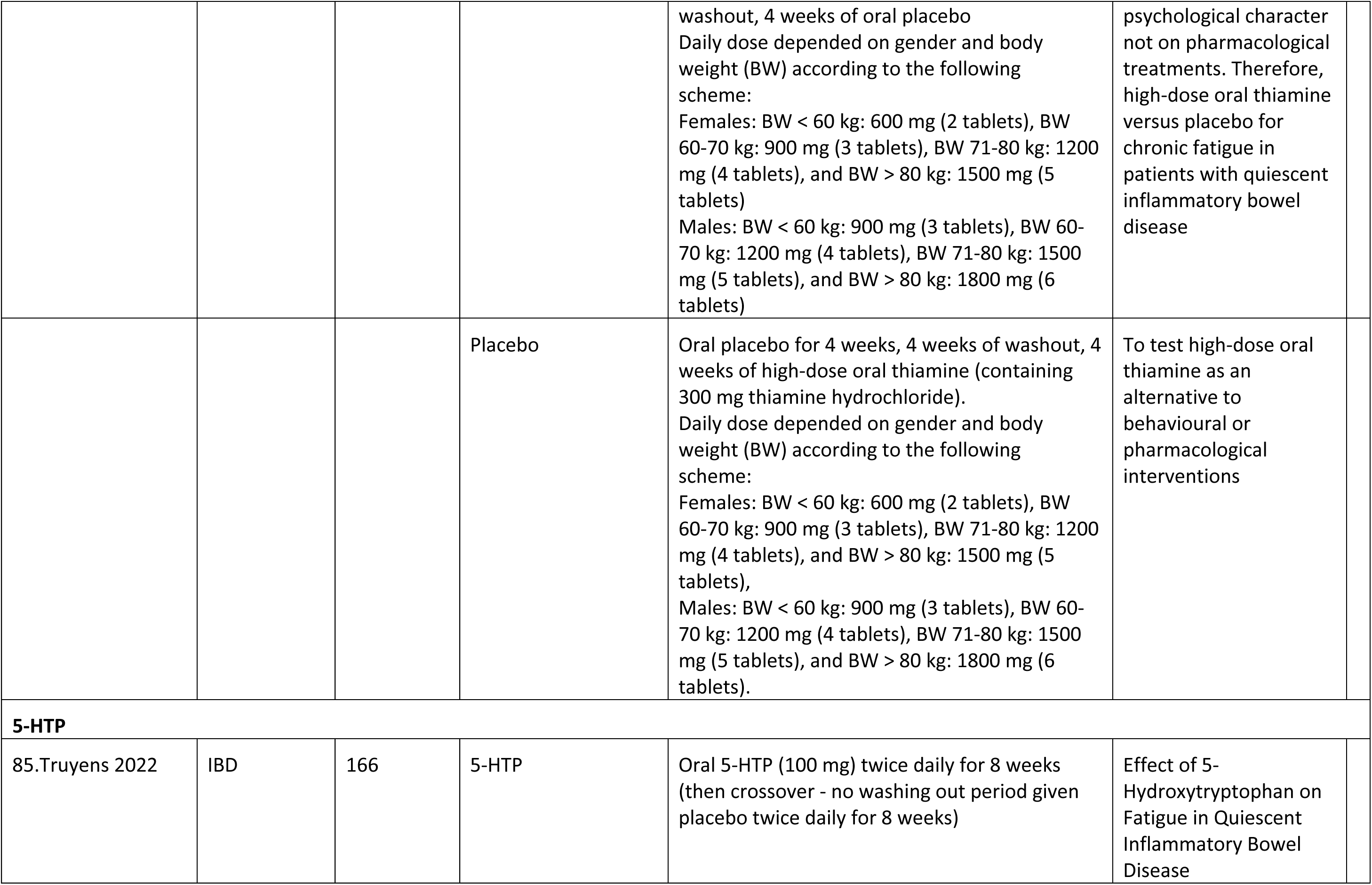
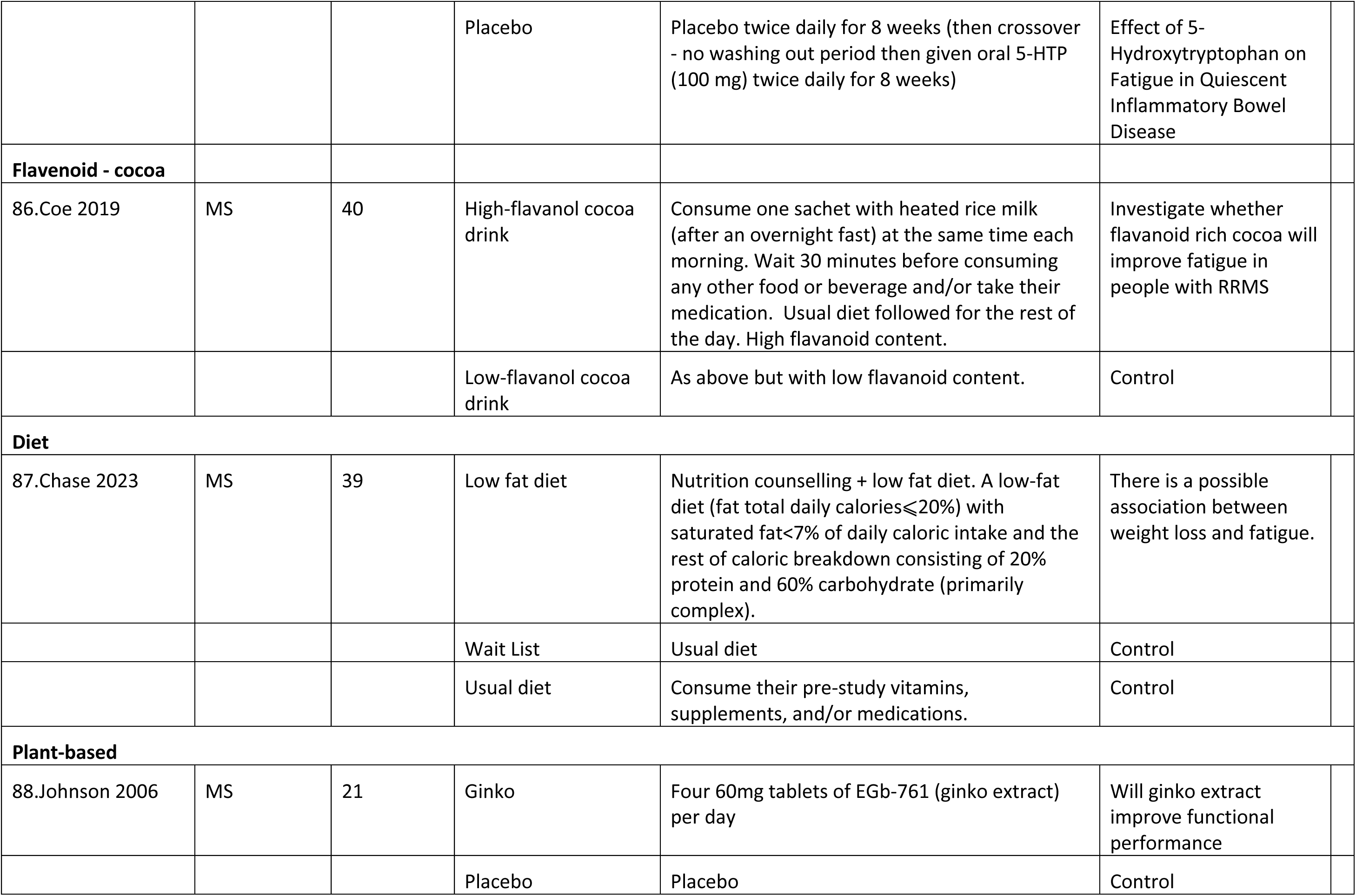

### Supplemental Results 6. Intervention delivery characteristics for studies in the NMA

#### 6.1 Behavioural Interventions

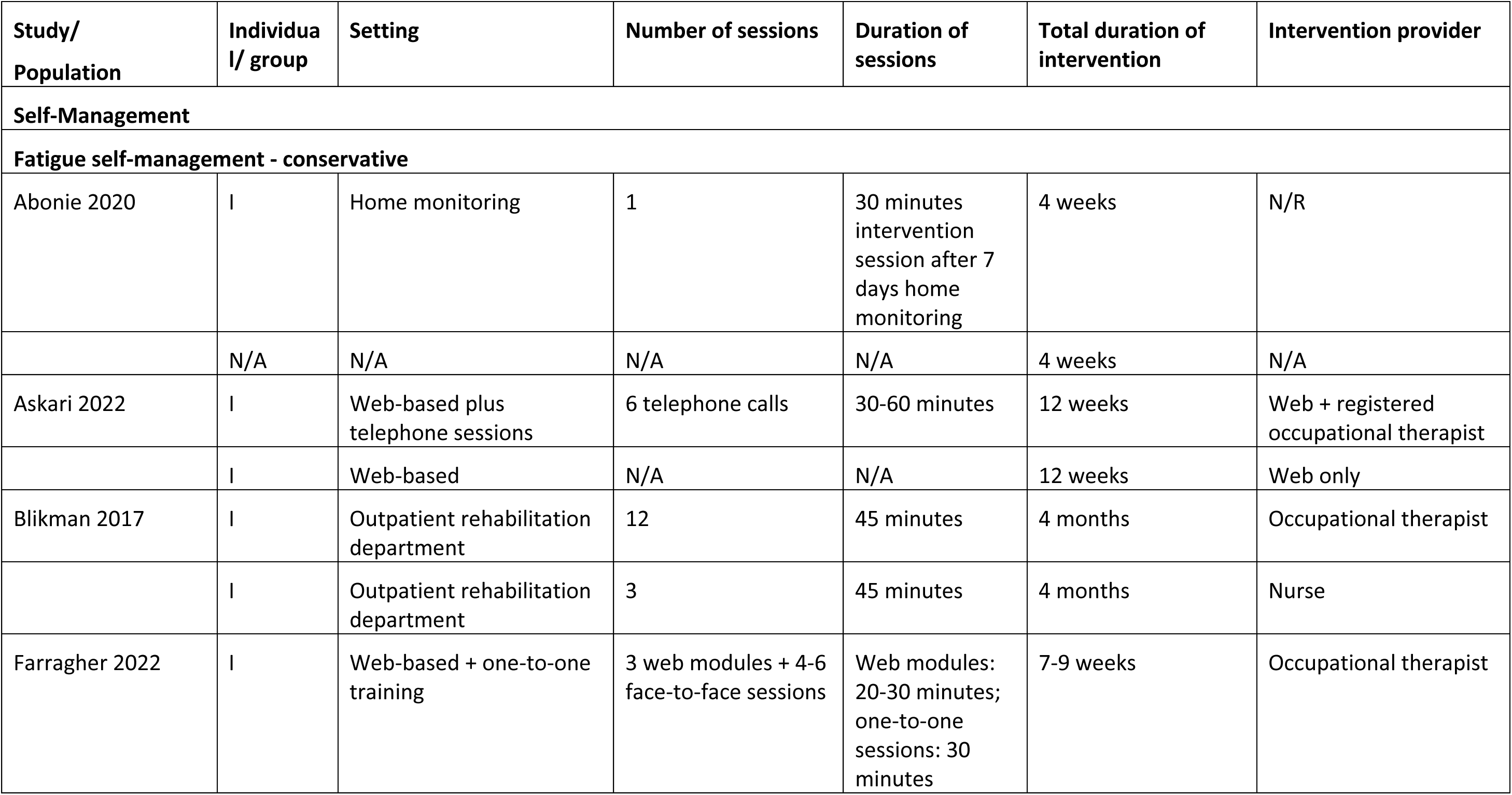
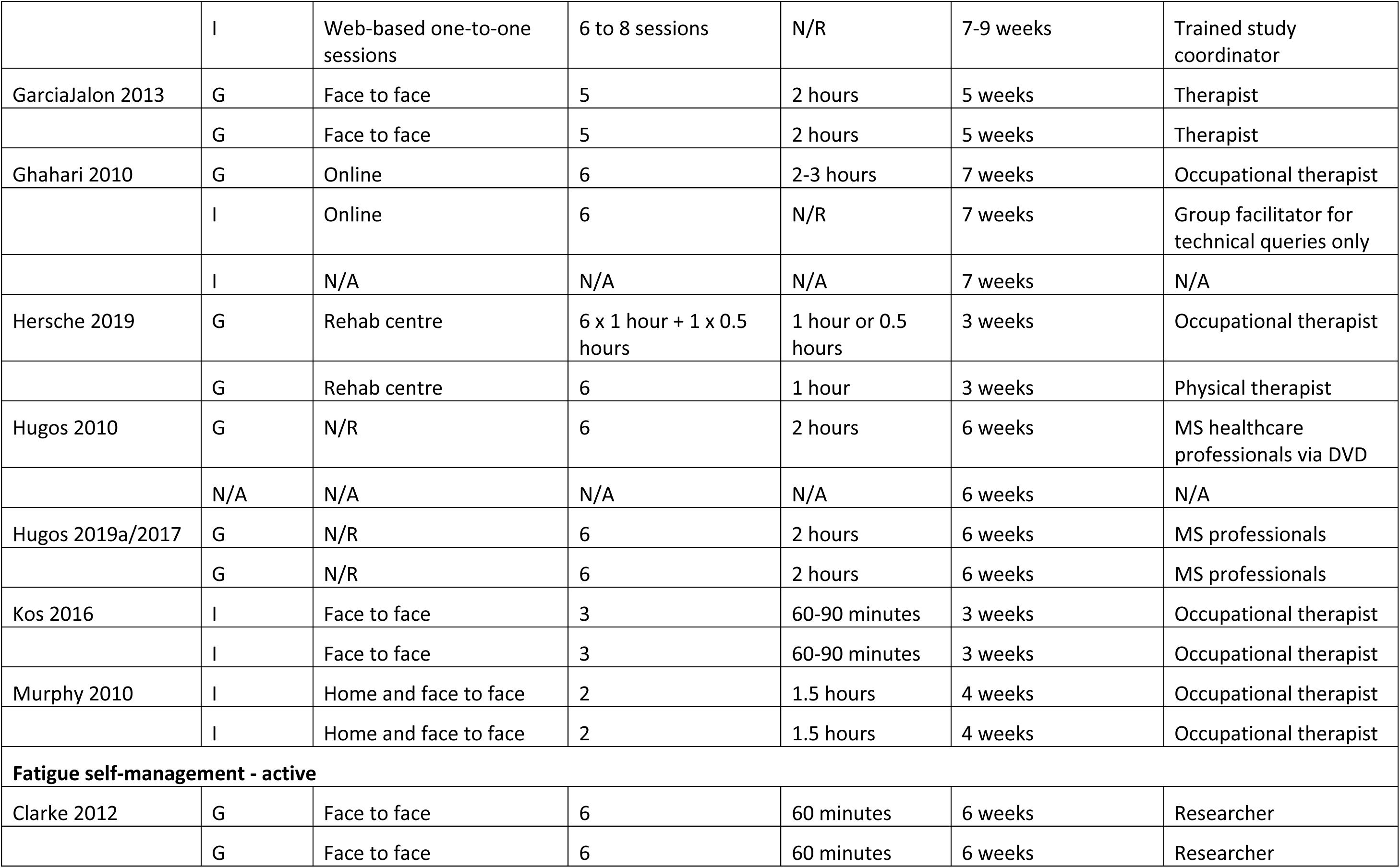
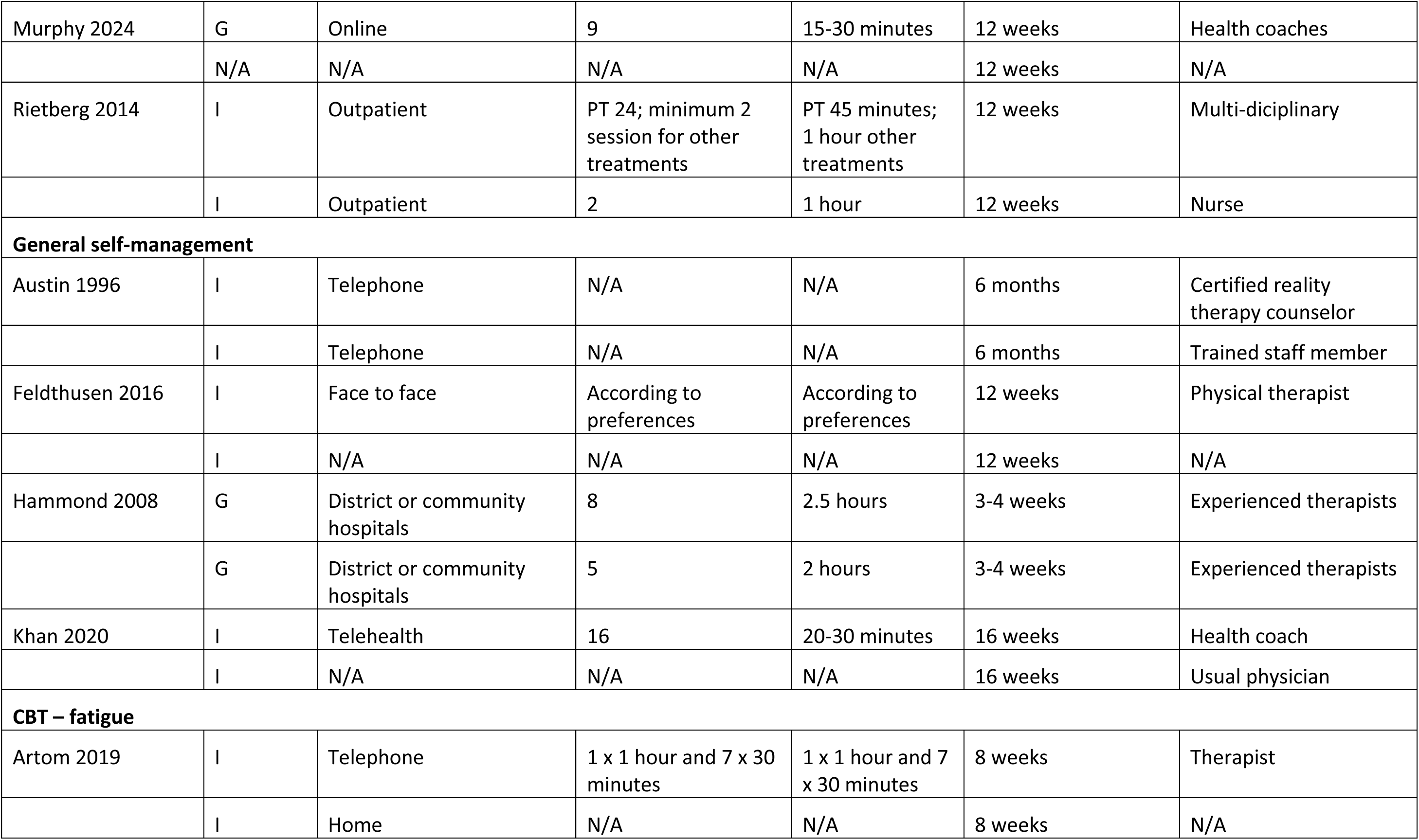
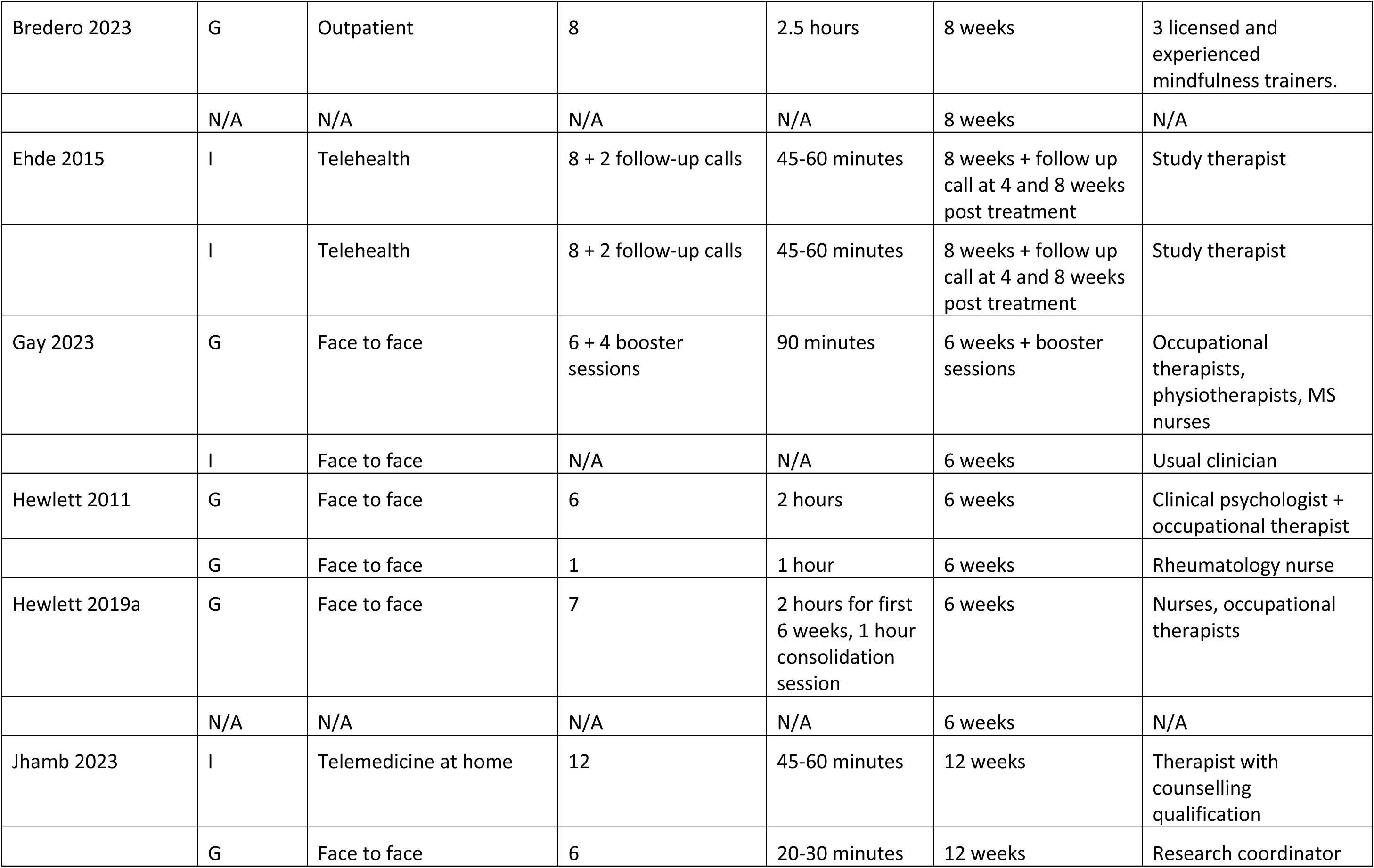
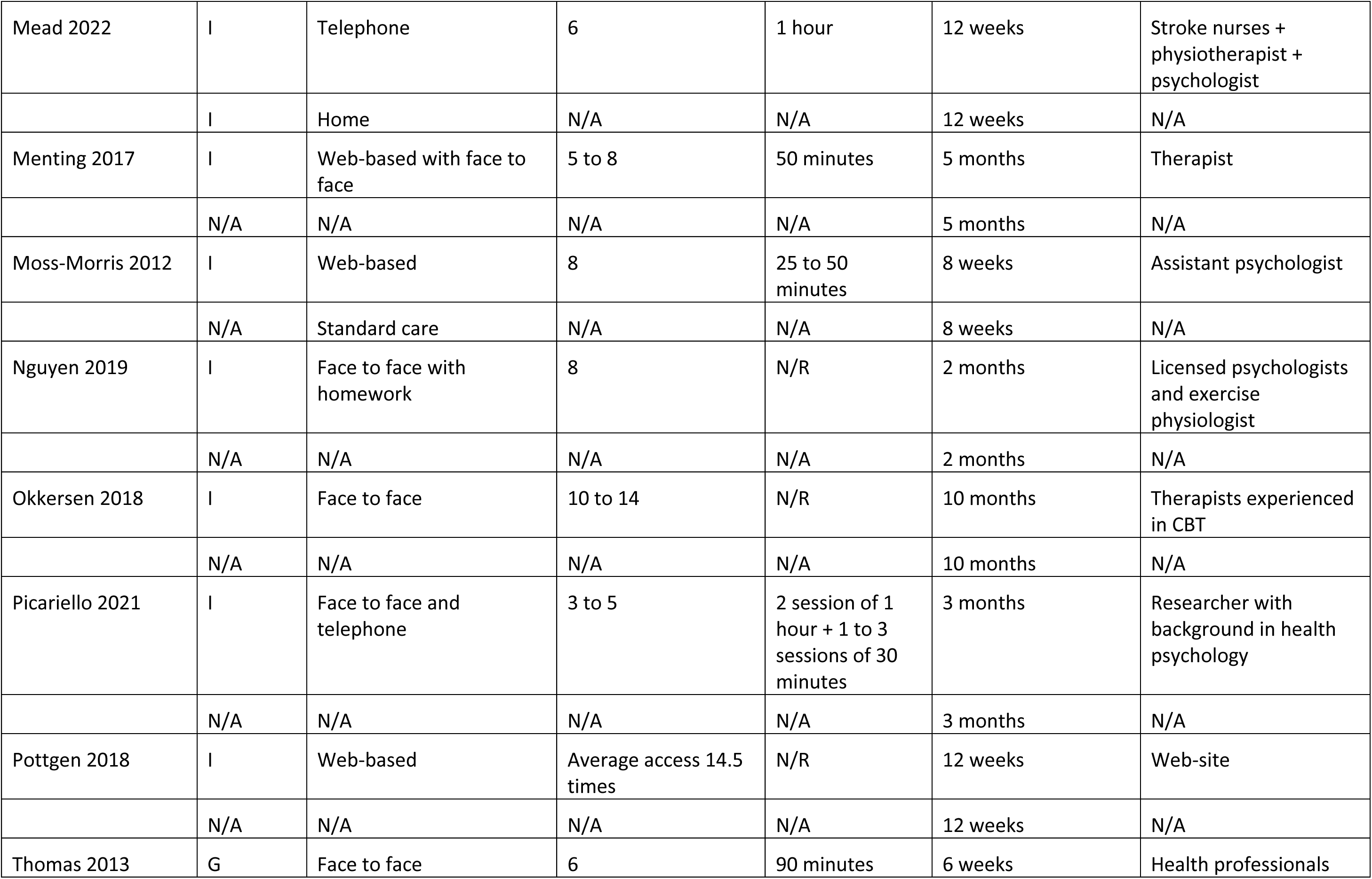
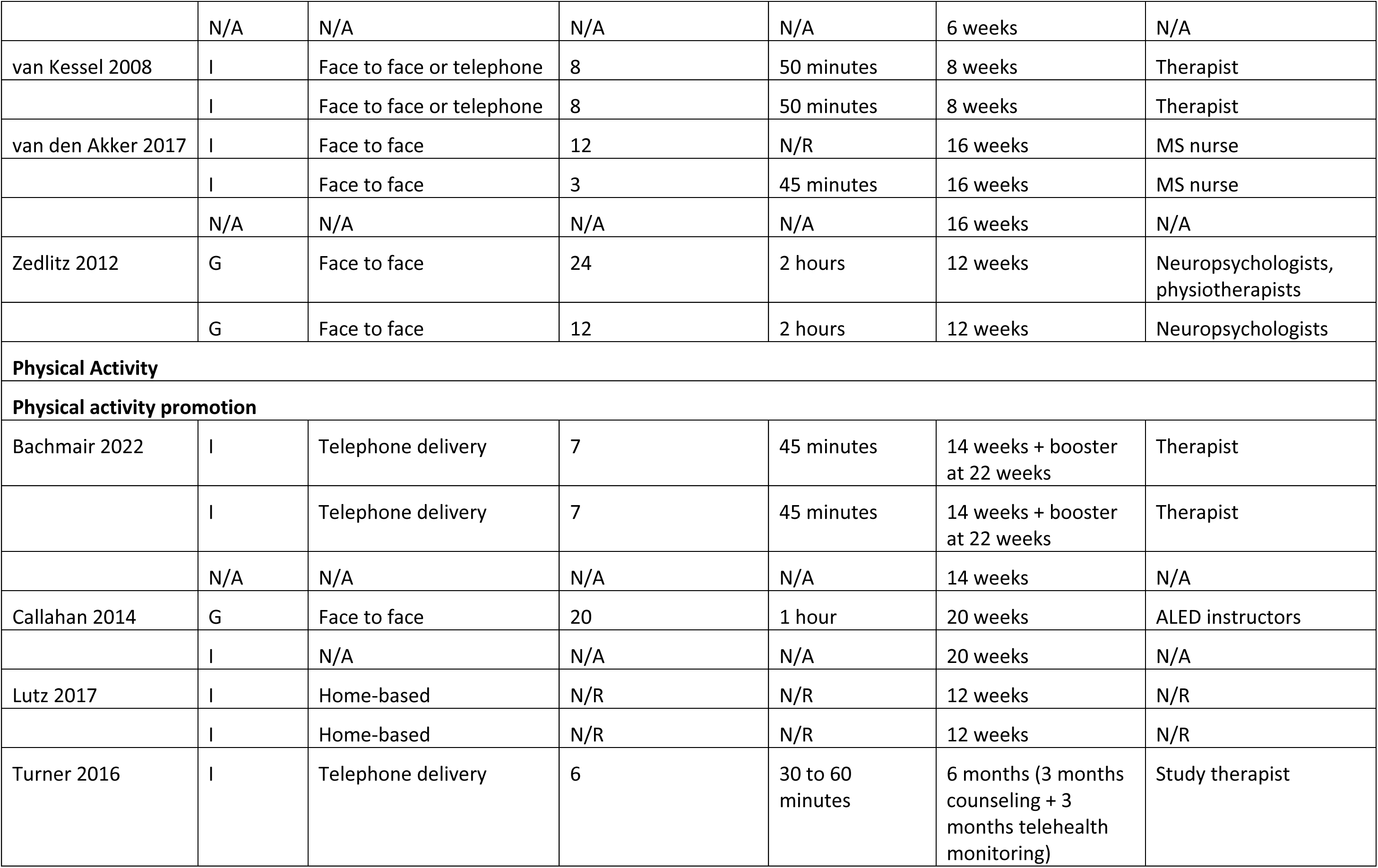
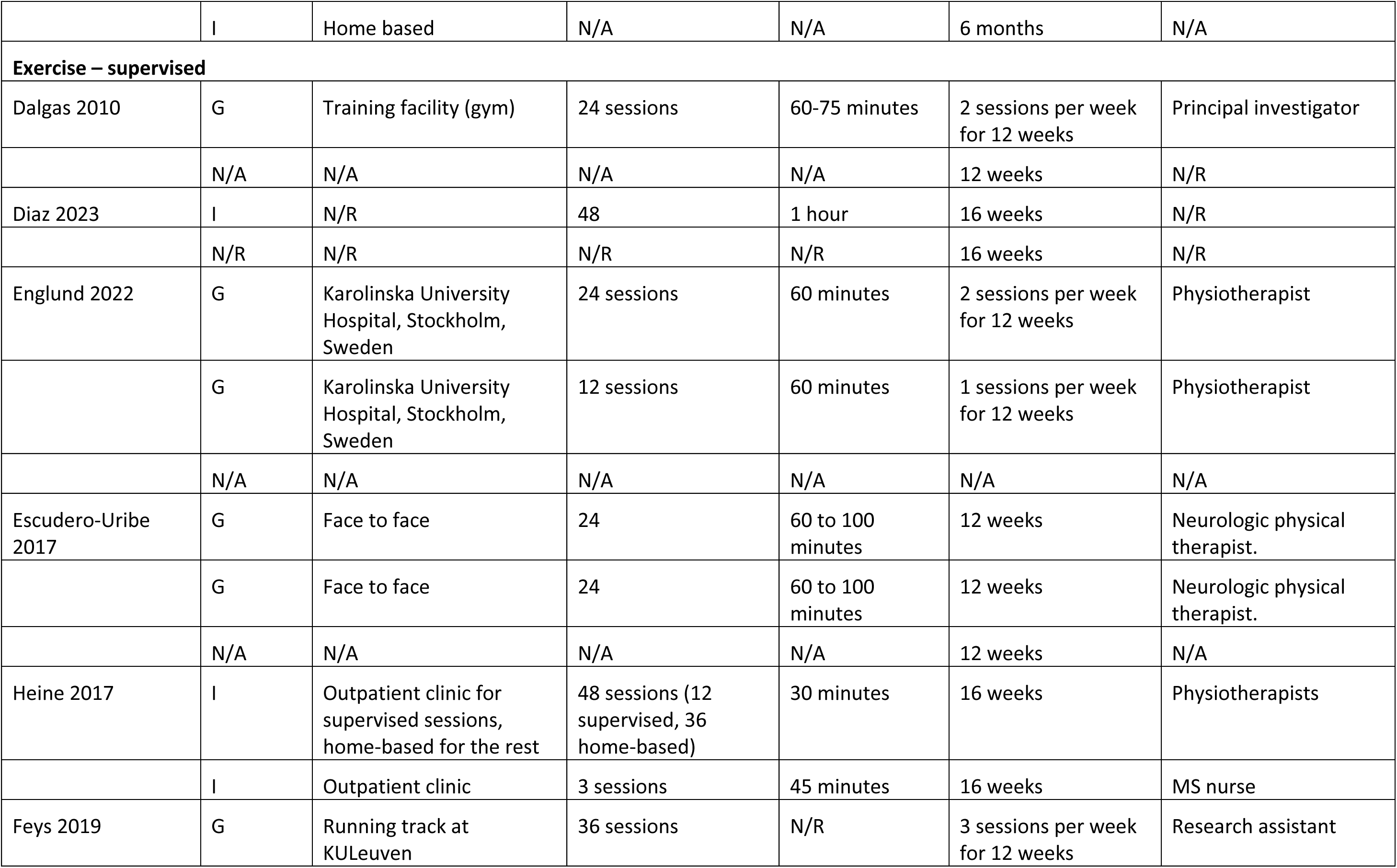
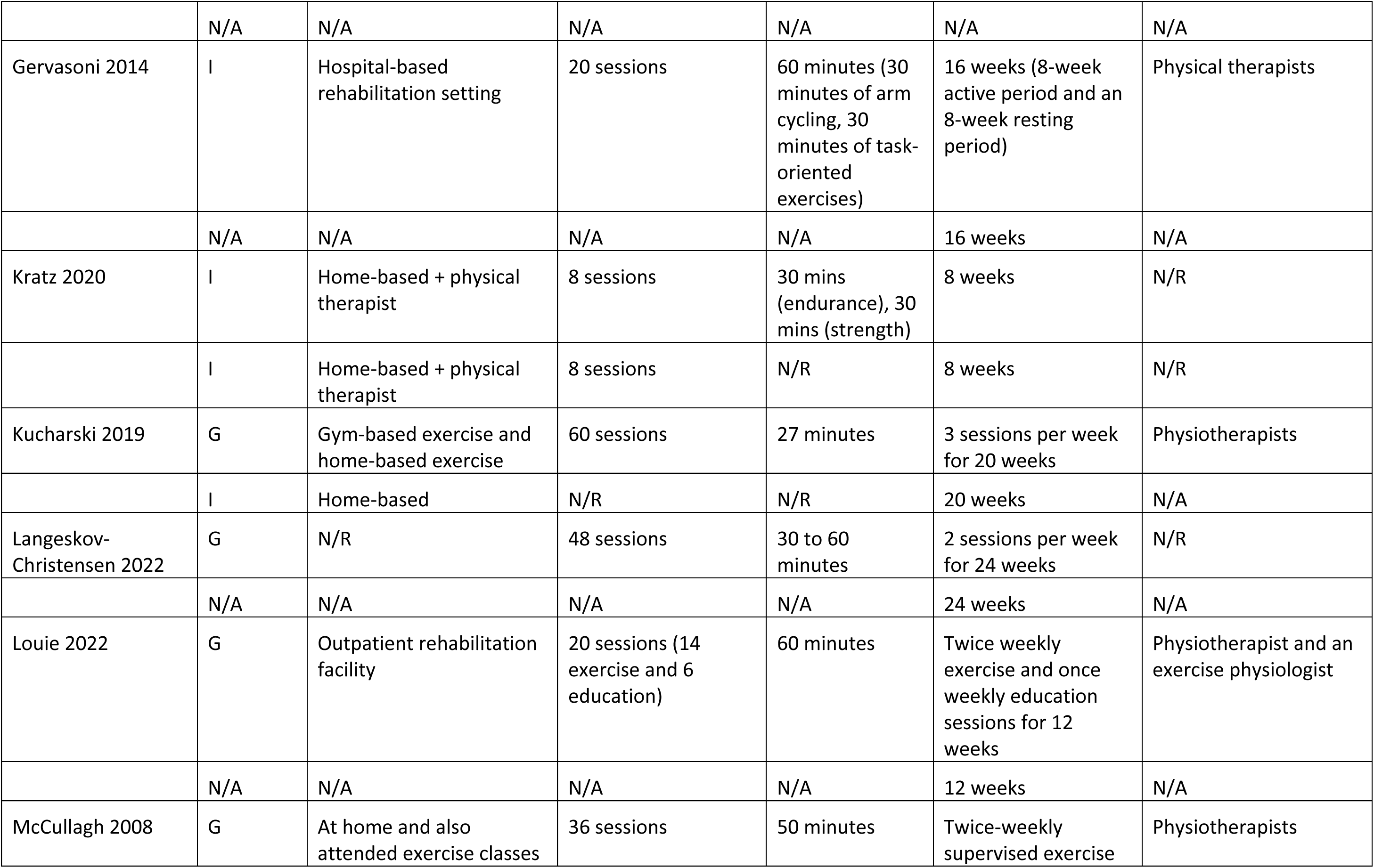
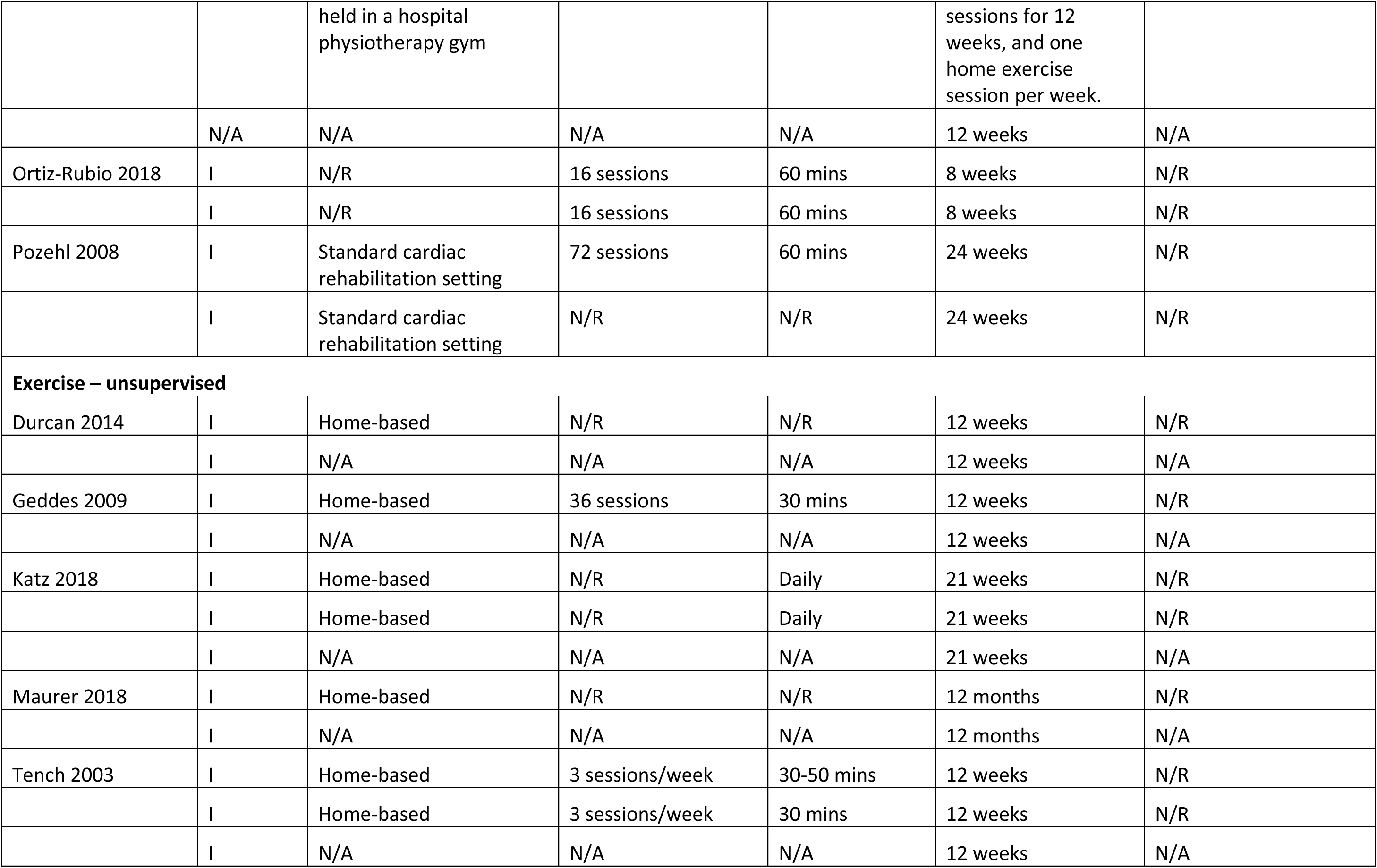
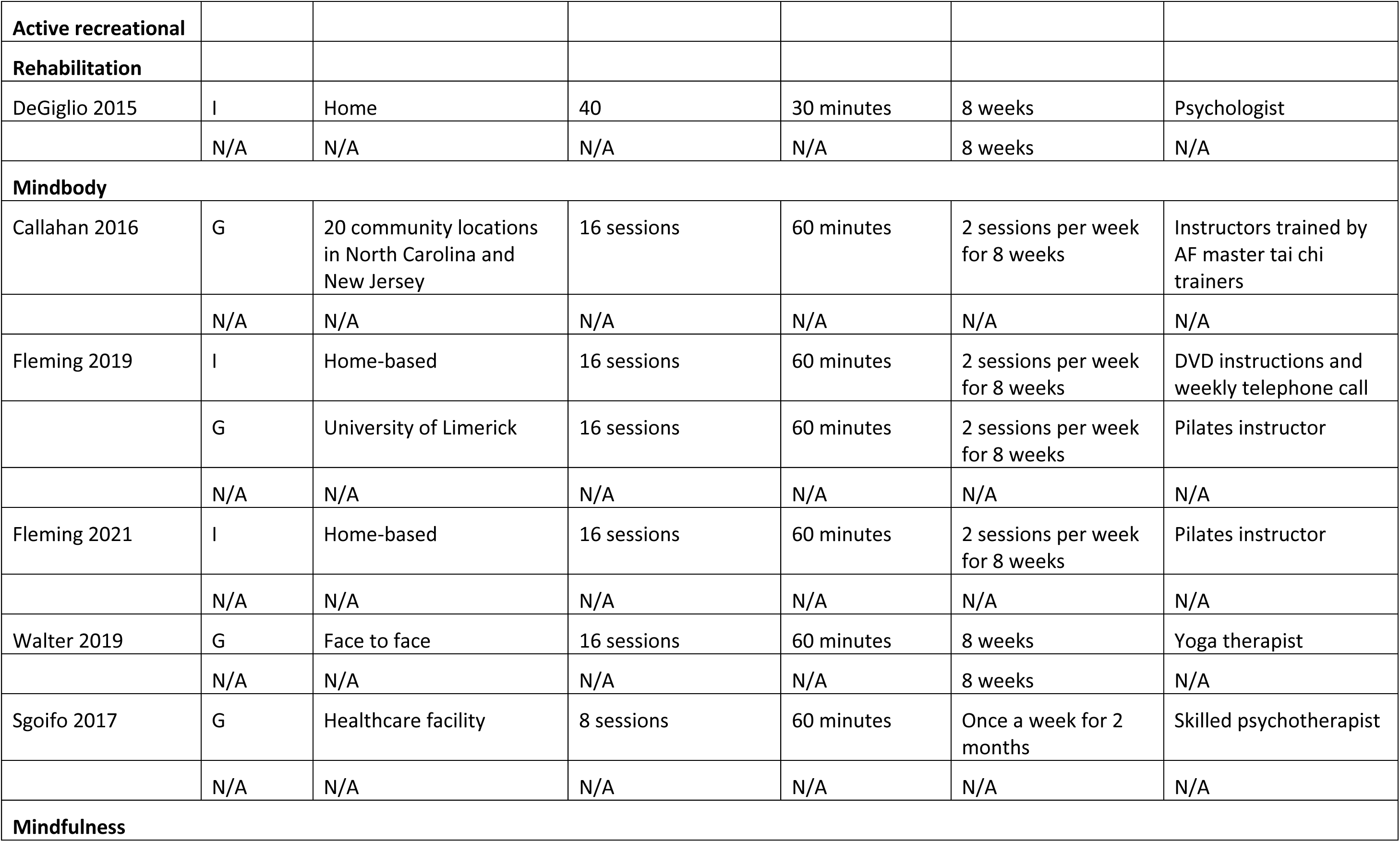
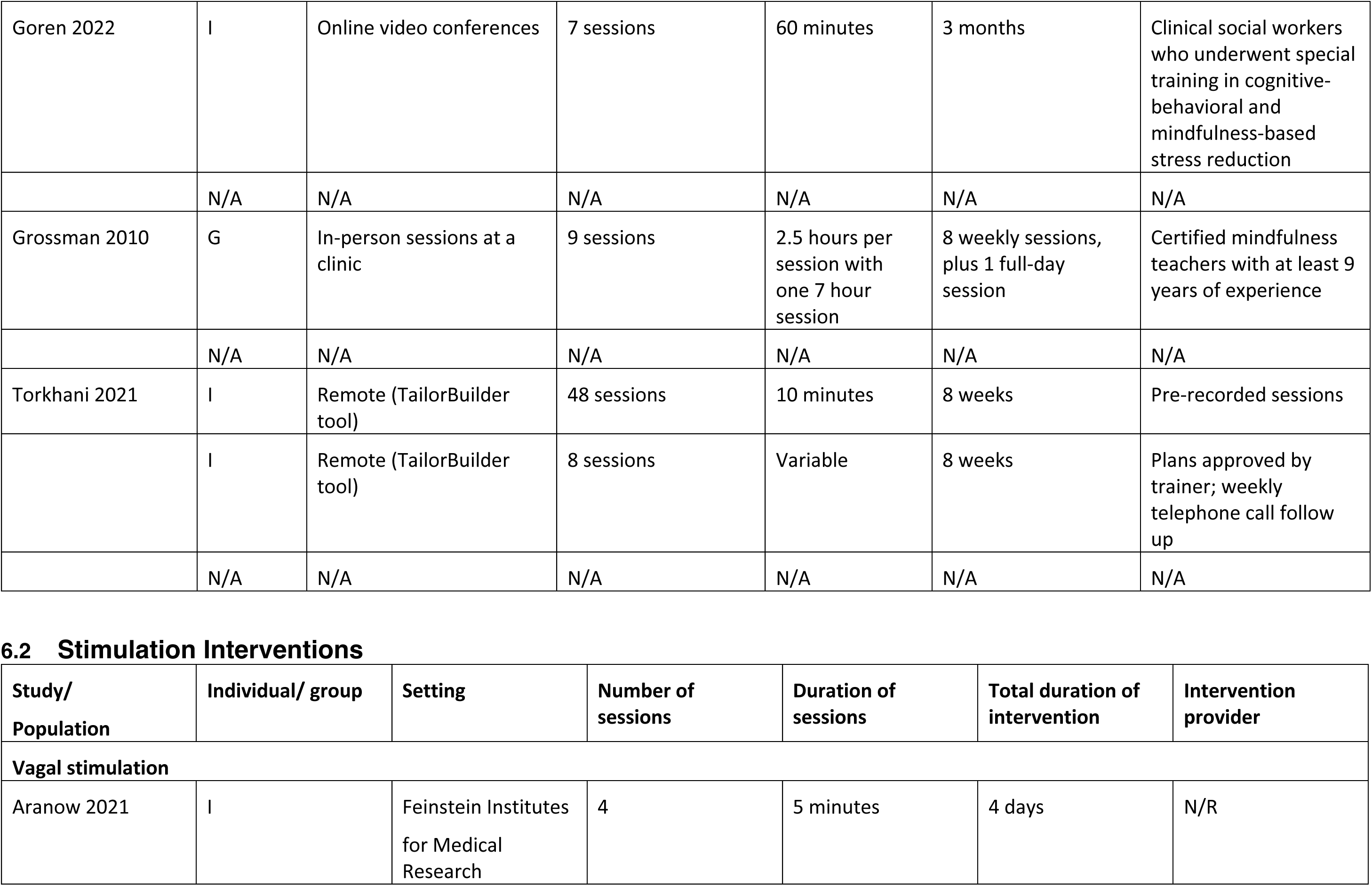
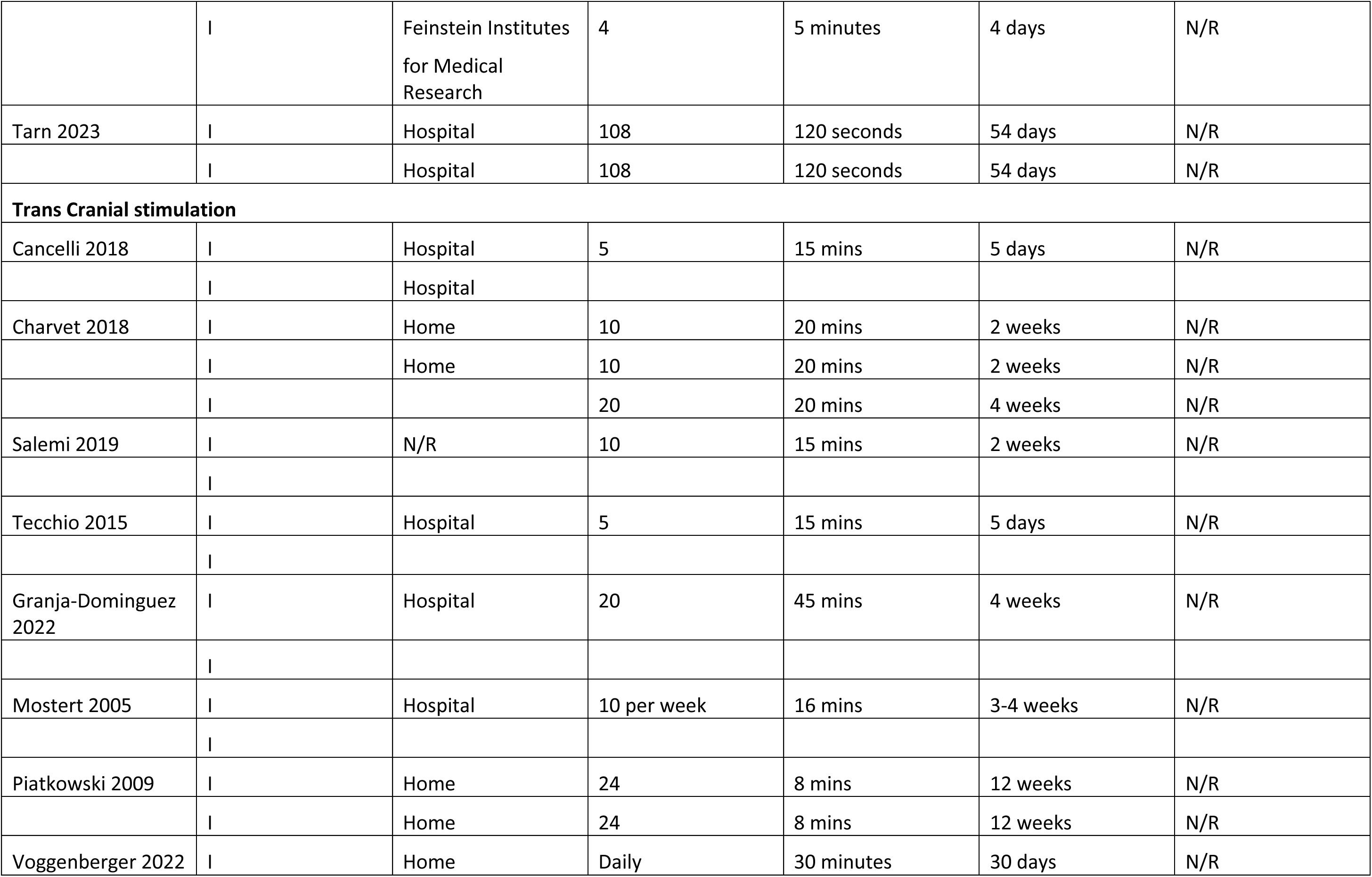
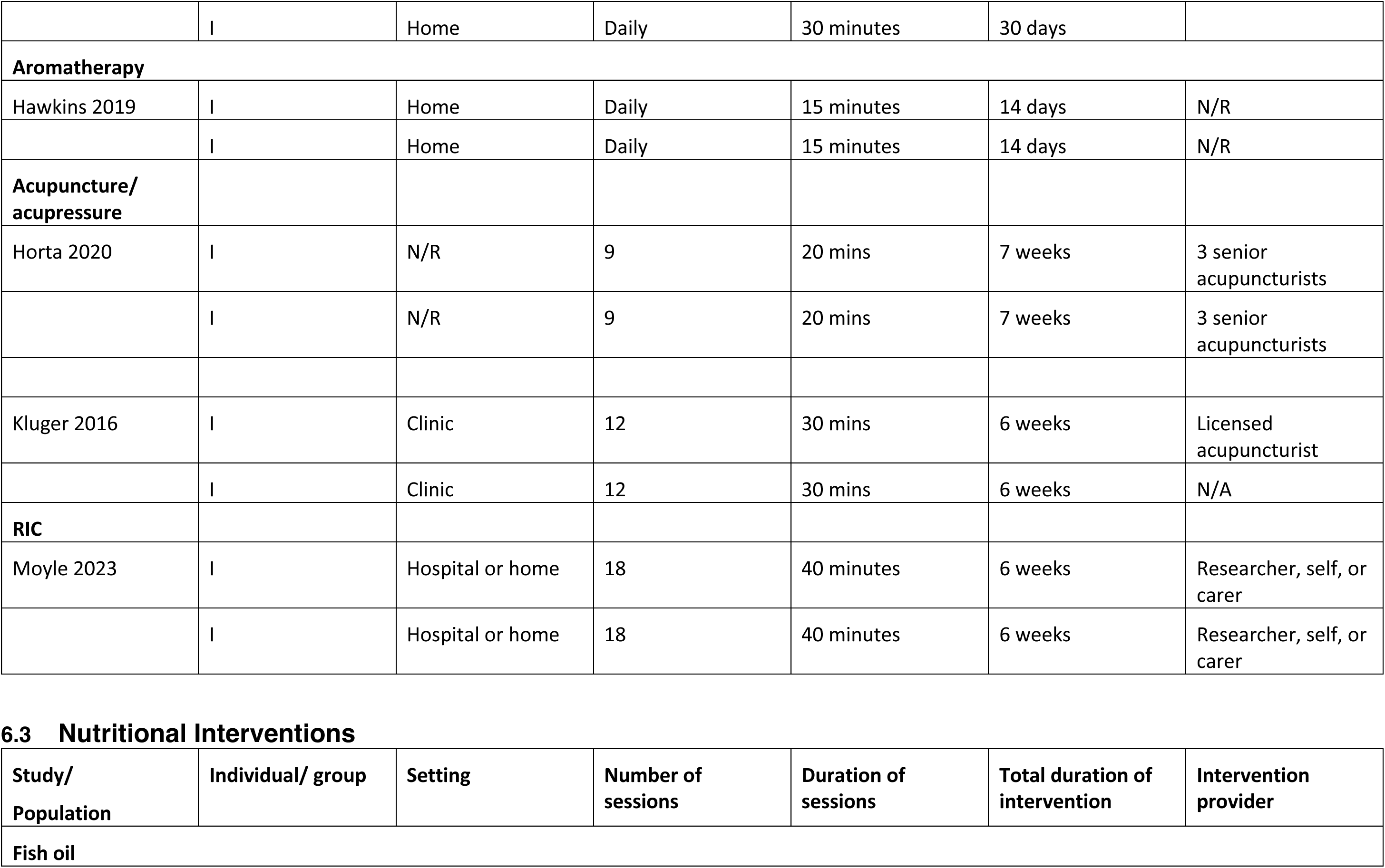
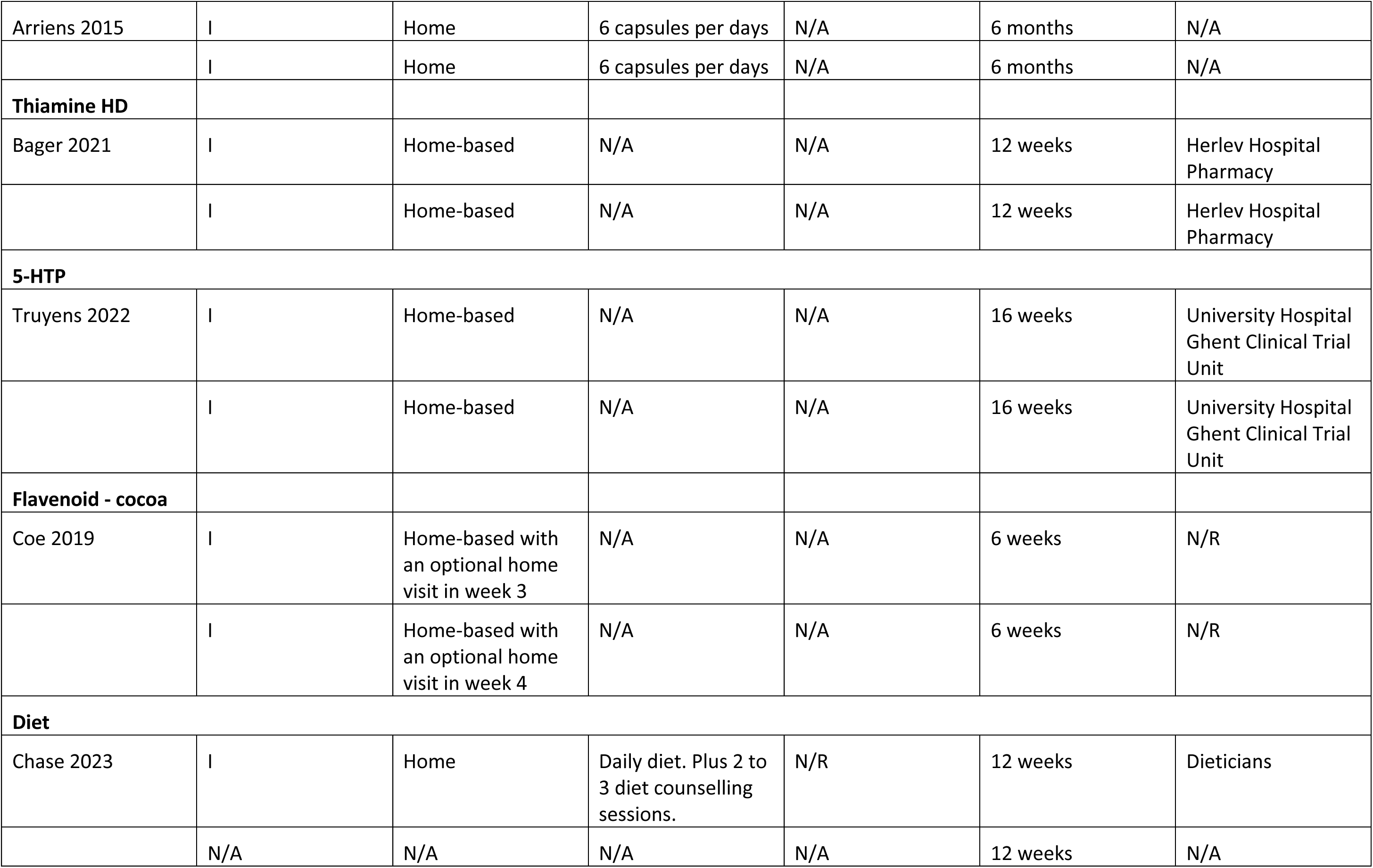
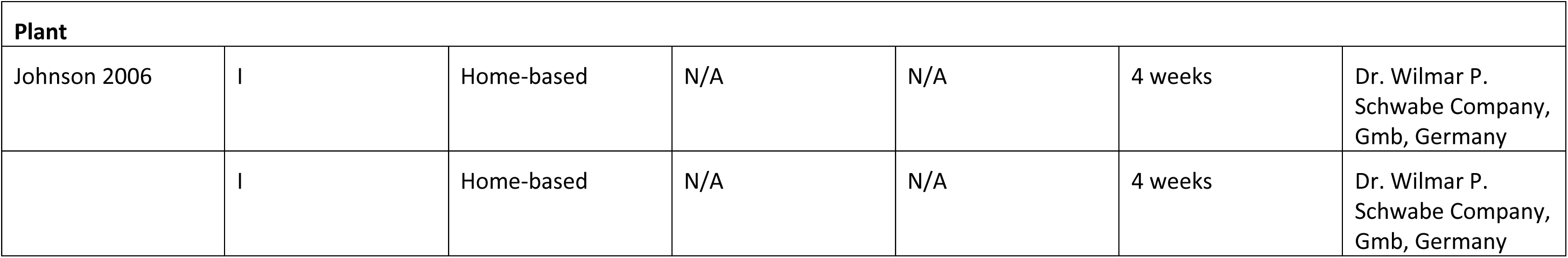

| Study/<br>Population | Individual/<br>group | Setting | Number of sessions | Duration of<br>sessions | Total duration of<br>intervention | Intervention provider |
| --- | --- | --- | --- | --- | --- | --- |
| **Self-Management** |  |  |  |  |  |  |
| **Fatigue self-management - conservative** |  |  |  |  |  |  |
| Abonie 2020 | I | Home monitoring | 1 | 30 minutes<br>intervention<br>session after 7<br>days home<br>monitoring | 4 weeks | N/R |
|  | N/A | N/A | N/A | N/A | 4 weeks | N/A |
| Askari 2022 | I | Web-based plus<br>telephone sessions | 6 telephone calls | 30-60 minutes | 12 weeks | Web + registered<br>occupational therapist |
|  | I | Web-based | N/A | N/A | 12 weeks | Web only |
| Blikman 2017 | I | Outpatient rehabilitation<br>department | 12 | 45 minutes | 4 months | Occupational therapist |
|  | I | Outpatient rehabilitation<br>department | 3 | 45 minutes | 4 months | Nurse |
| Farragher 2022 | I | Web-based + one-to-one<br>training | 3 web modules + 4-6<br>face-to-face sessions | Web modules:<br>20-30 minutes;<br>one-to-one<br>sessions: 30<br>minutes | 7-9 weeks | Occupational therapist |
|  | I | Web-based one-to-one sessions | 6 to 8 sessions | N/R | 7-9 weeks | Trained study coordinator |
| GarciaJalon 2013 | G | Face to face | 5 | 2 hours | 5 weeks | Therapist |
|  | G | Face to face | 5 | 2 hours | 5 weeks | Therapist |
| Ghahari 2010 | G | Online | 6 | 2-3 hours | 7 weeks | Occupational therapist |
|  | I | Online | 6 | N/R | 7 weeks | Group facilitator for technical queries only |
|  | I | N/A | N/A | N/A | 7 weeks | N/A |
| Hersche 2019 | G | Rehab centre | 6 x 1 hour + 1 x 0.5 hours | 1 hour or 0.5 hours | 3 weeks | Occupational therapist |
|  | G | Rehab centre | 6 | 1 hour | 3 weeks | Physical therapist |
| Hugos 2010 | G | N/R | 6 | 2 hours | 6 weeks | MS healthcare professionals via DVD |
|  | N/A | N/A | N/A | N/A | 6 weeks | N/A |
| Hugos 2019a/2017 | G | N/R | 6 | 2 hours | 6 weeks | MS professionals |
|  | G | N/R | 6 | 2 hours | 6 weeks | MS professionals |
| Kos 2016 | I | Face to face | 3 | 60-90 minutes | 3 weeks | Occupational therapist |
|  | I | Face to face | 3 | 60-90 minutes | 3 weeks | Occupational therapist |
| Murphy 2010 | I | Home and face to face | 2 | 1.5 hours | 4 weeks | Occupational therapist |
|  | I | Home and face to face | 2 | 1.5 hours | 4 weeks | Occupational therapist |
| **Fatigue self-management - active** |  |  |  |  |  |  |
| Clarke 2012 | G | Face to face | 6 | 60 minutes | 6 weeks | Researcher |
|  | G | Face to face | 6 | 60 minutes | 6 weeks | Researcher |
| Murphy 2024 | G | Online | 9 | 15-30 minutes | 12 weeks | Health coaches |
|  | N/A | N/A | N/A | N/A | 12 weeks | N/A |
| Rietberg 2014 | I | Outpatient | PT 24; minimum 2 session for other treatments | PT 45 minutes; 1 hour other treatments | 12 weeks | Multi-diciplinary |
|  | I | Outpatient | 2 | 1 hour | 12 weeks | Nurse |
| **General self-management** |  |  |  |  |  |  |
| Austin 1996 | I | Telephone | N/A | N/A | 6 months | Certified reality therapy counselor |
|  | I | Telephone | N/A | N/A | 6 months | Trained staff member |
| Feldthusen 2016 | I | Face to face | According to preferences | According to preferences | 12 weeks | Physical therapist |
|  | I | N/A | N/A | N/A | 12 weeks | N/A |
| Hammond 2008 | G | District or community hospitals | 8 | 2.5 hours | 3-4 weeks | Experienced therapists |
|  | G | District or community hospitals | 5 | 2 hours | 3-4 weeks | Experienced therapists |
| Khan 2020 | I | Telehealth | 16 | 20-30 minutes | 16 weeks | Health coach |
|  | I | N/A | N/A | N/A | 16 weeks | Usual physician |
| **CBT – fatigue** |  |  |  |  |  |  |
| Artom 2019 | I | Telephone | 1 x 1 hour and 7 x 30 minutes | 1 x 1 hour and 7 x 30 minutes | 8 weeks | Therapist |
|  | I | Home | N/A | N/A | 8 weeks | N/A |
| Bredero 2023 | G | Outpatient | 8 | 2.5 hours | 8 weeks | 3 licensed and experienced mindfulness trainers. |
|  | N/A | N/A | N/A | N/A | 8 weeks | N/A |
| Ehde 2015 | I | Telehealth | 8 + 2 follow-up calls | 45-60 minutes | 8 weeks + follow up call at 4 and 8 weeks post treatment | Study therapist |
|  | I | Telehealth | 8 + 2 follow-up calls | 45-60 minutes | 8 weeks + follow up call at 4 and 8 weeks post treatment | Study therapist |
| Gay 2023 | G | Face to face | 6 + 4 booster sessions | 90 minutes | 6 weeks + booster sessions | Occupational therapists, physiotherapists, MS nurses |
|  | I | Face to face | N/A | N/A | 6 weeks | Usual clinician |
| Hewlett 2011 | G | Face to face | 6 | 2 hours | 6 weeks | Clinical psychologist + occupational therapist |
|  | G | Face to face | 1 | 1 hour | 6 weeks | Rheumatology nurse |
| Hewlett 2019a | G | Face to face | 7 | 2 hours for first 6 weeks, 1 hour consolidation session | 6 weeks | Nurses, occupational therapists |
|  | N/A | N/A | N/A | N/A | 6 weeks | N/A |
| Jhamb 2023 | I | Telemedicine at home | 12 | 45-60 minutes | 12 weeks | Therapist with counselling qualification |
|  | G | Face to face | 6 | 20-30 minutes | 12 weeks | Research coordinator |
| Mead 2022 | I | Telephone | 6 | 1 hour | 12 weeks | Stroke nurses + physiotherapist + psychologist |
|  | I | Home | N/A | N/A | 12 weeks | N/A |
| Menting 2017 | I | Web-based with face to face | 5 to 8 | 50 minutes | 5 months | Therapist |
|  | N/A | N/A | N/A | N/A | 5 months | N/A |
| Moss-Morris 2012 | I | Web-based | 8 | 25 to 50 minutes | 8 weeks | Assistant psychologist |
|  | N/A | Standard care | N/A | N/A | 8 weeks | N/A |
| Nguyen 2019 | I | Face to face with homework | 8 | N/R | 2 months | Licensed psychologists and exercise physiologist |
|  | N/A | N/A | N/A | N/A | 2 months | N/A |
| Okkersen 2018 | I | Face to face | 10 to 14 | N/R | 10 months | Therapists experienced in CBT |
|  | N/A | N/A | N/A | N/A | 10 months | N/A |
| Picariello 2021 | I | Face to face and telephone | 3 to 5 | 2 session of 1 hour + 1 to 3 sessions of 30 minutes | 3 months | Researcher with background in health psychology |
|  | N/A | N/A | N/A | N/A | 3 months | N/A |
| Pottgen 2018 | I | Web-based | Average access 14.5 times | N/R | 12 weeks | Web-site |
|  | N/A | N/A | N/A | N/A | 12 weeks | N/A |
| Thomas 2013 | G | Face to face | 6 | 90 minutes | 6 weeks | Health professionals |
|  | N/A | N/A | N/A | N/A | 6 weeks | N/A |
| van Kessel 2008 | I | Face to face or telephone | 8 | 50 minutes | 8 weeks | Therapist |
|  | I | Face to face or telephone | 8 | 50 minutes | 8 weeks | Therapist |
| van den Akker 2017 | I | Face to face | 12 | N/R | 16 weeks | MS nurse |
|  | I | Face to face | 3 | 45 minutes | 16 weeks | MS nurse |
|  | N/A | N/A | N/A | N/A | 16 weeks | N/A |
| Zedlitz 2012 | G | Face to face | 24 | 2 hours | 12 weeks | Neuropsychologists,<br>physiotherapists |
|  | G | Face to face | 12 | 2 hours | 12 weeks | Neuropsychologists |
| **Physical Activity** |  |  |  |  |  |  |
| **Physical activity promotion** |  |  |  |  |  |  |
| Bachmair 2022 | I | Telephone delivery | 7 | 45 minutes | 14 weeks + booster<br>at 22 weeks | Therapist |
|  | I | Telephone delivery | 7 | 45 minutes | 14 weeks + booster<br>at 22 weeks | Therapist |
|  | N/A | N/A | N/A | N/A | 14 weeks | N/A |
| Callahan 2014 | G | Face to face | 20 | 1 hour | 20 weeks | ALED instructors |
|  | I | N/A | N/A | N/A | 20 weeks | N/A |
| Lutz 2017 | I | Home-based | N/R | N/R | 12 weeks | N/R |
|  | I | Home-based | N/R | N/R | 12 weeks | N/R |
| Turner 2016 | I | Telephone delivery | 6 | 30 to 60<br>minutes | 6 months (3 months<br>counseling + 3<br>months telehealth<br>monitoring) | Study therapist |
|  | I | Home based | N/A | N/A | 6 months | N/A |
| **Exercise – supervised** |  |  |  |  |  |  |
| Dalgas 2010 | G | Training facility (gym) | 24 sessions | 60-75 minutes | 2 sessions per week for 12 weeks | Principal investigator |
|  | N/A | N/A | N/A | N/A | 12 weeks | N/R |
| Diaz 2023 | I | N/R | 48 | 1 hour | 16 weeks | N/R |
|  | N/R | N/R | N/R | N/R | 16 weeks | N/R |
| Englund 2022 | G | Karolinska University Hospital, Stockholm, Sweden | 24 sessions | 60 minutes | 2 sessions per week for 12 weeks | Physiotherapist |
|  | G | Karolinska University Hospital, Stockholm, Sweden | 12 sessions | 60 minutes | 1 sessions per week for 12 weeks | Physiotherapist |
|  | N/A | N/A | N/A | N/A | N/A | N/A |
| Escudero-Uribe 2017 | G | Face to face | 24 | 60 to 100 minutes | 12 weeks | Neurologic physical therapist. |
|  | G | Face to face | 24 | 60 to 100 minutes | 12 weeks | Neurologic physical therapist. |
|  | N/A | N/A | N/A | N/A | 12 weeks | N/A |
| Heine 2017 | I | Outpatient clinic for supervised sessions, home-based for the rest | 48 sessions (12 supervised, 36 home-based) | 30 minutes | 16 weeks | Physiotherapists |
|  | I | Outpatient clinic | 3 sessions | 45 minutes | 16 weeks | MS nurse |
| Feys 2019 | G | Running track at KULeuven | 36 sessions | N/R | 3 sessions per week for 12 weeks | Research assistant |
|  | N/A | N/A | N/A | N/A | N/A | N/A |
| Gervasoni 2014 | I | Hospital-based rehabilitation setting | 20 sessions | 60 minutes (30 minutes of arm cycling, 30 minutes of task-oriented exercises) | 16 weeks (8-week active period and an 8-week resting period) | Physical therapists |
|  | N/A | N/A | N/A | N/A | 16 weeks | N/A |
| Kratz 2020 | I | Home-based + physical therapist | 8 sessions | 30 mins (endurance), 30 mins (strength) | 8 weeks | N/R |
|  | I | Home-based + physical therapist | 8 sessions | N/R | 8 weeks | N/R |
| Kucharski 2019 | G | Gym-based exercise and home-based exercise | 60 sessions | 27 minutes | 3 sessions per week for 20 weeks | Physiotherapists |
|  | I | Home-based | N/R | N/R | 20 weeks | N/A |
| Langeskov-Christensen 2022 | G | N/R | 48 sessions | 30 to 60 minutes | 2 sessions per week for 24 weeks | N/R |
|  | N/A | N/A | N/A | N/A | 24 weeks | N/A |
| Louie 2022 | G | Outpatient rehabilitation facility | 20 sessions (14 exercise and 6 education) | 60 minutes | Twice weekly exercise and once weekly education sessions for 12 weeks | Physiotherapist and an exercise physiologist |
|  | N/A | N/A | N/A | N/A | 12 weeks | N/A |
| McCullagh 2008 | G | At home and also attended exercise classes | 36 sessions | 50 minutes | Twice-weekly supervised exercise | Physiotherapists |
|  |  | held in a hospital physiotherapy gym |  |  | sessions for 12 weeks, and one home exercise session per week. |  |
|  | N/A | N/A | N/A | N/A | 12 weeks | N/A |
| Ortiz-Rubio 2018 | I | N/R | 16 sessions | 60 mins | 8 weeks | N/R |
|  | I | N/R | 16 sessions | 60 mins | 8 weeks | N/R |
| Pozehl 2008 | I | Standard cardiac rehabilitation setting | 72 sessions | 60 mins | 24 weeks | N/R |
|  | I | Standard cardiac rehabilitation setting | N/R | N/R | 24 weeks | N/R |
| **Exercise – unsupervised** |  |  |  |  |  |  |
| Durcan 2014 | I | Home-based | N/R | N/R | 12 weeks | N/R |
|  | I | N/A | N/A | N/A | 12 weeks | N/A |
| Geddes 2009 | I | Home-based | 36 sessions | 30 mins | 12 weeks | N/R |
|  | I | N/A | N/A | N/A | 12 weeks | N/A |
| Katz 2018 | I | Home-based | N/R | Daily | 21 weeks | N/R |
|  | I | Home-based | N/R | Daily | 21 weeks | N/R |
|  | I | N/A | N/A | N/A | 21 weeks | N/A |
| Maurer 2018 | I | Home-based | N/R | N/R | 12 months | N/R |
|  | I | N/A | N/A | N/A | 12 months | N/A |
| Tench 2003 | I | Home-based | 3 sessions/week | 30-50 mins | 12 weeks | N/R |
|  | I | Home-based | 3 sessions/week | 30 mins | 12 weeks | N/R |
|  | I | N/A | N/A | N/A | 12 weeks | N/A |
| **Active recreational** |  |  |  |  |  |  |
| **Rehabilitation** |  |  |  |  |  |  |
| DeGiglio 2015 | I | Home | 40 | 30 minutes | 8 weeks | Psychologist |
|  | N/A | N/A | N/A | N/A | 8 weeks | N/A |
| **Mindbody** |  |  |  |  |  |  |
| Callahan 2016 | G | 20 community locations in North Carolina and New Jersey | 16 sessions | 60 minutes | 2 sessions per week for 8 weeks | Instructors trained by AF master tai chi trainers |
|  | N/A | N/A | N/A | N/A | N/A | N/A |
| Fleming 2019 | I | Home-based | 16 sessions | 60 minutes | 2 sessions per week for 8 weeks | DVD instructions and weekly telephone call |
|  | G | University of Limerick | 16 sessions | 60 minutes | 2 sessions per week for 8 weeks | Pilates instructor |
|  | N/A | N/A | N/A | N/A | N/A | N/A |
| Fleming 2021 | I | Home-based | 16 sessions | 60 minutes | 2 sessions per week for 8 weeks | Pilates instructor |
|  | N/A | N/A | N/A | N/A | N/A | N/A |
| Walter 2019 | G | Face to face | 16 sessions | 60 minutes | 8 weeks | Yoga therapist |
|  | N/A | N/A | N/A | N/A | 8 weeks | N/A |
| Sgoifo 2017 | G | Healthcare facility | 8 sessions | 60 minutes | Once a week for 2 months | Skilled psychotherapist |
|  | N/A | N/A | N/A | N/A | N/A | N/A |
| **Mindfulness** |  |  |  |  |  |  |
| Goren 2022 | I | Online video conferences | 7 sessions | 60 minutes | 3 months | Clinical social workers who underwent special training in cognitive-behavioral and mindfulness-based stress reduction |
|  | N/A | N/A | N/A | N/A | N/A | N/A |
| Grossman 2010 | G | In-person sessions at a clinic | 9 sessions | 2.5 hours per session with one 7 hour session | 8 weekly sessions, plus 1 full-day session | Certified mindfulness teachers with at least 9 years of experience |
|  | N/A | N/A | N/A | N/A | N/A | N/A |
| Torkhani 2021 | I | Remote (TailorBuilder tool) | 48 sessions | 10 minutes | 8 weeks | Pre-recorded sessions |
|  | I | Remote (TailorBuilder tool) | 8 sessions | Variable | 8 weeks | Plans approved by trainer; weekly telephone call follow up |
|  | N/A | N/A | N/A | N/A | N/A | N/A |

6.2 Stimulation Interventions
| Study/<br>Population | Individual/ group | Setting | Number of<br>sessions | Duration of<br>sessions | Total duration of<br>intervention | Intervention<br>provider |
| --- | --- | --- | --- | --- | --- | --- |
| **Vagal stimulation** |  |  |  |  |  |  |
| Aranow 2021 | I | Feinstein Institutes<br>for Medical<br>Research | 4 | 5 minutes | 4 days | N/R |
|  | I | Feinstein Institutes<br>for Medical<br>Research | 4 | 5 minutes | 4 days | N/R |
| Tarn 2023 | I | Hospital | 108 | 120 seconds | 54 days | N/R |
|  | I | Hospital | 108 | 120 seconds | 54 days | N/R |
| **Trans Cranial stimulation** |  |  |  |  |  |  |
| Cancelli 2018 | I | Hospital | 5 | 15 mins | 5 days | N/R |
|  | I | Hospital |  |  |  |  |
| Charvet 2018 | I | Home | 10 | 20 mins | 2 weeks | N/R |
|  | I | Home | 10 | 20 mins | 2 weeks | N/R |
|  | I |  | 20 | 20 mins | 4 weeks | N/R |
| Salemi 2019 | I | N/R | 10 | 15 mins | 2 weeks | N/R |
|  | I |  |  |  |  |  |
| Tecchio 2015 | I | Hospital | 5 | 15 mins | 5 days | N/R |
|  | I |  |  |  |  |  |
| Granja-Dominguez<br>2022 | I | Hospital | 20 | 45 mins | 4 weeks | N/R |
|  | I |  |  |  |  |  |
| Mostert 2005 | I | Hospital | 10 per week | 16 mins | 3-4 weeks | N/R |
|  | I |  |  |  |  |  |
| Piatkowski 2009 | I | Home | 24 | 8 mins | 12 weeks | N/R |
|  | I | Home | 24 | 8 mins | 12 weeks | N/R |
| Voggenberger 2022 | I | Home | Daily | 30 minutes | 30 days | N/R |
|  | I | Home | Daily | 30 minutes | 30 days |  |
| **Aromatherapy** |  |  |  |  |  |  |
| Hawkins 2019 | I | Home | Daily | 15 minutes | 14 days | N/R |
|  | I | Home | Daily | 15 minutes | 14 days | N/R |
| **Acupuncture/**<br>**acupressure** |  |  |  |  |  |  |
| Horta 2020 | I | N/R | 9 | 20 mins | 7 weeks | 3 senior<br>acupuncturists |
|  | I | N/R | 9 | 20 mins | 7 weeks | 3 senior<br>acupuncturists |
| Kluger 2016 | I | Clinic | 12 | 30 mins | 6 weeks | Licensed<br>acupuncturist |
|  | I | Clinic | 12 | 30 mins | 6 weeks | N/A |
| **RIC** |  |  |  |  |  |  |
| Moyle 2023 | I | Hospital or home | 18 | 40 minutes | 6 weeks | Researcher, self, or<br>carer |
|  | I | Hospital or home | 18 | 40 minutes | 6 weeks | Researcher, self, or<br>carer |

6.3 Nutritional Interventions
| Study/<br>Population | Individual/ group | Setting | Number of<br>sessions | Duration of<br>sessions | Total duration of<br>intervention | Intervention<br>provider |
| --- | --- | --- | --- | --- | --- | --- |
| **Fish oil** |  |  |  |  |  |  |
| Arriens 2015 | I | Home | 6 capsules per days | N/A | 6 months | N/A |
|  | I | Home | 6 capsules per days | N/A | 6 months | N/A |
| **Thiamine HD** |  |  |  |  |  |  |
| Bager 2021 | I | Home-based | N/A | N/A | 12 weeks | Herlev Hospital Pharmacy |
|  | I | Home-based | N/A | N/A | 12 weeks | Herlev Hospital Pharmacy |
| **5-HTP** |  |  |  |  |  |  |
| Truyens 2022 | I | Home-based | N/A | N/A | 16 weeks | University Hospital Ghent Clinical Trial Unit |
|  | I | Home-based | N/A | N/A | 16 weeks | University Hospital Ghent Clinical Trial Unit |
| **Flavenoid - cocoa** |  |  |  |  |  |  |
| Coe 2019 | I | Home-based with an optional home visit in week 3 | N/A | N/A | 6 weeks | N/R |
|  | I | Home-based with an optional home visit in week 4 | N/A | N/A | 6 weeks | N/R |
| **Diet** |  |  |  |  |  |  |
| Chase 2023 | I | Home | Daily diet. Plus 2 to 3 diet counselling sessions. | N/R | 12 weeks | Dieticians |
|  | N/A | N/A | N/A | N/A | 12 weeks | N/A |

| Plant |  |  |  |  |  |  |
| --- | --- | --- | --- | --- | --- | --- |
| Johnson 2006 | I | Home-based | N/A | N/A | 4 weeks | Dr. Wilmar P. Schwabe Company, Gmb, Germany |
|  | I | Home-based | N/A | N/A | 4 weeks | Dr. Wilmar P. Schwabe Company, Gmb, Germany |

### Supplemental Results 7. Intervention characteristics, studies not included in NMA

#### 7.1 Intervention content

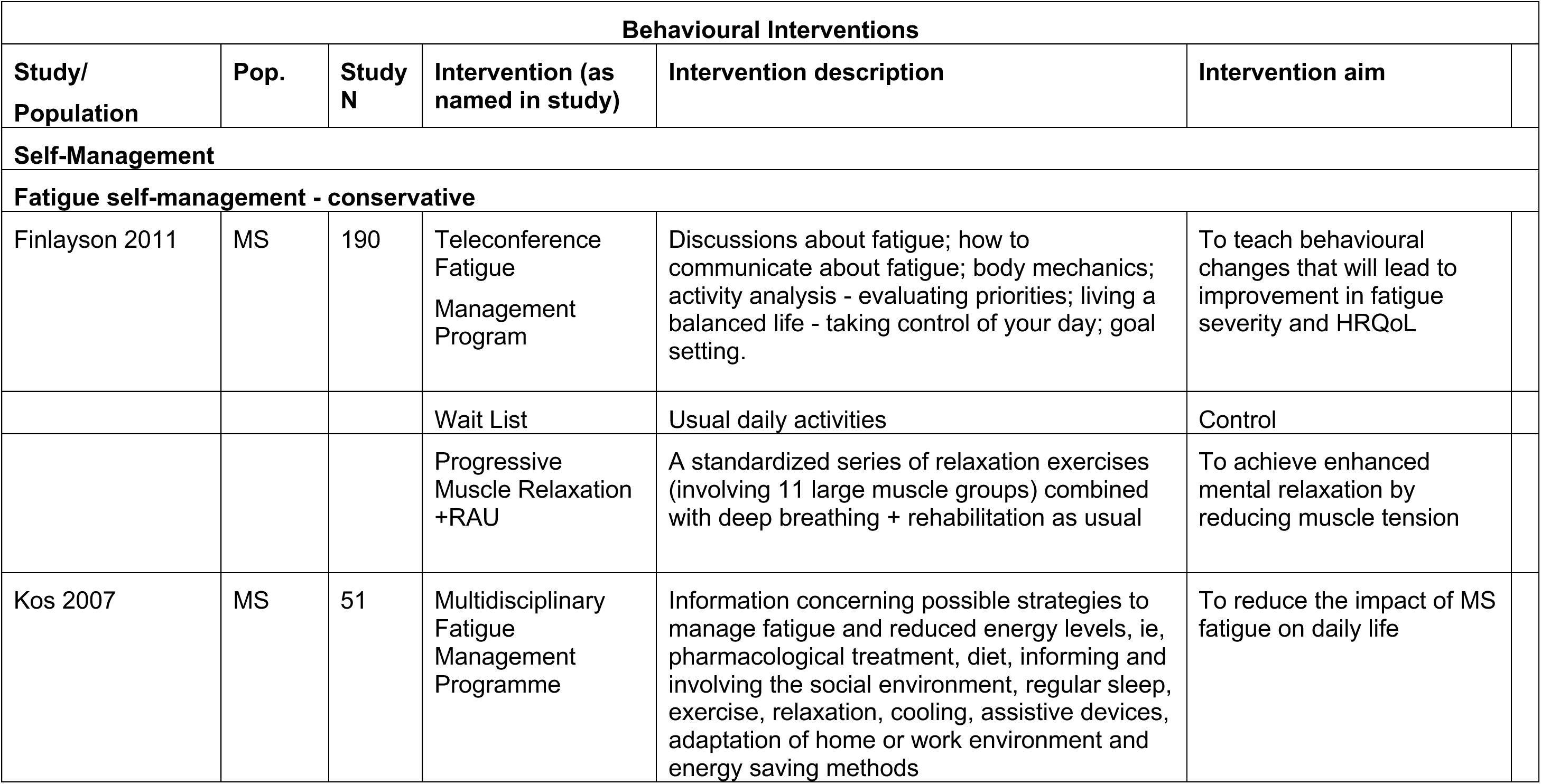
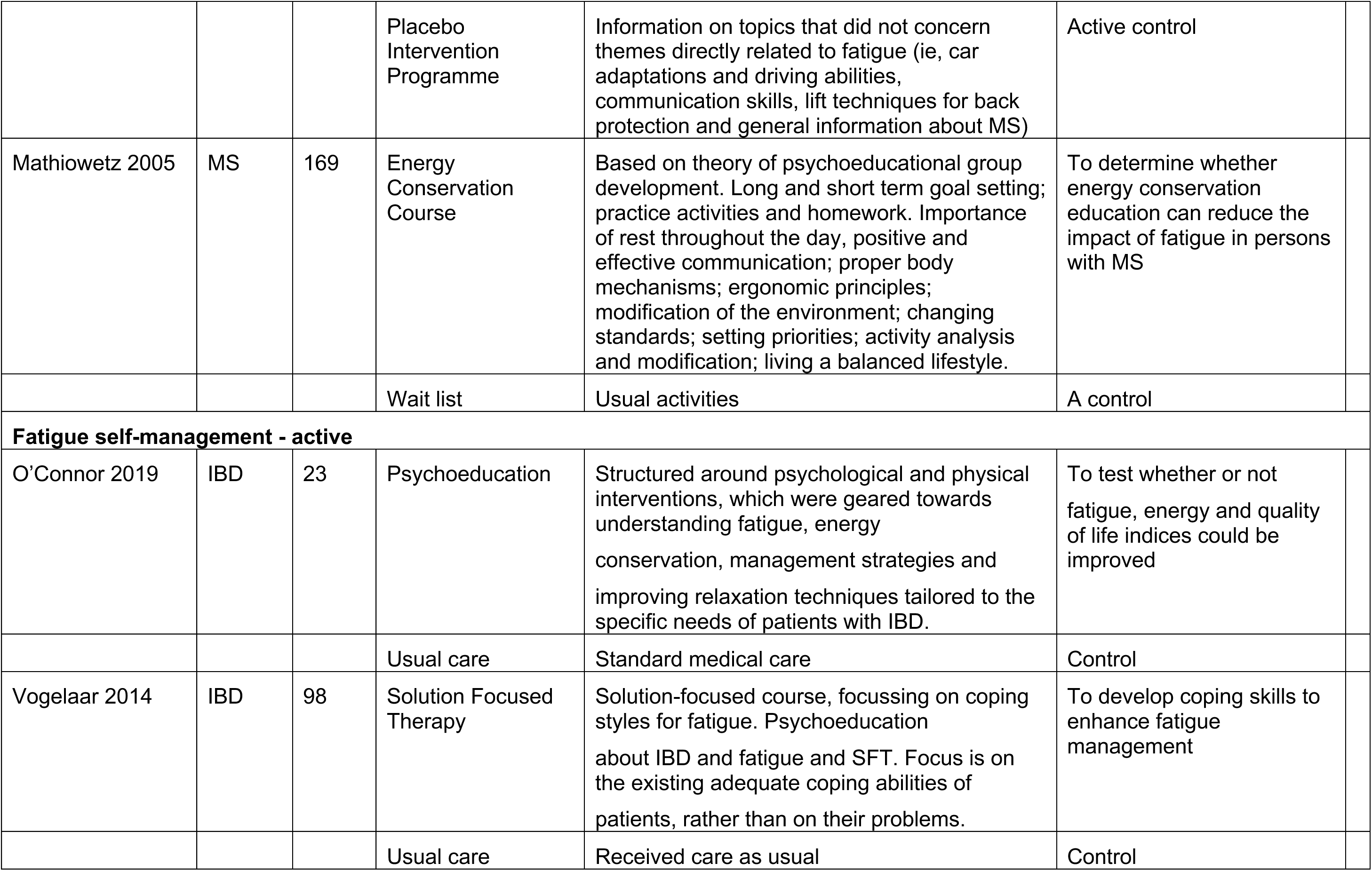
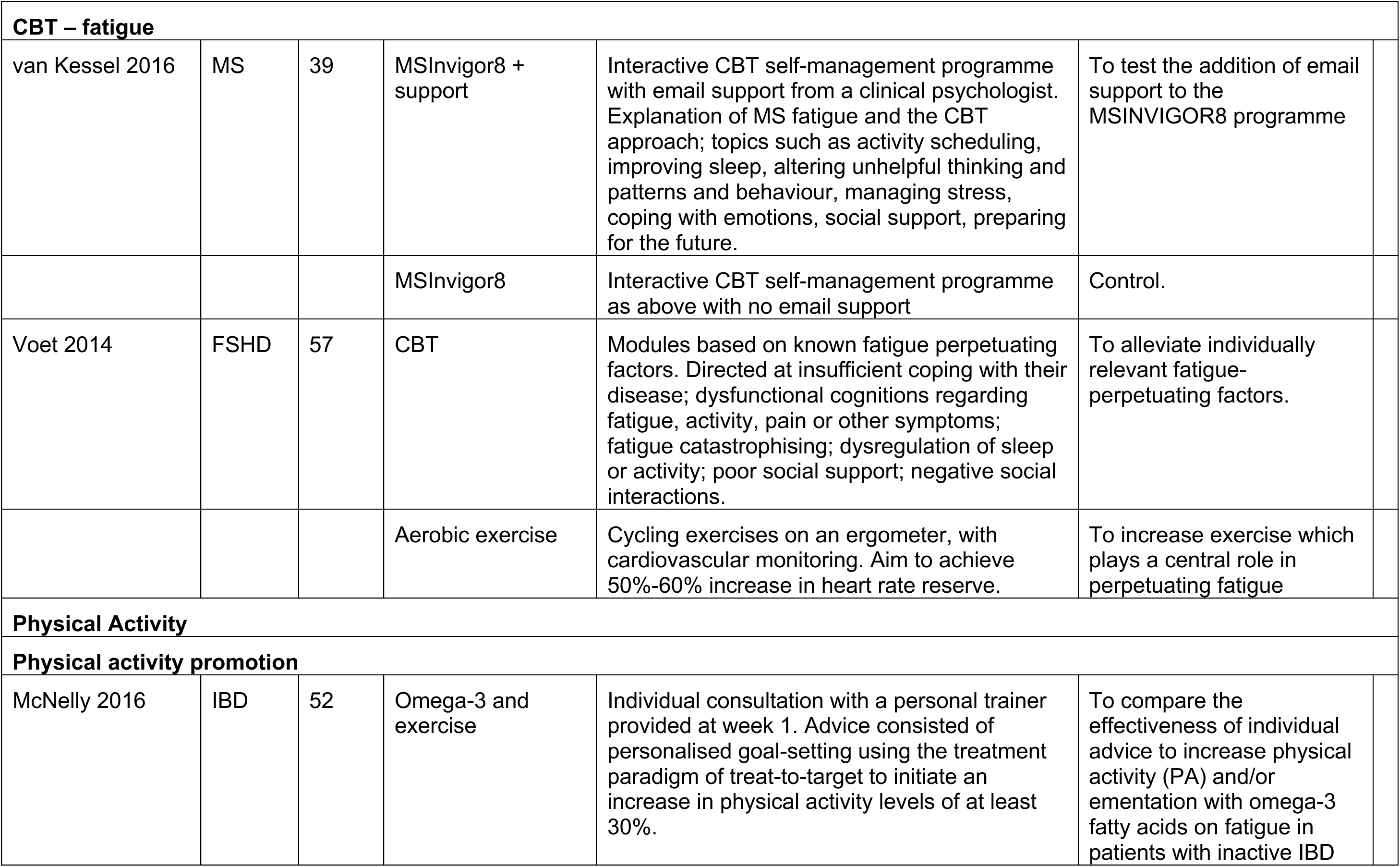
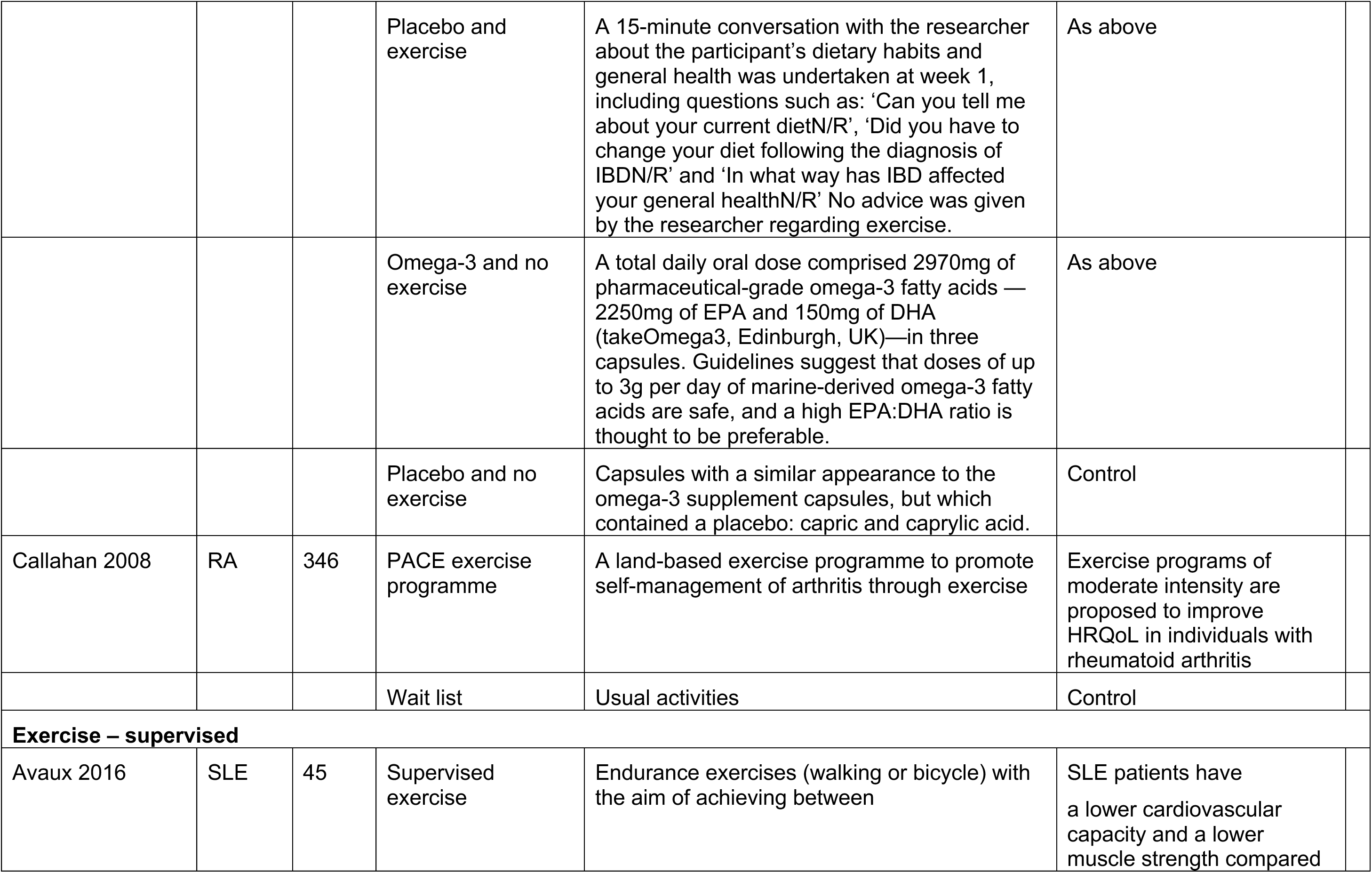
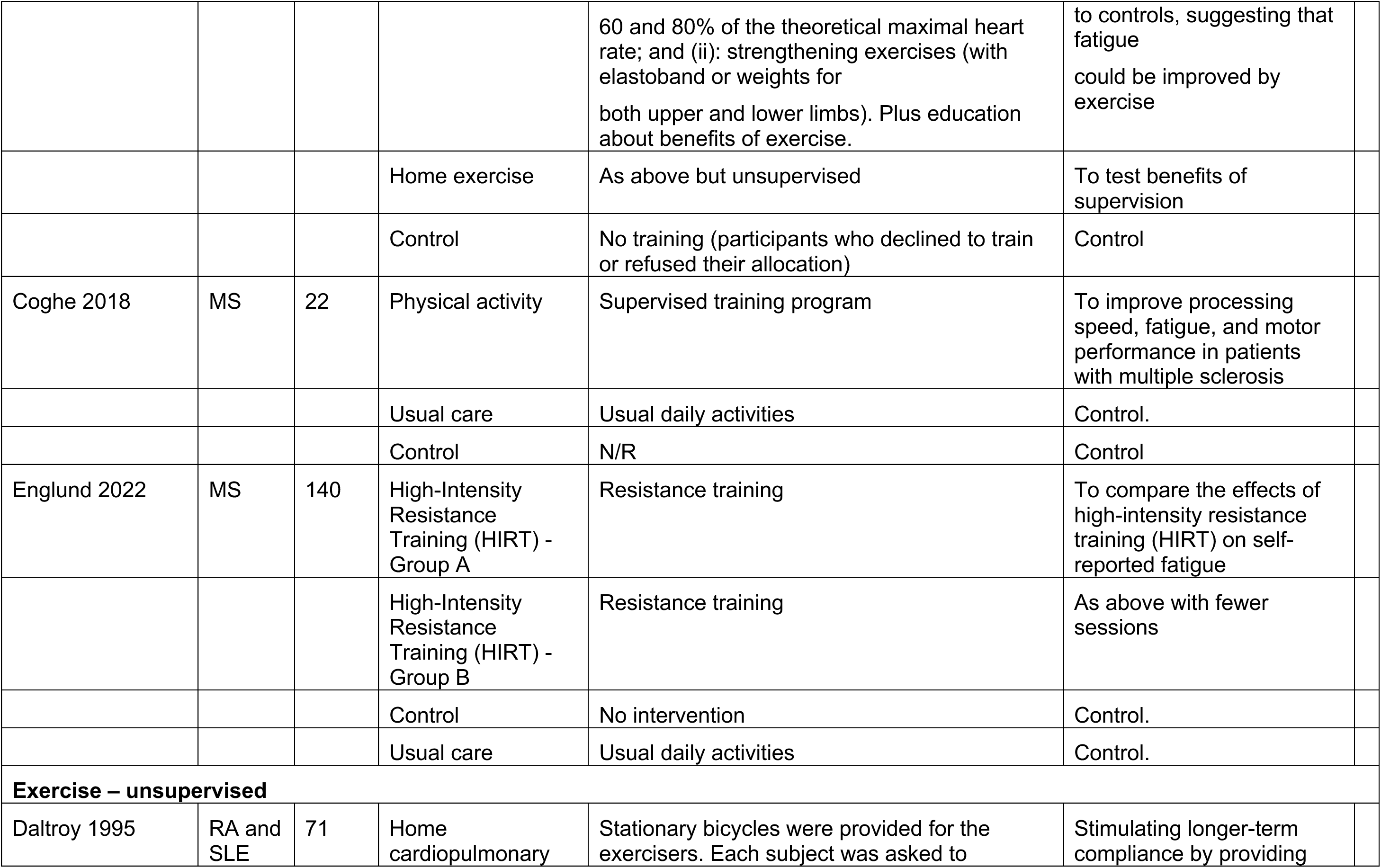
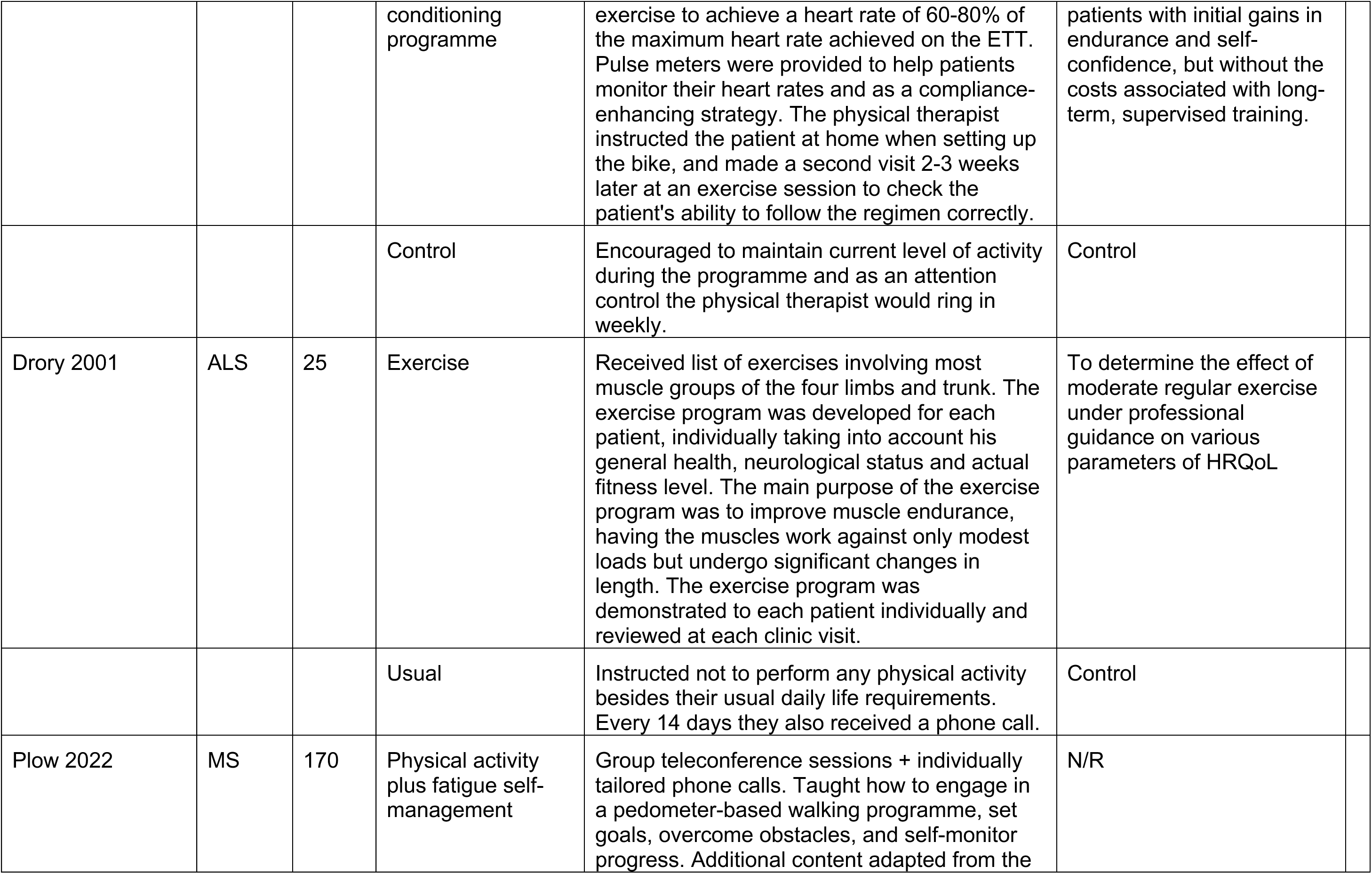
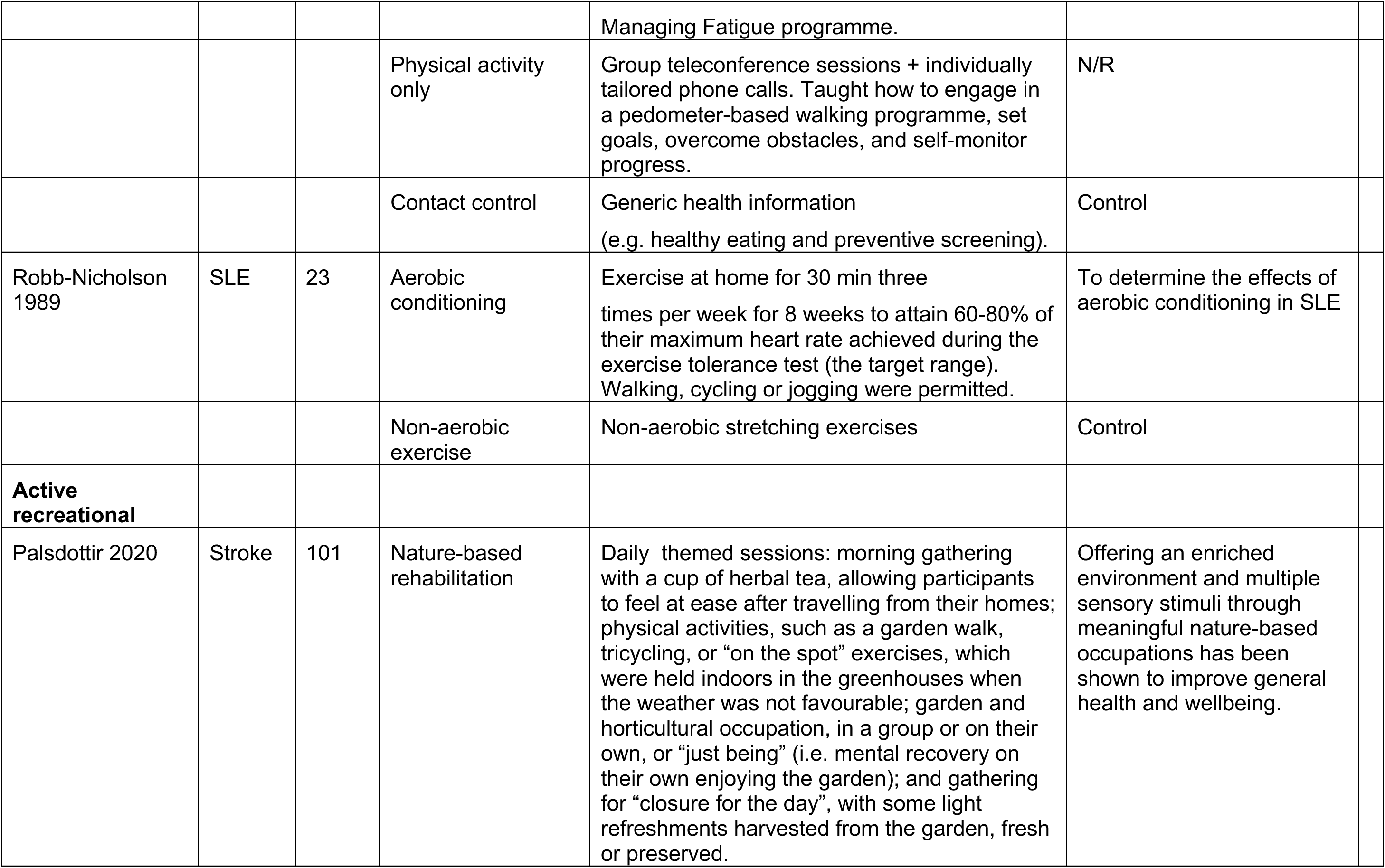
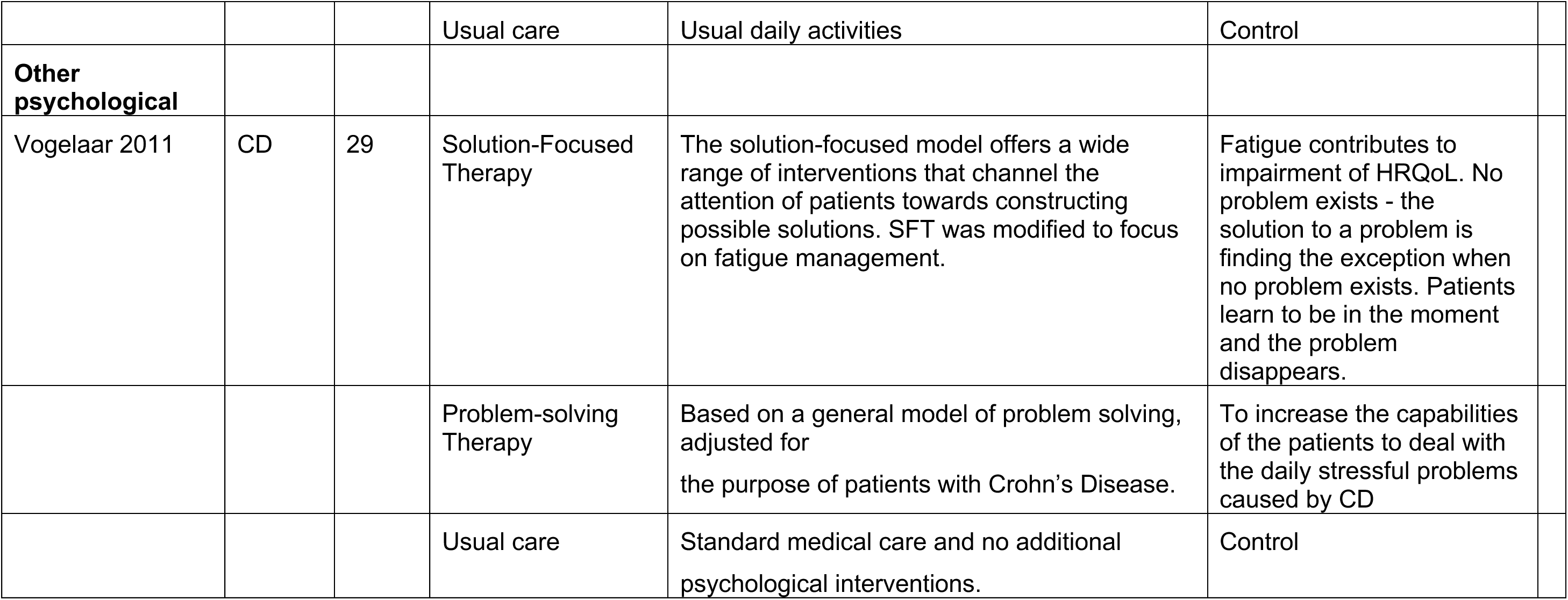
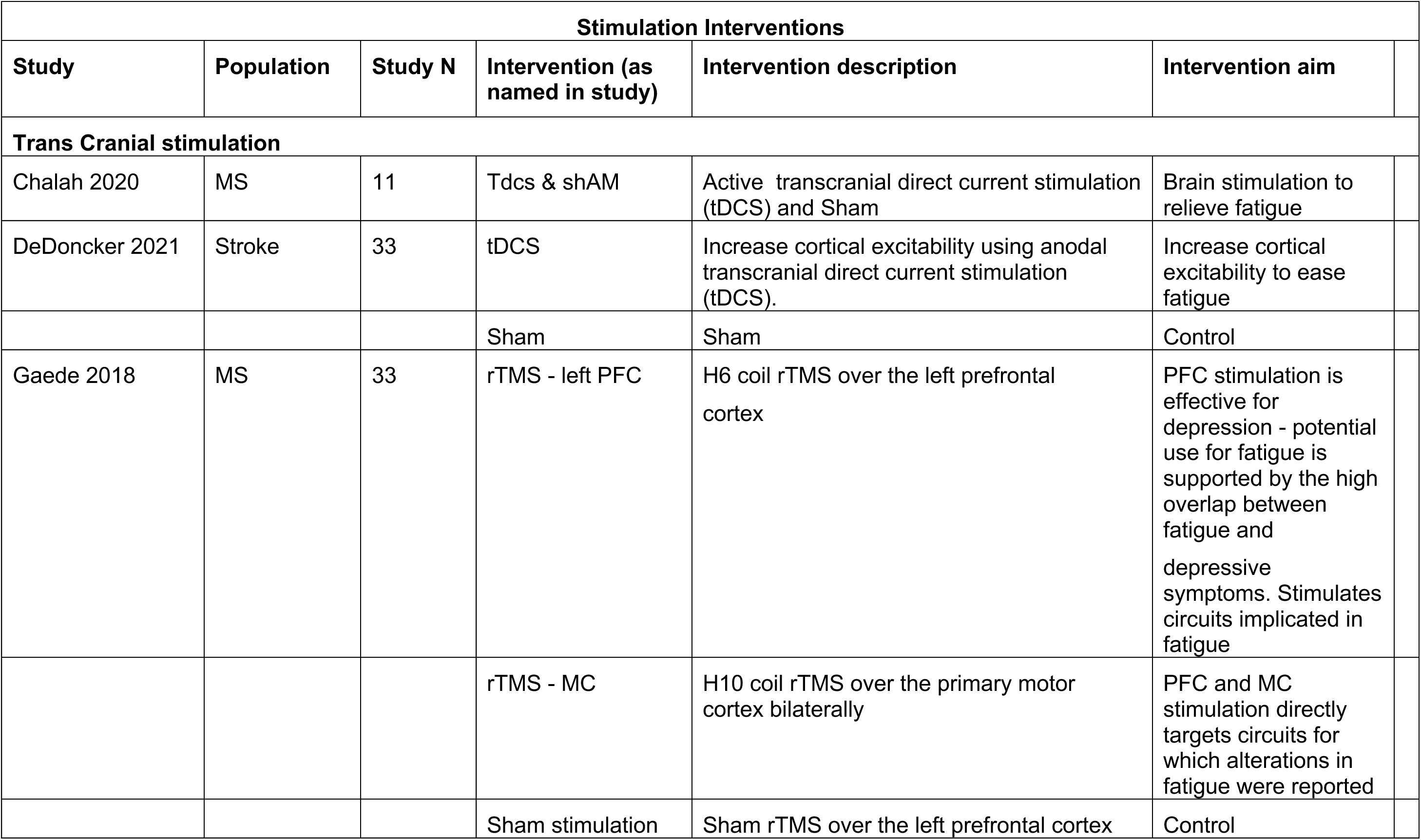
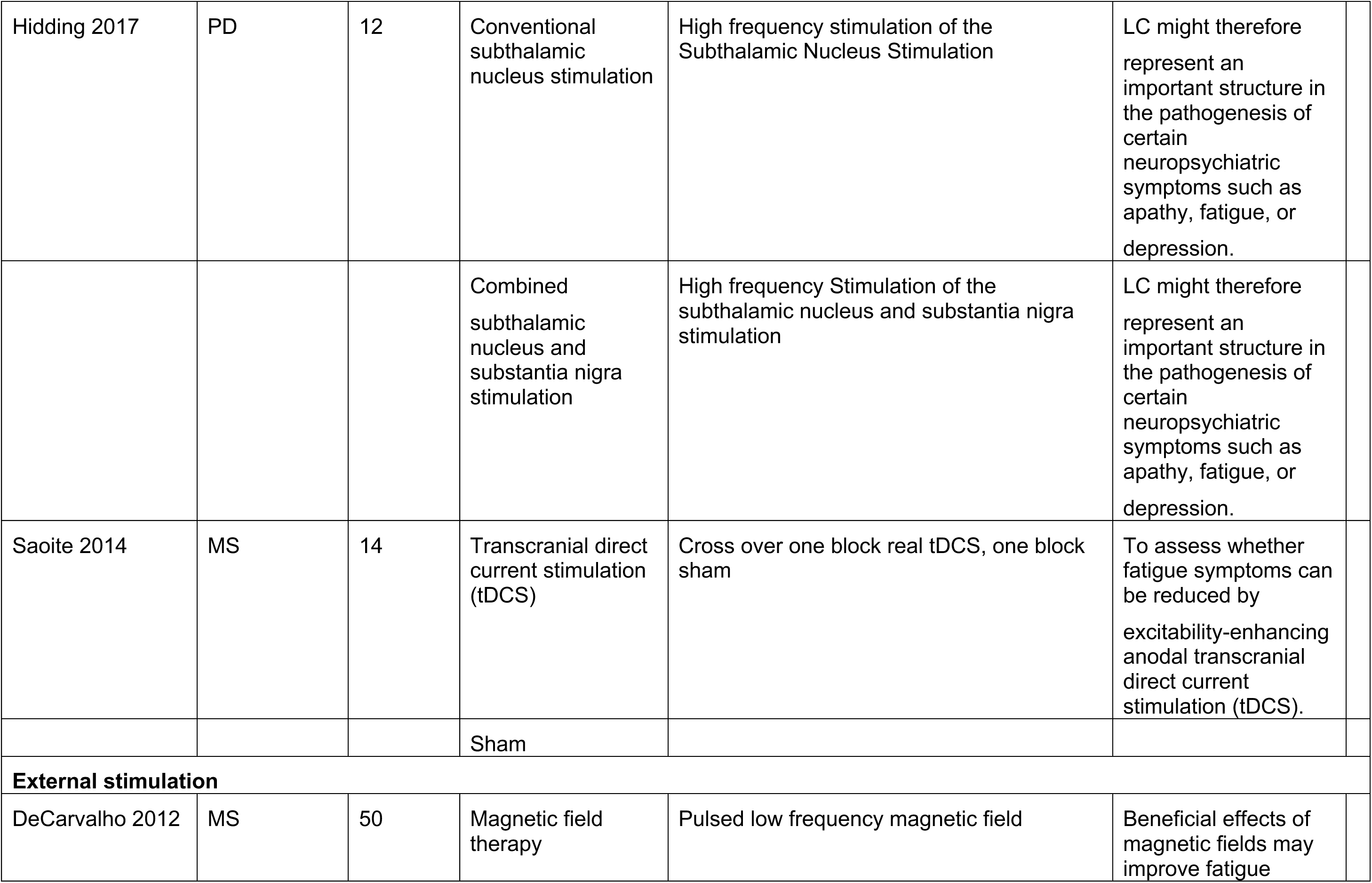
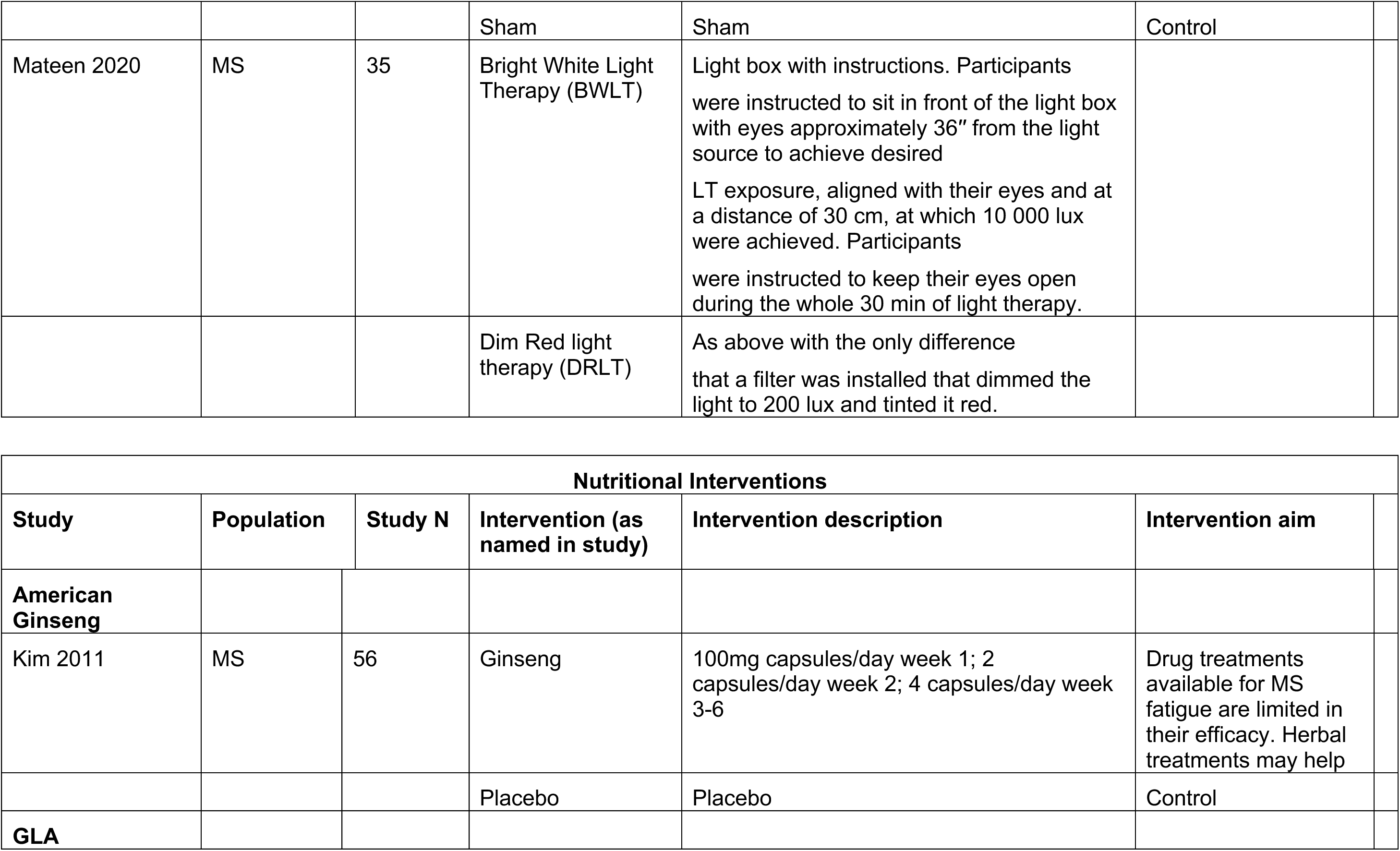
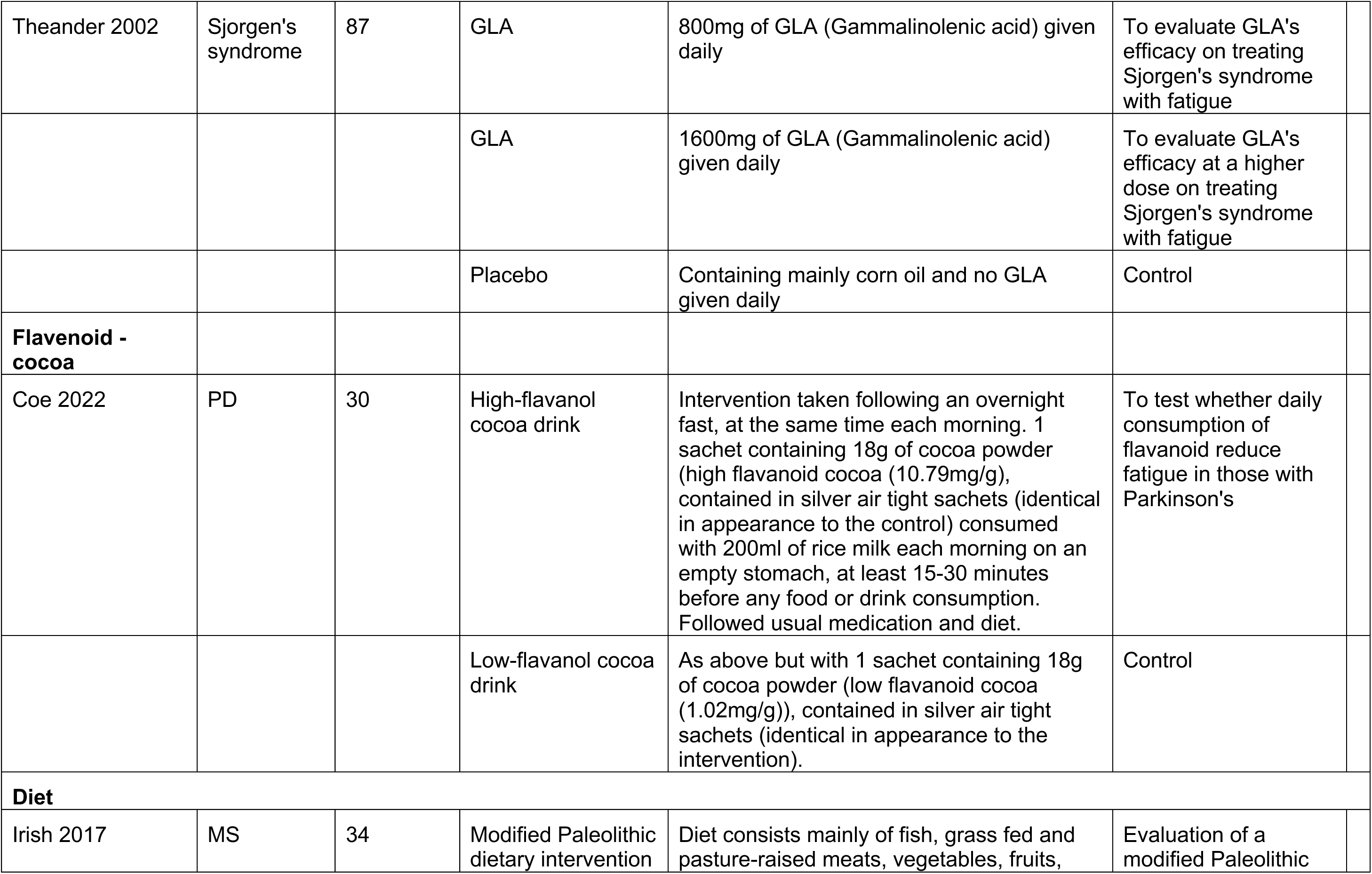
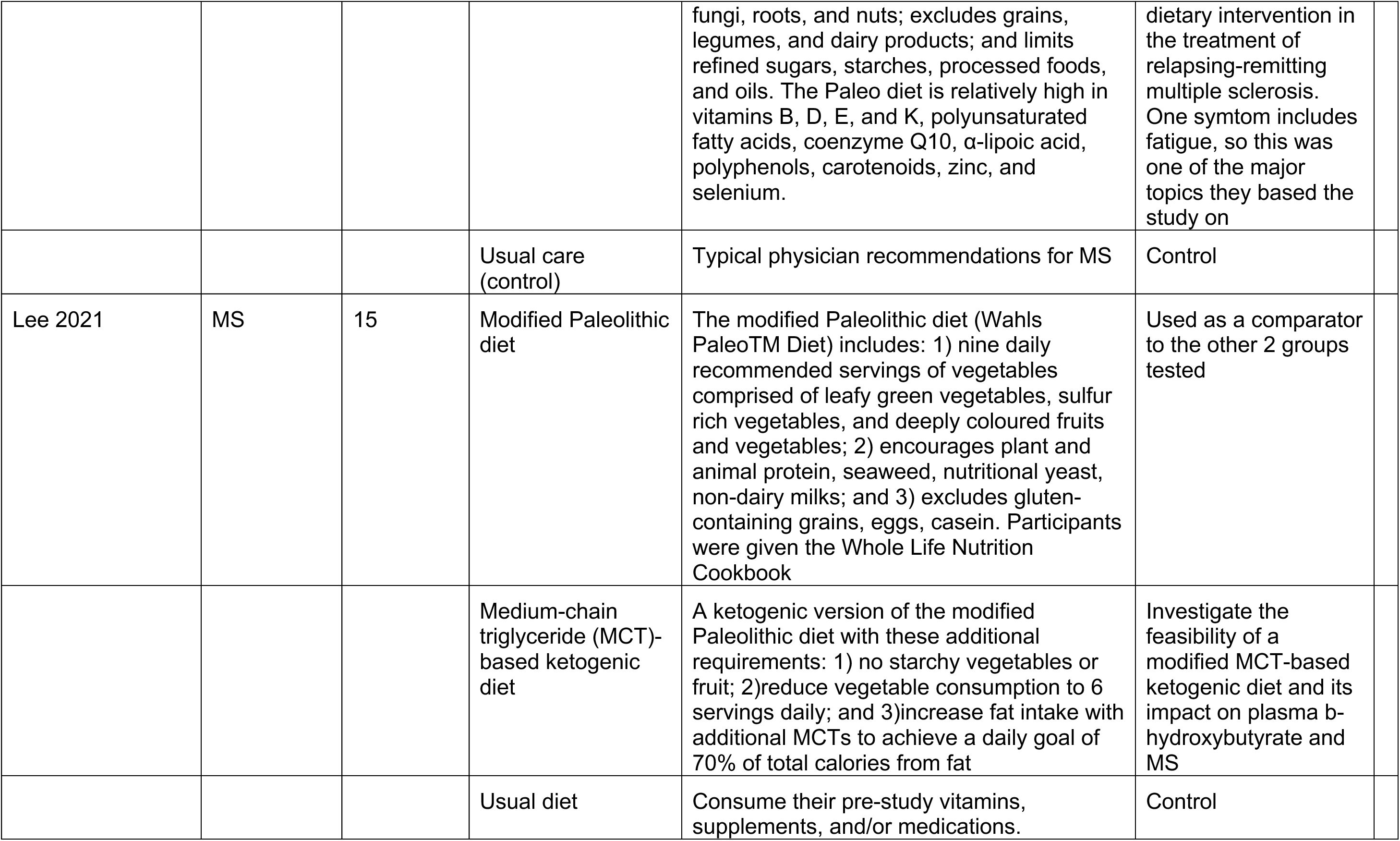

| Behavioural Interventions |  |  |  |  |  |
| --- | --- | --- | --- | --- | --- |
| Study/<br>Population | Pop. | Study<br>N | Intervention (as<br>named in study) | Intervention description | Intervention aim |
| **Self-Management** |  |  |  |  |  |
| **Fatigue self-management - conservative** |  |  |  |  |  |
| Finlayson 2011 | MS | 190 | Teleconference<br>Fatigue<br>Management<br>Program | Discussions about fatigue; how to communicate about fatigue; body mechanics; activity analysis - evaluating priorities; living a balanced life - taking control of your day; goal setting. | To teach behavioural changes that will lead to improvement in fatigue severity and HRQoL |
|  |  |  | Wait List | Usual daily activities | Control |
|  |  |  | Progressive<br>Muscle Relaxation<br>+RAU | A standardized series of relaxation exercises (involving 11 large muscle groups) combined with deep breathing + rehabilitation as usual | To achieve enhanced mental relaxation by reducing muscle tension |
| Kos 2007 | MS | 51 | Multidisciplinary<br>Fatigue<br>Management<br>Programme | Information concerning possible strategies to manage fatigue and reduced energy levels, ie, pharmacological treatment, diet, informing and involving the social environment, regular sleep, exercise, relaxation, cooling, assistive devices, adaptation of home or work environment and energy saving methods | To reduce the impact of MS fatigue on daily life |
|  |  |  | Placebo Intervention Programme | Information on topics that did not concern themes directly related to fatigue (ie, car adaptations and driving abilities, communication skills, lift techniques for back protection and general information about MS) | Active control |
| Mathiowetz 2005 | MS | 169 | Energy Conservation Course | Based on theory of psychoeducational group development. Long and short term goal setting; practice activities and homework. Importance of rest throughout the day, positive and effective communication; proper body mechanisms; ergonomic principles; modification of the environment; changing standards; setting priorities; activity analysis and modification; living a balanced lifestyle. | To determine whether energy conservation education can reduce the impact of fatigue in persons with MS |
|  |  |  | Wait list | Usual activities | A control |
| **Fatigue self-management - active** |  |  |  |  |  |
| O'Connor 2019 | IBD | 23 | Psychoeducation | Structured around psychological and physical interventions, which were geared towards understanding fatigue, energy conservation, management strategies and improving relaxation techniques tailored to the specific needs of patients with IBD. | To test whether or not fatigue, energy and quality of life indices could be improved |
|  |  |  | Usual care | Standard medical care | Control |
| Vogelaar 2014 | IBD | 98 | Solution Focused Therapy | Solution-focused course, focussing on coping styles for fatigue. Psychoeducation about IBD and fatigue and SFT. Focus is on the existing adequate coping abilities of patients, rather than on their problems. | To develop coping skills to enhance fatigue management |
|  |  |  | Usual care | Received care as usual | Control |

| **CBT – fatigue** |  |  |  |  |  |
| --- | --- | --- | --- | --- | --- |
| van Kessel 2016 | MS | 39 | MSInvigor8 + support | Interactive CBT self-management programme with email support from a clinical psychologist. Explanation of MS fatigue and the CBT approach; topics such as activity scheduling, improving sleep, altering unhelpful thinking and patterns and behaviour, managing stress, coping with emotions, social support, preparing for the future. | To test the addition of email support to the MSINVIGOR8 programme |
|  |  |  | MSInvigor8 | Interactive CBT self-management programme as above with no email support | Control. |
| Voet 2014 | FSHD | 57 | CBT | Modules based on known fatigue perpetuating factors. Directed at insufficient coping with their disease; dysfunctional cognitions regarding fatigue, activity, pain or other symptoms; fatigue catastrophising; dysregulation of sleep or activity; poor social support; negative social interactions. | To alleviate individually relevant fatigue-perpetuating factors. |
|  |  |  | Aerobic exercise | Cycling exercises on an ergometer, with cardiovascular monitoring. Aim to achieve 50%-60% increase in heart rate reserve. | To increase exercise which plays a central role in perpetuating fatigue |
| **Physical Activity** |  |  |  |  |  |
| **Physical activity promotion** |  |  |  |  |  |
| McNelly 2016 | IBD | 52 | Omega-3 and exercise | Individual consultation with a personal trainer provided at week 1. Advice consisted of personalised goal-setting using the treatment paradigm of treat-to-target to initiate an increase in physical activity levels of at least 30%. | To compare the effectiveness of individual advice to increase physical activity (PA) and/or supplementation with omega-3 fatty acids on fatigue in patients with inactive IBD |
|  |  |  | Placebo and exercise | A 15-minute conversation with the researcher about the participant's dietary habits and general health was undertaken at week 1, including questions such as: 'Can you tell me about your current dietN/R', 'Did you have to change your diet following the diagnosis of IBDN/R' and 'In what way has IBD affected your general healthN/R' No advice was given by the researcher regarding exercise. | As above |
|  |  |  | Omega-3 and no exercise | A total daily oral dose comprised 2970mg of pharmaceutical-grade omega-3 fatty acids — 2250mg of EPA and 150mg of DHA (takeOmega3, Edinburgh, UK)—in three capsules. Guidelines suggest that doses of up to 3g per day of marine-derived omega-3 fatty acids are safe, and a high EPA:DHA ratio is thought to be preferable. | As above |
|  |  |  | Placebo and no exercise | Capsules with a similar appearance to the omega-3 supplement capsules, but which contained a placebo: capric and caprylic acid. | Control |
| Callahan 2008 | RA | 346 | PACE exercise programme | A land-based exercise programme to promote self-management of arthritis through exercise | Exercise programs of moderate intensity are proposed to improve HRQoL in individuals with rheumatoid arthritis |
|  |  |  | Wait list | Usual activities | Control |
| **Exercise – supervised** |  |  |  |  |  |
| Avaux 2016 | SLE | 45 | Supervised exercise | Endurance exercises (walking or bicycle) with the aim of achieving between | SLE patients have a lower cardiovascular capacity and a lower muscle strength compared |
|  |  |  |  | 60 and 80% of the theoretical maximal heart rate; and (ii): strengthening exercises (with elastoband or weights for both upper and lower limbs). Plus education about benefits of exercise. | to controls, suggesting that fatigue could be improved by exercise |
|  |  |  | Home exercise | As above but unsupervised | To test benefits of supervision |
|  |  |  | Control | No training (participants who declined to train or refused their allocation) | Control |
| Coghe 2018 | MS | 22 | Physical activity | Supervised training program | To improve processing speed, fatigue, and motor performance in patients with multiple sclerosis |
|  |  |  | Usual care | Usual daily activities | Control. |
|  |  |  | Control | N/R | Control |
| Englund 2022 | MS | 140 | High-Intensity Resistance Training (HIRT) - Group A | Resistance training | To compare the effects of high-intensity resistance training (HIRT) on self-reported fatigue |
|  |  |  | High-Intensity Resistance Training (HIRT) - Group B | Resistance training | As above with fewer sessions |
|  |  |  | Control | No intervention | Control. |
|  |  |  | Usual care | Usual daily activities | Control. |
| **Exercise – unsupervised** |  |  |  |  |  |
| Daltroy 1995 | RA and SLE | 71 | Home cardiopulmonary | Stationary bicycles were provided for the exercisers. Each subject was asked to | Stimulating longer-term compliance by providing |
|  |  |  | conditioning programme | exercise to achieve a heart rate of 60-80% of the maximum heart rate achieved on the ETT. Pulse meters were provided to help patients monitor their heart rates and as a compliance-enhancing strategy. The physical therapist instructed the patient at home when setting up the bike, and made a second visit 2-3 weeks later at an exercise session to check the patient's ability to follow the regimen correctly. | patients with initial gains in endurance and self-confidence, but without the costs associated with long-term, supervised training. |
|  |  |  | Control | Encouraged to maintain current level of activity during the programme and as an attention control the physical therapist would ring in weekly. | Control |
| Drory 2001 | ALS | 25 | Exercise | Received list of exercises involving most muscle groups of the four limbs and trunk. The exercise program was developed for each patient, individually taking into account his general health, neurological status and actual fitness level. The main purpose of the exercise program was to improve muscle endurance, having the muscles work against only modest loads but undergo significant changes in length. The exercise program was demonstrated to each patient individually and reviewed at each clinic visit. | To determine the effect of moderate regular exercise under professional guidance on various parameters of HRQoL |
|  |  |  | Usual | Instructed not to perform any physical activity besides their usual daily life requirements. Every 14 days they also received a phone call. | Control |
| Plow 2022 | MS | 170 | Physical activity plus fatigue self-management | Group teleconference sessions + individually tailored phone calls. Taught how to engage in a pedometer-based walking programme, set goals, overcome obstacles, and self-monitor progress. Additional content adapted from the | N/R |
|  |  |  |  | Managing Fatigue programme. |  |
|  |  |  | Physical activity only | Group teleconference sessions + individually tailored phone calls. Taught how to engage in a pedometer-based walking programme, set goals, overcome obstacles, and self-monitor progress. | N/R |
|  |  |  | Contact control | Generic health information (e.g. healthy eating and preventive screening). | Control |
| Robb-Nicholson 1989 | SLE | 23 | Aerobic conditioning | Exercise at home for 30 min three times per week for 8 weeks to attain 60-80% of their maximum heart rate achieved during the exercise tolerance test (the target range). Walking, cycling or jogging were permitted. | To determine the effects of aerobic conditioning in SLE |
|  |  |  | Non-aerobic exercise | Non-aerobic stretching exercises | Control |
| **Active recreational** |  |  |  |  |  |
| Palsdottir 2020 | Stroke | 101 | Nature-based rehabilitation | Daily themed sessions: morning gathering with a cup of herbal tea, allowing participants to feel at ease after travelling from their homes; physical activities, such as a garden walk, tricycling, or “on the spot” exercises, which were held indoors in the greenhouses when the weather was not favourable; garden and horticultural occupation, in a group or on their own, or “just being” (i.e. mental recovery on their own enjoying the garden); and gathering for “closure for the day”, with some light refreshments harvested from the garden, fresh or preserved. | Offering an enriched environment and multiple sensory stimuli through meaningful nature-based occupations has been shown to improve general health and wellbeing. |
|  |  |  | Usual care | Usual daily activities | Control |
| **Other psychological** |  |  |  |  |  |
| Vogelaar 2011 | CD | 29 | Solution-Focused Therapy | The solution-focused model offers a wide range of interventions that channel the attention of patients towards constructing possible solutions. SFT was modified to focus on fatigue management. | Fatigue contributes to impairment of HRQoL. No problem exists - the solution to a problem is finding the exception when no problem exists. Patients learn to be in the moment and the problem disappears. |
|  |  |  | Problem-solving Therapy | Based on a general model of problem solving, adjusted for the purpose of patients with Crohn's Disease. | To increase the capabilities of the patients to deal with the daily stressful problems caused by CD |
|  |  |  | Usual care | Standard medical care and no additional psychological interventions. | Control |

| Stimulation Interventions |  |  |  |  |  |
| --- | --- | --- | --- | --- | --- |
| Study | Population | Study N | Intervention (as named in study) | Intervention description | Intervention aim |
| **Trans Cranial stimulation** |  |  |  |  |  |
| Chalah 2020 | MS | 11 | Tdcs & shAM | Active transcranial direct current stimulation (tDCS) and Sham | Brain stimulation to relieve fatigue |
| DeDoncker 2021 | Stroke | 33 | tDCS | Increase cortical excitability using anodal transcranial direct current stimulation (tDCS). | Increase cortical excitability to ease fatigue |
|  |  |  | Sham | Sham | Control |
| Gaede 2018 | MS | 33 | rTMS - left PFC | H6 coil rTMS over the left prefrontal cortex | PFC stimulation is effective for depression - potential use for fatigue is supported by the high overlap between fatigue and depressive symptoms. Stimulates circuits implicated in fatigue |
|  |  |  | rTMS - MC | H10 coil rTMS over the primary motor cortex bilaterally | PFC and MC stimulation directly targets circuits for which alterations in fatigue were reported |
|  |  |  | Sham stimulation | Sham rTMS over the left prefrontal cortex | Control |
| Hidding 2017 | PD | 12 | Conventional subthalamic nucleus stimulation | High frequency stimulation of the Subthalamic Nucleus Stimulation | LC might therefore represent an important structure in the pathogenesis of certain neuropsychiatric symptoms such as apathy, fatigue, or depression. |
|  |  |  | Combined subthalamic nucleus and substantia nigra stimulation | High frequency Stimulation of the subthalamic nucleus and substantia nigra stimulation | LC might therefore represent an important structure in the pathogenesis of certain neuropsychiatric symptoms such as apathy, fatigue, or depression. |
| Saoite 2014 | MS | 14 | Transcranial direct current stimulation (tDCS) | Cross over one block real tDCS, one block sham | To assess whether fatigue symptoms can be reduced by excitability-enhancing anodal transcranial direct current stimulation (tDCS). |
|  |  |  | Sham |  |  |
| **External stimulation** |  |  |  |  |  |
| DeCarvalho 2012 | MS | 50 | Magnetic field therapy | Pulsed low frequency magnetic field | Beneficial effects of magnetic fields may improve fatigue |
|  |  |  | Sham | Sham | Control |
| Mateen 2020 | MS | 35 | Bright White Light Therapy (BWLT) | Light box with instructions. Participants were instructed to sit in front of the light box with eyes approximately 36" from the light source to achieve desired<br>LT exposure, aligned with their eyes and at a distance of 30 cm, at which 10 000 lux were achieved. Participants<br>were instructed to keep their eyes open during the whole 30 min of light therapy. |  |
|  |  |  | Dim Red light therapy (DRLT) | As above with the only difference that a filter was installed that dimmed the light to 200 lux and tinted it red. |  |

| Nutritional Interventions |  |  |  |  |  |
| --- | --- | --- | --- | --- | --- |
| Study | Population | Study N | Intervention (as named in study) | Intervention description | Intervention aim |
| **American Ginseng** |  |  |  |  |  |
| Kim 2011 | MS | 56 | Ginseng | 100mg capsules/day week 1; 2 capsules/day week 2; 4 capsules/day week 3-6 | Drug treatments available for MS fatigue are limited in their efficacy. Herbal treatments may help |
|  |  |  | Placebo | Placebo | Control |
| **GLA** |  |  |  |  |  |
| Theander 2002 | Sjorgen's syndrome | 87 | GLA | 800mg of GLA (Gammalinolenic acid) given daily | To evaluate GLA's efficacy on treating Sjorgen's syndrome with fatigue |
|  |  |  | GLA | 1600mg of GLA (Gammalinolenic acid) given daily | To evaluate GLA's efficacy at a higher dose on treating Sjorgen's syndrome with fatigue |
|  |  |  | Placebo | Containing mainly corn oil and no GLA given daily | Control |
| **Flavenoid - cocoa** |  |  |  |  |  |
| Coe 2022 | PD | 30 | High-flavanol cocoa drink | Intervention taken following an overnight fast, at the same time each morning. 1 sachet containing 18g of cocoa powder (high flavanoid cocoa (10.79mg/g), contained in silver air tight sachets (identical in appearance to the control) consumed with 200ml of rice milk each morning on an empty stomach, at least 15-30 minutes before any food or drink consumption. Followed usual medication and diet. | To test whether daily consumption of flavanoid reduce fatigue in those with Parkinson's |
|  |  |  | Low-flavanol cocoa drink | As above but with 1 sachet containing 18g of cocoa powder (low flavanoid cocoa (1.02mg/g)), contained in silver air tight sachets (identical in appearance to the intervention). | Control |
| **Diet** |  |  |  |  |  |
| Irish 2017 | MS | 34 | Modified Paleolithic dietary intervention | Diet consists mainly of fish, grass fed and pasture-raised meats, vegetables, fruits, | Evaluation of a modified Paleolithic |

|  |  |  |  |  |  |  |
| --- | --- | --- | --- | --- | --- | --- |
| | | | | fungi, roots, and nuts; excludes grains, legumes, and dairy products; and limits refined sugars, starches, processed foods, and oils. The Paleo diet is relatively high in vitamins B, D, E, and K, polyunsaturated fatty acids, coenzyme Q10, α-lipoic acid, polyphenols, carotenoids, zinc, and selenium. | dietary intervention in the treatment of relapsing-remitting multiple sclerosis. One symptom includes fatigue, so this was one of the major topics they based the study on | |
|  |  |  | Usual care (control) | Typical physician recommendations for MS | Control |  |
| Lee 2021 | MS | 15 | Modified Paleolithic diet | The modified Paleolithic diet (Wahls Paleo<sup>TM</sup> Diet) includes: 1) nine daily recommended servings of vegetables comprised of leafy green vegetables, sulfur rich vegetables, and deeply coloured fruits and vegetables; 2) encourages plant and animal protein, seaweed, nutritional yeast, non-dairy milks; and 3) excludes gluten-containing grains, eggs, casein. Participants were given the Whole Life Nutrition Cookbook | Used as a comparator to the other 2 groups tested |  |
| | | | Medium-chain triglyceride (MCT)-based ketogenic diet | A ketogenic version of the modified Paleolithic diet with these additional requirements: 1) no starchy vegetables or fruit; 2) reduce vegetable consumption to 6 servings daily; and 3) increase fat intake with additional MCTs to achieve a daily goal of 70% of total calories from fat | Investigate the feasibility of a modified MCT-based ketogenic diet and its impact on plasma β-hydroxybutyrate and MS | |
|  |  |  | Usual diet | Consume their pre-study vitamins, supplements, and/or medications. | Control |  |

#### 7.2 Intervention delivery

| Behavioural Interventions |  |  |  |  |  |  |
| --- | --- | --- | --- | --- | --- | --- |
| Study/<br>Population | Individual/<br>group | Setting | Number of<br>sessions | Duration of<br>sessions | Total duration of<br>intervention | Intervention<br>provider |
| **Self-Management** |  |  |  |  |  |  |
| **Fatigue self-management - conservative** |  |  |  |  |  |  |
| Finlayson 2011 | G | Teleconference | 6 | 70 minutes | 6 weeks | Licensed<br>occupational<br>therapist |
|  | I | N/A | N/A | N/A | 6 weeks | N/A |
| Kos 2007 | G | Face to face | 4 | 2 hours | 4 weeks | Occupational<br>therapist |
|  | G | Face to face | 4 | 2 hours | 4 weeks | Occupational<br>therapist |
| Mathiowetz 2005 | G | Community | 6 | 2 hours | 6 weeks | Occupational<br>therapists |
|  | N/A | N/A | N/A | N/A | 6 weeks | N/A |
| **Fatigue self-management - active** |  |  |  |  |  |  |
| O'Connor 2019 | G | Face to face | 3 | 1 hour | 6 months | Occupational<br>therapist |
|  | N/A | N/A | N/A | N/A | 6 months | N/A |
| Vogelaar 2014 | G | Face to face | 6 + booster at<br>month 6 | 1.5 hours | 3 months | N/R |
|  | N/A | N/A | N/A | N/A | 3 months | N/A |
| **CBT – fatigue** |  |  |  |  |  |  |
| van Kessel 2016 | I | Web-based | 8 | 25 to 50 minutes | 8 weeks | Web-site with email support from a skilled clinical psychologist |
|  | I | Web-based | 8 | 25 to 50 minutes | 8 weeks | Web-site with no therapeutic contact |
| Voet 2014 | I | Face to face | Minimum 3 sessions | 50 minutes | 16 weeks | Cognitive behaviour therapist |
|  | I | Home and supervised | 3 supervised sessions and minimum 40 home sessions | 30 minutes | 16 weeks | Physical therapist |
|  | N/A | N/A | N/A | N/A | 16 weeks | N/A |
| **Physical Activity** |  |  |  |  |  |  |
| **Physical activity promotion** |  |  |  |  |  |  |
| McNelly 2016 | I | Home-based | N/R | N/R | 12 weeks | N/R |
|  | I | Home-based | N/R | N/R | 12 weeks | N/R |
|  | I | Home-based | N/R | N/R | 12 weeks | N/R |
|  | I | Home-based | N/R | N/R | 12 weeks | N/R |
| Callahan 2008 | G | Community | At least one class | N/R | 8 weeks | Exercise and health professionals |
|  | N/A | N/A | N/A | N/A | 8 weeks | N/A |
| **Exercise – supervised** |  |  |  |  |  |  |
| Avaux 2016 | I | Hospital validation centre | Individualised | 3 hours per week | 12 weeks | Multidisciplinary team |
|  | I | Home | Individualised | 3 hours per week | 12 weeks | Unsupervised |
|  | N/A | N/A | N/A | N/A | 12 weeks | N/A |
| Coghe 2018 | G | N/R | 72 sessions | 60 minutes | 3 sessions per week for 24 weeks | Two coaches specializing in physical activity |
|  | N/A | N/A | N/A | N/A | 24 weeks | N/A |
| Englund 2022 | G | Karolinska University Hospital, Stockholm, Sweden | 24 sessions | 60 minutes | 2 sessions per week for 12 weeks | Physiotherapist |
|  | G | Karolinska University Hospital, Stockholm, Sweden | 12 sessions | 60 minutes | 1 sessions per week for 12 weeks | Physiotherapist |
|  | N/A | N/A | N/A | N/A | N/A | N/A |
| **Exercise – unsupervised** |  |  |  |  |  |  |
| Daltroy 1995 | I | Home-based + physical therapist | 36 sessions | 30 mins | 12 weeks | N/R |
|  | I | Home-based + physical therapist | N/R | N/R | 12 weeks | N/R |
| Drory 2001 | I | Home-based | 2 sessions/day | 15 mins | 12 months | N/R |
|  | I | N/A | N/A | N/A | 12 months | N/A |
| Plow 2022 | G | Telephone | 10 | N/R | 12 weeks | Occupational therapist + research assistant |
|  | G | Telephone | 10 | N/R | 12 weeks | Occupational therapist + research assistant |
|  | G | Telephone | 10 | N/R | 12 weeks | Occupational therapist + research assistant |
| Robb-Nicholson 1989 | I | Home-based | 24 | 30 minutes | 8 weeks | Unsupervised |
|  | I | Home-based | 24 | 30 minutes | 8 weeks | N/A |
| **Active recreational** |  |  |  |  |  |  |
| Palsdottir 2020 | G | Alnarp Rehabilitation garden | 20 | 3.5 hours | 10 weeks | Occupational therapist; horticulturalist; psychologist; physiotherapist |
|  |  | N/A | N/A | N/A | 10 weeks | Individualised multidisciplinary care |
| **Other psychological** |  |  |  |  |  |  |
| Vogelaar 2011 | N/R | Outpatients | 5 sessions | N/R | 3 months | Experienced psychotherapist |
|  | N/R | Outpatients | 10 sessions | N/R | 3 months | Experienced psychotherapist |
|  | N/A | N/A | N/A | N/A | 3 months | N/A |

| Stimulation Interventions |  |  |  |  |  |  |
| --- | --- | --- | --- | --- | --- | --- |
| Study/<br>Population | Individual/ group | Setting | Number of<br>sessions | Duration of<br>sessions | Total duration of<br>intervention | Intervention<br>provider |
| **Transcranial Stimulation** |  |  |  |  |  |  |
| Chalah 2020 | I | Hospital | 5 real, 5 sham | 20 mins | 5 days of each with 3 week washout period in-between |  |
| DeDoncker 2021 | I |  | 2 | 20 mins | 1 day |  |
|  | I |  | 2 | 20 mins | 1 day |  |
| Gaede 2018 | I | Clinic | 3 sessions per week | 16 minutes | 6 weeks | N/R |
|  | I | Clinic | 3 sessions per week | 16 minutes | 6 weeks | N/R |
|  | I | Clinic | 3 sessions per week | 16 minutes | 6 weeks | N/R |
| Hidding 2017 | I | Clinic | N/R | N/R | 3 weeks | N/R |
|  | I | Clinic | N/R | N/R | 3 weeks | N/R |
| Saoite 2014 | I | Clinic | 5 | 20 minutes | 5 days/ 2 weeks wash-out/ 5 days |  |
|  | I |  |  |  |  |  |
| **External stimulation** |  |  |  |  |  |  |
| DeCarvalho 2012 | I | Outpatient dept. | 24 | 24 mins | 8 weeks |  |
|  | I |  |  |  |  |  |
| Mateen 2020 | I | Clinic/Home | 15 | 30 mins | 15 days |  |
|  | I |  |  |  |  |  |

| **Nutritional Interventions** |  |  |  |  |  |  |
| --- | --- | --- | --- | --- | --- | --- |
| **Study/**<br>**Population** | **Individual/ group** | **Setting** | **Number of**<br>**sessions** | **Duration of**<br>**sessions** | **Total duration of**<br>**intervention** | **Intervention**<br>**provider** |
| **American**<br>**Ginseng** |  |  |  |  |  |  |
| Kim 2011 | I | Home-based | N/A | N/A | 6 weeks | Afexa Life Sciences, Edmonton, Canada |
|  | I | Home-based | N/A | N/A | 6 weeks | Afexa Life Sciences, Edmonton, Canada |
| **GLA** |  |  |  |  |  |  |
| Theander 2002 | I | Home-based | N/A | N/A | 6 months | Scotia Pharmaceutical Ltd., Guilford, Surrey, UK |
|  | I | Home-based | N/A | N/A | 6 months | Scotia Pharmaceutical Ltd., Guilford, Surrey, UK |
|  | I | Home-based | N/A | N/A | 6 months | Scotia Pharmaceutical Ltd., Guilford, Surrey, UK |
| **Flavanoid - cocoa** |  |  |  |  |  |  |
| Coe 2022 | I | A hotel | N/A | N/A | 6 days | OBU in the Oxford Brookes Centre for Nutrition and Health (OxBCNH) kitchen |
|  | I | A hotel | N/A | N/A | 6 days | OBU in the Oxford Brookes Centre for Nutrition and Health (OxBCNH) kitchen |
| **Diet** |  |  |  |  |  |  |
| Irish 2017 | I | Home-based plus visit every 2 weeks | Visit every 2 weeks visit | N/R | 3 months | N/R |
|  | I | Home-based plus visit every 2 weeks | Visit every 2 weeks visit | N/R | 3 months | N/R |
| Lee 2021 | I | Home based | Nutritional ketosis monitoring every 4 weeks | N/R | 12 weeks | Wahls Paleo Diet + dietician |
|  | I | Home based | Nutritional ketosis monitoring every 4 weeks | N/R | 12 weeks | Wahls Paleo Plus + dietician |
|  | I | Home based | Nutritional ketosis monitoring every 4 weeks | N/R | 12 weeks | Dietician |

### Supplemental Results 8. Risk of Bias summary plots by intervention group

#### 8.1 CBT-based interventions

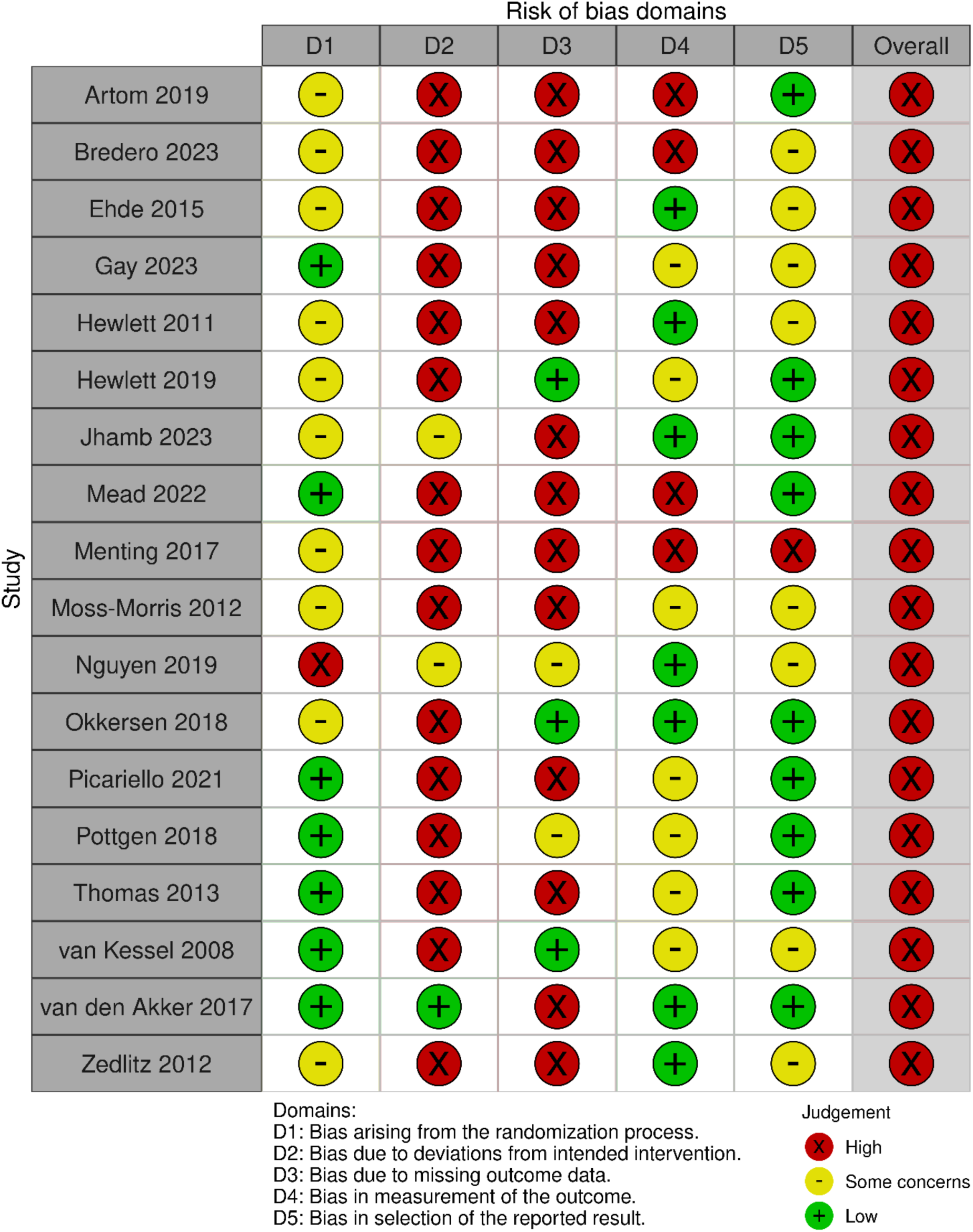

#### 8.2 Fatigue self-management interventions

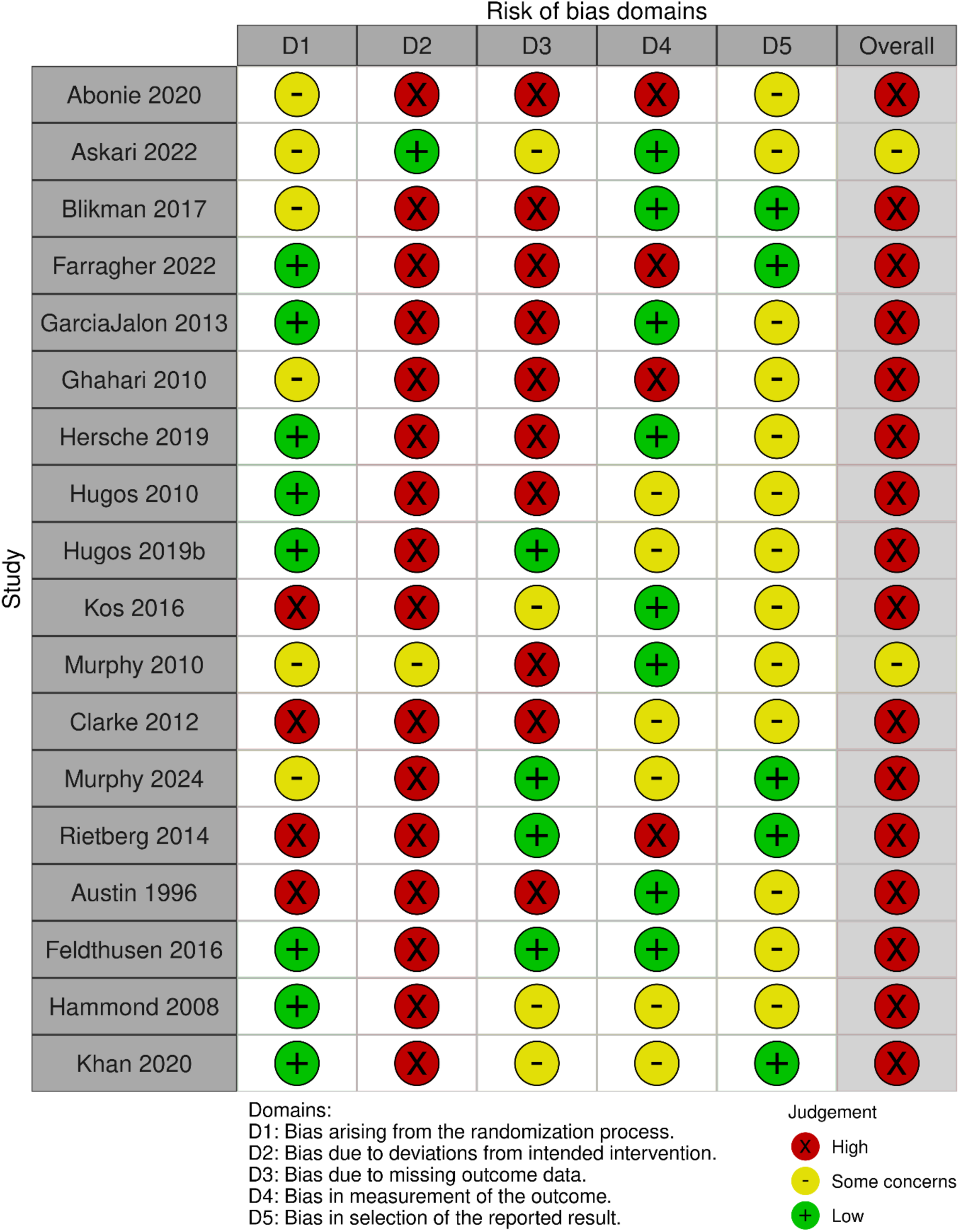

#### 8.3 Mind-body interventions

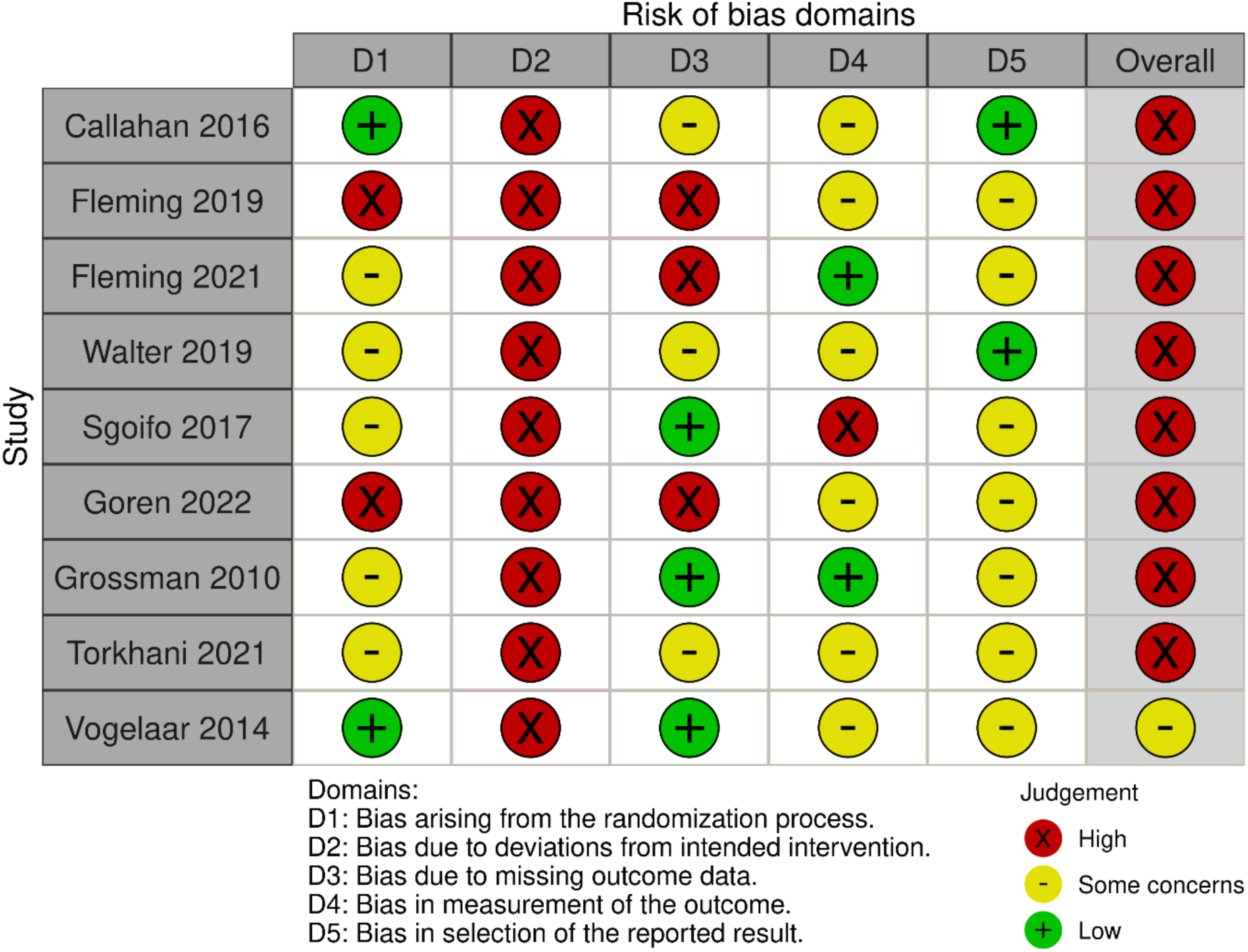

#### 8.4 Physical Activity Promotion Interventions

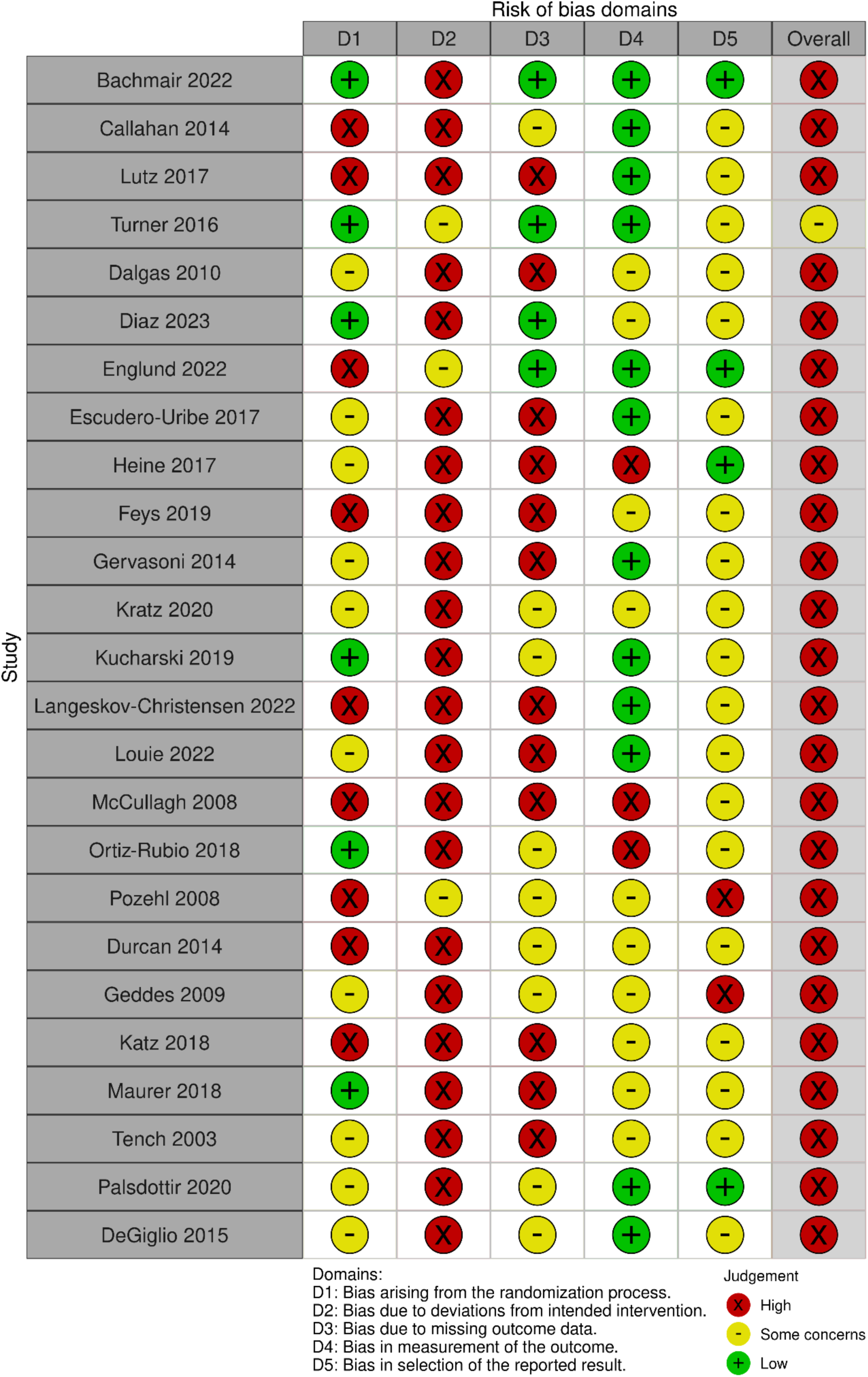

#### 8.5 External stimulation interventions

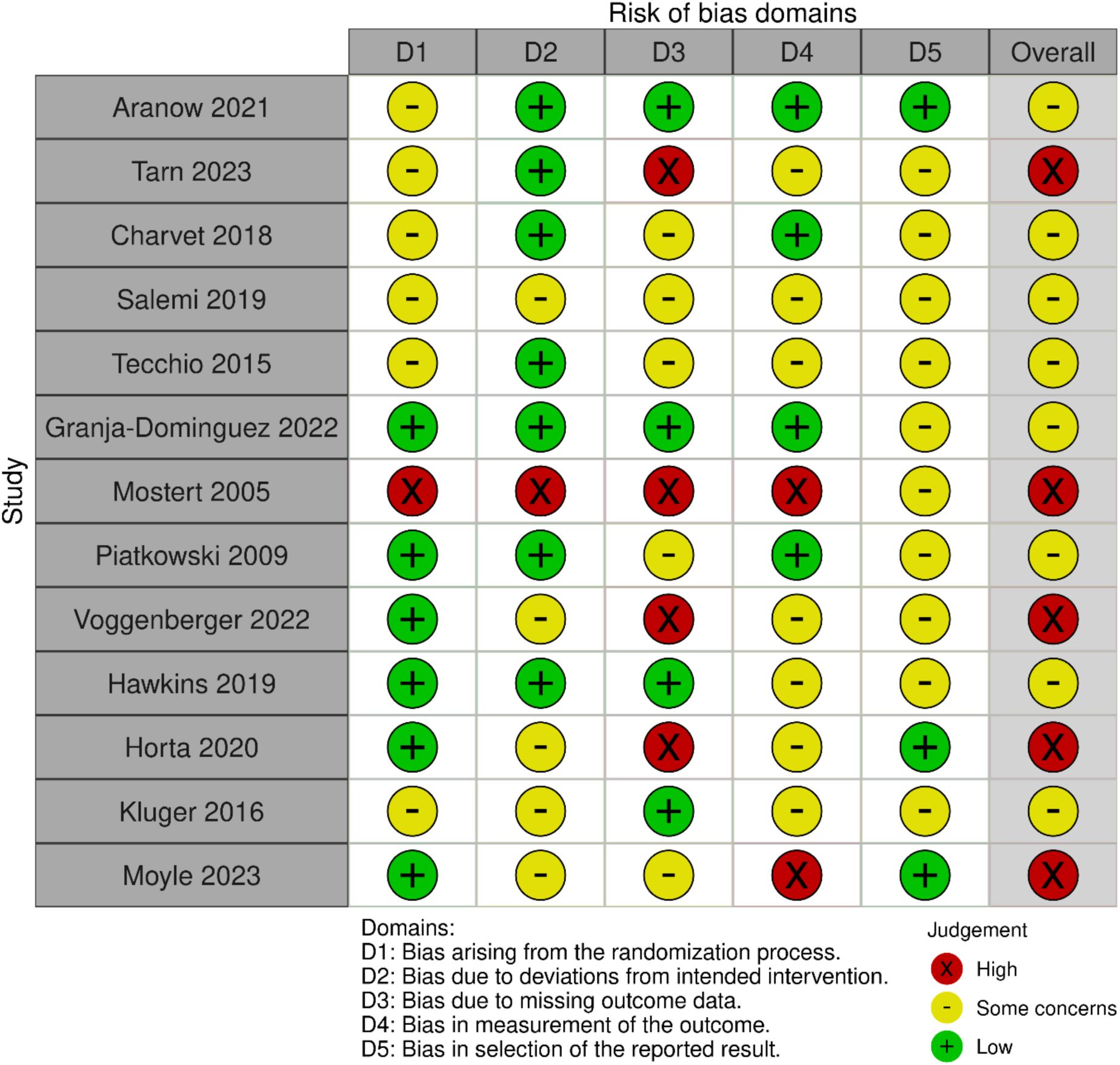

#### 8.6 Nutritional and other supplement interventions

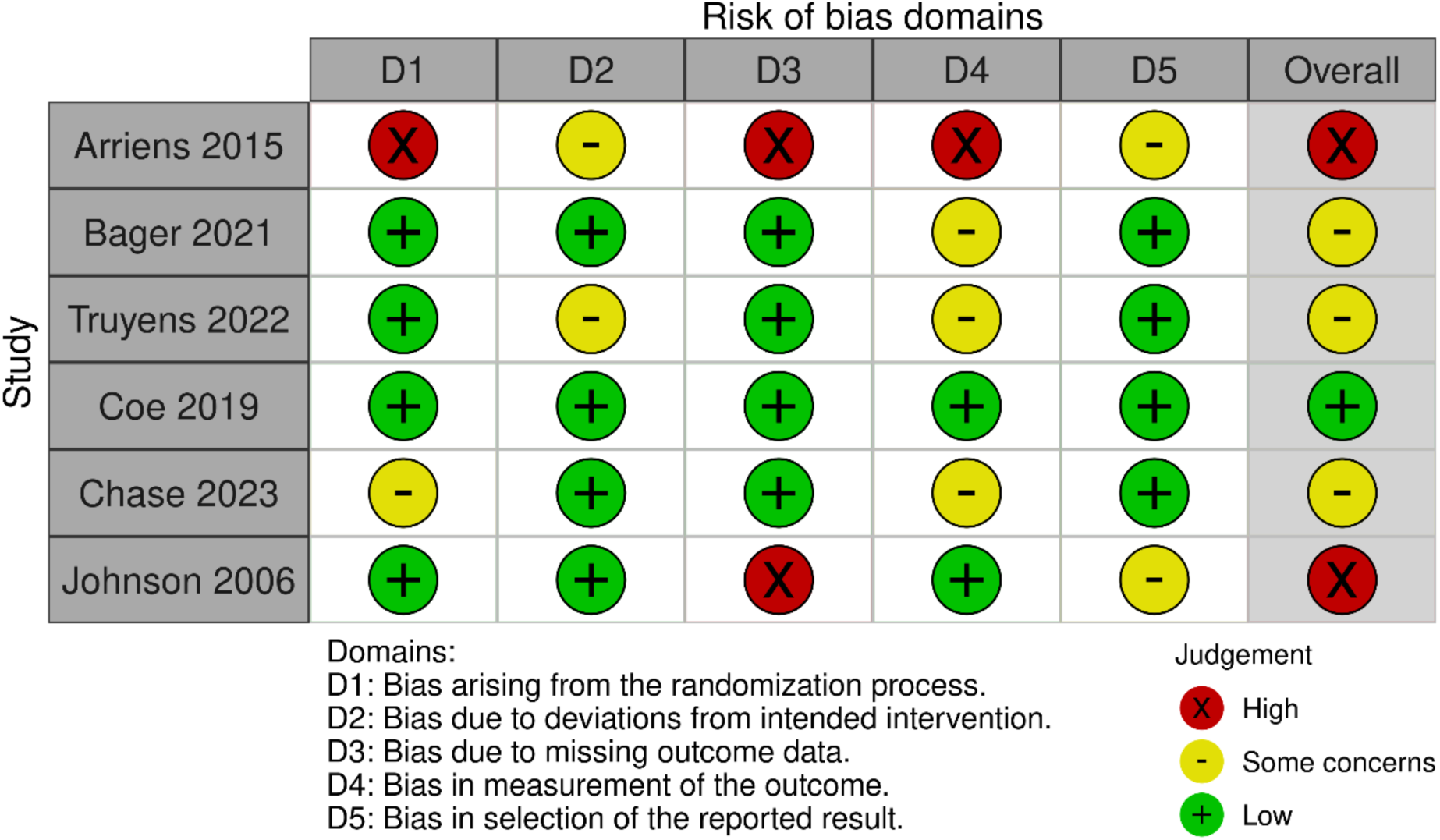

### EIFFEL Supplementary Statistical Analysis Results

All network diagrams were labelled with three letter intervention identifiers, as summarized in Table 1.

**Table 1.**
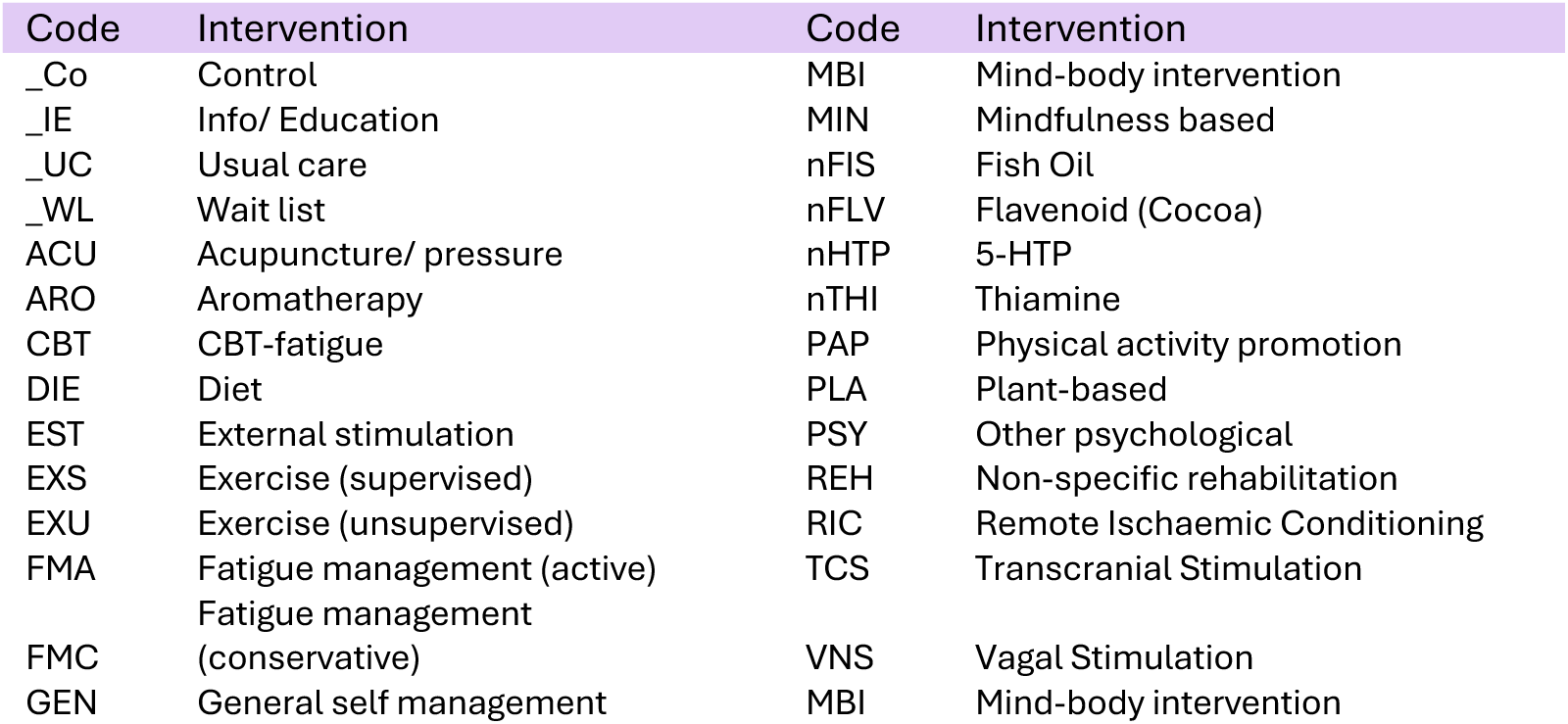
Three letter intervention identifier codes included within network diagrams.

#### Primary analysis: inconsistency checks

##### End of treatment (EOT)

The mean posterior residual deviances were compared between the unrelated mean effects model and NMA models, Figure 1. The following studies were identified as being below the *y* = *x* line indicating potential inconsistency within the network; Louie 2022^1^, Fleming 2021^2^ Menting 2017^3^, Langeskov-Christensen 2022^4^, Turner 2016^5^ and Horta 2020^6^. The studies were checked for any errors in data extraction or noticeable population differences, but none were identified. Node-splitting was subsequently used to assess whether there was any statistically significant inconsistency within the network, none was identified and thus no further action was taken.

**Figure 1.**
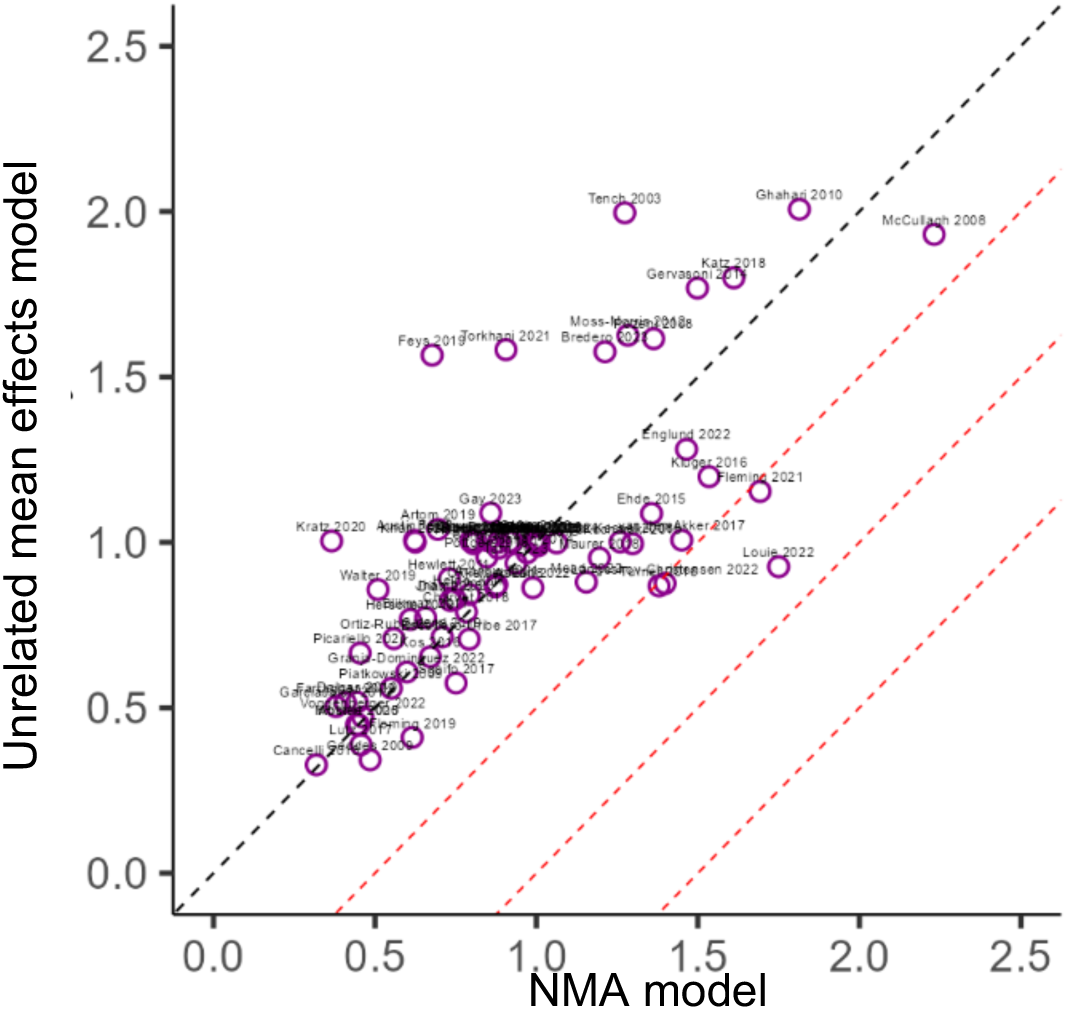
Mean posterior residual deviances according to the unrelated mean effects model and the NMA model, at end of treatment. Black dashed line is given by *y* = *x*, red dashed lines represent contours separated by differences of 0.5 between the two models. Any studies below the first red dashed line indicative of potential inconsistency.

##### Short term (ST)

The mean posterior residual deviances were compared between the unrelated mean effects model and NMA models, Figure 2. The following study was identified as being below the *y* = *x* line; Clarke 2012^7^ The study was inspected for any errors that may have occurred during data extraction or noticeable population differences, but none were found. Node-splitting was subsequently used to assess whether there was any statistically significant inconsistency within the network, none was identified and thus no further action was taken.

**Figure 2.**
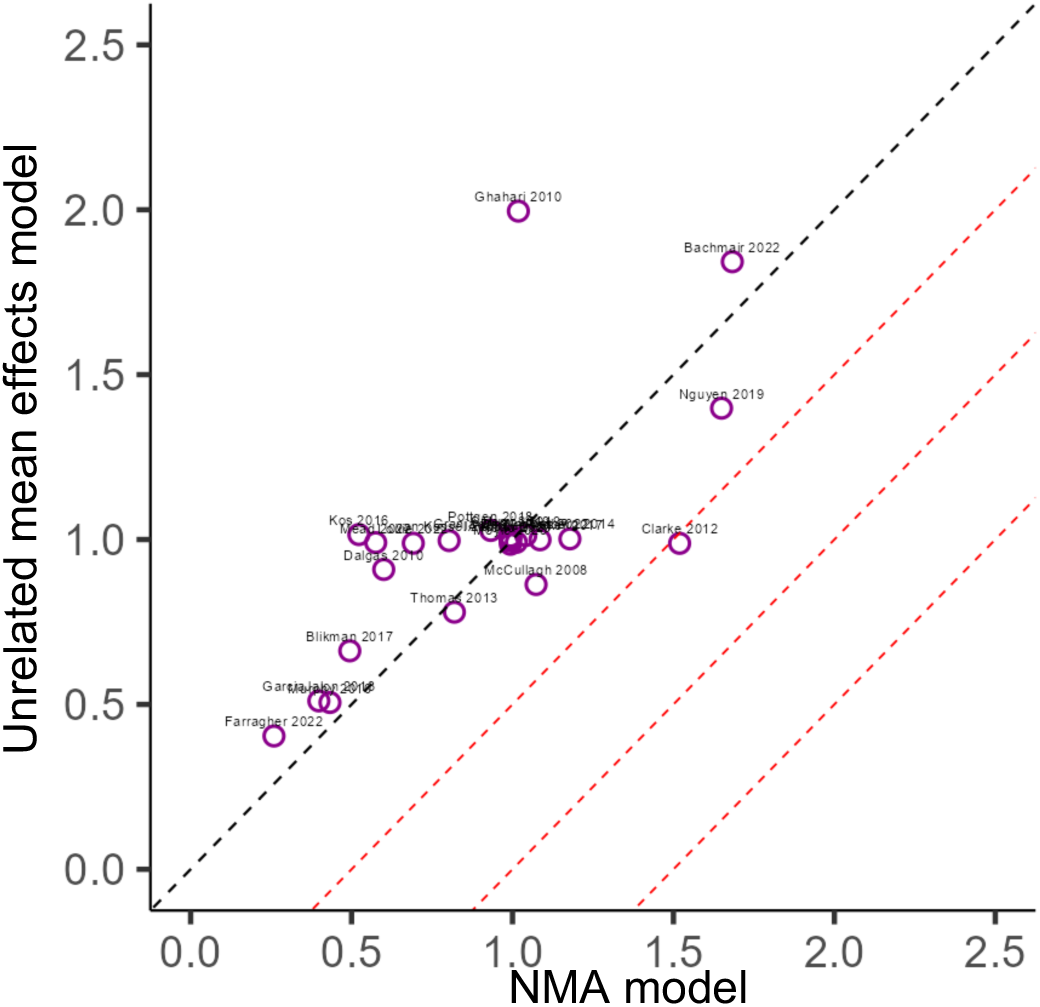
Mean posterior residual deviances according to the unrelated mean effects model versus the NMA model, at short term. Black dashed line is given by *y* = *x*, red dashed lines represent contours separated by differences of 0.5 between the two models. Any studies below the first red dashed line indicative of potential inconsistency.

##### Longer term (LT)

The mean posterior residual deviances were compared between the inconsistency and consistency models, Figure 3. No studies were identified as being below the *y* = *x* line, suggesting that there is no evidence of inconsistency within the network.

**Figure 3.**
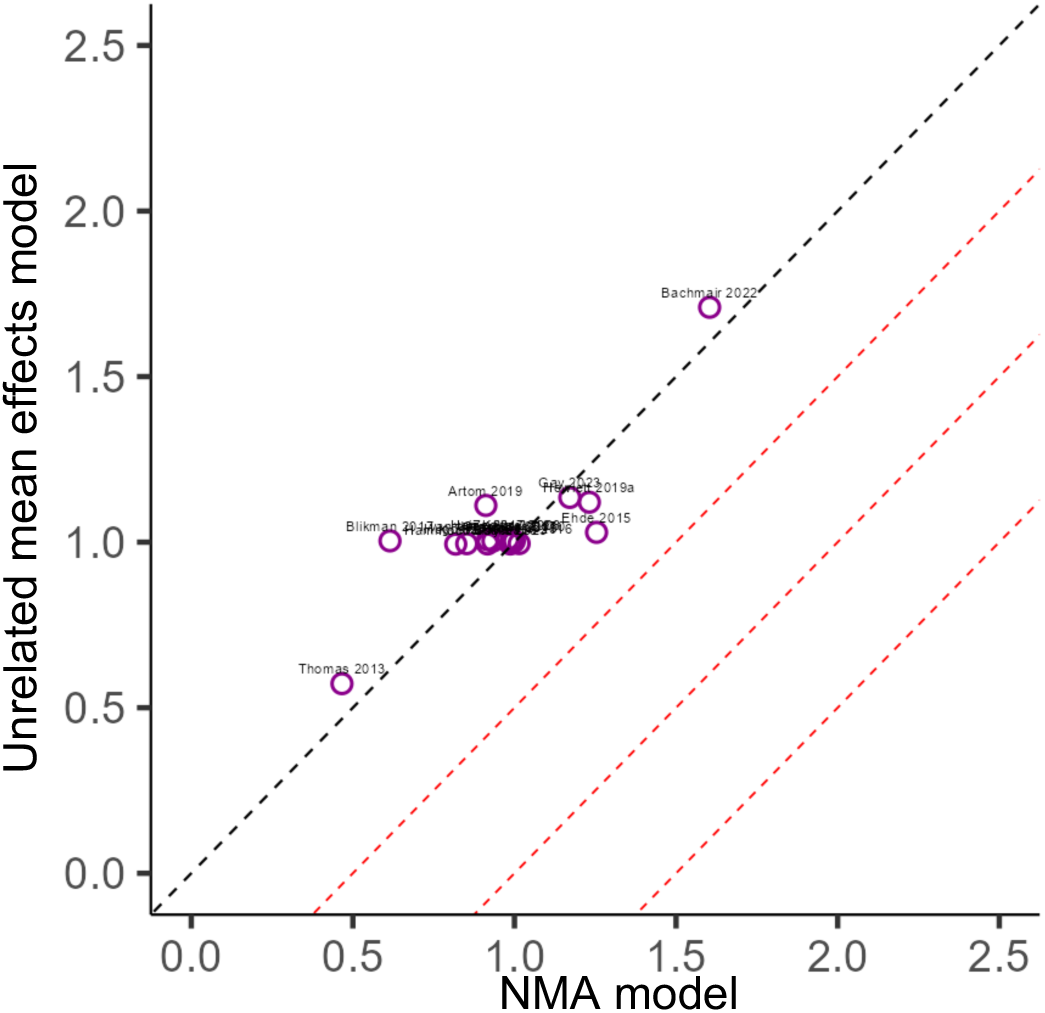
Mean posterior residual deviances according to the unrelated mean effects model and NMA model, at long term. Black dashed line is given by *y* = *x* red dashed lines represent contours separated by differences of 0.5 between the two models. Any studies below the first red dashed line indicative of potential inconsistency.

#### NMA scenario analysis: use of alternative data to inform the LT analysis

Data were available from 18 studies presenting a graded fatigue outcome at LT follow up. Five studies (Artom 2019^8^, Ehde 2015^9^, Gay 2023^10^, Hammond 2008^11^ and Hewlett 2019a^12^) presented alternative data for the LT follow-up at a time point closer to 3 months. These studies in the primary analysis had a follow up time of 10 months, 10 months, 12 months, 12 months and 46 weeks respectively. Within this scenario analysis, data collected at 4 months, 4 months, 6 months, 6 months and 20 weeks was instead used for each study in order to assess the potential impact of our decision to extract the longest available time points for the LT analysis. The network of evidence remains the same as that presented in Figure 2 of the main text.

The figure below shows the updated forest plot for the scenario analysis using alternative data for these five studies. There were no changes to which interventions were identified as statistically significant, though some minor differences were observed in the 95% credible intervals (CrIs). The between study heterogeneity was slightly increased in this analysis, compared with the LT primary analysis; 0.137 [95% CrI 0.009, 0.436] versus 0.096 [95% CrI 0.005, 0.356]. Both indicate moderate heterogeneity, but the slight increase is likely due to the inclusion of more variable follow-up times within this scenario.

**Figure 4.**
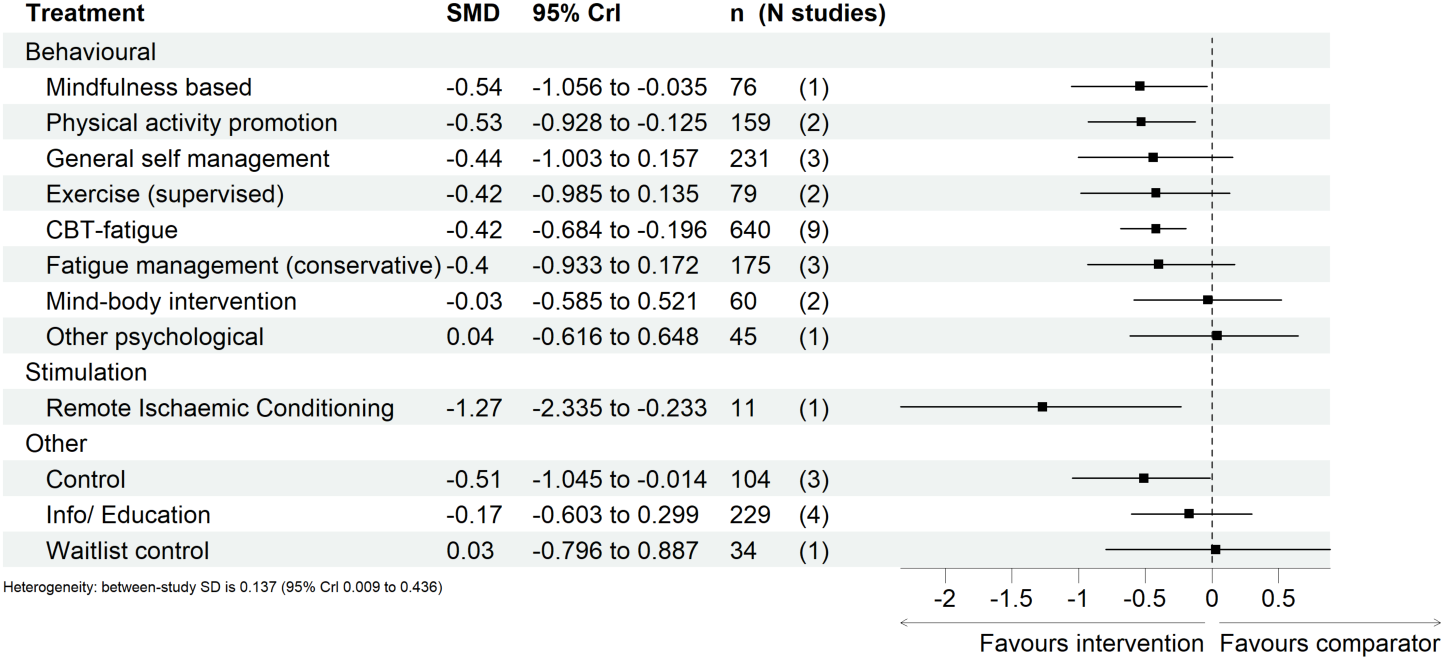
Predicted effects on fatigue outcomes of interventions, relative to usual care, at long term^†^, with 95% credible intervals (CrI). The number of participants (n) and the number of studies (N studies) are given for context. Broad intervention categorisation is also presented to aid interpretation (Behavioural, Stimulation, Nutritional, and Other). The “control” node is displayed as this functioned to ensure connectivity of the network, but this is not an active intervention for consideration/recommendation. ^†^Data for five studies changed to use earlier follow-up data within the long-term analysis time window.

#### NMA scenario analysis: relaxation of the transdiagnostic assumption

Condition group specific networks were constructed to help assess any potential differences in treatment effects across different condition groups. Due to the sparsity of evidence, networks could only be constructed using: EOT data for multiple sclerosis (MS), musculoskeletal conditions (MSK), inflammatory bowel disease (IBD), Kidney-related and Stroke-related conditions; ST for MS; and LT for MS and MSK. The results for each of these networks are presented below.

##### EOT

For the EOT condition-specific networks, Figure 5, there were 6 viable networks relating to the following condition groups: MS, MSK (two disconnected networks), IBD, Kidney, and Stroke. The largest EOT network was for MS with 19 interventions across 44 studies; the network originally included 46 studies, but statistically significant inconsistency was detected via node-splitting, which led to the removal of two studies, Fleming (2021)^2^ and Turner (2016)^5^ which provided direct evidence for the interventions flagged with statistically significant inconsistency. Two disconnected EOT networks were constructed for MSK: the first, “MSK #1”, included 10 interventions over 12 studies; the second, “MSK #2”, included 3 interventions over 3 studies. The EOT network for IBD included 8 interventions over 6 studies. Whilst the EOT networks for Kidney and Stroke each contained 4 interventions across 3 studies. Note that no inconsistency checking via node-splitting was not feasible for the following networks: MSK #2, Kidney, and Stroke, because the networks contained no closed loops of evidence.

The point estimates and 95% CrIs for the EOT condition-specific networks are shown in Figures 6-11; the treatment effect is relative to usual care unless otherwise stated. Several differences can be seen between the primary analysis where the transdiagnostic assumption is upheld and the condition group specific networks – these are detailed below.

**Figure 5.**
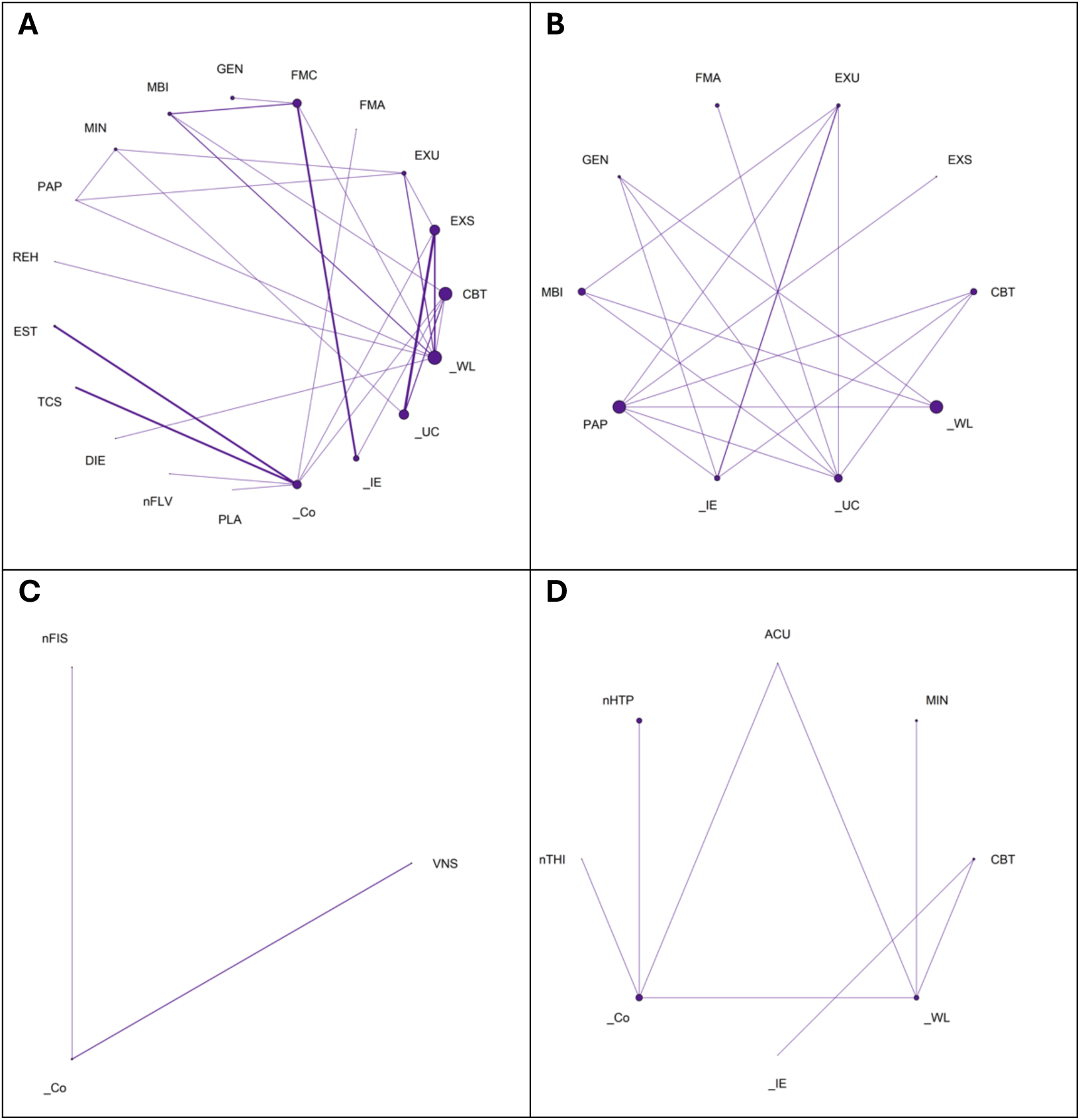
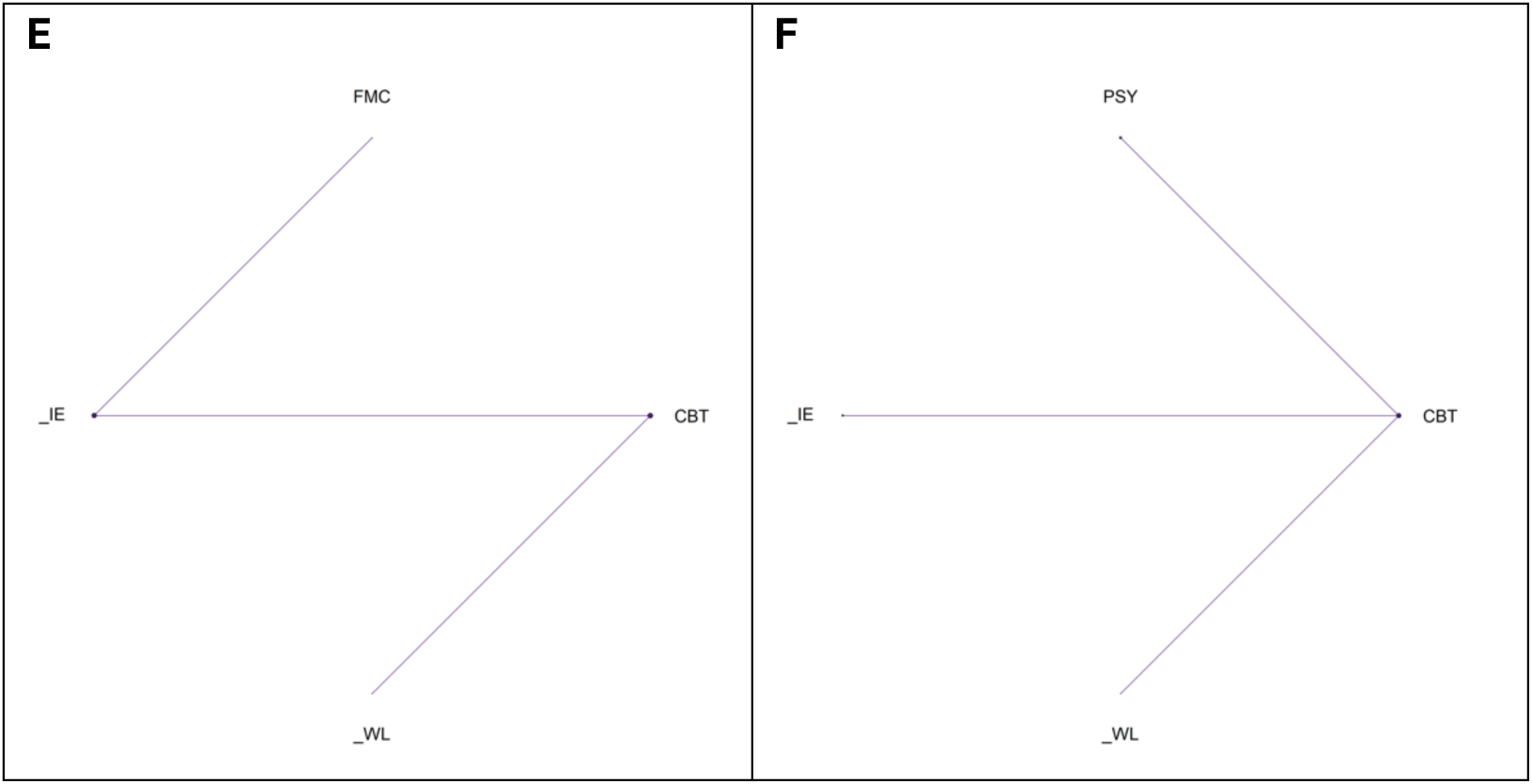
Network geometry for A) MS, B) MSK #1, C) MSK #2, D) IBD, E) Kidney, and F) Stroke condition-specific analyses, at end of treatment, respectively, indicating the number of participants who received each intervention (size of node) and the number of studies contributing to the direct evidence and comparisons between interventions (thickness of line).

In the EOT MS-specific network, all behavioural interventions (except physical activity promotion) were found to have beneficial, statistically significant effects on fatigue outcomes. Transcranial and external stimulation were also shown to have beneficial predicted effects.

Of the nutritional interventions, only Flavenoid (cocoa) supplements were shown to have a potentially beneficial, statistically significant effect on fatigue outcomes, but as in the primary analysis, this was only evidenced by one study and should be interpreted with caution.

A number of treatments were found to have statistically significant effects which were not identified in the primary analysis, these included: non-specific rehabilitation, fatigue management (conservative), general self management, mind-body intervention and Flavenoid (Cocoa), suggesting these interventions may have improved effects on fatigue for individuals with MS. Conversely, physical activity promotion is not shown to have a statistically significant beneficial effect on fatigue outcomes. Furthermore, the treatment effect of waitlist control relative to usual care was non-beneficial for fatigue outcomes within the primary analysis but within the MS-specific network was shown to have a statistically significant, beneficial effect. Generally, treatment effects observed in the MS-specific network were indicative of greater treatment effects on fatigue outcomes when compared to the primary analysis, however, the evidence base of the MS-specific network is approximately half of that analysed within the primary analysis (44 vs. 84 studies) and results should therefore be interpreted with appropriate caution.

The between study heterogeneity was found to be 0.12 [95% CrI 0.007, 0.312] which indicates moderate heterogeneity within the network.

**Figure 6.**
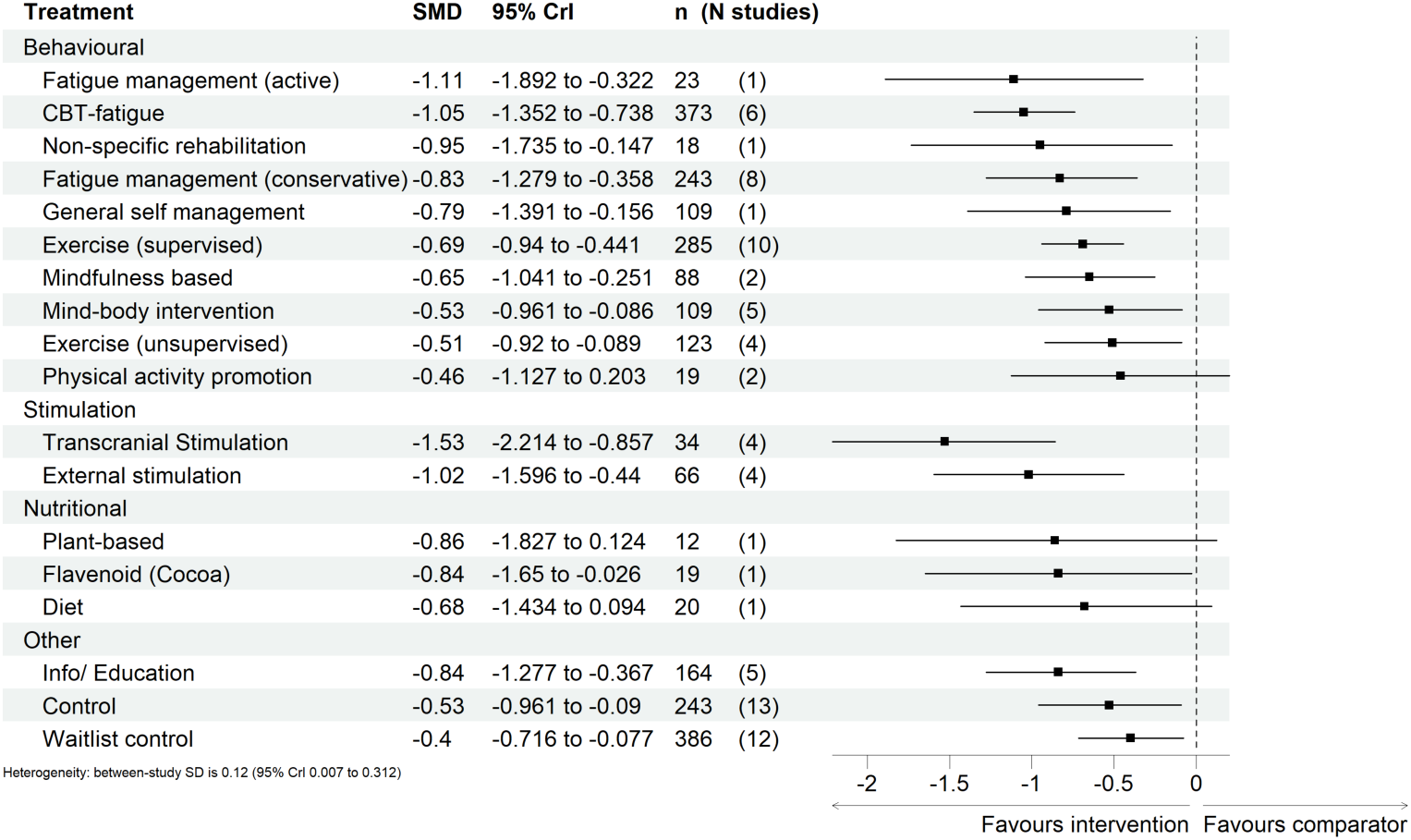
Predicted effects on fatigue outcomes of interventions within the MS-specific network, relative to usual care, at end of treatment, with 95% credible intervals (CrI). The number of participants (n) and the number of studies (N studies) are given for context. Broad intervention categorisation is also presented to aid interpretation (Behavioural, Stimulation, Nutritional, and Other). The “control” node is displayed as this functioned to ensure connectivity of the network, but this is not an active intervention for consideration/recommendation.

In the EOT MSK #1 network, exercise (supervised) and fatigue management (active) had statistically significant, beneficial effects on fatigue outcomes, as in the primary analysis. However, although CBT-fatigue and physical activity promotion were found to be statistically significantly beneficial in the primary analysis, in the MSK #1 analysis, they were no longer found to be statistically significant. The evidence base of the MSK #1 network is however much smaller than the primary analysis network (12 vs 84 studies) and therefore the treatment effects presented should be interpreted with caution as no intervention featured across more than 4 MSK studies.

The between study heterogeneity was 0.138 [95% CrI 0.007, 0.498], which indicates a moderate study heterogeneity within the network.

**Figure 7.**
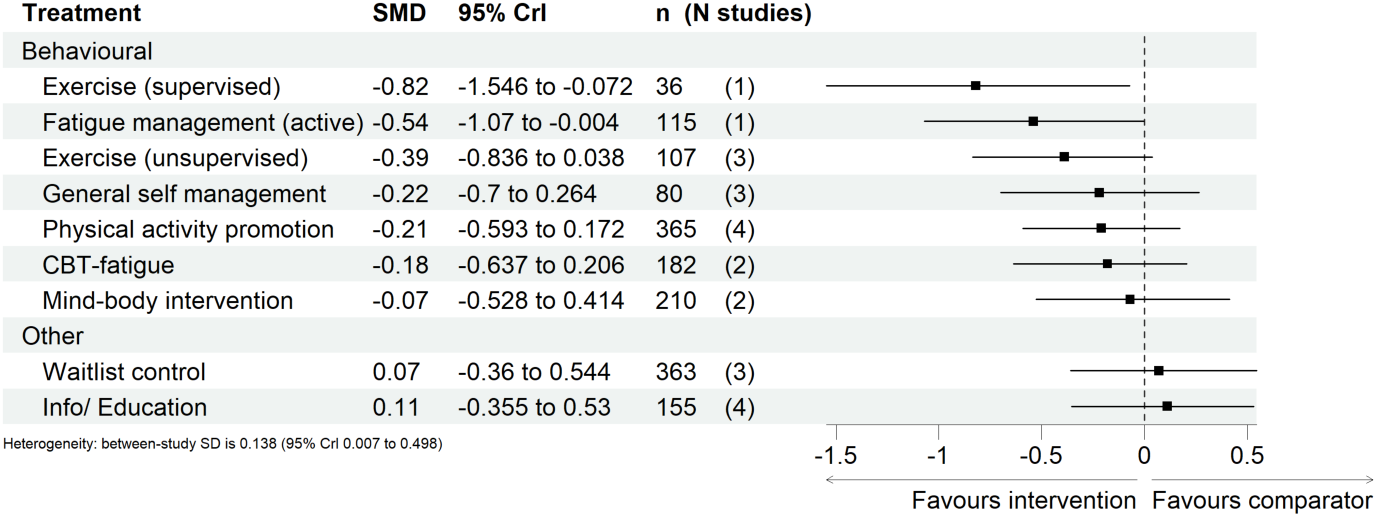
Predicted effects on fatigue outcomes of interventions within the MSK-specific network (#1), relative to usual care, at end of treatment, with 95% credible intervals (CrI). The number of participants (n) and the number of studies (N studies) are given for context. Broad intervention categorisation is also presented to aid interpretation (Behavioural, Stimulation, Nutritional, and Other).

The EOT MSK #2 network treatment effects are presented relative to control and only included comparison of vagal stimulation and fish oil, Figure 8. As there were fewer than 5 studies within the 2^nd^ network for MSK-related conditions, an informative prior on the between study heterogeneity was used. Vagal stimulation was found to be statistically significant with a positive effect on fatigue outcomes, however, this network only consisted of 3 studies, two of which influenced the vagal stimulation node, results should therefore be interpreted with caution. The between study heterogeneity was predicted to be 0.14 [95%CrI 0.027, 0.477] indicating moderate heterogeneity.

**Figure 8.**
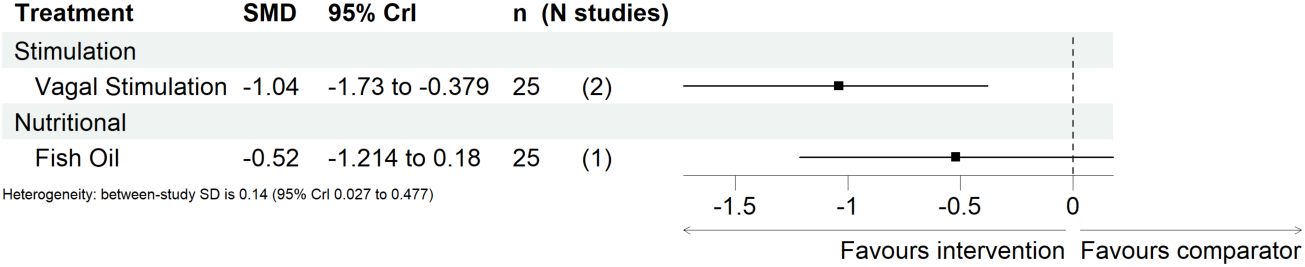
Predicted effects on fatigue outcomes of interventions within the MSK-specific network (#2), relative to control, at end of treatment, with 95% credible intervals (CrI). The number of participants (n) and the number of studies (N studies) are given for context. Broad intervention categorisation is also presented to aid interpretation (Behavioural, Stimulation, Nutritional, and Other).

The EOT analysis for IBD conditions showed that none of the interventions were identified to have statistically significant effects relative to waitlist control. However, these interventions were informed by a maximum of 2 studies and thus have minimal evidence. In addition to the low number of studies, the between study heterogeneity standard deviation was found to be 1.385 [95% CrI 0.071, 2.694], which indicates extremely high heterogeneity amongst the 6 studies included within the network, and therefore these results should be interpreted accordingly.

**Figure 9.**
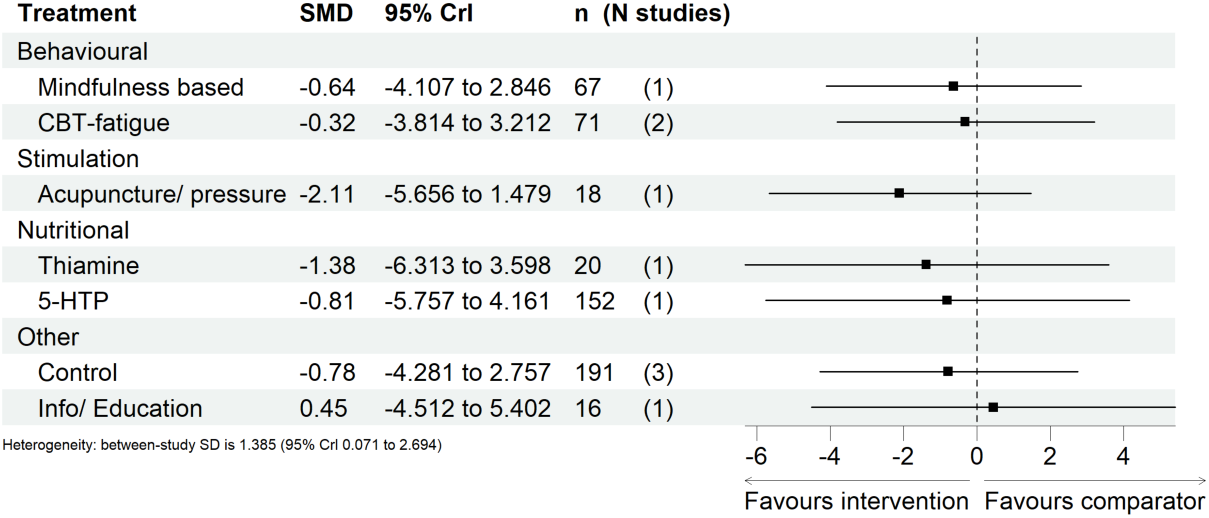
Predicted effects on fatigue outcomes of interventions within the IBD-specific network, relative to wait list, at end of treatment, with 95% credible intervals (CrI). The number of participants (n) and the number of studies (N studies) are given for context. Broad intervention categorisation is also presented to aid interpretation (Behavioural, Stimulation, Nutritional, and Other). The “control” node is displayed as this functioned to ensure connectivity of the network, but this is not an active intervention for consideration/recommendation.

As there were fewer than 5 studies within the network for Kidney-related conditions, an informative prior on the between study heterogeneity was used. The analysis showed that neither CBT-fatigue nor fatigue management (conservative) were found to be statistically significantly beneficial for fatigue outcomes relative to wait list controls. This network was informed by only four studies and is likely underinformed. The between study heterogeneity was found to be moderate (0.143 [95%CrI 0.027, 0.483]).

**Figure 10.**
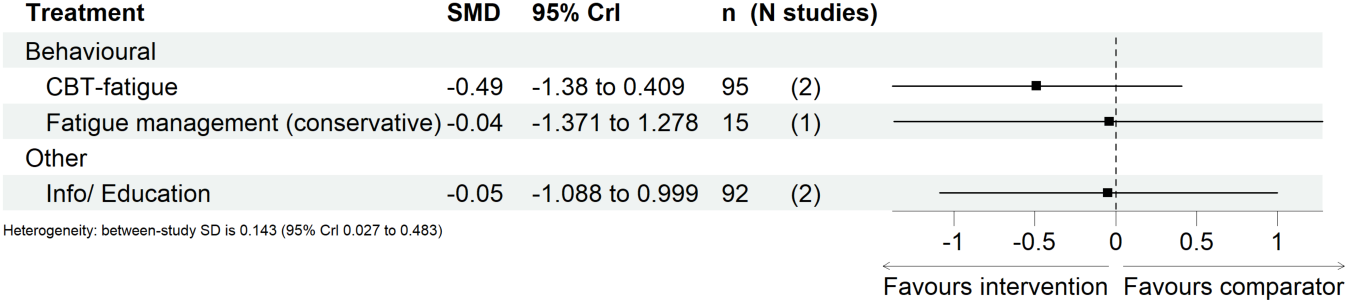
Predicted effects on fatigue outcomes of interventions within the Kidney-specific network, relative to wait list, at end of treatment, with 95% credible intervals (CrI). The number of participants (n) and the number of studies (N studies) are given for context. Broad intervention categorisation is also presented to aid interpretation (Behavioural, Stimulation, Nutritional, and Other).

Finally, the analysis of Stroke-related conditions had again less than 5 studies informing the network and thus the informative prior was used. Three studies provided evidence for CBT- fatigue, for which a statistically significant beneficial effect was identified relative to wait list control. A statistically significant, beneficial effect was also identified for “other psychological” interventions. Due to the low numbers of studies, these results should be interpreted with caution. The between study heterogeneity was again found to be moderate, with the standard deviation equal to 0.143 [95% CrI 0.027, 0.484].

**Figure 11.**
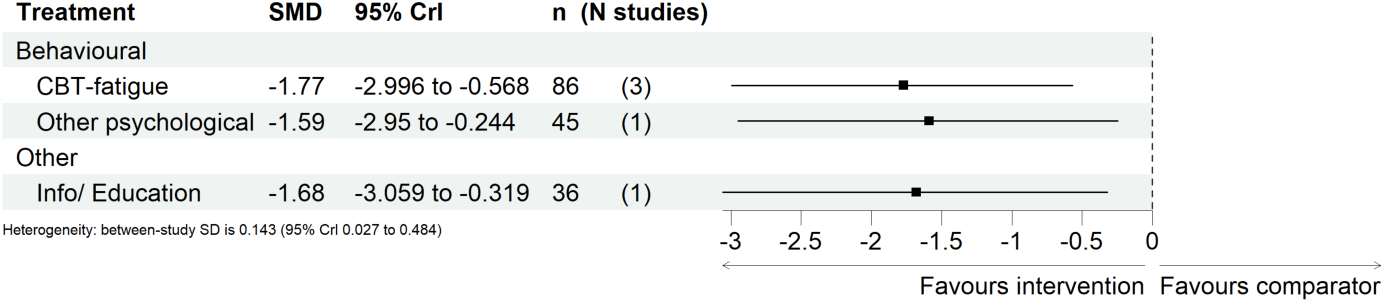
Predicted effects on fatigue outcomes of interventions within the Stroke-specific network, relative to wait list, at end of treatment, with 95% credible intervals (CrI). The number of participants (n) and the number of studies (N studies) are given for context. Broad intervention categorisation is also presented to aid interpretation (Behavioural, Stimulation, Nutritional, and Other).

Inconsistency was assessed for the EOT condition-specific networks with closed loops using the posterior mean residual deviances, followed by node-splitting, Figure 12. Following removal of Fleming (2021)^2^ and Turner (2016)^5^ from the EOT MS-specific network, no statistically significant inconsistency was detected in the EOT analyses.

**Figure 12.**
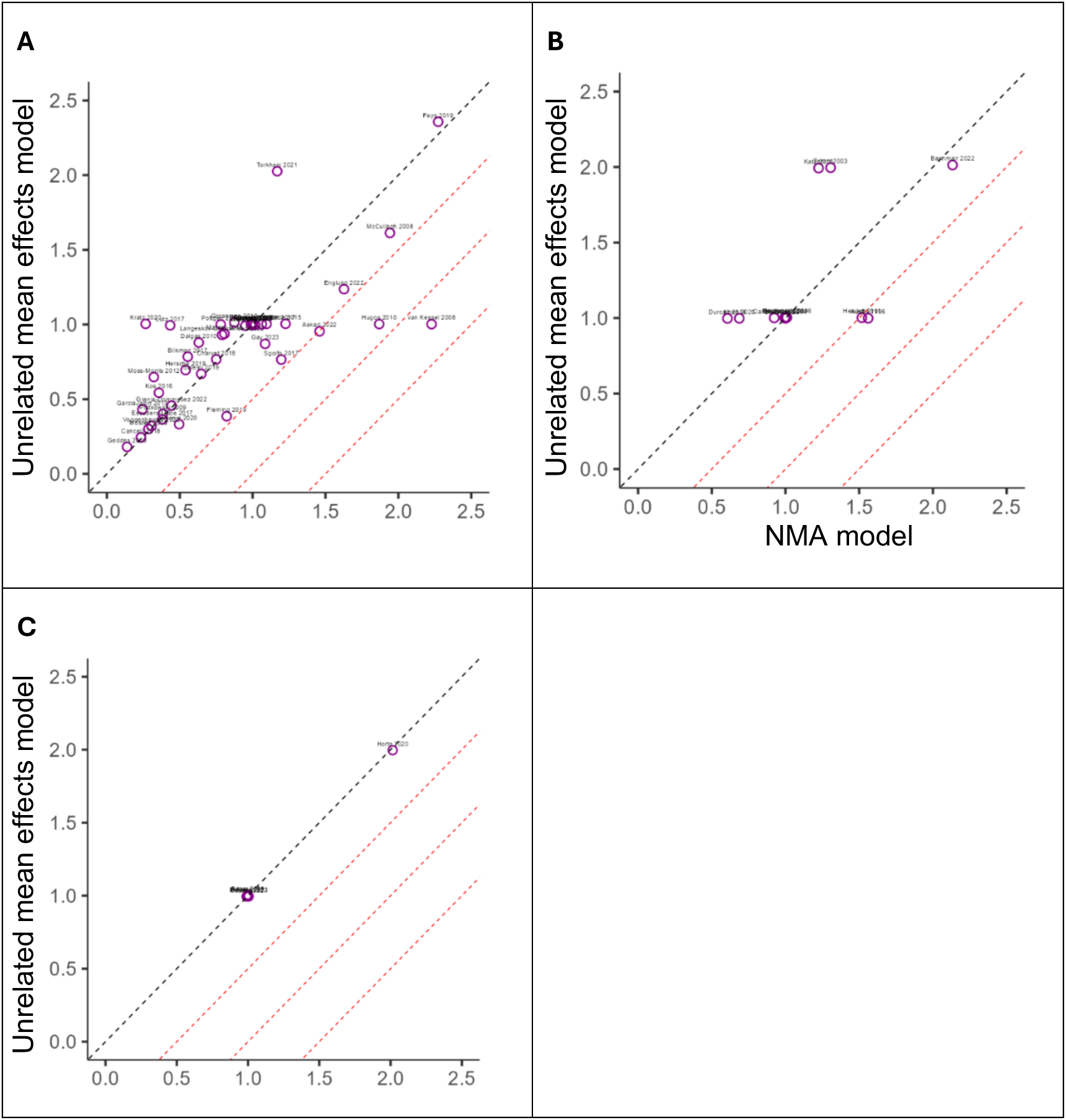
Mean posterior residual deviances according to the unrelated mean effects model versus the NMA model for A) MS, B) MSK #1, and C) IBD condition-specific analyses, at end of treatment, respectively. Black dashed line is given by *y* = *x*, red dashed lines represent contours separated by differences of 0.5 between the two models. Any studies below the first red dashed line indicative of potential inconsistency.

##### ST

For the ST condition-specific analysis, only one viable network could be constructed which summarised evidence for studies in MS, shown in Figure 13 A. This network included 12 interventions across 14 studies. The point estimates and 95% CrIs for the ST MS network are shown in Figure 13 B where the treatment effect is relative to usual care. Inconsistency was assessed for the ST MS network via comparison of the posterior mean residual deviances from the unrelated mean effects model and NMA model, followed by node-splitting, Figure 13 C; no statistically significant inconsistency was detected.

In the ST MS-specific network, no treatment was identified to have a statistically significant effect on fatigue outcomes, however, none of the included treatments were found to be statistically significant in the ST primary analysis. Additionally, the evidence base of the MS-specific network is smaller than the evidence within the primary analysis (14 vs. 24 studies).

In general, the treatment effects should be interpreted with caution as no intervention featured in more than 4 studies.

The between study heterogeneity standard deviation was 0.243 [95% CrI 0.01, 1.528] which indicates moderate heterogeneity within the network.

**Figure 13.**
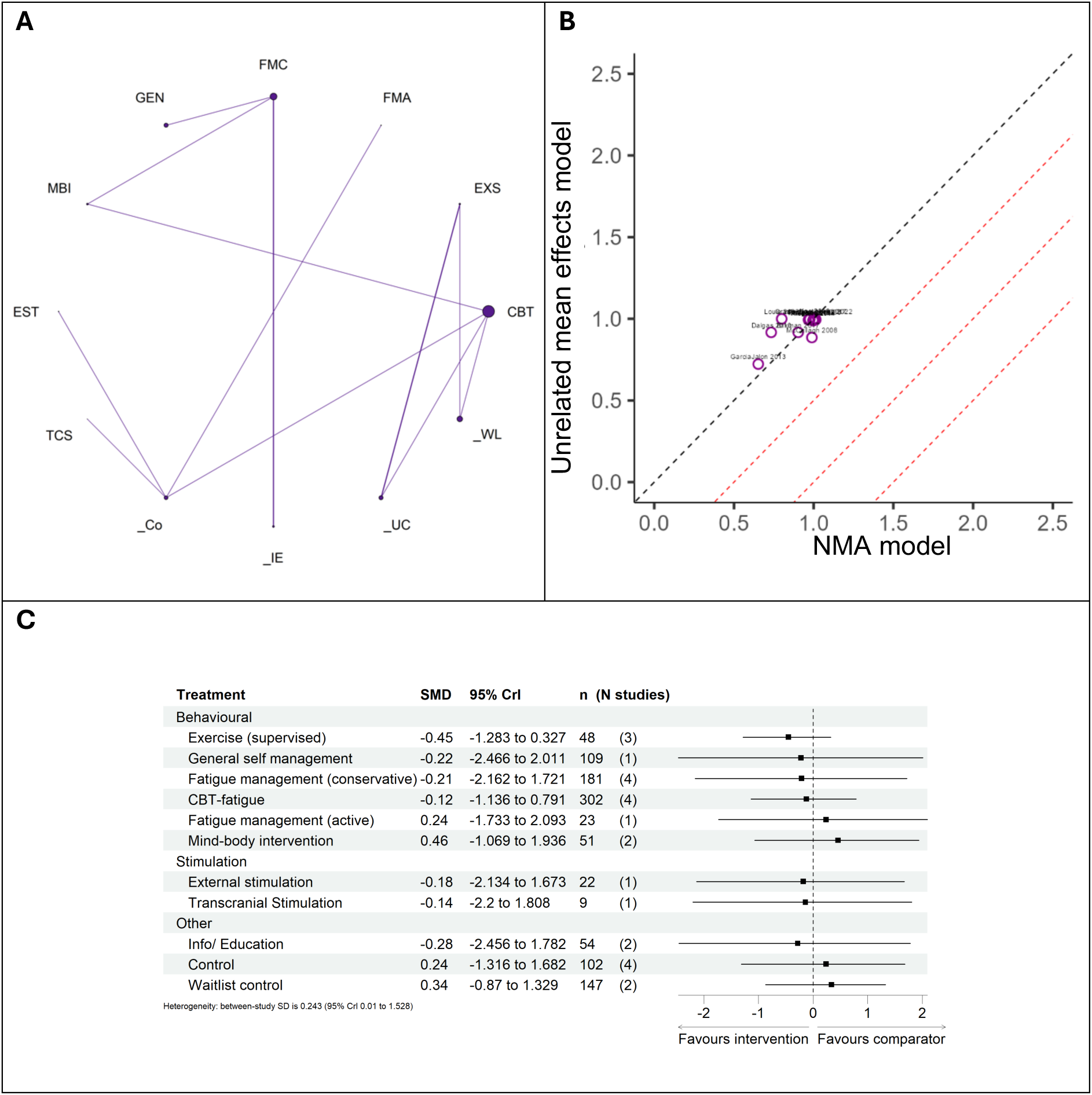
A) Network geometry, B) posterior residual deviances according to the unrelated mean effects model and the NMA model, and C) predicted effects on fatigue outcomes of interventions, relative to usual care, with 95% credible intervals (CrI); for the MS condition-specific analysis, at short term, respectively. The number of participants (n) and the number of studies (N studies) are given for context. The “control” node is displayed as this functioned to ensure connectivity of the network, but this is not an active intervention for consideration/recommendation.

##### LT

For the LT condition-specific analysis, there were two viable networks relating to: MS, shown in Figure 14 A, and MSK, shown in Figure 15 A. These networks included: 9 interventions across 10 studies, and 4 interventions across 3 studies, respectively. The point estimates and 95% CrIs for the LT networks are shown in Figure 14 B and 15 B, respectively; the treatment effects are relative to usual care. Inconsistency was assessed for the LT networks by comparing the posterior mean residual deviances from the unrelated mean effects model and the NMA model, followed by node-splitting, Figure 14 C and 15 C; no statistically significant inconsistency was detected in either network.

In the LT MS-specific network, no treatment was shown to have a statistically significant effect on fatigue outcomes. In the LT primary analysis, mindfulness and CBT-fatigue were shown to have statistically significant, beneficial effects on fatigue outcomes, but this was not mirrored in the MS-specific analysis. As for the MS-specific EOT and ST networks, the evidence base is approximately half of the transdiagnostic case (10 vs. 18 studies); though there is some consensus between treatment effects seen in the MS-specific network and the primary analysis, however the broadening of the 95% CrIs resulted in non-significance.

The between study heterogeneity standard deviation was 0.317 [95% CrI 0.015, 2.119], indicating moderate to high heterogeneity within the network.

**Figure 14.**
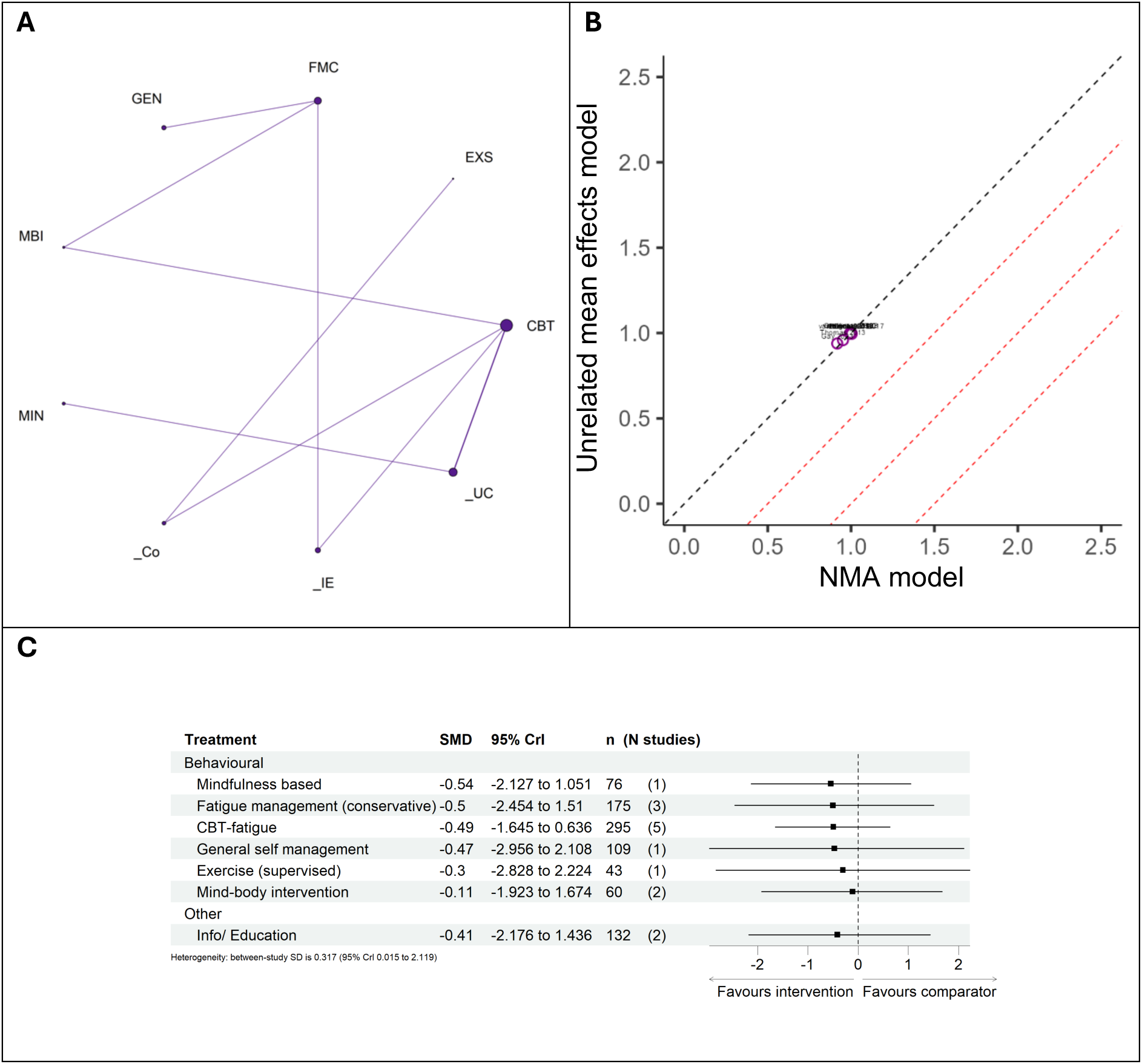
A) Network geometry, B) posterior residual deviances according to the unrelated model effects model and the NMA model, and C) predicted effects on fatigue outcomes of interventions, relative to usual care, with 95% credible intervals (CrI); for the MS condition-specific analysis, at long term, respectively. The number of participants (n) and the number of studies (N studies) are given for context. The “control” node is displayed as this functioned to ensure connectivity of the network, but this is not an active intervention for consideration/recommendation.

In the LT MSK-specific network, two treatments were shown to have statistically significant treatment effects relative to usual care: physical activity promotion and CBT-fatigue.

Exercise (supervised) was found not to be statistically significant – these results are consistent with the LT primary analysis.

The between study heterogeneity standard deviation was moderate, 0.121 [95% CrI 0.025, 0.435].

**Figure 15.**
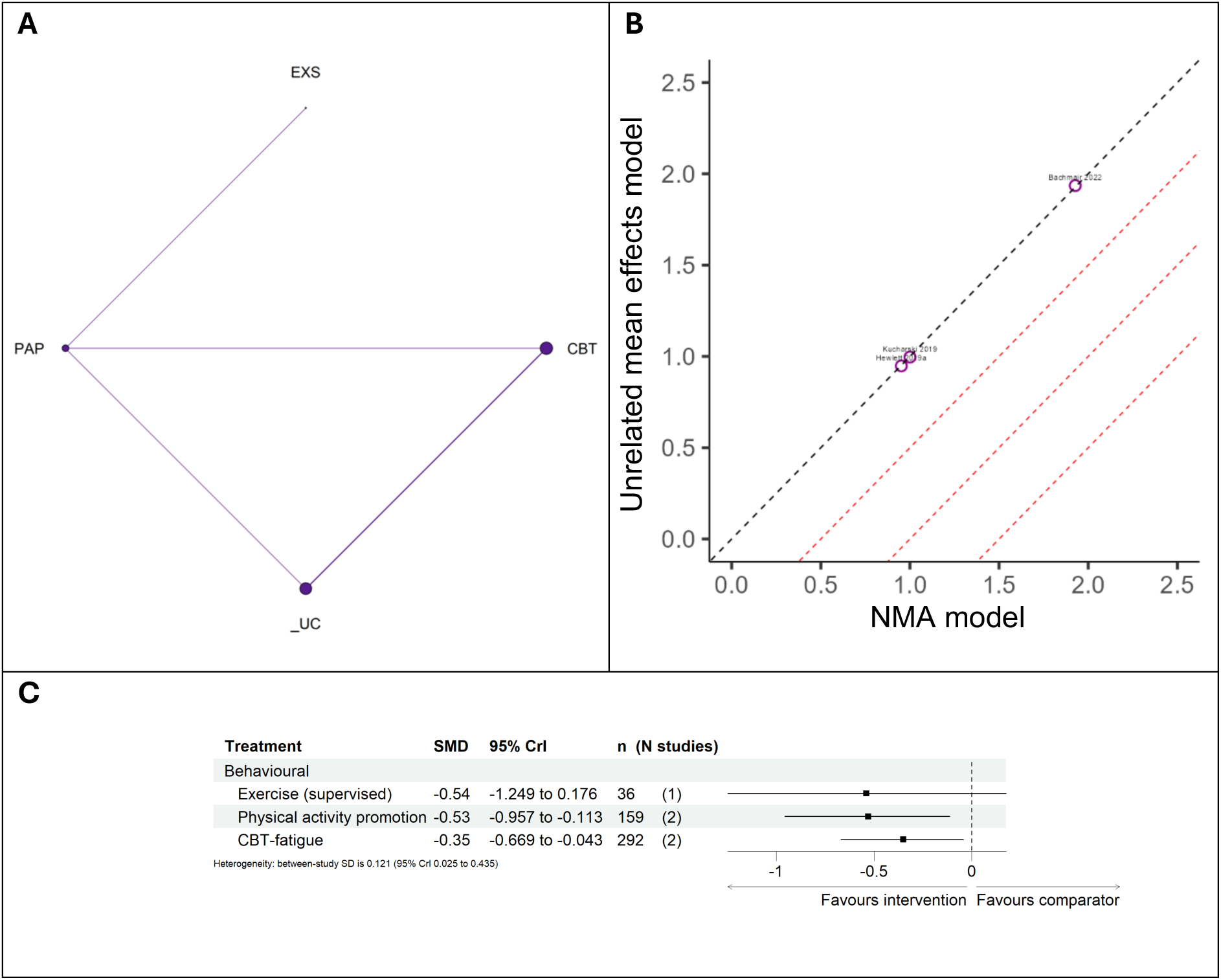
A) Network geometry, B) posterior residual deviances according to the unrelated mean effects model and the NMA model, and C) predicted effects on fatigue outcomes of interventions, relative to usual care, with 95% credible intervals (CrI); for the MSK condition-specific analysis, at long term, respectively. The number of participants (n) and the number of studies (N studies) are given for context. The “control” node is displayed as this functioned to ensure connectivity of the network, but this is not an active intervention for consideration/recommendation.

#### NMA scenario analysis: exclusion of pilot and feasibility studies

The evidence base for the primary analysis consists of studies reporting results from RCTs as well as pilot and feasibility trials. To assess the potential impact of the inclusion of pilot and feasibility studies within the NMAs, we re-constructed the networks for EOT, ST and LT follow up, omitting any pilot or feasibility studies. This resulted in networks with 23, 13 and 12 connected interventions, informed by 65, 15 and 15 studies.

##### EOT

Generally, the NMA results when excluding pilot/feasibility studies were similar to the primary analysis. Four interventions were no longer included in the network; remote ischaemic conditioning, fish oil supplements, plant based supplements, and flavonoid (cocoa) derived supplements. In the primary analysis, acupuncture was shown to exhibit statistically significant beneficial effects for fatigue outcomes, this was however only directly informed by two studies. In this scenario analysis, one of these studies was excluded and the treatment effect of acupuncture was no longer shown to be statistically significant. Similarly, thiamine-based supplements in this scenario analysis were directly informed by two fewer studies than the primary analysis and no longer found to be statistically significant. The between study variance was comparable between this analysis and the primary analysis. Other than the changes listed above, there was generally a minor impact on treatment effect estimates and the associated 95% CrIs at EOT.

**Figure 16.**
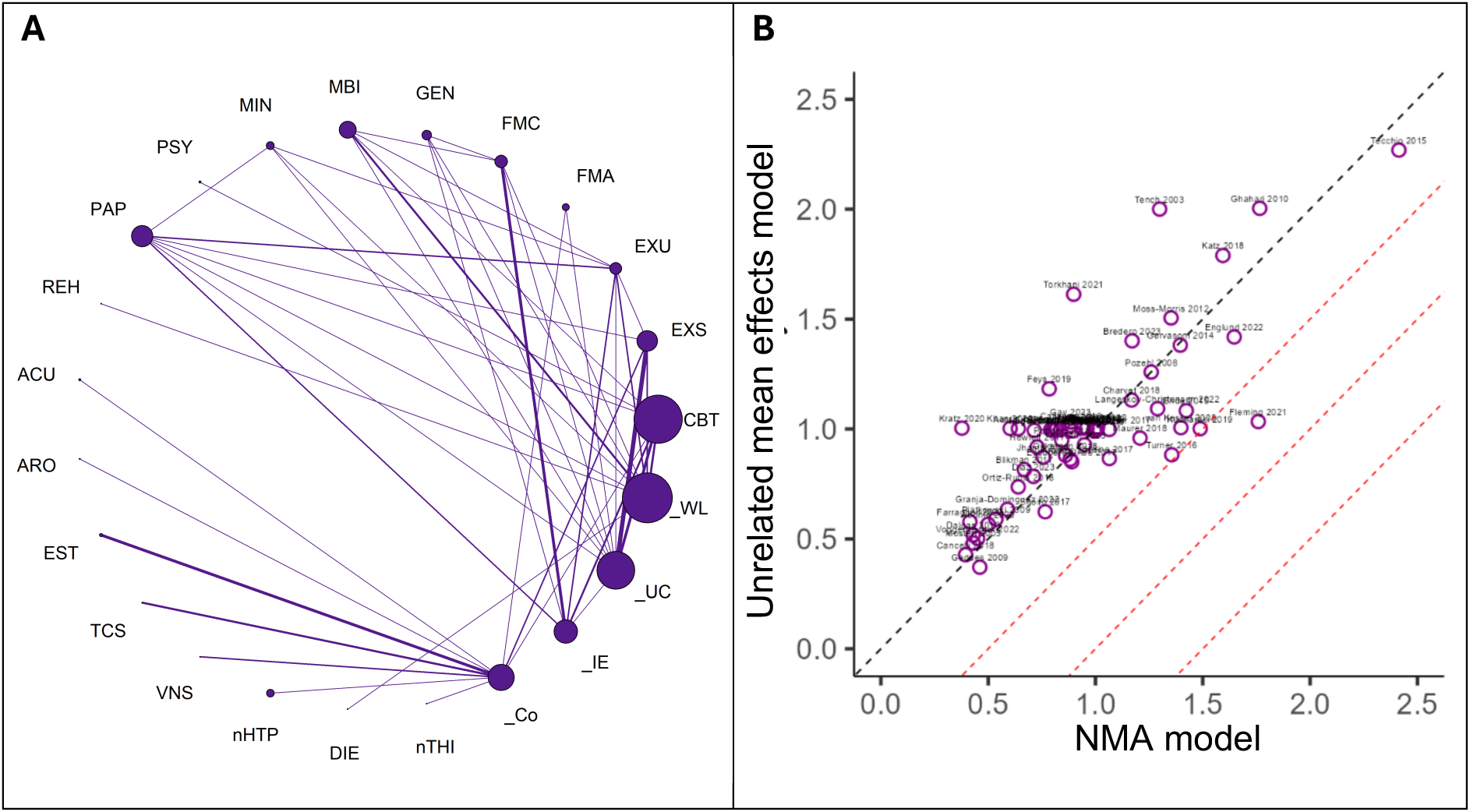
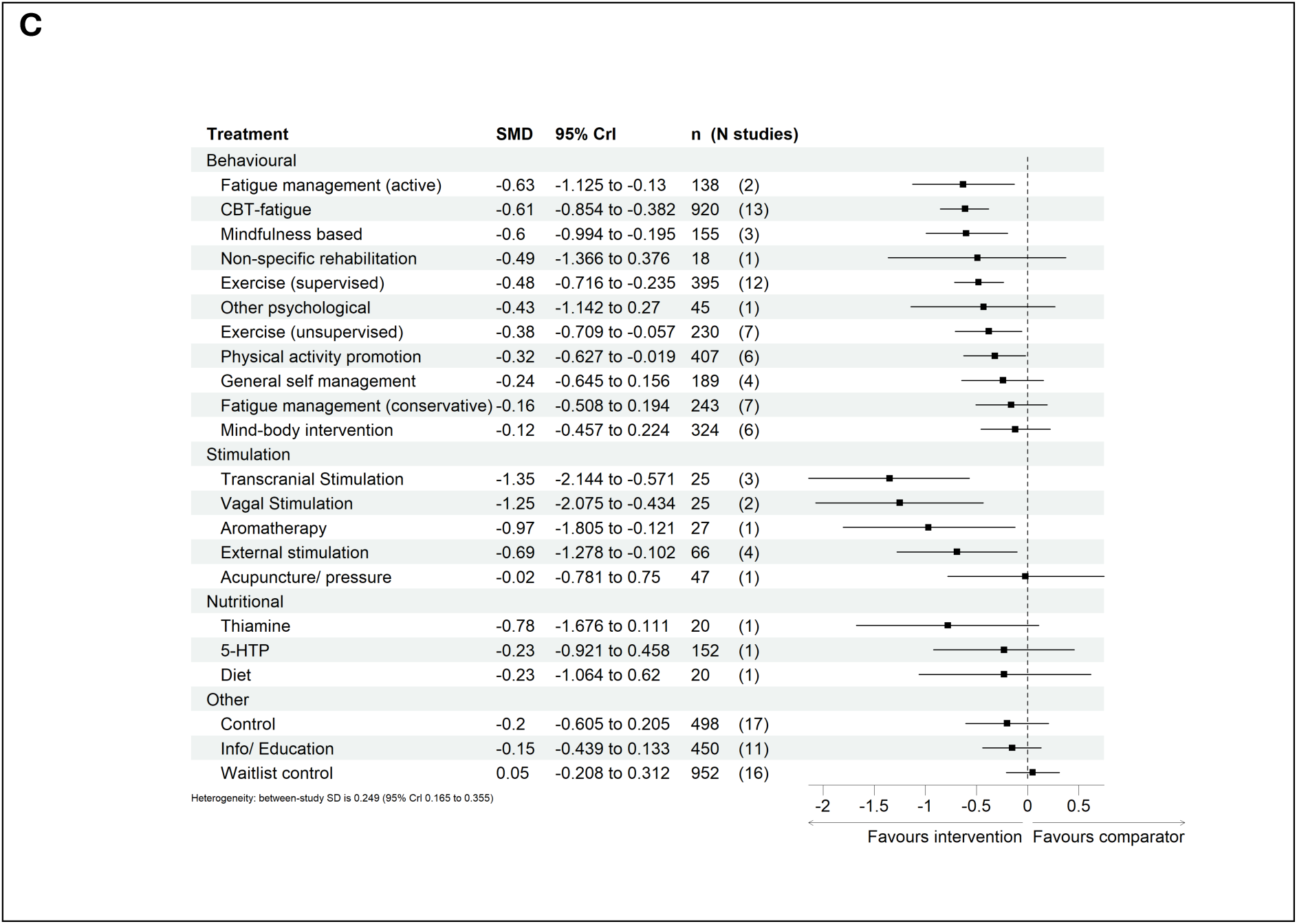
A) Network geometry, B) posterior residual deviances according to the unrelated mean effects model and NMA model, and C) predicted effects on fatigue outcomes of interventions, relative to usual care, with 95% credible intervals (CrI); for the end of treatment analysis†, respectively. The number of participants (n) and the number of studies (N studies) are given for context. The “control” node is displayed as this functioned to ensure connectivity of the network, but this is not an active intervention for consideration/recommendation. †Data from pilot/feasibility studies were excluded in this analysis.

##### ST

The SMDs and 95% credible intervals were similar at ST follow-up when pilot and feasibility studies were excluded with the primary analysis, with no changes in statistical significance for the included interventions. Three interventions were however no longer included in the network including: remote ischaemic conditioning, transcranial stimulation, and acupuncture/pressure based interventions. Between study variance was comparable between the two analyses.

**Figure 17.**
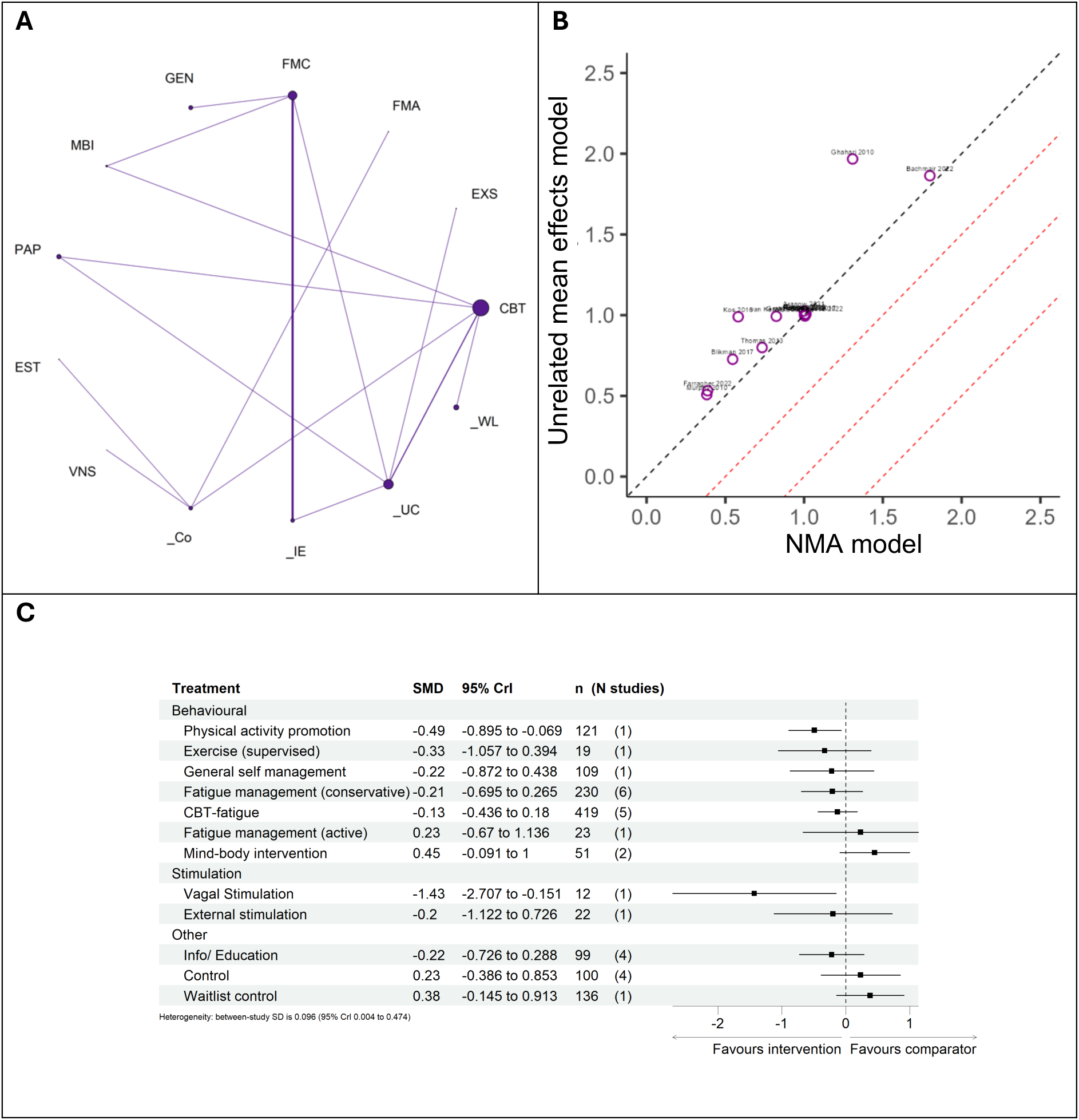
A) Network geometry, B) posterior residual deviances according to the unrelated mean effects model versus the NMA model, and C) predicted effects on fatigue outcomes of interventions, relative to usual care, with 95% credible intervals (CrI); for the short term analysis^†^, respectively. The number of participants (n) and the number of studies (N studies) are given for context. The “control” node is displayed as this functioned to ensure connectivity of the network, but this is not an active intervention for consideration/recommendation. ^†^Data from pilot/feasibility studies were excluded in this analysis.

##### LT

In the LT follow-up analysis, when pilot studies were not included, broader 95% CrIs were evident for the majority of interventions compared to the primary analysis. Despite this, two interventions were found to exhibit statistically significant, beneficial effects for fatigue, which were not found to be statistically significant in the primary analysis, including: conservative fatigue management approaches, and general self management. Only one study directly evidencing conservative fatigue management was a pilot study in the primary analysis, and thus the changes in the scenario analysis results appear to be an indirect effect resulting from changes to other interventions within the network. Despite this, other intervention treatment effect point estimates, appear to be similar to the primary analysis. As in the EOT and ST follow-up analyses, remote ischaemic conditioning was no longer included within the network. As with the other time points, between study variance was comparable to that observed in the primary analyses.

**Figure 18.**
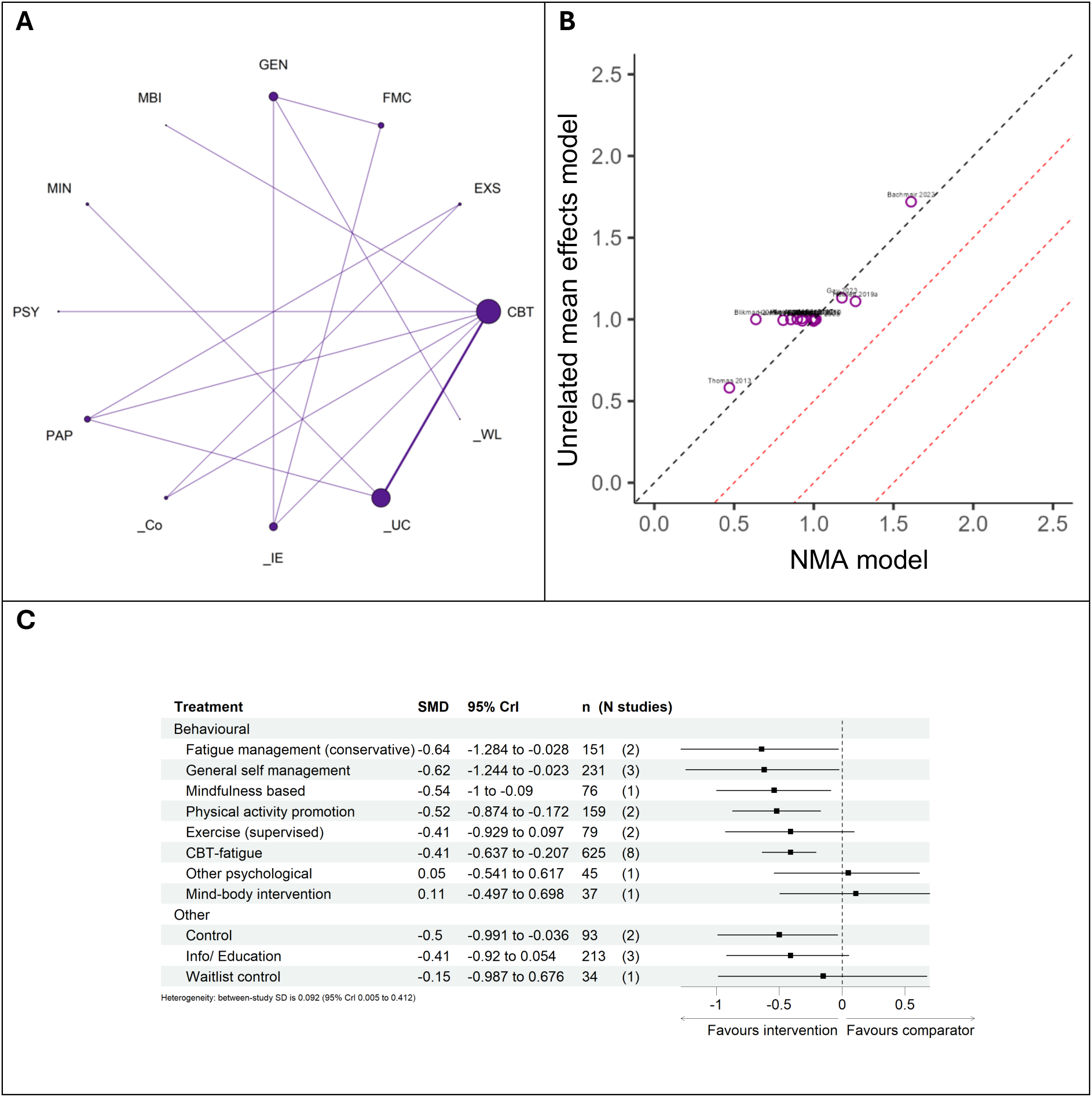
A) Network geometry, B) posterior residual deviances according to the unrelated mean effects model and the NMA model, and C) predicted effects on fatigue outcomes of interventions, relative to usual care, with 95% credible intervals (CrI); for the long term analysis^†^, respectively. The number of participants (n) and the number of studies (N studies) are given for context. The “control” node is displayed as this functioned to ensure connectivity of the network, but this is not an active intervention for consideration/recommendation. ^†^Data from pilot/feasibility studies were excluded in this analysis.

---

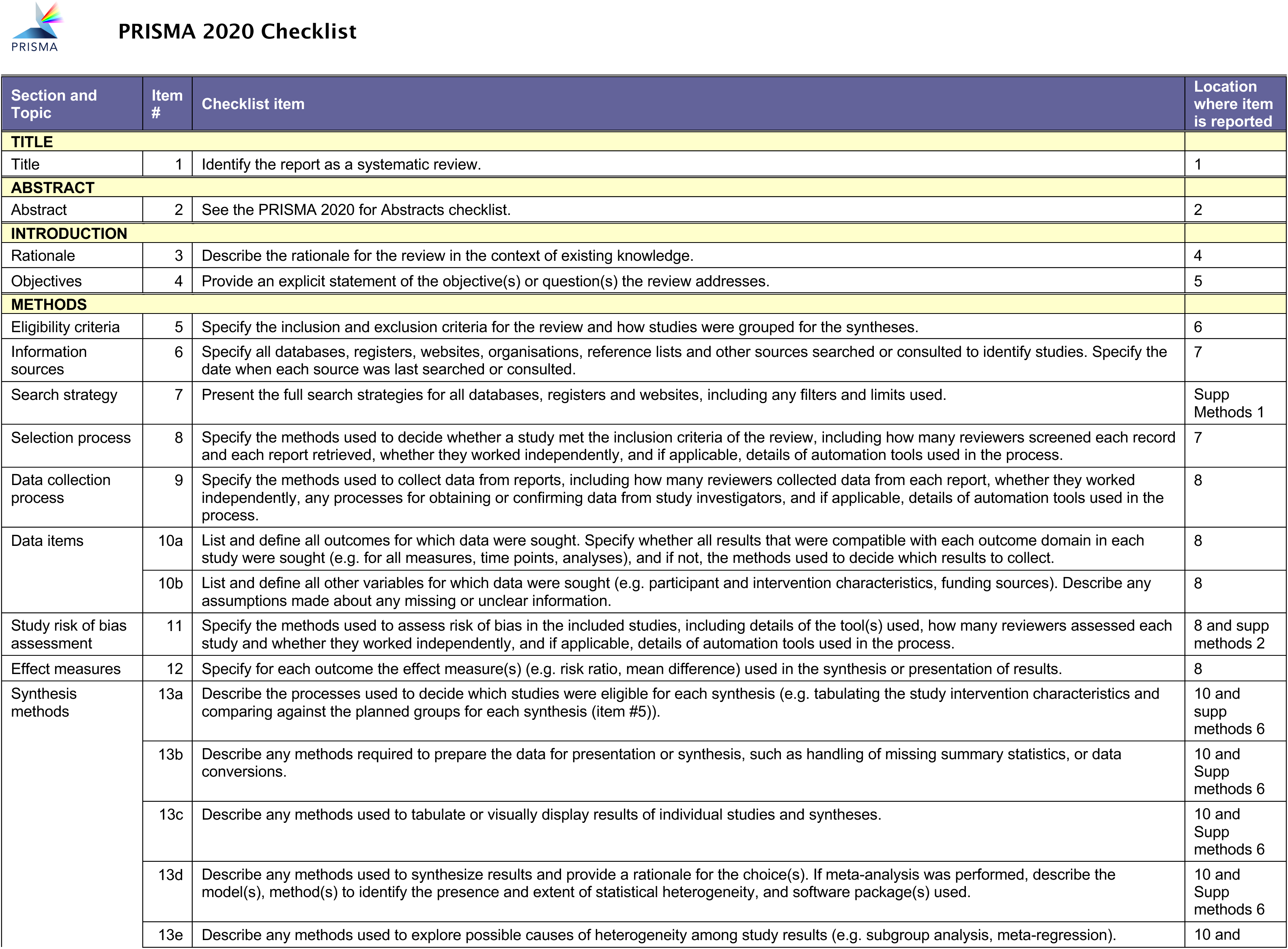
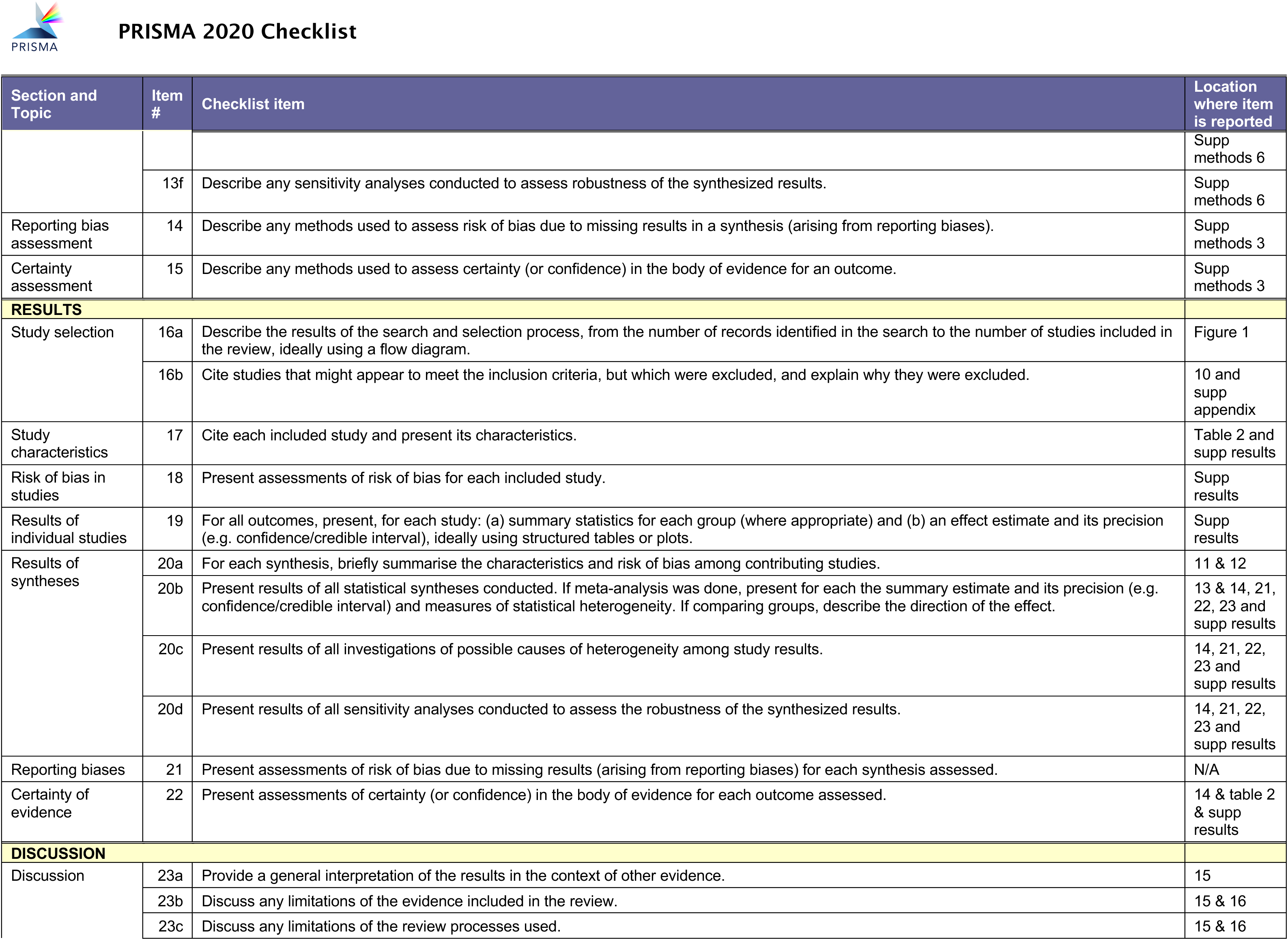
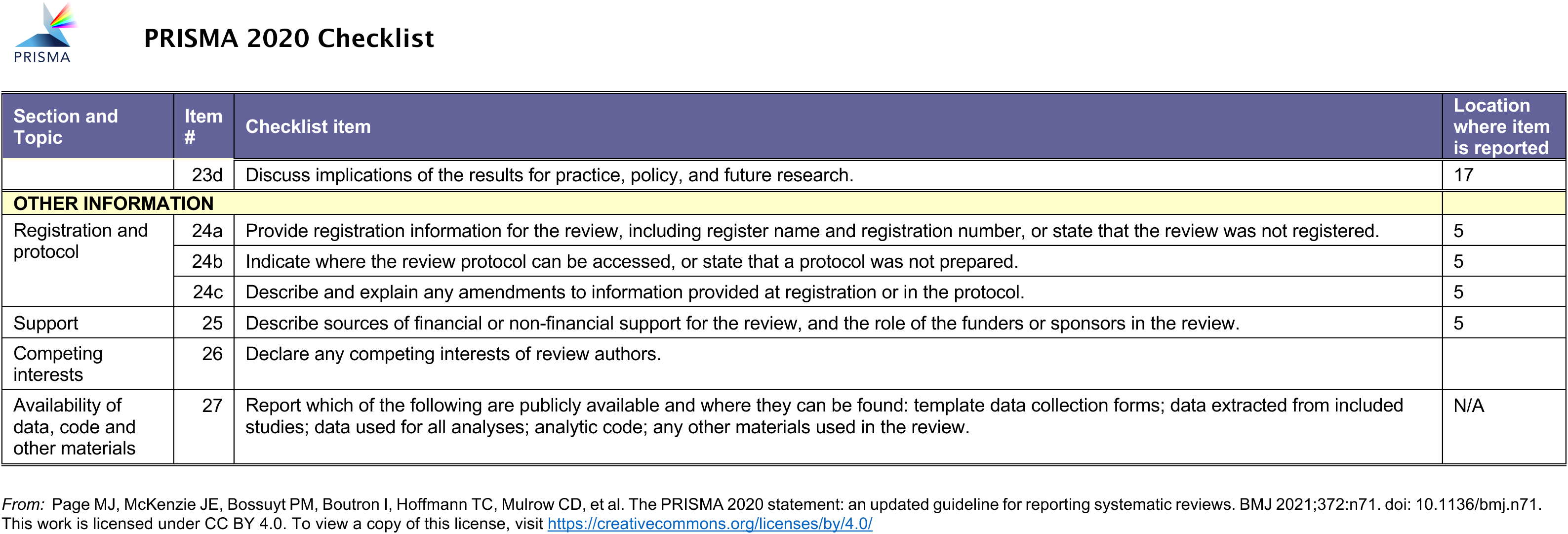

## Notes

### Competing Interest Statement

The authors have declared no competing interest.

### Clinical Protocols

https://www.crd.york.ac.uk/PROSPERO/view/CRD42023440141

### Funding Statement

This study was funded by the National Institute of Health and Care Research Health Technology Assessment (HTA), reference NIHR154660. The views expressed in this publication are those of the authors and not necessarily those of the NHS, the NIHR or the Department of Health.

